# What Interventions have been used in the UK in response to Winter Pressures? Mapping review of Studies/Initiatives relating to Discharge Planning and/or Integrated Care

**DOI:** 10.1101/2023.02.22.23286290

**Authors:** Anna Cantrell, Duncan Chambers, Andrew Booth

**Author notes:** Funding and registration: NIHR Health Services & Delivery Research Programme (project number NIHR 135773).

## Abstract

**Background:** Winter pressures are a familiar phenomenon within the NHS and represent the most extreme of many regular demands placed on health and social care service provision. This review focuses on a part of the pathway that is particularly problematic: the discharge process from hospital to social care and the community. Although studies of discharge are plentiful, we identified a need to focus on identifying interventions and initiatives that are a specific response to “winter pressures”. This mapping review will focus on interventions or initiatives in relation to winter pressures in the United Kingdom with either discharge planning to increase smart discharge (both a reduction in patients waiting to be discharged and patients being discharged to the most appropriate place) and/or integrated care.

**Methods:** We conducted a mapping review of UK evidence published 2018-2022. Initially, we searched MEDLINE, HMIC, Social Care Online, Social Sciences Citation Index and Kings Fund Library to find relevant interventions in conjunction with winter pressures. From these interventions we created a taxonomy of intervention types and draft map. A second broader stage of searching was then undertaken for named candidate interventions on Google Scholar. For each taxonomy heading we produced a table with definition, findings from research studies, local initiatives and systematic reviews, and evidence gaps.

**Results:** The taxonomy developed was split into structural, changing staff behaviour, changing community provision, integrated care, targeting carers, modelling and workforce planning. The last two categories were excluded from the scope. Within the different taxonomy sections we generated a total of 41 headings. These were further organised into the different stages of the patient pathway: hospital avoidance, alternative delivery site, facilitated discharge and cross-cutting. The evidence for each heading was summarised in tables and evidence gaps were identified.

**Conclusions:** Few initiatives identified were specifically identified as a response to winter pressures. Discharge to assess and Hospital at home interventions are heavily used and well-supported by the evidence but other responses, while also heavily used, were based on limited evidence. There is a lack of studies considering patient, family and provider needs when developing interventions aimed at improving delayed discharge. Additionally, there is a shortage of studies that measure the longer-term impact of interventions. Hospital avoidance and discharge planning are whole system approaches. Considering the whole health and social care system is imperative to ensure that implementing an initiative in one setting does not just move the problem to another setting.

**Limitations:** Time limitations for completing the review constrained the time for additional searches. This may carry implications for the completeness of the evidence base identified.

**Future work:** Further research to consider a realist review that views approaches across the different sectors within a whole system evaluation frame.

## Plain English Summary

The NHS struggles with winter pressures every year due to rises in respiratory and cardiovascular problems during the winter months. This mapping review aimed to map the evidence around what has been done to minimise winter pressures related to discharge planning and integrated care to make suggestions for future research. Good research evidence was identified for acute medical units, discharge to assess and hospital at home. The evidence for many other activities to minimise the impact of winter pressures was case studies, conference presentations or small low methodological quality research studies. The research has identified many different initiatives with diverse labels and the need to consider the effect implementing an initiative in one setting could have on another setting. It would be useful to consider further review work around what works for whom in what circumstances and why based on common mechanisms within the different initiatives and across the whole health and social care system.

## Scientific summary

### Introduction

The term “winter pressures” refers to “how hospitals cope with the challenges of maintaining regular service over the winter period”. Attention often focuses on additional demands on Accident and Emergency Services but pressures are exerted in terms of increased demand across the entire health and social care system. Increased prevalence of respiratory and cardiovascular illnesses during the winter places severe demands upon systems that already face difficulties in matching service provision to demand. Better or earlier planning and/or increasing service provision across integrated care systems, and related sectors such as housing, constitute just two of the possible responses to increased demand.

Winter pressures are a familiar phenomenon within the NHS and represent the most extreme of many regular demands placed on health service provision. The impact of winter pressures is pervasive and operates along a continuum from public health, in mitigation via immunisation campaigns, through to discharge into social care and the community. Interventions target multiple points along this continuum, as well as whole system approaches.

This mapping review focuses on one stage of the pathway, considered particularly problematic, namely the discharge process from hospital. Studies of discharge interventions and transfers of care are plentiful and international initiatives are reviewed in a recent scoping review. However, a considerable challenge lies in identifying interventions that specifically are articulated as an explicit response to “winter pressures”. There is strong rationale for focusing on the sub-group of “winter pressures” interventions as specific interventions implemented in response to acute and severe system pressure. This contrasts to interventions that seek to improve discharge within a system in “steady state”. While commentators on the current context articulate that the system experiences “winter pressures” all year round, until recently responses have targeted the context of winter pressures. Potentially, this set of studies therefore represents a highly relevant evidence base on discharge related interventions to inform current NHS planning.

This mapping review aims to chart and document the evidence in relation to winter pressures in the United Kingdom together with either discharge planning to increase discharge (both a reduction in patients waiting to be discharged and patients being discharged to the most appropriate place) and/or integrated care. For the purposes of this review “integrated care” involves partnerships of organisations that come together to plan and deliver joined up health and care services (NHS England).

The primary objective of the mapping review is to address the question:

- Which interventions in relation to discharge planning/integrated care have been suggested, tried or evaluated in seeking to address winter pressures in the United Kingdom?

The secondary review question is:

- Which research or evaluation gaps exist in relation to service- or system-level interventions as a response to discharge planning/integrated care in the context of winter pressures?

In addressing this question the mapping review will seek:

- To identify a potential winter pressures research agenda in relation to discharge planning/integrated care to inform commissioning of future research.

### Methods

We conducted a mapping review of UK evidence published 2018-2022. For the mapping review we used a two stage search process to search for the evidence. Initially, we searched MEDLINE, HMIC, Social Care Online, Social Sciences Citation Index and Kings Fund Library to find relevant interventions. The search was broad for terms for winter pressures. Searches on google scholar which search the full-text instead of just title and abstract included terms for discharge and integrated care. Study screening and selection was undertaken in excel by three reviewers who independently screened the title and abstracts of the 723 references which were retrieved by the search. Study eligibility was based on following (Population-Exposure-Comparative-Outcome(s)-Study Types i.e. PECOS) aspects:

- Users of UK health and/or social care systems (Population)
- Winter pressures impacting on discharge, to social care and the community, and integrated care (Exposure)
- Other foreseeable, unusual or exceptional periods of demand (if appropriate) (Comparison – may or may not be present)
- Increased smart discharge (both a reduction in patients waiting to be discharged and patients being discharged to the most appropriate place), system effects, health and health service outcomes, effects on patients, carers and staff (Outcomes)
- Eligible types of study design (primary research study, evidence synthesis or research report) (Study types)

To classify within the broader thematic groups of interventions, we developed a taxonomy (see the Excel spreadsheet) documenting the candidate interventions together with other relevant supporting literature. Our team started from categories developed by the Cochrane EPOC (Effective Practice and Organisation of Care) Group for their systematic reviews of discharge planning. These were further expanded using categories from a rapid review produced by the Centre for Clinical Effectiveness, Monash University. This process resulted in the following broad groupings: Structural (S), Changing Staff Behaviour (CSB), Changing Community Provision (CCP), Integrated Care, and Targeting Carers (TC). The draft taxonomy was reviewed for parsimony (to minimise duplication of concepts) and comprehensiveness (to include all named interventions identified to date). However, published commentary has documented the non-exclusivity and lack of precision of existing labels. Following the production of the draft map using the taxonomy we decided to further split the taxonomy headings to represent contributions to the patient pathway: hospital avoidance, alternate delivery site, facilitated discharge and cross-cutting. The modelling and workforce planning groupings were not considered as within scope and were therefore discarded.

The second stage of searching consisted of searches for named candidate interventions from the literature and current practice on Google Scholar. The second stage was to identify where possible reviews, ideally systematic reviews and these searches were broader than winter pressures but were limited to research published from 2012-2022. The second stage searches helped in completing the intervention tables and identifying the evidence gaps. Research priorities were classified as high, moderate or low and further classified by the nature of the evidence gap(s) identified (research gap, synthesis gap and/or implementation gap).

### Results

The taxonomy consists of a total of 41 headings. These were further organised into the different contributions to the patient pathway: hospital avoidance, alternative delivery site, facilitated discharge and cross-cutting. The evidence for each heading was provided and this helped with identification of the evidence gaps. Within structural interventions for the hospital avoidance part of the patient pathway research gap were identified for same day emergency care and research and implementation gaps for surgical hubs. The alternative delivery sites subsection is populated by systematic review evidence for the effectiveness of acute medical units, other specialist units developing using winter funding need to be fully evaluated. Models based on ‘discharge to assess’ (D2A, also ‘Home First’ and others) within facilitated discharge are relatively well researched. Some taxonomy headings, e.g. ‘bed management’ and ‘discharge co-ordinators’, were often evaluated within a broader process of ‘discharge planning’. ‘Patient flow’ is another broad heading with some overlap with both ‘bed management’ and discharge planning. The concept of ‘patient flow’ is also broader than facilitated discharge, although its ultimate goal is ensuring safe discharge as soon as is clinically appropriate. The evidence base for initiatives defined as ‘cross-cutting’ varied widely and was characterised by case studies with a lack of research studies. Community provision initiatives and integrated care were heterogeneous and characterised by multiple diverse initiatives, largely unevaluated, and by involvement of multiple contributors and sectors.

Overall, the evidence base is characterised by large numbers of case studies, often published online or presented at conferences, and relatively few peer-reviewed journal articles. Case studies are often accompanied by guidance to support implementation of changes to services. This distribution of evidence probably reflects the urgent need to develop and implement solutions to the ever increasing winter (and increasingly year-round) pressures on the health and care system. The majority of evaluations report positive effects on important outcomes such as length of hospital stay but many are uncontrolled or based on small samples, meaning that they need to be interpreted with caution.

### Conclusions

Few initiatives identified were specifically implemented as a response to winter pressures. Hospital at home, as a heavily used intervention, was well-supported by the evidence but other responses while also heavily used were based on limited evidence. There is a lack of studies considering patient, family and provider needs when developing interventions aimed at improving delayed discharge. Additionally, few studies measure the impact of interventions over a long time, short-term results can appear promising but evidence for longer-term sustainability is notably absent. Hospital avoidance and delayed discharge requires a whole system approach. It is imperative to consider the whole system to ensure that implementing an initiative in one setting does not just move the problem to another setting.

### Limitations

Time limitations for completing the review constrained the time for additional searches with a focus on systematic reviews and high-profile studies. This carries implications for the variability of coverage and completeness of the evidence base identified.

### Implications for service delivery

Effective interventions to avoid hospital admission, deliver services in different settings and facilitate discharge are key to reducing short-term acute pressures on health and social care. These pressures are generally associated with the winter period but have increasingly been experienced throughout the year and are particularly acute at the time of writing (January 2023). Longer-term improvements to service delivery may require policy changes related to investment and workforce planning that are outside the scope of this review.

### Implications for research

We identified high priority topics for primary research and evaluation in all the broad groupings of taxonomy headings as follows:

Changing community provision (CCP): Private sector, Step up facilities
Changing staff behaviour (CSB): Clinical audit, QI programmes, Protocols/guidelines, Quality management systems
Integrated care (IC): Integrated care discharge huddle
Structural (S): Bed management, Extra service delivery, Governance, Monitoring and review, Same day services, Specialist units, Volunteers

In terms of evidence synthesis, our detailed exploration further supports the need for a realist review that views approaches across the different sectors within a whole system evaluation frame. Further evidence synthesis should consider identified synthesis gaps in research within the aforementioned areas.

## Introduction

### Background

The term “winter pressures” refers to “how hospitals cope with the challenges of maintaining regular service over the winter period”. Although attention typically focuses on additional demands on Accident and Emergency Services, pressures are exerted in terms of increased demand across the entire health and social care system. Increased prevalence of respiratory and cardiovascular illnesses during the winter places severe demands upon systems that already face difficulties in matching service provision to demand. Better or earlier planning and/or increasing service provision across integrated care systems, and related sectors such as housing, constitute just two of the possible responses to increased demand.

The Covid 19 pandemic response illustrates many issues that need to be addressed in planning for hospital discharge when the health system faces specific expected or unexpected system pressure. Whilst very large numbers were rapidly discharged to care home beds in the early months of the pandemic, the discharge process generated significant concerns about whether the associated risks to both the discharged patients and the care homes had been sufficiently considered in the discharge planning process.

Winter pressures are a familiar phenomenon within the NHS and represent the most extreme of many regular demands placed on health service provision. The impact of winter pressures is pervasive and operates along a continuum from public health, in mitigation via immunisation campaigns, through to discharge into social care and the community. Interventions target multiple points along this continuum, as well as whole system approaches.

This mapping review focuses on one stage of the pathway, considered particularly problematic, namely the discharge process from hospital. Studies of discharge interventions and transfers of care are plentiful and international initiatives are reviewed in a recent scoping review. However, a considerable challenge lies in identifying interventions that specifically are articulated as an explicit response to “winter pressures”. There is strong rationale for focusing on the sub-group of “winter pressures” interventions because these may represent interventions implemented in response to particularly acute and severe system pressure. This contrasts to interventions that seek to improve discharge within a system in “steady state”. While commentators on the current context articulate that the system experiences “winter pressures” all year round, until recently responses have targeted the context of winter pressures. Potentially, this set of studies therefore represents a highly relevant evidence base on discharge related interventions to inform current NHS planning.

This mapping review aims to chart and document the evidence in relation to winter pressures in the United Kingdom together with either discharge planning to increase discharge (both a reduction in patients waiting to be discharged and patients being discharged to the most appropriate place) and/or integrated care^1^. The objectives are:

- To document and describe potential and evaluated interventions that seek to address issues and challenges in relation to discharge planning/integrated care in the context of winter pressures in the United Kingdom;
- To identify research or evaluation gaps relating to service- or system-level interventions in response to discharge planning/integrated care in the context of winter pressures;
- To identify a potential winter pressures research agenda in relation to discharge planning/integrated care to inform commissioning of future research.

### Objective of the Review

The primary objective of the mapping review is to address the question:

- Which interventions in relation to discharge planning/integrated care have been suggested, tried or evaluated in seeking to address winter pressures in the United Kingdom?

The secondary review question is:

- Which research or evaluation gaps exist in relation to service- or system-level interventions as a response to discharge planning/integrated care in the context of winter pressures?

In addressing this question the mapping review will seek:

- To identify a potential winter pressures research agenda in relation to discharge planning/integrated care to inform commissioning of future research.

## Methods

We undertook a systematic mapping review to map the winter pressure literature, specifically in relation to discharge planning and integrated care and, to help to determine opportunities for further research. The mapping review will aim to explore the volume and characteristics of the available evidence in relation to discharge planning or integrated care in response to winter pressures.

The systematic mapping review closely adhered to published methods for a mapping review. The methodology described by James and colleagues^4^ was used to guide the five stages of the review; setting the scope and inclusion criteria; searching for evidence; screening evidence; coding and database production; describing and visualising the findings.

### 1. Setting the scope and inclusion criteria

This mapping review focuses on interventions designed to reduce clinically unnecessary occupancy of hospital beds and demand on health services through discharge and transfer to social care or the community (e.g. “hospital at home”). Such interventions typically require improved linkages and communication between various health and social care services, captured in the concept of integrated care. Integrated care systems seek to capitalise on the benefits of such “joined-up” systems.

Key to feasibility is the requirement for study authors to have linked their research or data explicitly to “winter pressures”. Numerous interventions have been proposed to handle discharge planning but not all have been explored within the context of a pressurised health or social care system. Insisting on this requirement helped in identifying interventions or mechanisms that offer promise within other pressurised care environments. Specifically, we based eligibility on the following (Population-Exposure-Comparative-Outcome(s)-Study Types i.e. PECOS) aspects:

- Users of UK health and/or social care systems (Population)
- Winter pressures impacting on discharge, to social care and the community, and integrated care (Exposure)
- Other foreseeable, unusual or exceptional periods of demand (if appropriate) (Comparison – may or may not be present)
- Increased smart discharge (both a reduction in patients waiting to be discharged and patients being discharged to the most appropriate place), system effects, health and health service outcomes, effects on patients, carers and staff (Outcomes)
- Eligible types of study design (primary research study, evidence synthesis or research report) (Study types)

### 2. Searching for evidence

The mapping review took a two pronged approach to identify the evidence for this review. The first stage search identified the interventions through explicit references to winter pressures. The second stage of further searching was undertaken of named interventions extending beyond winter pressures. on Google Scholar to find systematic reviews and guidance. More detail is provided below.

#### First Stage Search

For the first stage search the literature search employed two separate methods according to the type of source being utilised. We searched databases that offer title and abstract search facilities using terminology relating to winter pressures. We examined retrieved items identified as initially relevant at full-text to establish whether they contain data within the context of discharge planning or integrated care. We searched data sources that enable full-text searching e.g. Google Scholar and other web-searches by combining the winter pressure terms with the twin concepts of discharge planning and/or integrated care.

##### Search terms and languages

The literature search covered the period 2018-2022 and was restricted to English only. This search cut-off date was determined by the need to optimise the relevance to the current context and ways of working. However, we have documented UK studies published before 2018 referenced in evidence syntheses or research reports as still being of contemporary relevance. International intervention in systematic reviews as mapped. We extracted findings from these studies for completeness.

**Concept 1:** Winter pressure(s); winter plan(s)/planning; winter resilience; Winter protection plan.

**Concept 2:** discharge plan(s)/planning; delayed transfer of care; discharge to assess; better care fund, increased smart discharge (both a reduction in patients waiting to be discharged and patients being discharged to the most appropriate place)

**Concept 3:** integrated care

An initial search was conducted in October on Medline and then translated to the other databases, the Medline search strategy is provided in Appendix 1. The search utilised free-text terms for winter pressures and winter planning. We were unable to identify specific relevant thesaurus terms for winter pressures. To optimise sensitivity of the search we did not use specific discharge/integrated care terminology in the search strategy but decided that judgements on discharge planning/integrated care context be operationalized during title/abstract and full-text screening stages rather than rely on uneven application of database indexing.

##### Databases searched

- Medline Ovid Database: Ovid MEDLINE(R) and Epub Ahead of Print, In-Process, In-Data-Review & Other Non-Indexed Citations, Daily and Versions <1946 to October 11, 2022>
- HMIC Ovid: HMIC Health Management Information Consortium <1979 to July 2022>
- Social Care Online
- SSCI: Social Sciences Citation Index (SSCI)—1900-present
- Kings Fund Library

Google scholar was also searched through Publish and Perish

The following organisation websites were also searched to identify report about initiatives to improve discharge planning or integrated care process:

- British Medical Association
- Department of Health and Social Care
- Health Foundation
- Kings Fund digital archive
- The Nuffield Trust

Searches on Google were also undertaken for winter pressures and discharge OR integrated care limited to NHS, GOV or org sites.

#### Second stage search

The second stage of searching was searches of google scholar for named candidate interventions from the literature and current practice. The searches were on Google Scholar for evidence broader than winter pressures. These searches were intended to retrieve reviews, ideally systematic reviews and were limited to research published from 2012-2022. These searches helped with completing the intervention tables and identifying the evidence gaps.

### 3. Screening evidence

Study screening and selection was undertaken in Microsoft Excel. A team of three reviewers independently screened the titles and abstracts of the 723 references from the searches. Articles meeting the inclusion criteria for titles and abstracts or items with insufficient detail on content relating to discharge planning or integrated care were reviewed at full-text.

#### Study validity assessment

The primary purpose of the mapping review was to create a profile of the available evidence base and this was then used to identify potential evidence and synthesis gaps. We assessed individual studies at a study design level and not through individual critical appraisal. Initially identified items were categorised as primary research, evidence synthesis, or research report. The last of these identified reports from organisations such as the Nuffield Trust, King’s Fund, Health Foundation that combine critical commentary, usually with primary data. Outputs from governmental or professional organisations will be processed in a similar way. We identified research gaps from the distribution of primary studies and their research questions, summaries from evidence syntheses and recommendations from research reports. We then evaluated the authority and credibility of the research gaps accordingly.

### 4. Coding and database production

Meta-data extraction and coding for studies was undertaken using Microsoft Excel. For mapping purposes, references were categorised at title and abstract stage according to study design, characteristics and broad thematic content. A data coding spreadsheet was used to code data for study type, population, intervention (if present), outcomes or findings and conclusion. Following granular coding we then classified related studies within broader thematic categories that were be used to present study characteristics and to structure an accompanying narrative. Due to the time and resource constraints of the proposed work, missing or unclear information was not solicited from authors.

### 5. Describing and visualising the findings

#### Development of taxonomy

To classify into the broader thematic groups of interventions, we developed a taxonomy (see the Excel spreadsheet) documenting the candidate interventions together with other relevant supporting literature. Our team started from categories developed by the Cochrane EPOC (Effective Practice and Organisation of Care) Group for their systematic reviews of discharge planning (cited by Parker et al. 2002). These were further expanded using categories from a rapid review produced by the Centre for Clinical Effectiveness, Monash University.

This process resulted in the following broad groupings: Structural (S), Changing Staff Behaviour (CSB), Changing Community Provision (CCP), Integrated Care, and Targeting Carers (TC). The draft taxonomy was reviewed for parsimony (to minimise duplication of concepts) and comprehensiveness (to include all named interventions identified to date). However, published commentary has documented the non-exclusivity and lack of precision of existing labels. Following the production of the draft map using the taxonomy we decided that it would be useful to split the taxonomy headings into the following categories: hospital avoidance, alternate delivery site, facilitated discharge and cross-cutting. The Integrated Care and Targeting Carers groupings were not used and were therefore discarded,

#### Producing the map

Once the taxonomy was developed, tested and agreed we mapped candidate interventions to the taxonomy to produce the draft map. Following changes to the taxonomy we updated the draft map to produce the final map

#### Stakeholder involvement

During the mapping review, we consulted with stakeholders at the Department of Health and Social Care and the National Institute for Health and Care Research as well as with a standing advisory group of patients and carers. This ensured that the review of winter pressures was informed by the views and experiences of stakeholders. The aim of this consultation was to:

- illuminate the practical and personal challenges experienced by those planning or using health services during times of exceptional demand.
- inform the second phase of our search strategies by identifying specific strategies, initiatives and interventions that target discharge planning and integrated care in the context of winter pressures
- identify gaps in the literature with a view to compiling a future research agenda.

#### Patient and Public Involvement (PPI)

A PPI meeting with members of the Evidence Synthesis Centre standing public advisory group was conducted on 30^th^ November 2022 to discuss the mapping review. The meeting aimed to ascertain the groups’ thoughts and experiences around discharge planning and integrated care interventions to minimise winter pressures. To help bring out their thoughts and opinion we developed five scenarios around interventions found in the literature and asked what that liked about each approach and what are the drawbacks or what might have been overlooked. The five interventions used in developing the scenarios were Discharge to Assess, Same Day Emergency Care, Hospital Avoidance Response Team, Care & Repair and Telecare. The scenarios are provided in Appendix 2.

The PPI group met on 30^th^ November online and five members attended.

The groups’ thoughts and experiences are summarised and presented for each of the scenario interventions.

##### Discharge to Assess (D2A)

Good to free up bed space so not delaying planning operations because of shortage of beds. Concern about whether patients are properly assessed before discharge. Positive for patient to be out of hospital but there needs to be a care plan in place and they are time consuming and shouldn’t be rushed. Important to ensure that the patient will be safe in their own home or a care home.

##### Same Day Emergency Care (SDEC)

Concerned about discharge home with new medication who will follow-up, do GPs have time? If a patient is living alone who can chase for follow-up. Patients could need support in addition to a care. Care plans needed different if patient lives alone, has support. Ok if GP proactive response but usually need to keep asking due to pressure they are under. Potential side effects new medication. Concerns that the availability of services differ so much from area to area and we have to think about people living in rural areas

##### Hospital Avoidance Response Team

Rapid response good if reduces pressures on ambulances. Outcomes could depend upon paramedic training and expertise. Would paramedics be liaising with consultants from appropriate specialities? Would paramedics understanding medications and how can interact other medications. One group member discussed how paramedics specially trained in own area, very good, extra skills & knowledgeable. Issue of continuity of care after paramedic, carers need reliable ongoing support.

##### Care & repair

One group member discussed how a family member in Bristol had used the service 10 years ago for alterations after knee replacement. Not available everywhere particularly in rural areas. Repairs can take time, particularly given a shortage of workmen; typically work needs to be completed promptly. A real worry is cold homes and excess winter deaths. Energy bills and cost of living. Third sector – great if reliable but services in different areas are very variable.

##### Telecare

Monitors, wearables useful, good involve patients in reading good to use & assurances reduce usage of services. Good opportunity. Care home staff sending to A&E as not properly trained. Good idea but practicality staff not trained to use tech, staff need training in new equipment, depends care homes and staffing. Shortage care home staff. Staff training and turnover. Big role for telecare moving forward, unavoidable and important to embrace technology. Would assessment by diabetic nurse by zoom or skype be enough could need to be in person, can patient express herself thoroughly online, issue communication. Wearables/patient apps good maybe not for all conditions sometimes need F2F. Who would be monitoring in care home. Rising elderly population more demands tech but shouldn’t be fully dependent. Technology needs to be supported.

At the end the meeting the group voted on which intervention they thought it would be most useful to fund future research into, one group member chose not to vote. Three of the group voted for telecare believing that delivery of healthcare is moving in this direction and that it is important to have research to ensure it can be used effectively and to determine what works for different people and in which circumstances. The other group member voted for hospital avoidance response team emphasising their belief in the importance of paramedics in helping to mitigate winter pressures. Paramedics have been specially trained where they live and with the long waiting times for an ambulance to arrive or with patients at A&E waiting to be seen the role of paramedics is becoming more important.

## Results

The initial ‘winter pressures’ search identified 723 items, of which 117 were judged to be relevant and used for the draft map. Subsequent targeted searches identified a further 62 items related to the different taxonomy headings, together with references to guidance and web sites (see tables 2 to 83).

### Findings -Taxonomy

The final taxonomy is presented in Table 1. The taxonomy reflects the focus of the review on supporting early discharge where safe and appropriate but also includes interventions aimed at avoiding hospital admission through rapid response or delivering care in other settings. The ‘cross-cutting’ category groups interventions that could be delivered at different stages of the patient pathway and/or different applications of a particular technology (e.g. digital and data).

**Table 1.**
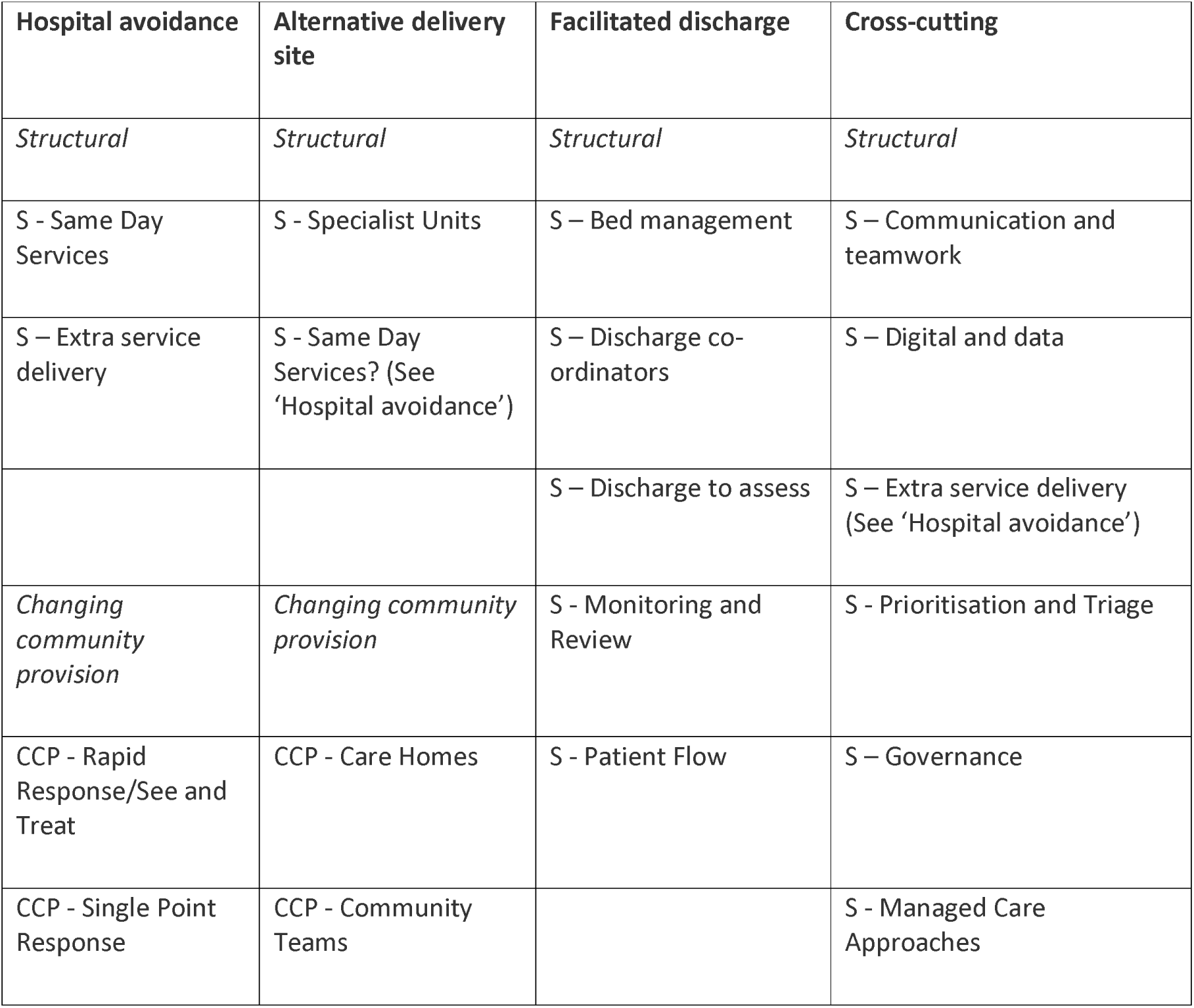
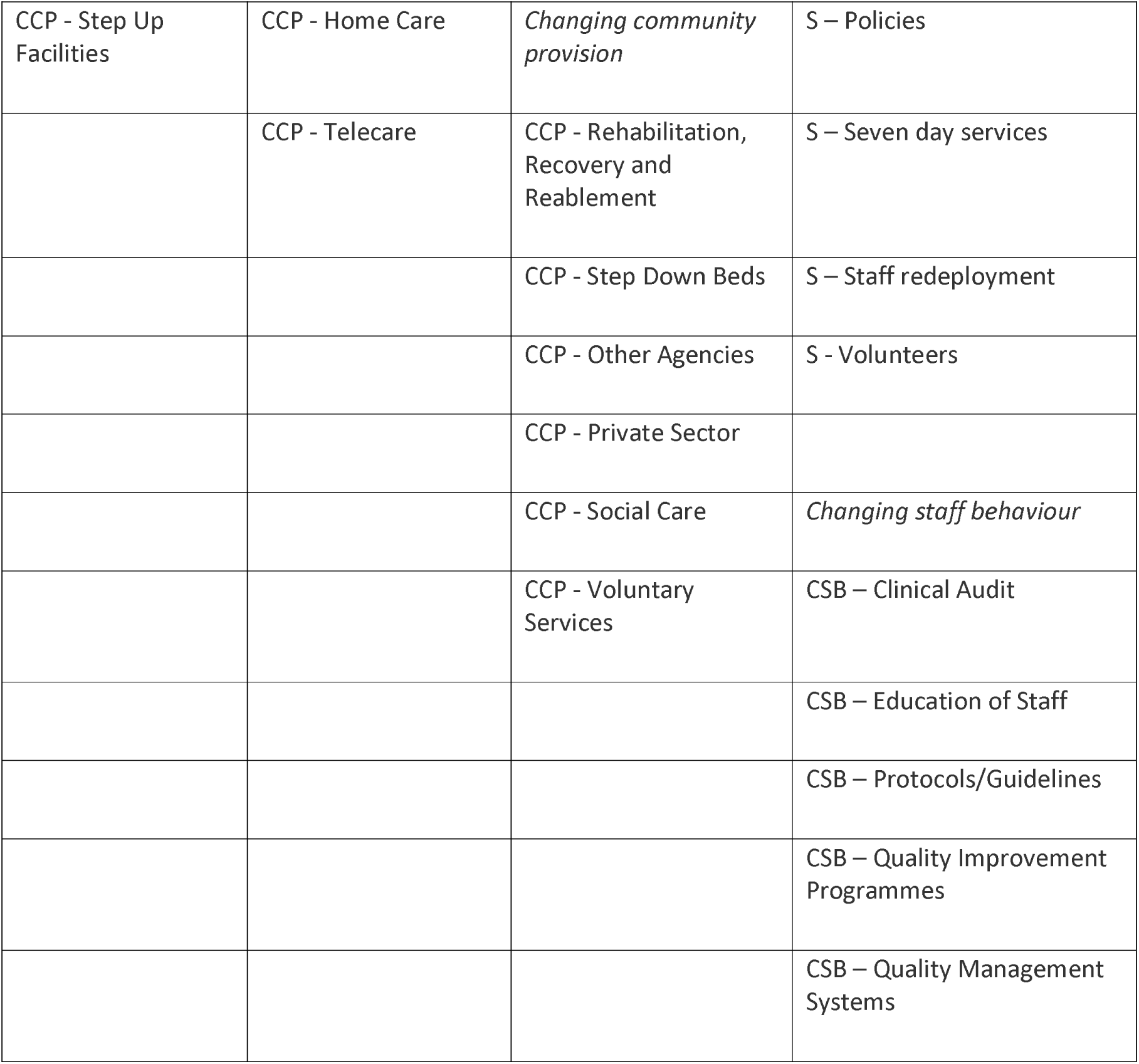
Final taxonomy of candidate interventions.

### Findings – Map

The final map is provided below and submitted as an accompanying PDF.

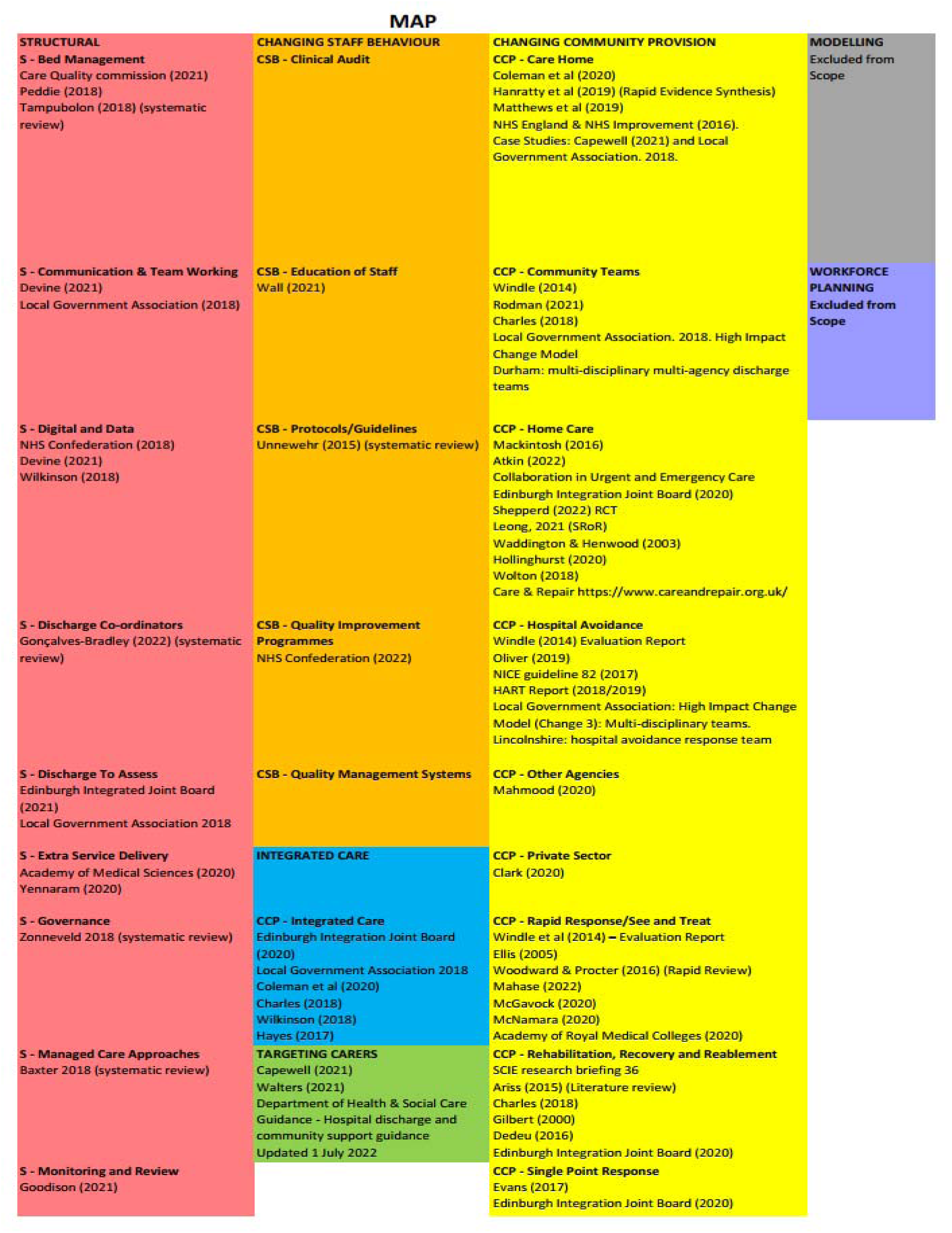

### Findings – Taxonomy Headings/Interventions

Findings are presented in the following tables following the order of the taxonomy presented in Table 1. Definitions and sample interventions are presented first, followed by a separate table of supporting evidence and identified research gaps under each taxonomy heading.

### Structural Interventions

Findings for taxonomy headings classified as structural are summarised in Tables 2 to 34. Tables 2 to 5 cover hospital avoidance; Tables 6 and 7 alternative delivery sites; Tables 8 to 17 cover facilitated discharge; and Tables 18 to 34 cover cross-cutting headings. Detailed information can be found in the tables, with brief overviews before and after each group.

#### Structural Interventions – Hospital Avoidance

The main taxonomy headings for hospital avoidance are same day services (Tables 2 and 3); extra service delivery (Tables 4 and 5). See also seven-day services (Tables 27 and 28) in the Structural Interventions – Cross-cutting section (page 61)).

##### S – Same Day Services

**Table 2.** Same Day Services: Definitions and Rationales.

| Taxonomy heading | Brief description/definition | Mechanism for minimising winter pressures |
| --- | --- | --- |
| S - Same Day Services | Services provided the same day without need for hospital admissions. | Same day services could help with winter pressures by reducing admissions. |
| Example interventions:<br>Same day emergency care (SDEC)/ Ambulatory emergency care (AEC)/ Ambulatory Care Unit Surgical Hubs/Elective surgical hubs/High Volume Low Complexity (HVLC) surgical hubs. | SDEC - provision of same day emergency care for patients otherwise admitted to hospital. Patients that present with relevant conditions can be rapidly assessed, diagnosed and treated without needing to be admitted to a ward and if a clinician deems clinically safe they can go home the same day. | SDEC aims to benefit patients and the healthcare system by reducing waiting times and hospital admissions where appropriate. Winter 2018/19 witnessed an increased use of ambulatory care units. |
|  | Surgical hubs - located at existing hospital sites and provide common procedures performed relatively quickly and effectively in one place, with patients able to return home the same day. | Surgical hubs help to deal with the rise in healthcare provision needed over the winter by enabling surgeons to continue to treat patients for routine operations preventing waiting list backlogs. |

**Table 3.**
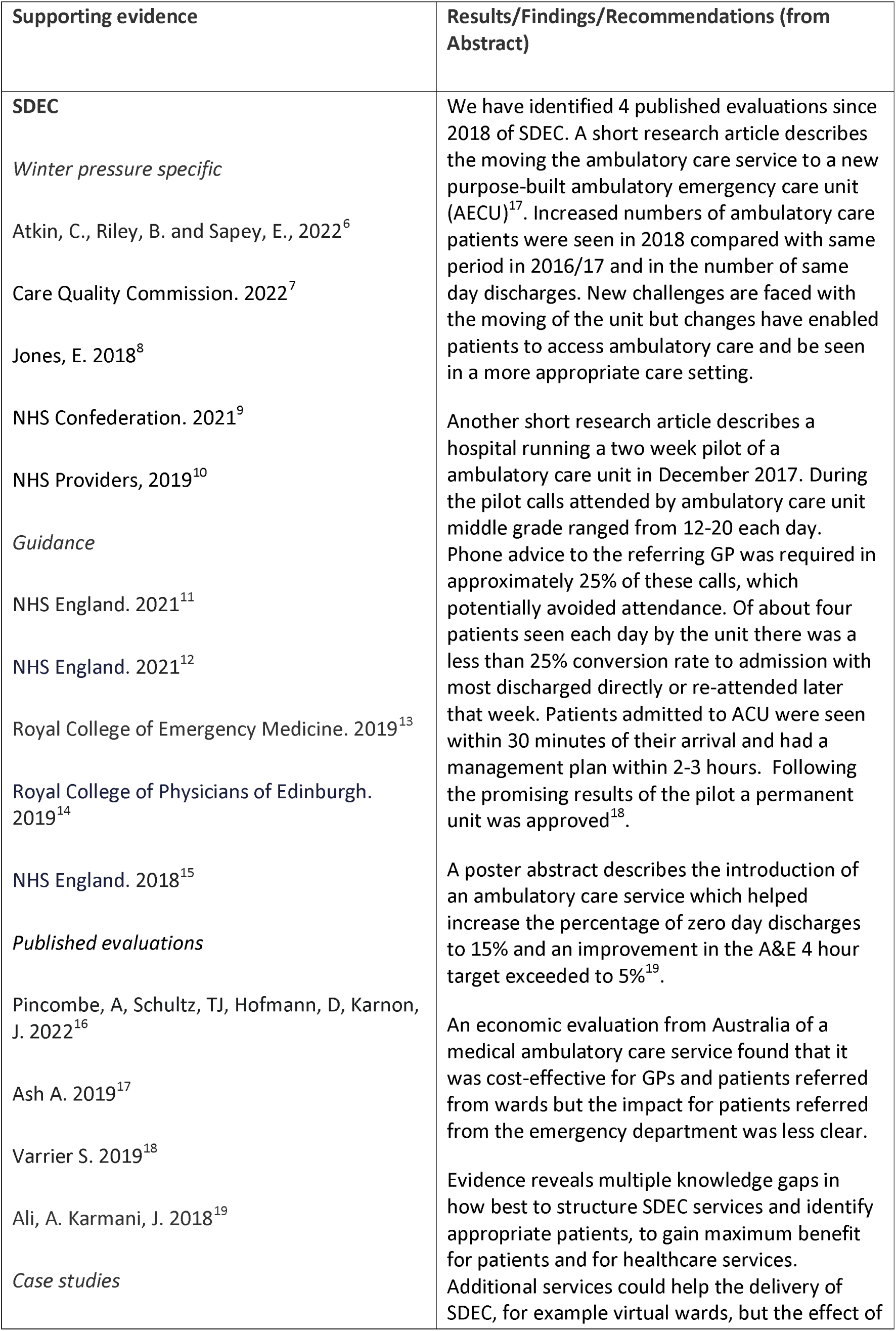
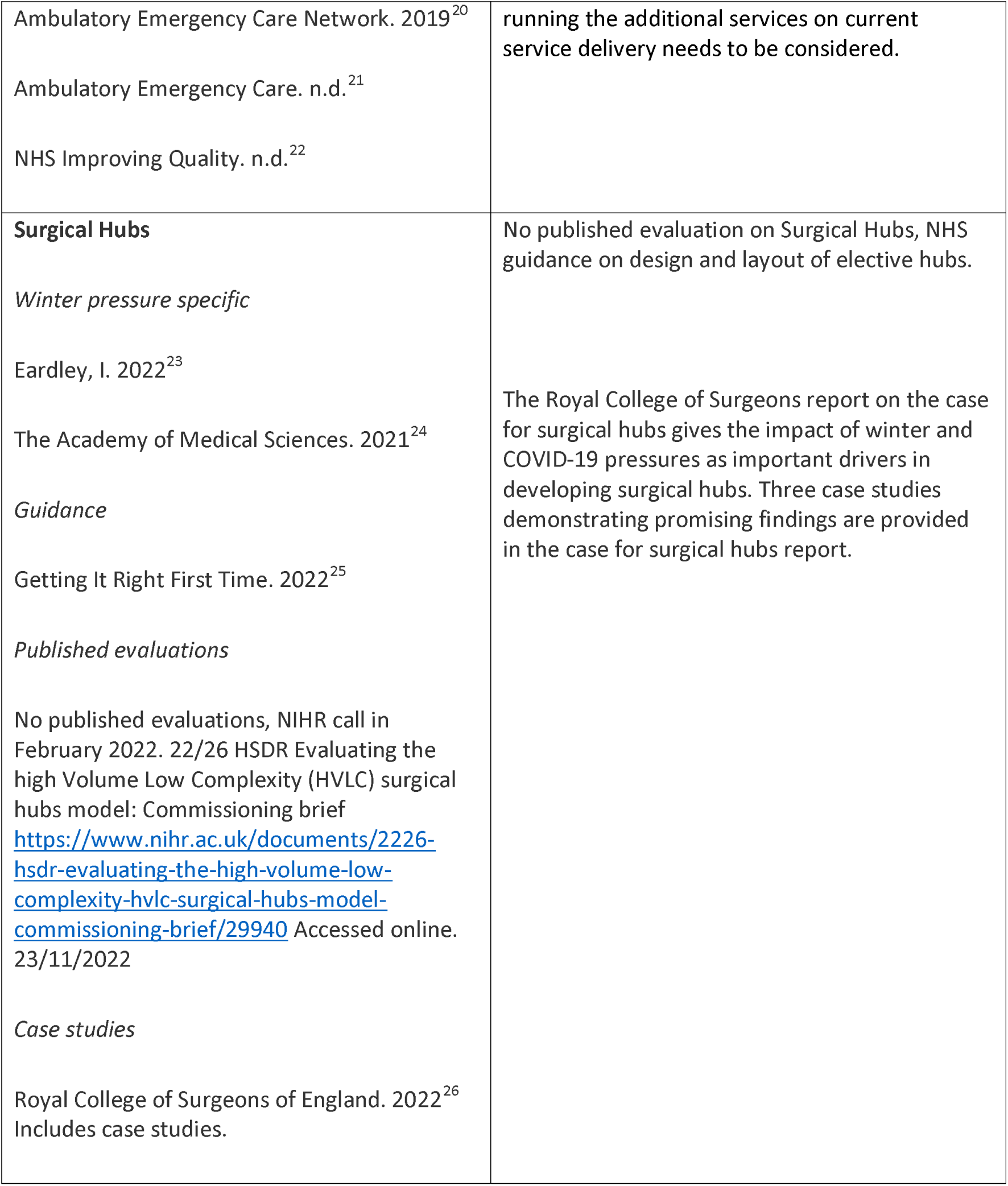
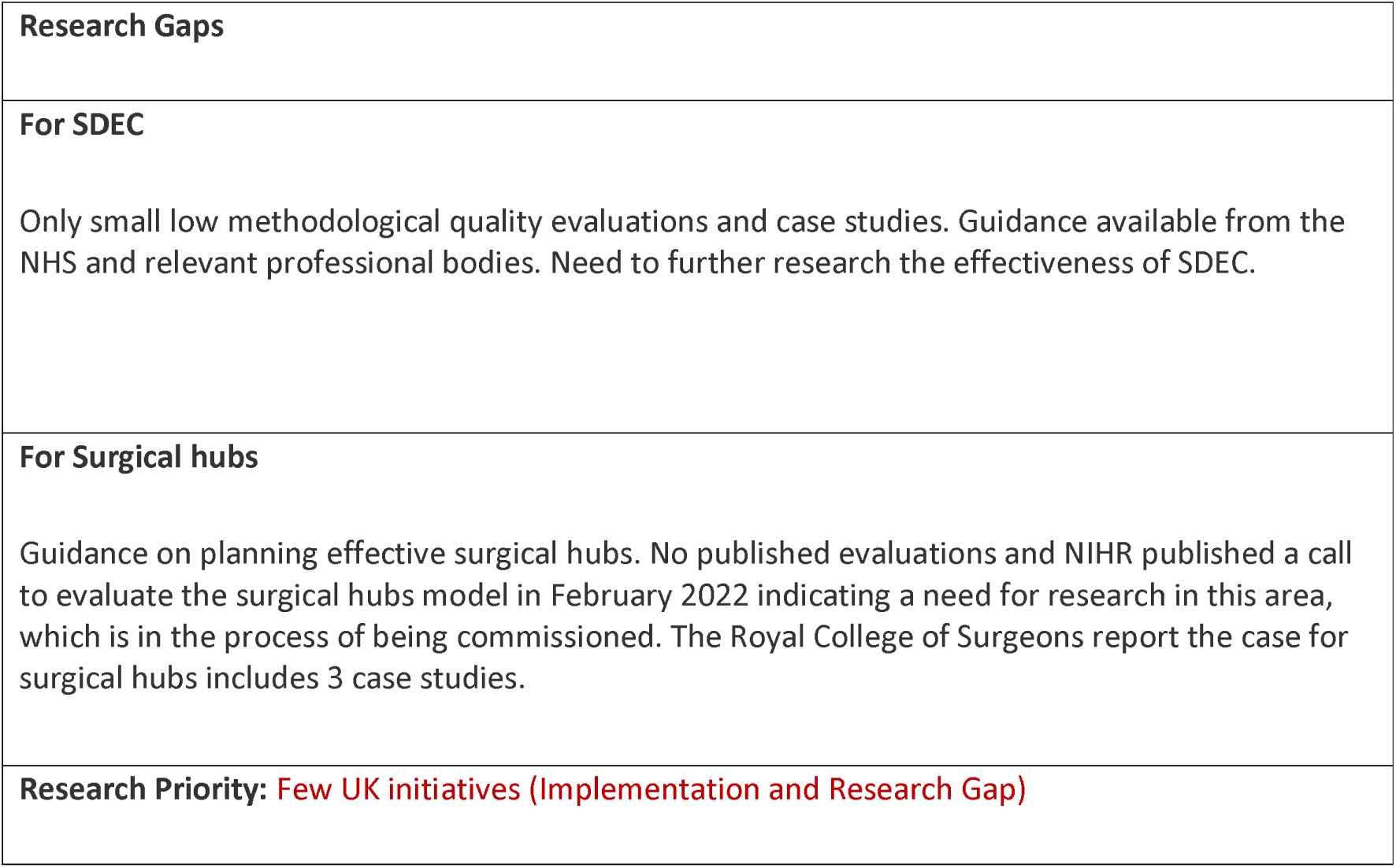
Same Day Services: Interventions and Supporting Evidence.

| Supporting evidence | Results/Findings/Recommendations (from Abstract) |
| --- | --- |
| **SDEC**<br>*Winter pressure specific*<br>Atkin, C., Riley, B. and Sapey, E., 2022<sup>6</sup><br>Care Quality Commission. 2022<sup>7</sup><br>Jones, E. 2018<sup>8</sup><br>NHS Confederation. 2021<sup>9</sup><br>NHS Providers, 2019<sup>10</sup><br>*Guidance*<br>NHS England. 2021<sup>11</sup><br>NHS England. 2021<sup>12</sup><br>Royal College of Emergency Medicine. 2019<sup>13</sup><br>Royal College of Physicians of Edinburgh. 2019<sup>14</sup><br>NHS England. 2018<sup>15</sup><br>*Published evaluations*<br>Pincombe, A, Schultz, TJ, Hofmann, D, Karnon, J. 2022<sup>16</sup><br>Ash A. 2019<sup>17</sup><br>Varrier S. 2019<sup>18</sup><br>Ali, A. Karmani, J. 2018<sup>19</sup><br>*Case studies* | We have identified 4 published evaluations since 2018 of SDEC. A short research article describes the moving the ambulatory care service to a new purpose-built ambulatory emergency care unit (AECU)<sup>17</sup>. Increased numbers of ambulatory care patients were seen in 2018 compared with same period in 2016/17 and in the number of same day discharges. New challenges are faced with the moving of the unit but changes have enabled patients to access ambulatory care and be seen in a more appropriate care setting.<br>Another short research article describes a hospital running a two week pilot of a ambulatory care unit in December 2017. During the pilot calls attended by ambulatory care unit middle grade ranged from 12-20 each day. Phone advice to the referring GP was required in approximately 25% of these calls, which potentially avoided attendance. Of about four patients seen each day by the unit there was a less than 25% conversion rate to admission with most discharged directly or re-attended later that week. Patients admitted to ACU were seen within 30 minutes of their arrival and had a management plan within 2-3 hours. Following the promising results of the pilot a permanent unit was approved<sup>18</sup>.<br>A poster abstract describes the introduction of an ambulatory care service which helped increase the percentage of zero day discharges to 15% and an improvement in the A&E 4 hour target exceeded to 5%<sup>19</sup>.<br>An economic evaluation from Australia of a medical ambulatory care service found that it was cost-effective for GPs and patients referred from wards but the impact for patients referred from the emergency department was less clear.<br>Evidence reveals multiple knowledge gaps in how best to structure SDEC services and identify appropriate patients, to gain maximum benefit for patients and for healthcare services. Additional services could help the delivery of SDEC, for example virtual wards, but the effect of |
| Ambulatory Emergency Care Network. 2019<sup>20</sup><br>Ambulatory Emergency Care. n.d.<sup>21</sup><br>NHS Improving Quality. n.d.<sup>22</sup> | running the additional services on current service delivery needs to be considered. |
| **Surgical Hubs**<br>*Winter pressure specific*<br>Eardley, I. 2022<sup>23</sup><br>The Academy of Medical Sciences. 2021<sup>24</sup><br>*Guidance*<br>Getting It Right First Time. 2022<sup>25</sup><br>*Published evaluations*<br>No published evaluations, NIHR call in February 2022. 22/26 HSDR Evaluating the high Volume Low Complexity (HVLC) surgical hubs model: Commissioning brief https://www.nihr.ac.uk/documents/2226-hsdr-evaluating-the-high-volume-low-complexity-hvlc-surgical-hubs-model-commissioning-brief/29940 Accessed online. 23/11/2022<br>*Case studies*<br>Royal College of Surgeons of England. 2022<sup>26</sup> Includes case studies. | No published evaluation on Surgical Hubs, NHS guidance on design and layout of elective hubs.<br>The Royal College of Surgeons report on the case for surgical hubs gives the impact of winter and COVID-19 pressures as important drivers in developing surgical hubs. Three case studies demonstrating promising findings are provided in the case for surgical hubs report. |

|  |
| --- |
| **Research Gaps** |
| **For SDEC**<br>Only small low methodological quality evaluations and case studies. Guidance available from the NHS and relevant professional bodies. Need to further research the effectiveness of SDEC. |
| **For Surgical hubs**<br>Guidance on planning effective surgical hubs. No published evaluations and NIHR published a call to evaluate the surgical hubs model in February 2022 indicating a need for research in this area, which is in the process of being commissioned. The Royal College of Surgeons report the case for surgical hubs includes 3 case studies. |
| **Research Priority:** Few UK initiatives (Implementation and Research Gap) |

##### S – Extra Service Delivery

**Table 4.** Extra Service Delivery: Definitions and Rationales.

| **Taxonomy heading** | **Brief description/definition** | **Mechanism for minimising winter pressures** |
| --- | --- | --- |
| S – Extra service delivery | Provision of extra resources to support service delivery at times of increased demand (as distinct from changing the time/place of service delivery) | Given the availability of staff, extra services can be planned, implemented and discontinued at relatively short notice in response to (expected) peaks/troughs in demand. |
| Example interventions: | ‘Super Saturday’ clinics (extra sessions to tackle elective treatment waiting lists; also used for COVID-19 and influenza vaccination); Rapid response team (late shift team to support same-day discharge from emergency care) | Reduction of elective waiting lists may prevent patients’ conditions worsening and reduce future demand. Extra resources in emergency care can allow earlier discharge and reduce demand for in-patient beds |

**Table 5.**
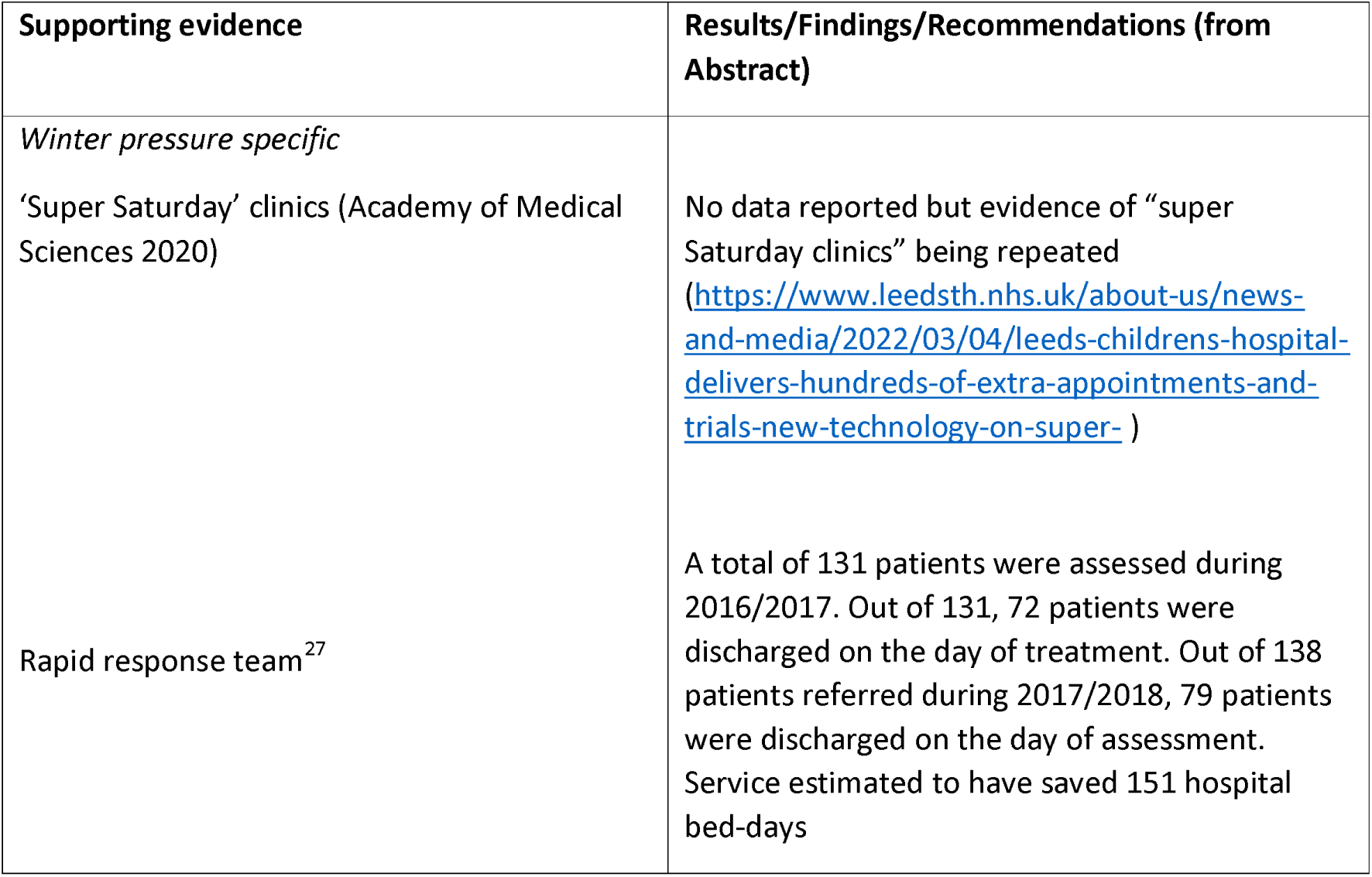
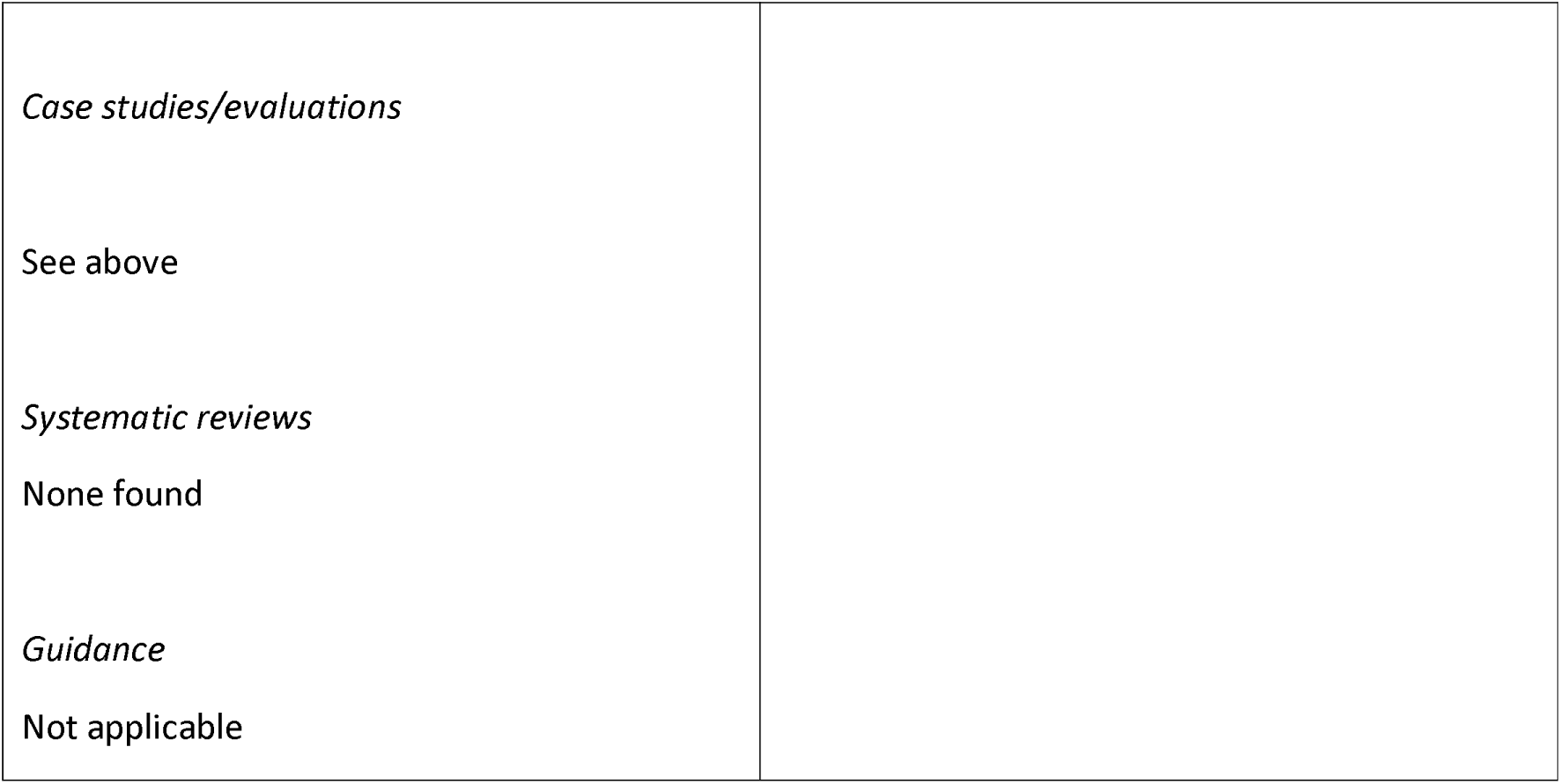
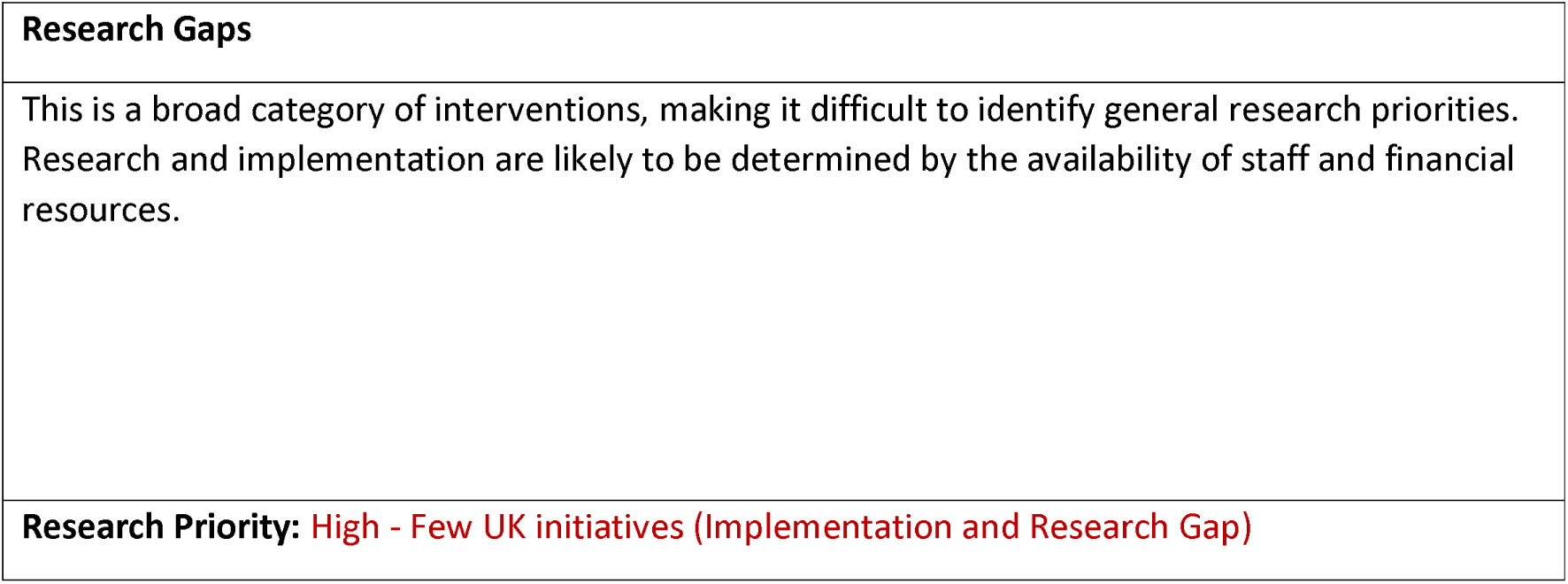
Extra Service Delivery: Interventions and Supporting Evidence.

| **Supporting evidence** | **Results/Findings/Recommendations (from Abstract)** |
| --- | --- |
| *Winter pressure specific*<br>‘Super Saturday’ clinics (Academy of Medical Sciences 2020)<br>Rapid response team<sup>27</sup> | No data reported but evidence of “super Saturday clinics” being repeated (https://www.leedsth.nhs.uk/about-us/news-and-media/2022/03/04/leeds-childrens-hospital-delivers-hundreds-of-extra-appointments-and-trials-new-technology-on-super- )<br>A total of 131 patients were assessed during 2016/2017. Out of 131, 72 patients were discharged on the day of treatment. Out of 138 patients referred during 2017/2018, 79 patients were discharged on the day of assessment. Service estimated to have saved 151 hospital bed-days |
| *Case studies/evaluations* |  |
| See above |  |
| *Systematic reviews* |  |
| None found |  |
| *Guidance* |  |
| Not applicable |  |

|  |
| --- |
| **Research Gaps** |
| This is a broad category of interventions, making it difficult to identify general research priorities. Research and implementation are likely to be determined by the availability of staff and financial resources. |
| **Research Priority:** High - Few UK initiatives (Implementation and Research Gap) |

Same day services fall into two broad categories: same day emergency care followed by discharge without hospital admission and routine surgery performed on a day case basis, with patients returning home on the day of surgery. Both have been widely adopted and are supported by relevant guidance. However a clear need remains for further research and evaluation (see Table 3).

Extra service delivery takes a variety of forms and some successes have been reported in both emergency and elective care (Table 5) but the evidence base remains limited.

#### Structural Interventions – Alternate delivery site

The heading ‘Alternative delivery site’ covers a variety of specialist units delivering services outside general hospitals or in demarcated areas within them. Examples are acute medical units and short stay units.

##### S – Specialist Units

**Table 6.** Specialist Units: Definitions and Rationales.

| **Taxonomy heading** | **Brief description/definition** | **Mechanism for minimising winter pressures** |
| --- | --- | --- |
| S – Specialist Units | Specialists units are units to deliver a particular service. | Specialist units can provide services separately to hospitals which reduce admissions, reduce infection by keeping patient groups separate, enable patients to receive the care they require. |
| Surgical Hubs/Elective surgical hubs/High Volume Low Complexity (HVLC) surgical hubs | Surgical hubs are located at existing hospital sites and provide common procedures that can be performed relatively quickly and effectively in one place or high volume low complexity surgeries. They will bring together staff with the required skills and expertise in one place. | Surgical hubs help to deal with the rise in healthcare provision needed over the winter by enabling surgeons to continue to treat patients for routine operations preventing waiting list backlogs. Improve infection control by keeping these patients separate from the emergency department and other wards in the acute hospital. Reduce number of operation being cancellations at short notice when hospitals are full. |
| Acute Medicine Urgent Access Clinic/Acute Medicine Clinic | The Clinic allows early outpatient review of symptoms and outstanding investigations. This means that clinicians can discharge patients sooner with an appointment to attend the clinic. | Help Drs to discharge patients earlier from ED, Acute Medical Units and expedite discharge from post-take ward round helping the manage increased healthcare usage during the winter months. |
| Acute Medical Unit (AMU)/Acute Assessment Unit (AAU)/Medical | AMUs provide rapid assessment investigating and treatment for acutely ill medical patients through referral by the emergency department, their GP or | Reduces patients going to the emergency department during this busy time of year which helps with patient flow |
| Assessment Unit (MMU) | outpatient services. Patient can receive acute medical assessment in one unit staffed by specialist medical and nursing personnel with equipment required for the assessment. Patients can be admitted for a short hospital stay for ongoing assessment or discharged home the same day. | throughout the hospital. |
| CRT+ & Long-COVID Single Point of Access (SPOA) | To provide a triage point to AHP rehabilitation services for people recovering from COVID-19. | Provide triage for people recovering from COVID-19 without them disrupting flow within health and social care. |
| Frail Elderly Short Stay Unit (FESSU) | Short-stay unit that assesses frail older people as soon as they come into hospital to ensure that they get the best possible care and treatment when they are in hospital and are supported (e.g. through comprehensive geriatric assessment) to recover when they leave. | Help with flow of frail older people through health and social care services, useful in winter when their healthcare usage could increase. |

**Table 7.** Specialist Units: Interventions and Supporting Evidence.

| Supporting evidence | Results/Findings/Recommendations (from Abstract) |
| --- | --- |
| Acute Medicine Urgent Access Clinic<br>*Winter pressure specific*<br>Cooper, M., 2019<sup>28</sup><br>*Additional evidence*<br>Case studies<br>Royal College of Physicians. 2018<sup>29</sup> | A recent survey of medical doctors at Warrington Hospital on the introduction of new acute medicine urgent access clinic found promising experiences. 93% of doctors believed that the introduction of the clinic allowed them to discharge patients earlier from the acute take, 77% believed that the clinic enabled quicker discharge from the emergency department, 79% of the doctors believed the clinic had sped up discharge from post-take ward round and 87% that it had sped up discharge from the acute medical unit. Further details of the introduction of the clinic at |
|  | Warrington and Halton hospitals NHS foundation Trust are provided in a case study in the RCP Chief Registrar Scheme 2017/18 year book. |
| Acute Medical Unit<br>*Winter pressure specific*<br>Peddie, G.J. and Gordon, C., 2018<sup>30</sup>.<br>*Additional evidence*<br>Systematic reviews<br>van Galen, L.S., Lammers, E.M.J., Schoonmade, L.J., Alam, N., Kramer, M.H.H. and Nanayakkara, P.W.B., 2017<sup>31</sup><br>Reid, L.E., Dinesen, L.C., Jones, M.C., Morrison, Z.J., Weir, C.J. and Lone, N.I., 2016<sup>32</sup><br>Guidance<br>UK, N.G.C., 2018<sup>33</sup><br>Published evaluations<br>Case studies | Two reviews published after 2012 were retrieved on acute medical units. A literature review from van Galen et al<sup>31</sup> concluded that current literature demonstrates an overall beneficial effect after implementation. The systematic review by Reid et al<sup>32</sup> concluded that acute medical units were associated with reductions in hospital length of stay and less convincingly mortality when implemented in European and Australasian settings.<br>A prospective cohort study<sup>30</sup> found results that suggest the median LoS, for the patients moved “out of hours” was significantly less than that of the patients moved “within” hours group. This could be due to the higher number of “boarders” in the “out of hours” group (36/57 patients = 63.2%) when compared to the “within hours” group (32/162 patients = 19.8%). Boarding while unacceptable still exists and it is not only related to winter months. |
| CRT+ & Long-COVID Single Point of Access (SPOA)/Post-COVID assessment services<br>*Winter pressure specific*<br>*Edinburgh Integration Joint Board. 2021<sup>34</sup>*<br>*Additional evidence* | The Edinburgh Integration Joint Boards evaluation of winter planning 2020/21 provides brief outcomes of funding project which includes the CRT+ and SPOA service. The CRT+ service received 23 referrals of which 16 (70%) were considered at risk of hospital admission. The service was able to support 100% admission avoidance at 48 hours and 83% at 7 days.<br>The SPOA service acted as a triage point to AHP rehabilitation services and 290 referrals were received during the period November 2020 to |
|  | end of March 2021. 314 onward referrals were made demonstrating the success of a SPOA to rehabilitation services.<br>No further evidence was found on these services. |
| Frail Elderly Short Stay Unit (FESSU)<br>*Winter pressure specific*<br>NHS Southport and Formby CCG. 2015<sup>35</sup><br>*Additional evidence*<br>Published evaluations<br>Michael, A. B., and F. Ijaola. 2014<sup>36</sup> (conference abstract)<br>Case studies | A conference abstract from 2014<sup>36</sup> details the outcomes of a 12 week pilot of a Frail Elderly Short Stay Unit. The average length of stay in the unit was 4.7 days and total length of stay (including subsequent inpatient care) was 6.5 days which had a positive impact on the Care of the Elderly wards.<br>Additional “winter funding” from central government in September 2013 enabled the establishment of a Frail Elderly Short Stay Unit (FESSU) in November 2013 as part of NHS Southport and Formby CCG their service developments to reduce admissions and length of stay for frail older people. |
| Ring-fenced arthroplasty unit<br>*Winter pressure specific*<br>Srikumar, B.et al 2021<sup>37</sup> (conference abstract)<br>*Additional evidence* | A conference abstract detailed the assessment of a ring-fenced arthroplasty unit performance during winter pressures and the pandemic (December-March 2018-19 and 2019-20)<sup>37</sup>. The unit performed 280 total hip and knee replacements in 2019-20 and the patients had a mean length of stay of 43 hours. This showed a reduction when compared with the length of stay in 2018-19 of 69 hours when 288 total hip and knee replacements were performed. Early discharge, defined as within 36 hours of surgery, occurred for 74% of patients in 2019-20 versus 24% in 2018-19 which add up to 333 inpatient days saved.<br>No further evidence was found on |
|  | ring-fenced arthroplasty units. |
| Speciality review at Rapid Access Clinics<br>*Winter pressure specific*<br>Edinburgh Integration Joint Board. 2021<sup>34</sup><br>*Additional evidence* | The Edinburgh Integration Joint Boards evaluation of winter planning 2020/21 details how NHS Lothian intended to use the additional funding for winter 2020/21 which including avoiding admission with services developed to provide certain initiatives including speciality review at rapid access clinics.<br>No further evidence was found on speciality review at rapid access clinics. |
| **Research Gaps** |  |
| Research in specialist units other than acute medical units could be helpful. |  |
| **Research Priority:** Acute medical units Low - Good quality Systematic Review(s) and plentiful UK research studies (No Gap)<br>Other High - Few UK initiatives (Implementation and Research Gap) |  |

In summary, acute medical units have a good evidence base overall, with some research specific to winter pressure conditions. Frail Elderly Short Stay Units have been supported by winter pressure funding but we found limited research on these and other types of specialist unit.

#### Structural Interventions – Facilitated discharge

Facilitated discharge is a key area for health and care services dealing with winter pressures. It includes interventions to optimise use of resources and minimise length of stay for patients in hospital, planning for discharge and discharge with support in place prior to a full assessment of health and care needs (discharge to assess).

##### S – Bed management

**Table 8.**
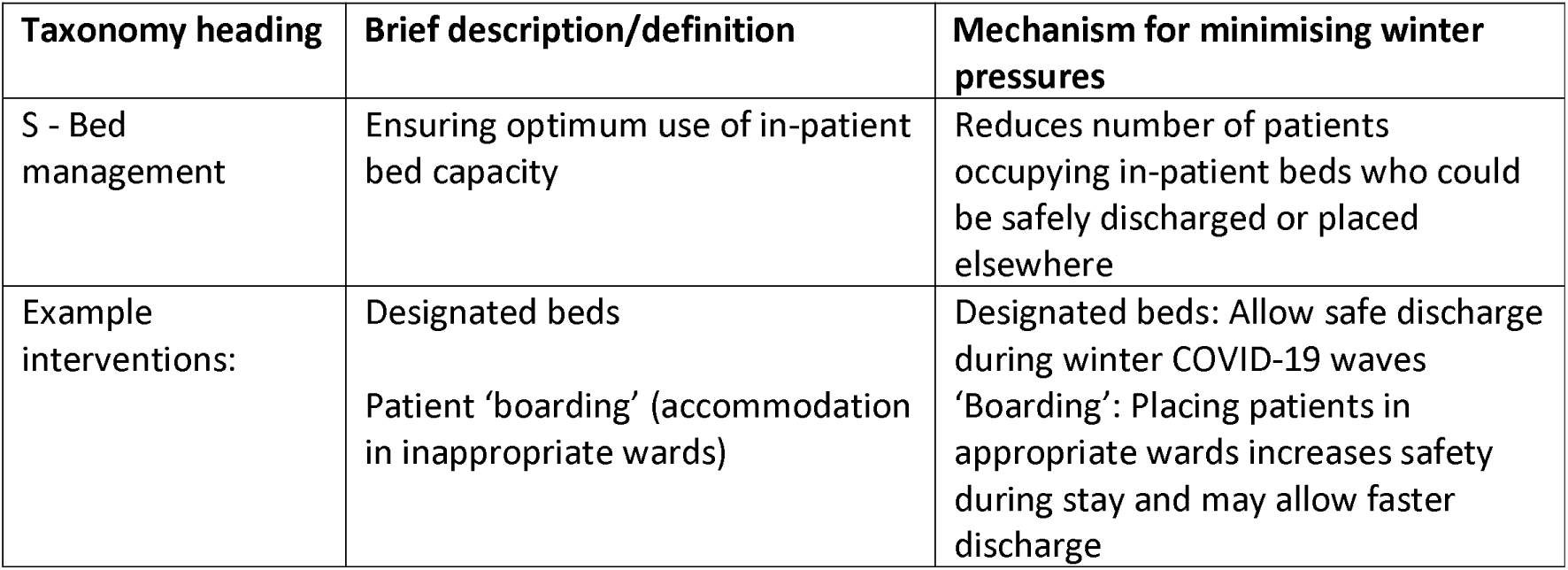
Bed Management: Definitions and Rationales.

**Table 9.** Bed Management: Interventions and Supporting Evidence.

| Supporting evidence | Results/Findings/Recommendations (from Abstract) |
| --- | --- |
| *Winter pressure specific*<br>Care Quality Commission 2021 (designated beds)<sup>38</sup><br>Peddie 2018 (patient 'boarding')<sup>30</sup> | February 2021: Over 150 'designated settings' within 100 local authorities<br>'Boarded' patients had shorter length of stay (perhaps selected because short stay expected) but care perceived to be of lower standard<br>Authors note that 'NHS Greater Glasgow and Clyde have reduced boarding by 70% since 2007 through focused discharge planning' (Beckett Daniel. Winter pressures in NHS Scotland 2008-2009: report for the Emergency Access Delivery team, Scottish Government Daniel Beckett. [2017-03-20]. http://www.shiftingthebalance.scot.nhs.uk/downloads/1251120568-Winter%20Pressures%20Report%20-%20final%20-%20rich%20text%20format.pdf) |
| *Systematic review*<br>Tampubolon, L (2018)<sup>39</sup> | Changing policies for hospital bed management are worth exploring to improve hospital patient flow and LOS. Downstream bed management is only one of the factors for potentially improving the overall organization function. Other factors include the need to reorganize channels upstream and strengthen the internal organization of emergency departments (EDs) |

| Research Gaps |
| --- |
| Systematic review included only five studies and poor quality (no quality/risk of bias assessment). Additional body of research based on mathematical modelling approaches |
| **Research Priority:** High - Few UK initiatives (Implementation and Research Gap) with caveat that interventions may be described as discharge planning/co-ordination |

##### S – Discharge co-ordinators

**Table 10.** Discharge co-ordinators: Definitions and Rationales.

| Taxonomy heading | Brief description/definition | Mechanism for minimising winter pressures |
| --- | --- | --- |
| S – Discharge co-ordinators | Individual with designated responsibility for discharge planning. Early discharge requires robust systems to be in place to develop plans for management and discharge, and to allow an expected date of discharge to be set within 48 hours (LGA 2018)<sup>40</sup> | Contributes to various forms of facilitated/supported discharge. Could be a temporary role or given extra support to cover winter pressures. |
| Example interventions: Often part of complex MDT interventions, e.g. Hospital at home (cited in Williams 2022)<sup>41</sup>; Oxfordshire Integrated Liaison Hub (LGA 2018)<sup>40</sup> | Integrated liaison hub staffed by MDT including qualified nurses and therapists with expertise in discharge planning, discharge planners, administrators and staff from adult social care (in-reaching). | Reduces delayed transfers of care and hence frees up hospital beds for new patients |

**Table 11.**
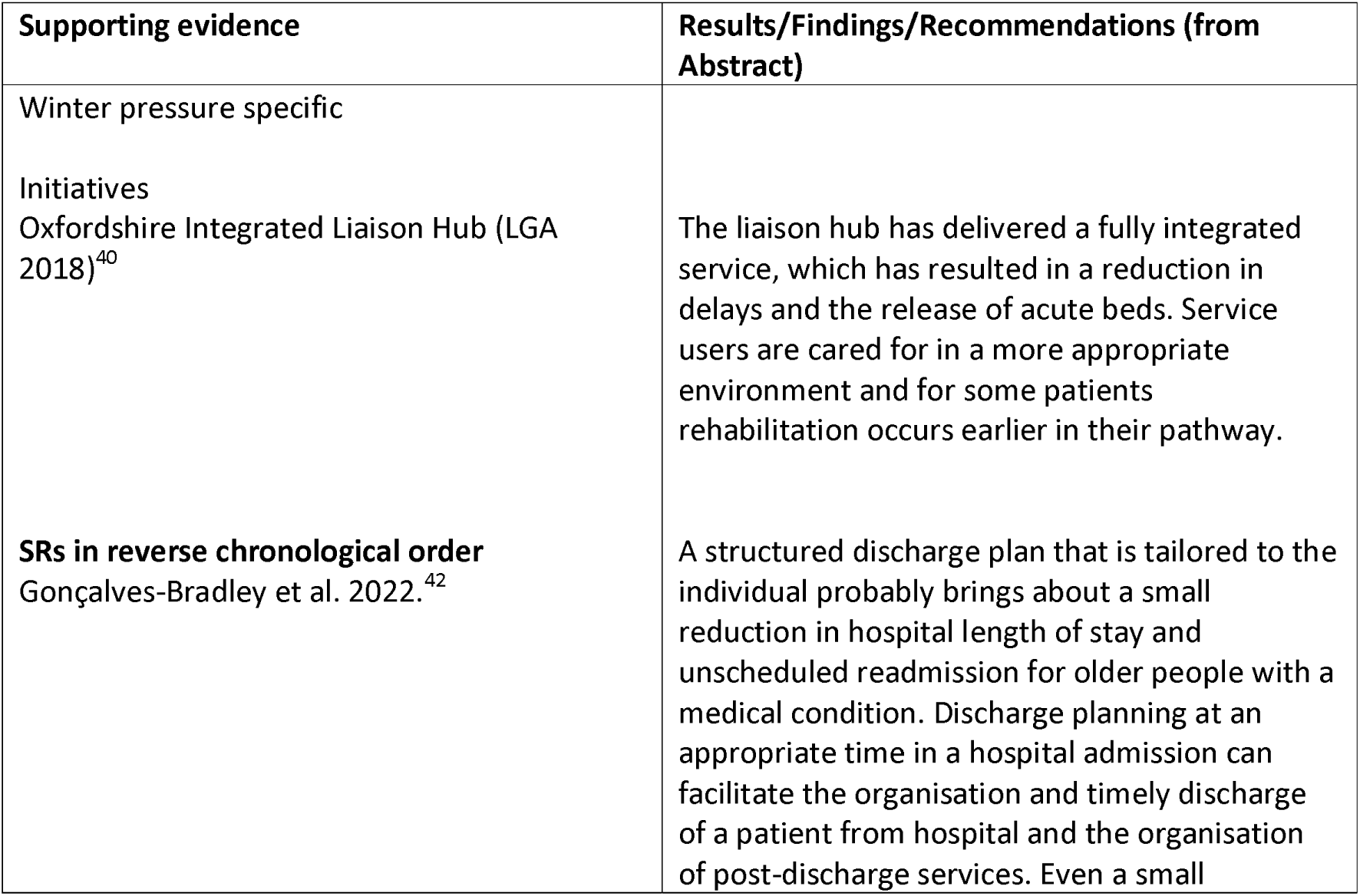
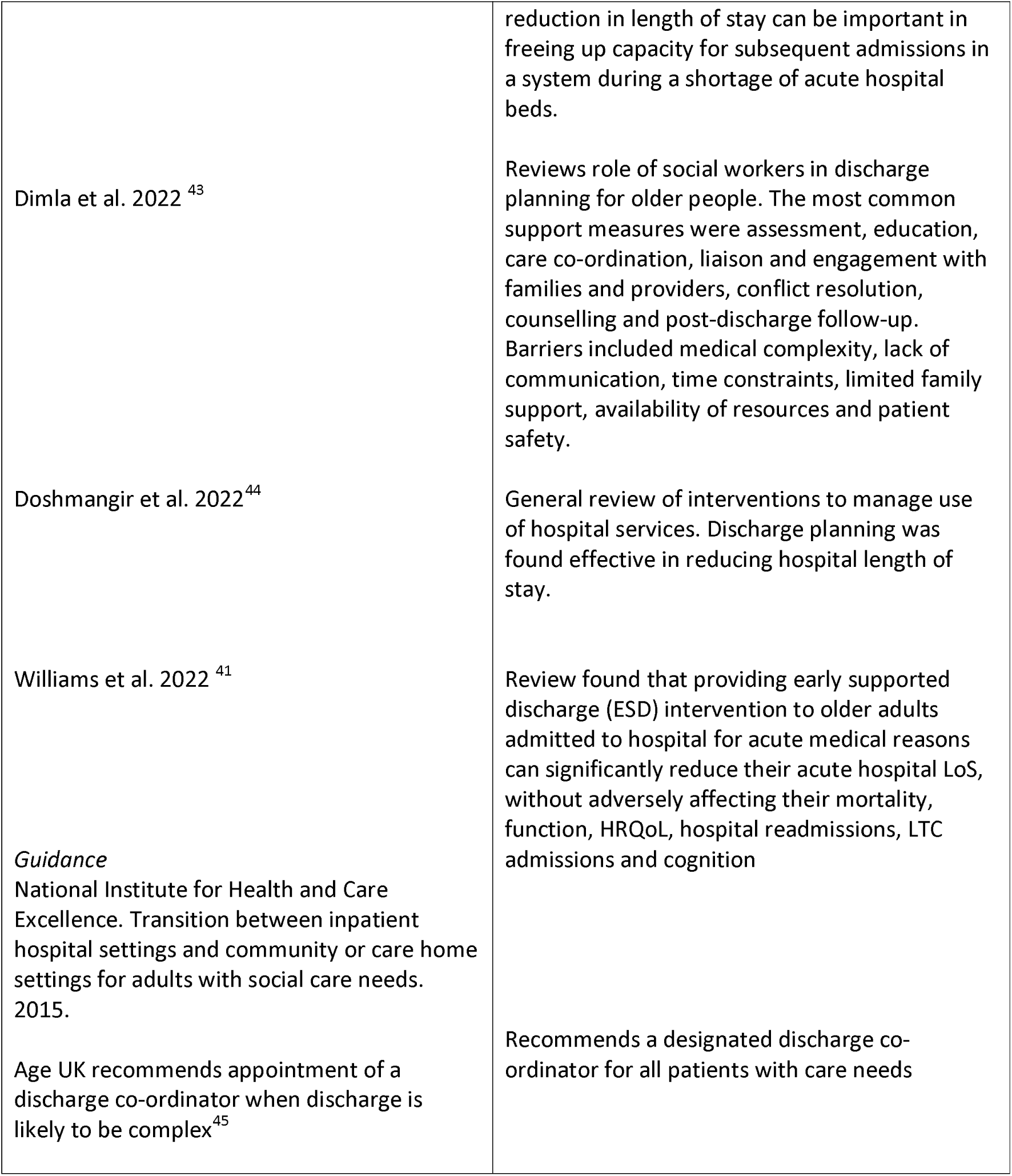
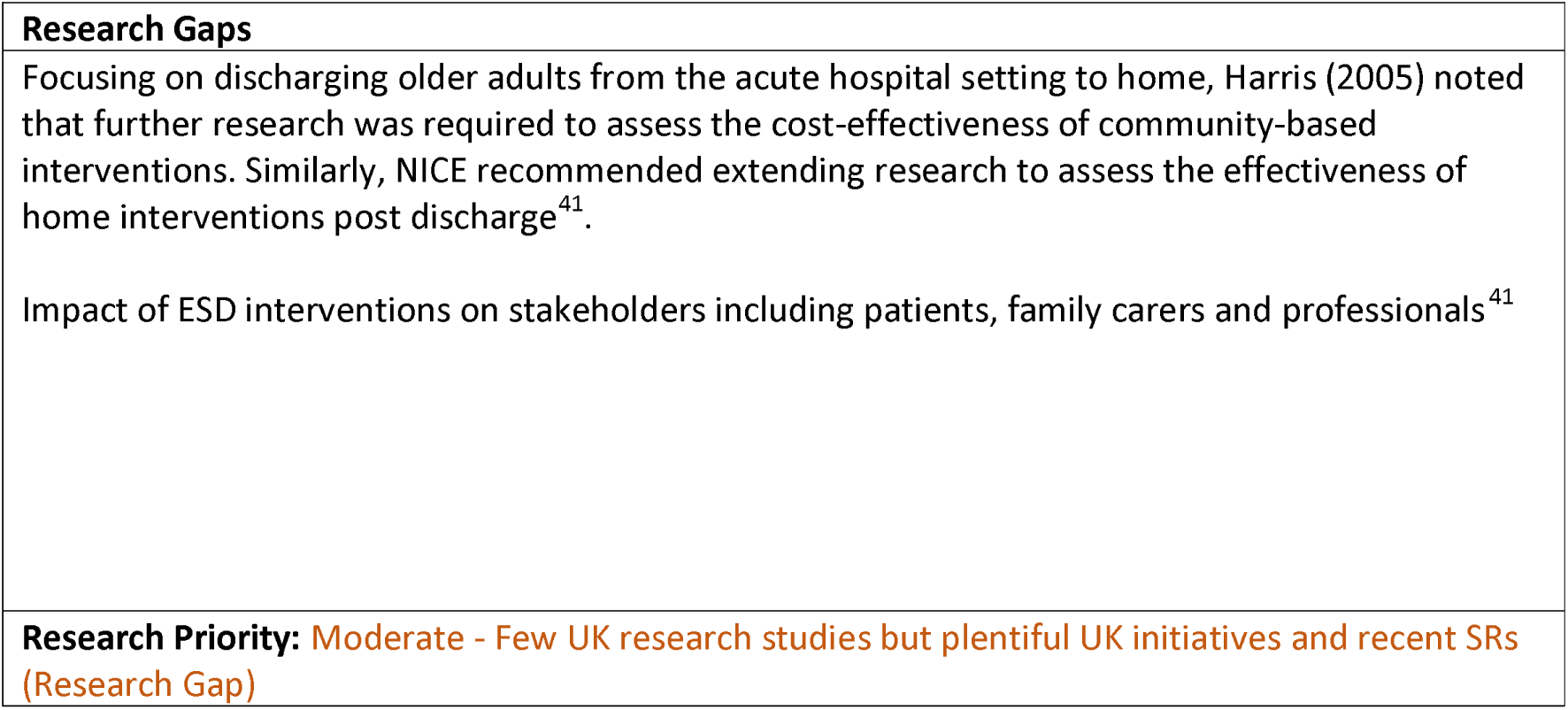
Discharge co-ordinators: Interventions and Supporting Evidence.

##### S – Discharge to assess

**Table 12.**
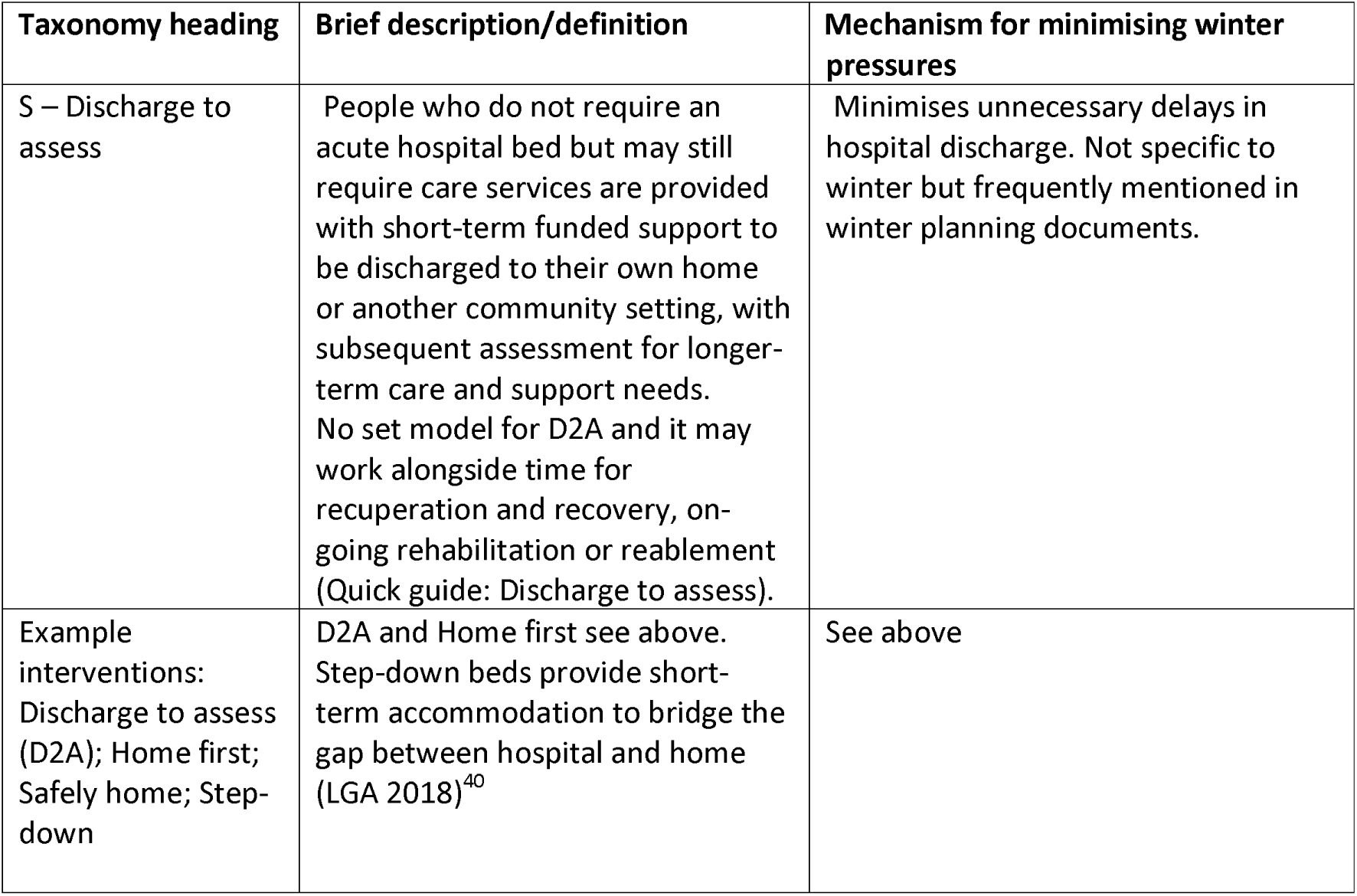
Discharge to assess: Definitions and Rationales.

**Table 13.**
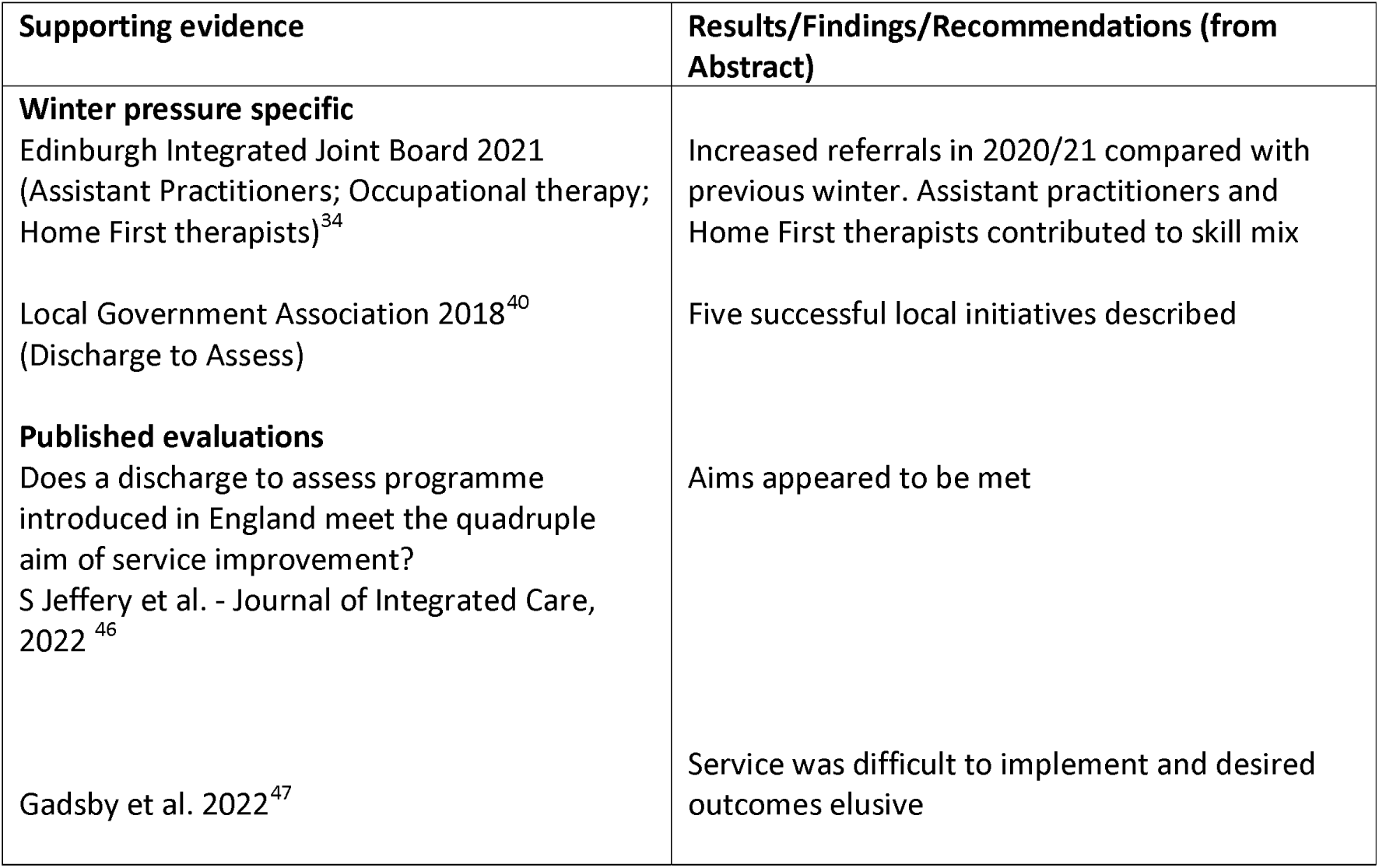
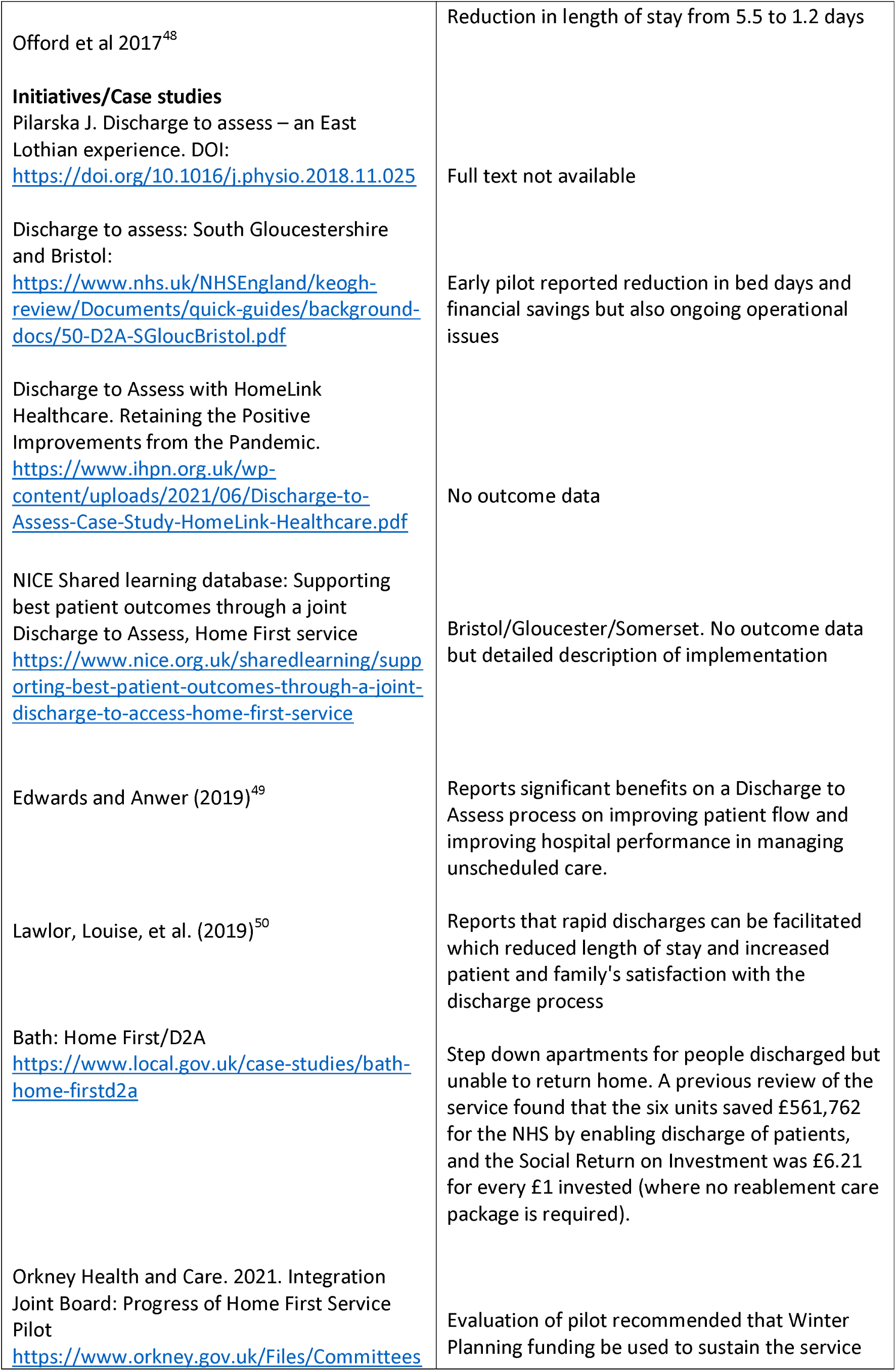
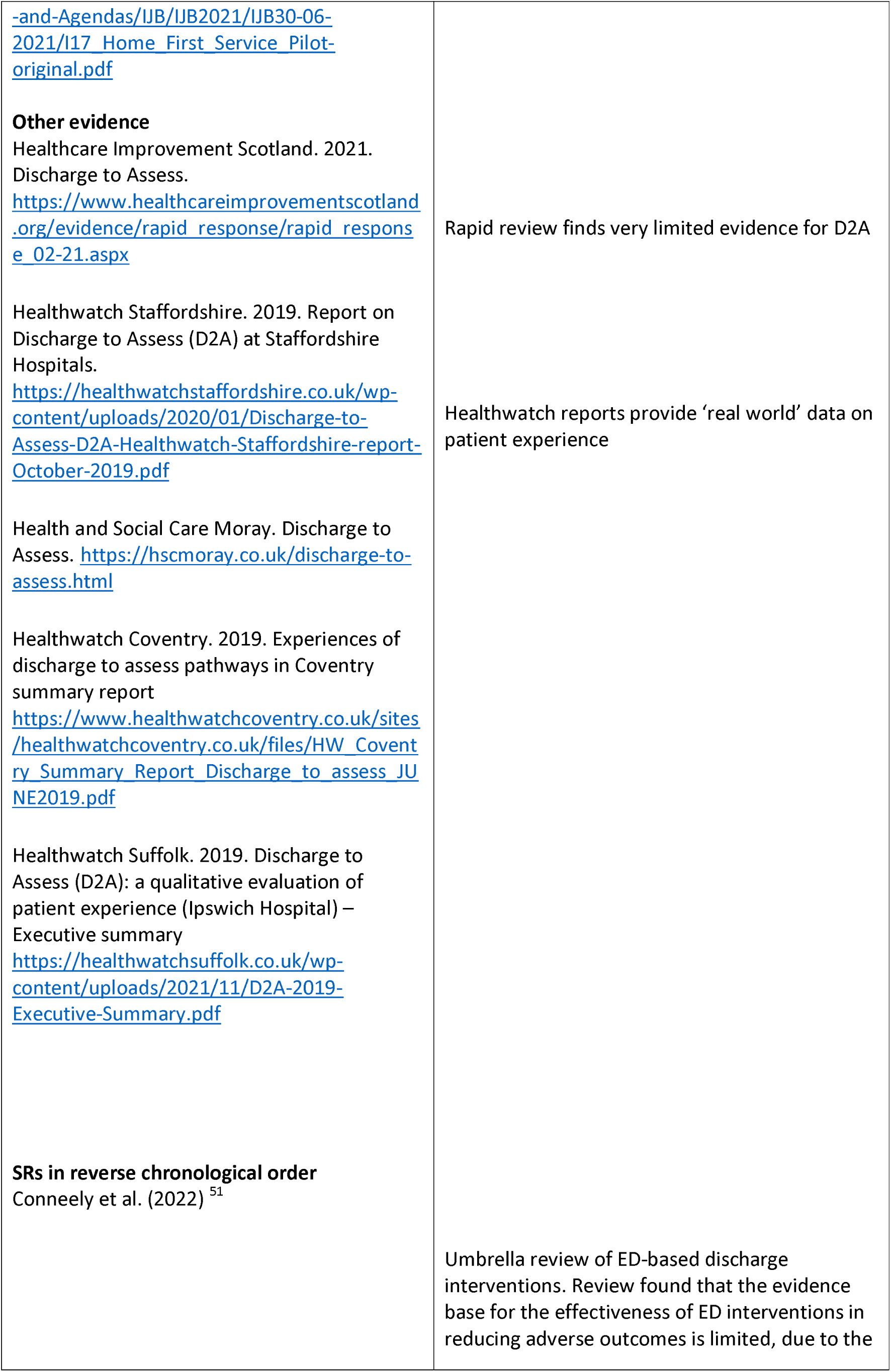
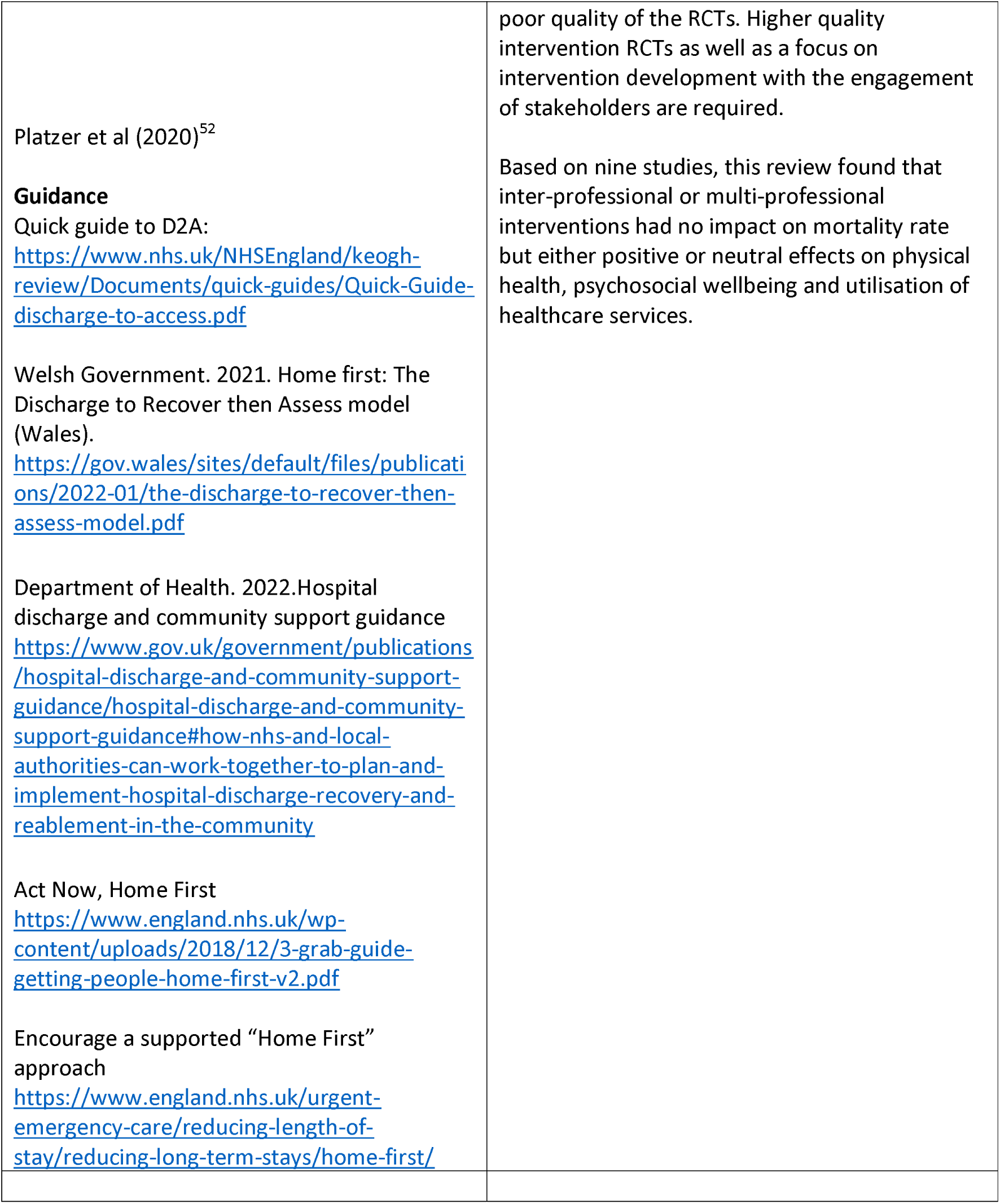
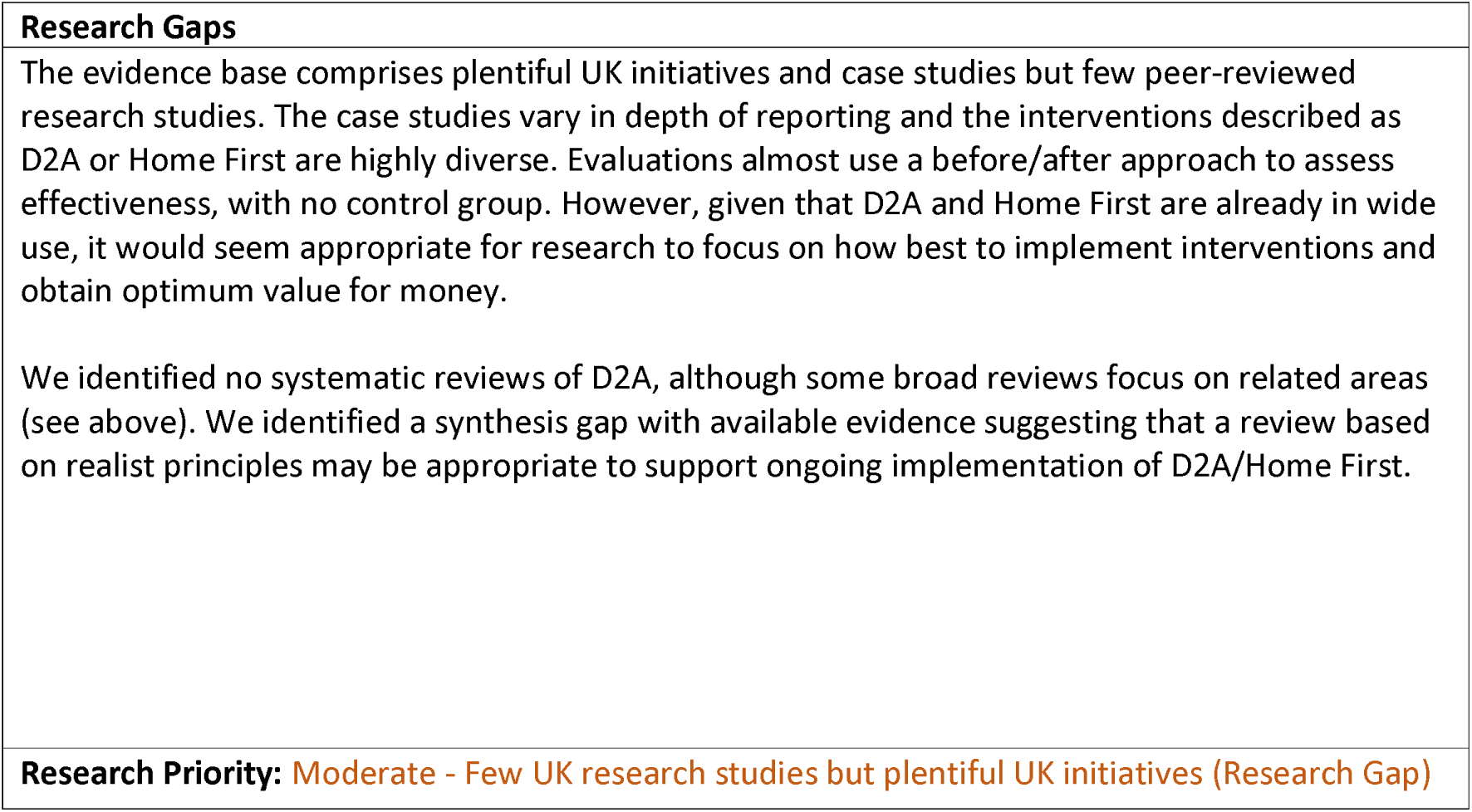
Discharge to assess: Interventions and Supporting Evidence.

##### S – Monitoring and Review

**Table 14.** Monitoring and Review: Definitions and Rationales.

| **Taxonomy heading** | **Brief description/definition** | **Mechanism for minimising winter pressures** |
| --- | --- | --- |
| S – Monitoring and review | Monitoring and review interventions aim to identify patients suitable for discharge or transfer from more to less intensive levels of support. Involvement of professionals from different disciplines supports a holistic assessment of the patient's needs. | Early review (e.g. 7 days/week) can identify patients who would benefit from transfer to a less intensive community setting and reduce pressure on the most specialist services. Closely related to 'patient flow' and 'prioritisation and triage'. Monitoring outside hospital is covered under 'telecare'. |
| Example interventions: Daily multi-disciplinary review | See Goodison (2021)<sup>53</sup> for application in a hyper-acute stroke unit | See above |

**Table 15.**
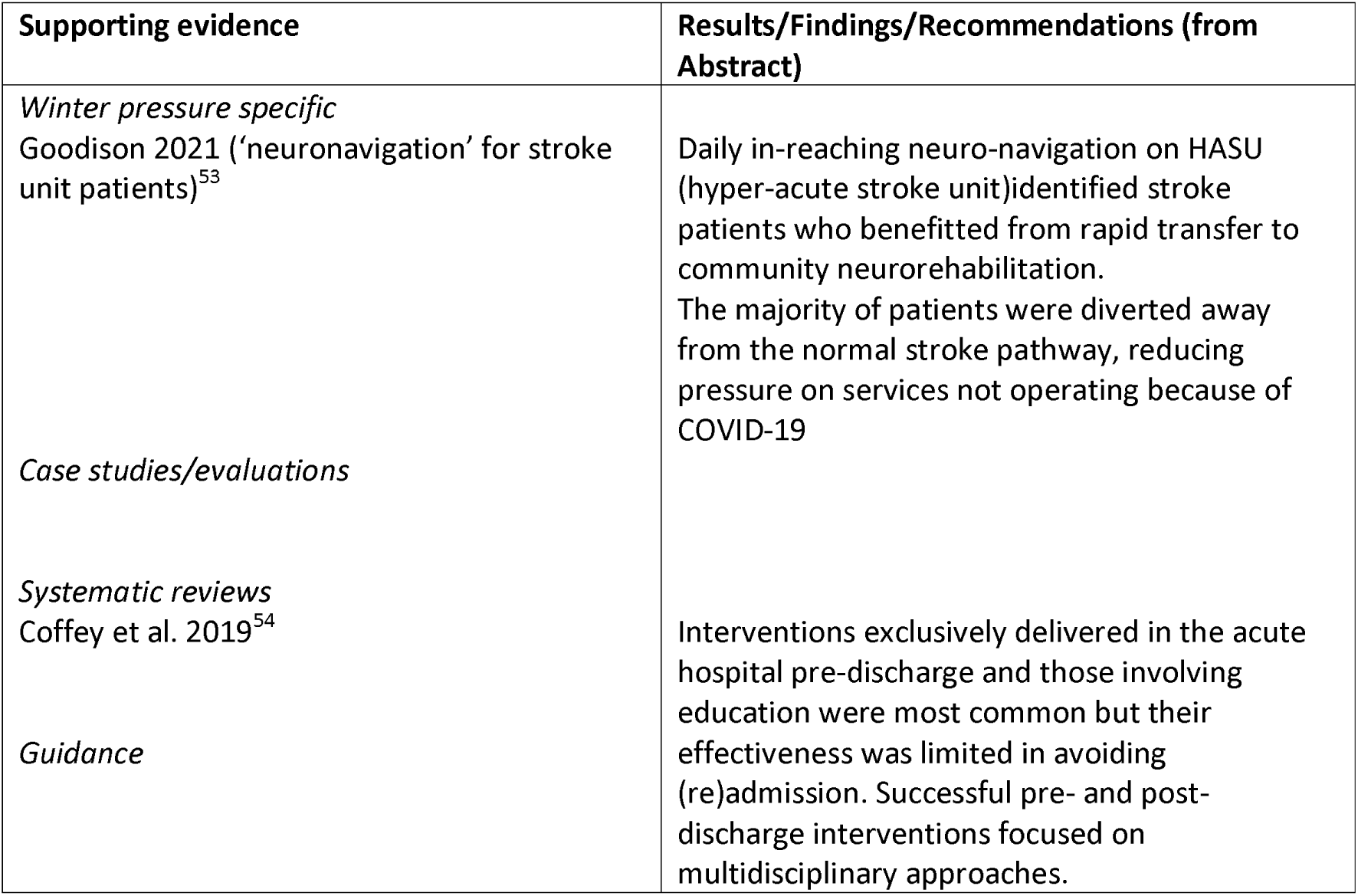
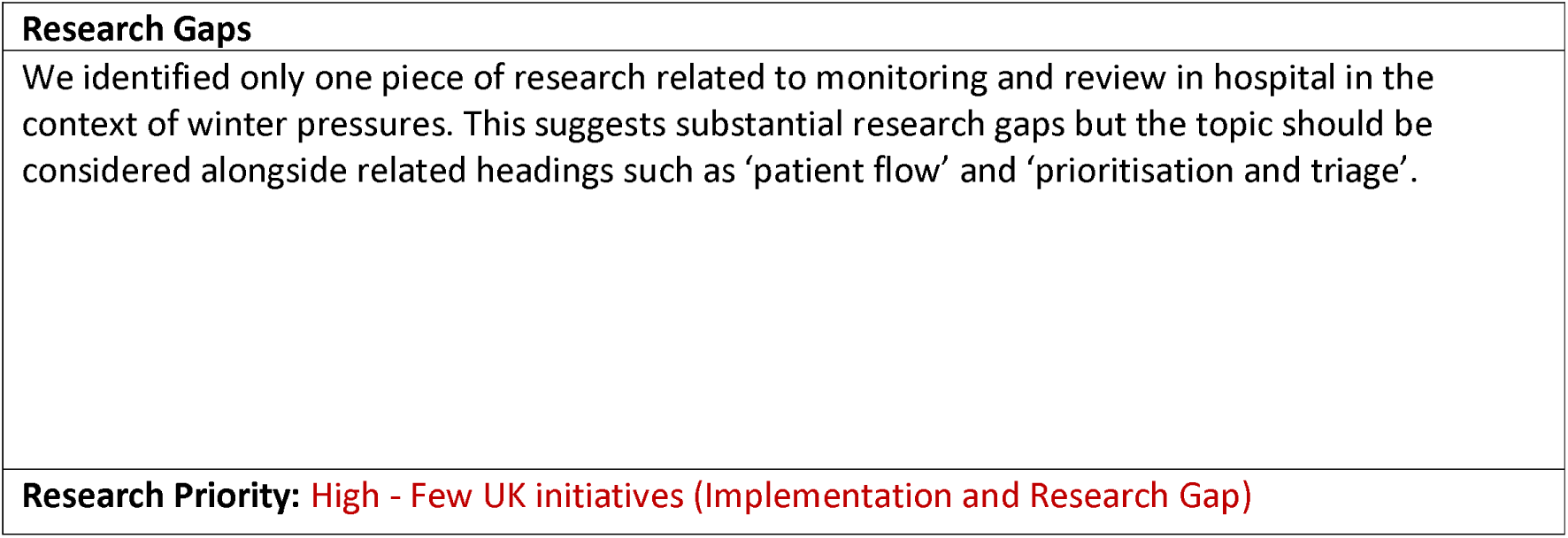
Monitoring and review: Interventions and Supporting Evidence.

| **Supporting evidence** | **Results/Findings/Recommendations (from Abstract)** |
| --- | --- |
| *Winter pressure specific*<br>Goodison 2021 ('neuronavigation' for stroke unit patients)<sup>53</sup><br>*Case studies/evaluations*<br>*Systematic reviews*<br>Coffey et al. 2019<sup>54</sup><br>*Guidance* | Daily in-reaching neuro-navigation on HASU (hyper-acute stroke unit) identified stroke patients who benefitted from rapid transfer to community neurorehabilitation. The majority of patients were diverted away from the normal stroke pathway, reducing pressure on services not operating because of COVID-19<br>Interventions exclusively delivered in the acute hospital pre-discharge and those involving education were most common but their effectiveness was limited in avoiding (re)admission. Successful pre- and post-discharge interventions focused on multidisciplinary approaches. |

|  |
| --- |
| **Research Gaps** |
| We identified only one piece of research related to monitoring and review in hospital in the context of winter pressures. This suggests substantial research gaps but the topic should be considered alongside related headings such as 'patient flow' and 'prioritisation and triage'. |
| **Research Priority:** High - Few UK initiatives (Implementation and Research Gap) |

##### S – Patient flow

**Table 16.** Patient flow: Definitions and Rationales.

| **Taxonomy heading** | **Brief description/definition** | **Mechanism for minimising winter pressures** |
| --- | --- | --- |
| S – Patient flow | Covers a range of interventions to ensure patients move through the health and care system as quickly and efficiently as possible while maintaining safety and patient focus | At periods of high demand, it is particularly important to minimise unnecessary contacts and use telephone or video appointments if appropriate to avoid patients choosing to attend the ED. Closely related to 'Monitoring and review' and 'Prioritisation and triage'. |
| Example interventions:<br>Patient flow bundle; Flow Navigation Centres | Flow Navigation Centres (hubs) receive referrals directly from NHS24 (Scotland). They offer rapid access to a senior clinical decision maker within the multidisciplinary team, via a telephone or video consultation where possible<sup>55</sup>.<br>The SAFER patient flow bundle is applicable to in-patient wards (excluding maternity). In highly specialist settings, daily review can identify patients who would benefit from transfer to a lower-intensity community setting<sup>10</sup>. | Optimising patient flow at the point of entry into the system (pre-hospital)<br>Optimising patient flow within hospitals |

**Table 17.**
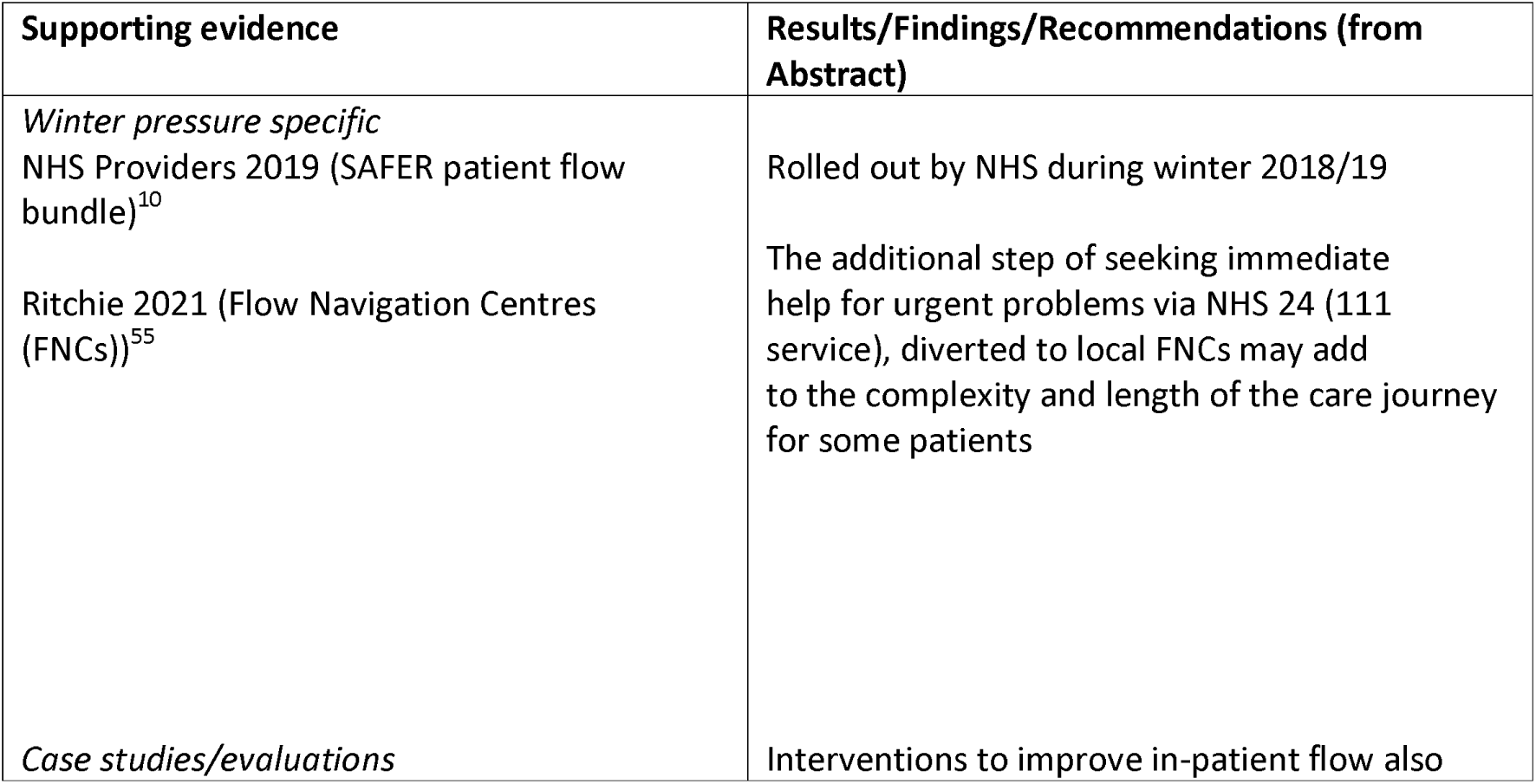
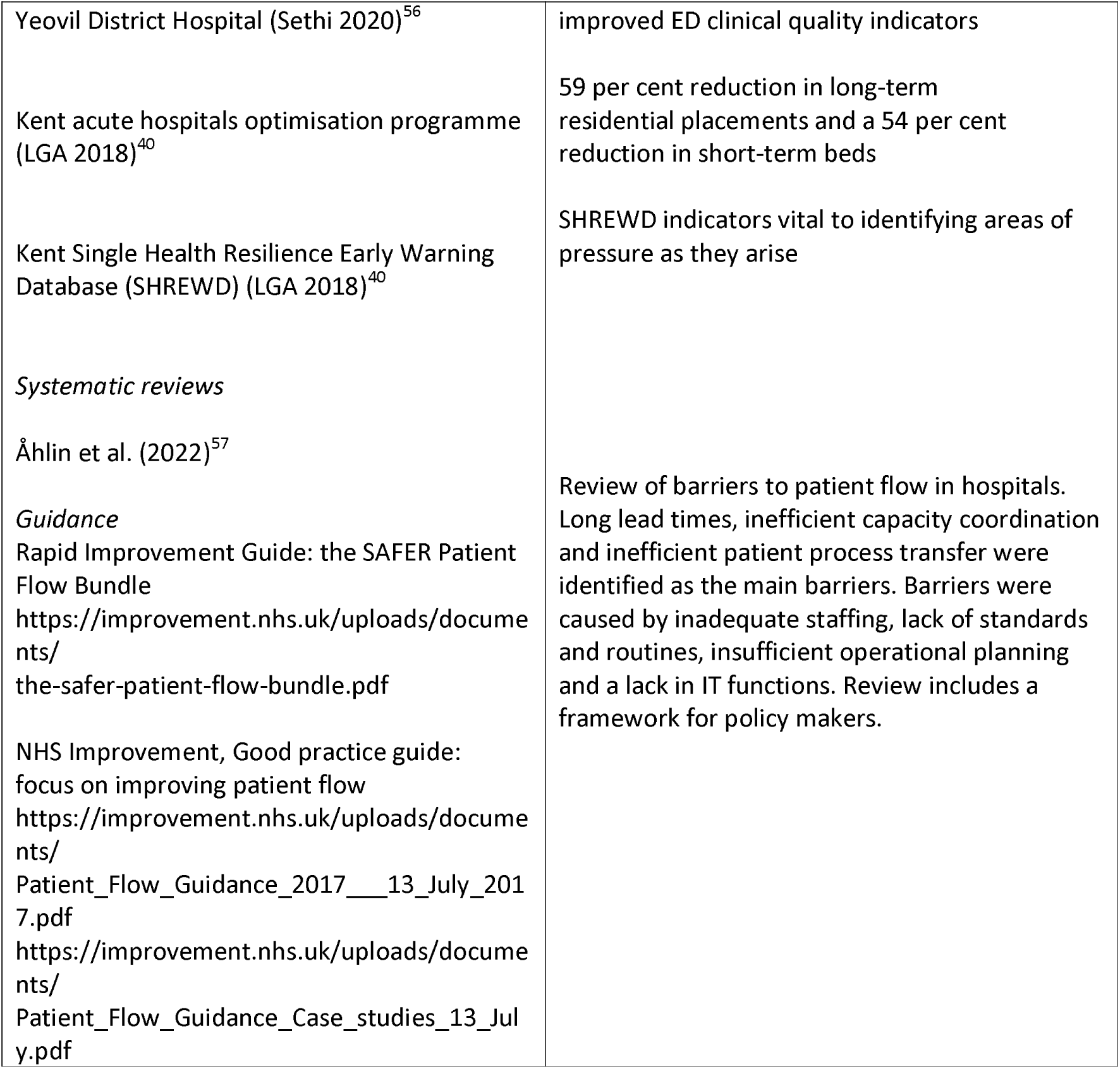
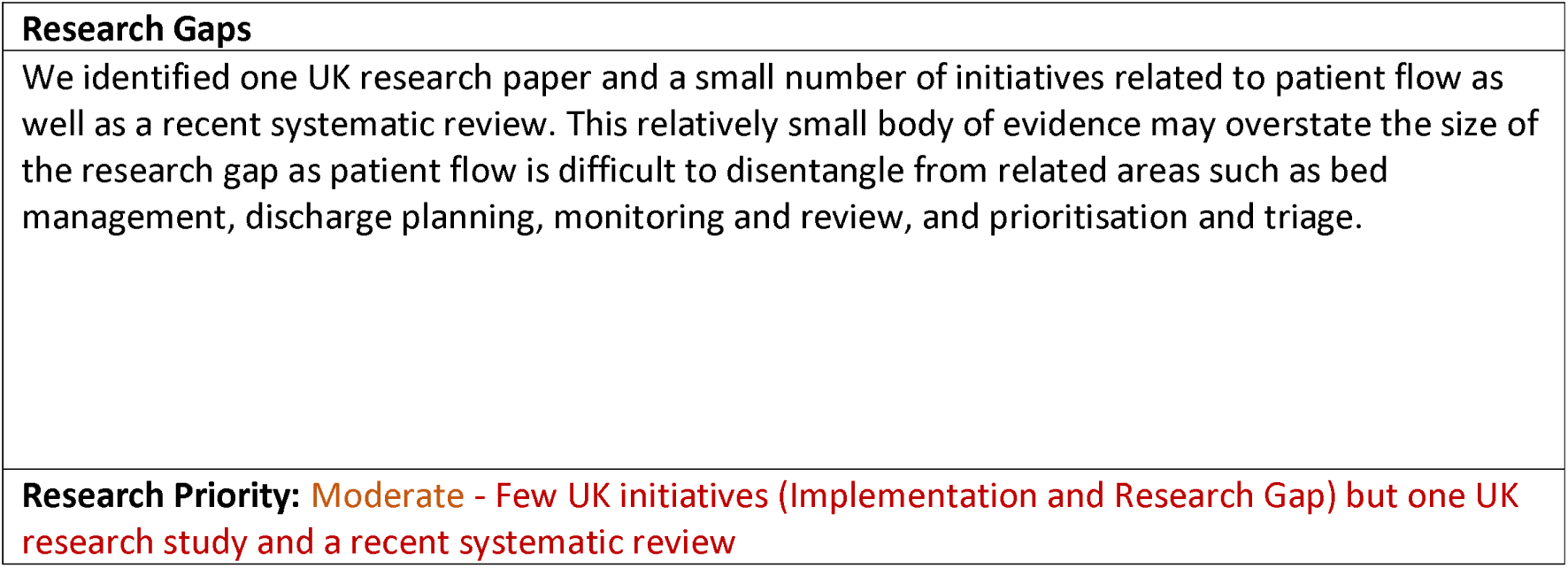
Patient flow: Interventions and Supporting Evidence.

We found that some aspects of facilitated discharge are relatively well researched, particularly discharge to assess and similar interventions (e.g. home first). Interventions with little UK evidence were sometimes supported by systematic reviews, although the quality of these varied. Some taxonomy headings, e.g. ‘bed management’ and ‘discharge co-ordinators’, were often evaluated as part of a broader process of ‘discharge planning’.

#### Structural Interventions – Cross-cutting

This section of the taxonomy covers a diverse group of structural changes with broad applicability, including multidisciplinary and multi-agency teams, use of digital technology and data, monitoring and accountability in the healthcare system, national and local policies, staff redeployment and the role of volunteers.

##### S – Communication and teamwork

**Table 18.** Communication and Teamwork: Definitions and Rationales.

| **Taxonomy heading** | **Brief description/definition** | **Mechanism for minimising winter pressures** |
| --- | --- | --- |
| S – Communication and teamwork | Tackles barriers at the interface between disciplines and or agencies (e.g. NHS and local government, hospitals and care homes) | Such barriers may become more pronounced as a defensive reaction to periods of exceptional demand/pressure on staff. |
| Example interventions:<br>Use of advice and guidance | Generalists and specialists working together to avoid admission | Unnecessary hospital admissions are avoided |
| Multidisciplinary/multi-agency discharge teams | Coordinated discharge planning based on joint assessment processes and protocols and on shared and agreed responsibilities | More efficient discharge process |

**Table 19.**
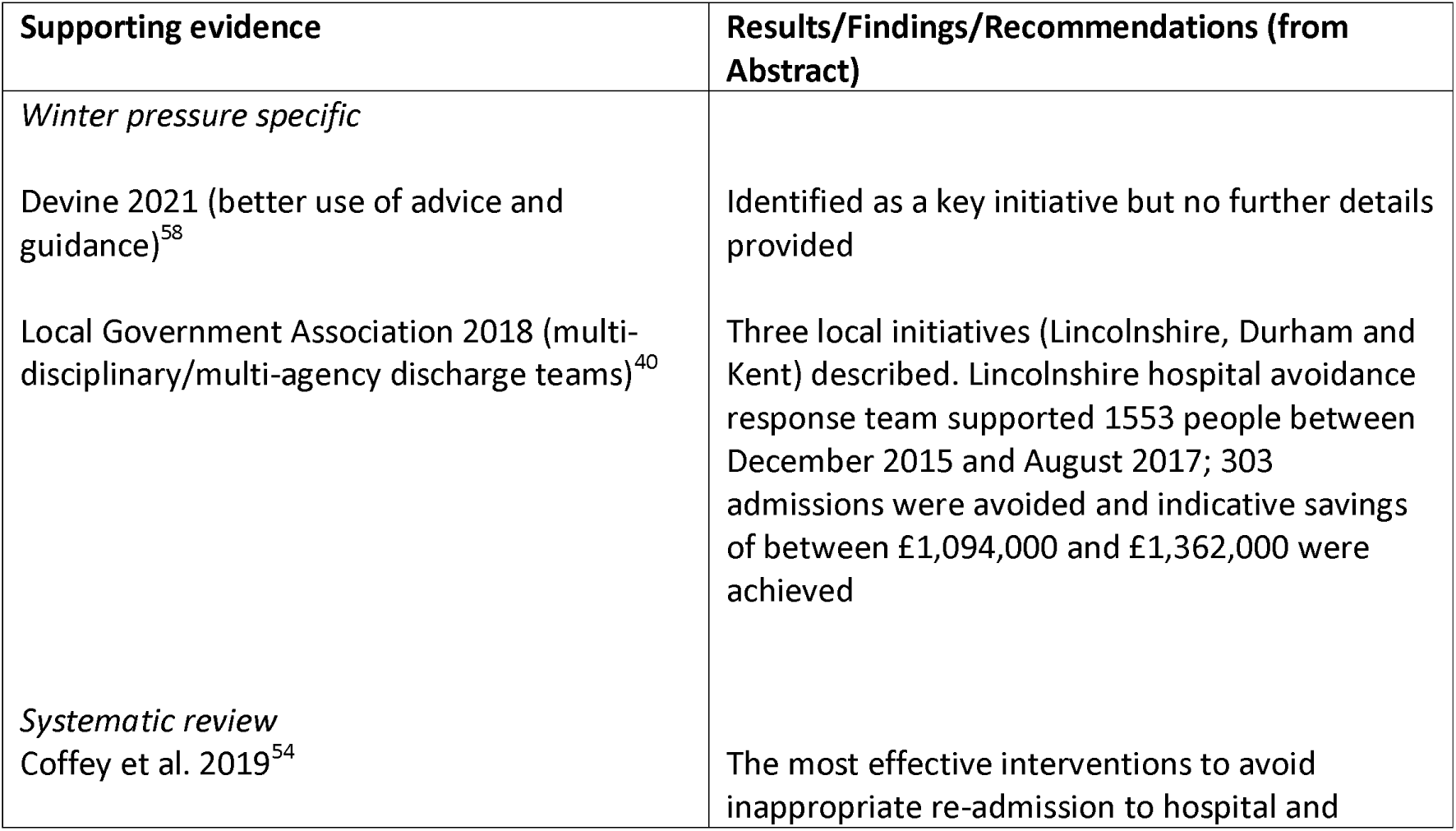
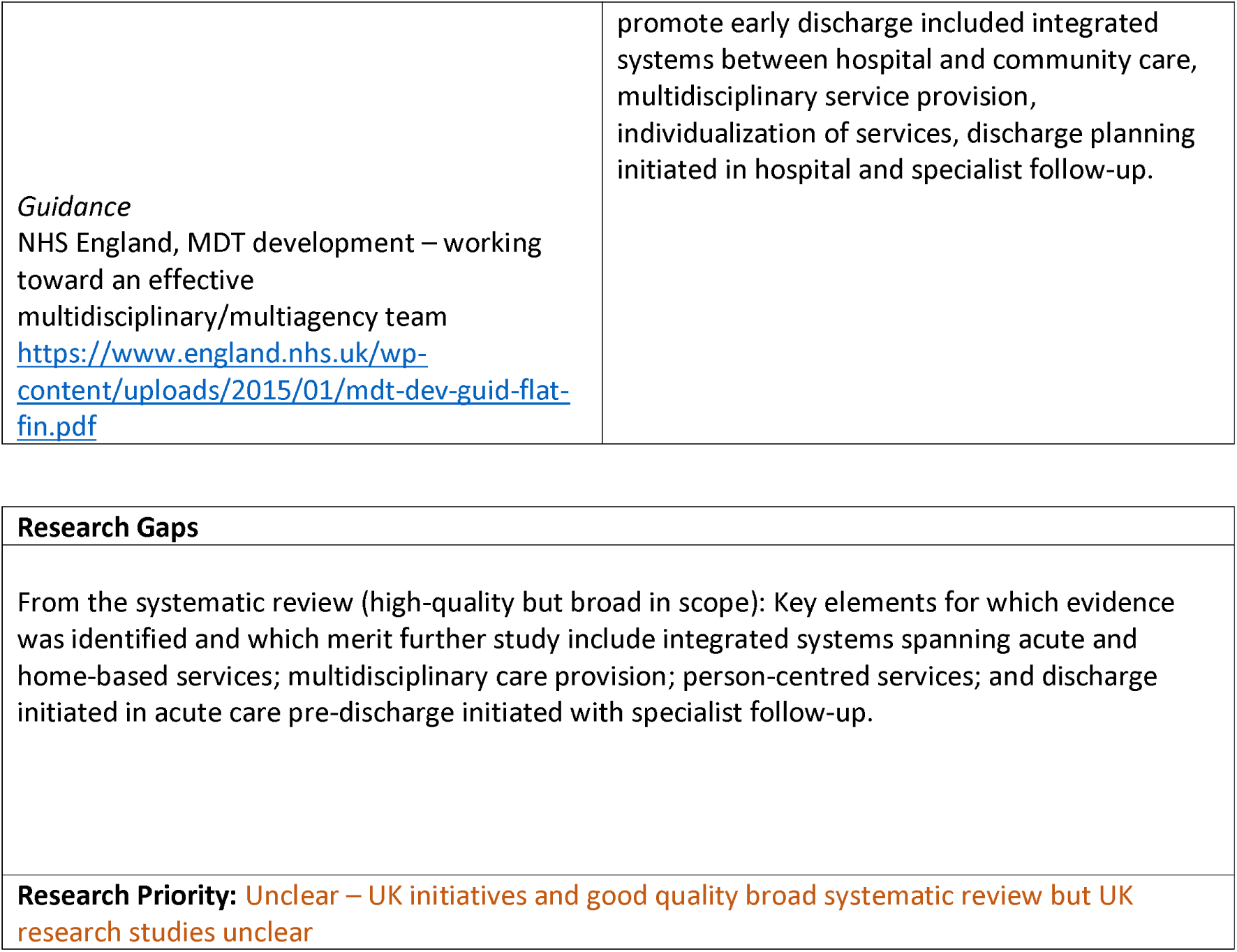
Communication and teamwork: Interventions and Supporting Evidence.

| **Supporting evidence** | **Results/Findings/Recommendations (from Abstract)** |
| --- | --- |
| *Winter pressure specific* |  |
| Devine 2021 (better use of advice and guidance)<sup>58</sup> | Identified as a key initiative but no further details provided |
| Local Government Association 2018 (multi-disciplinary/multi-agency discharge teams)<sup>40</sup> | Three local initiatives (Lincolnshire, Durham and Kent) described. Lincolnshire hospital avoidance response team supported 1553 people between December 2015 and August 2017; 303 admissions were avoided and indicative savings of between £1,094,000 and £1,362,000 were achieved |
| *Systematic review*<br>Coffey et al. 2019<sup>54</sup> | The most effective interventions to avoid inappropriate re-admission to hospital and |
| *Guidance*<br>NHS England, MDT development – working toward an effective multidisciplinary/multiagency team<br>https://www.england.nhs.uk/wp-content/uploads/2015/01/mdt-dev-guid-flat-fin.pdf | promote early discharge included integrated systems between hospital and community care, multidisciplinary service provision, individualization of services, discharge planning initiated in hospital and specialist follow-up. |

|  |
| --- |
| **Research Gaps**<br>From the systematic review (high-quality but broad in scope): Key elements for which evidence was identified and which merit further study include integrated systems spanning acute and home-based services; multidisciplinary care provision; person-centred services; and discharge initiated in acute care pre-discharge initiated with specialist follow-up. |
| **Research Priority:** Unclear – UK initiatives and good quality broad systematic review but UK research studies unclear |

##### S – Digital and data

**Table 20.**
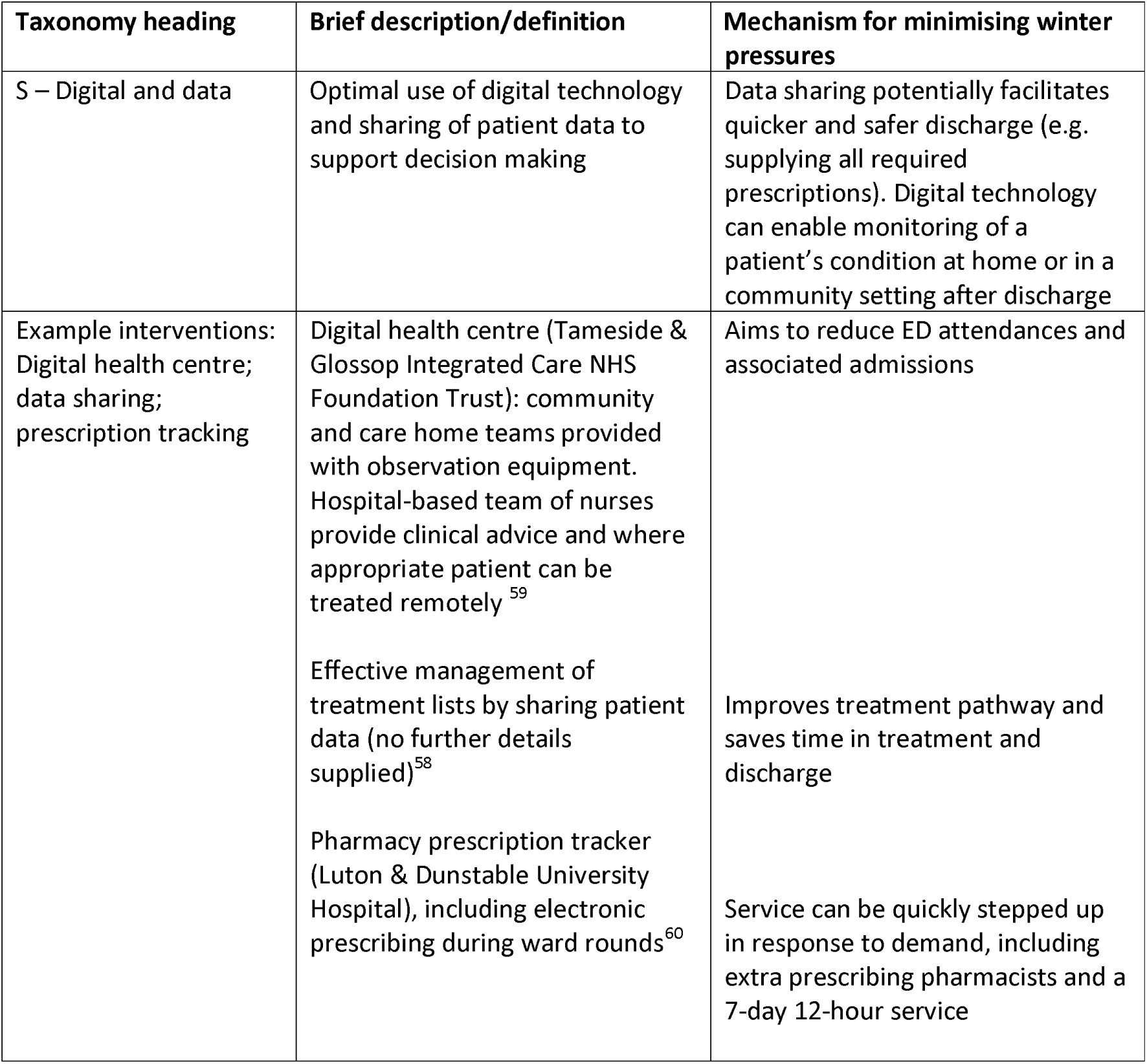
Digital and data: Definitions and Rationales.

**Table 21.**
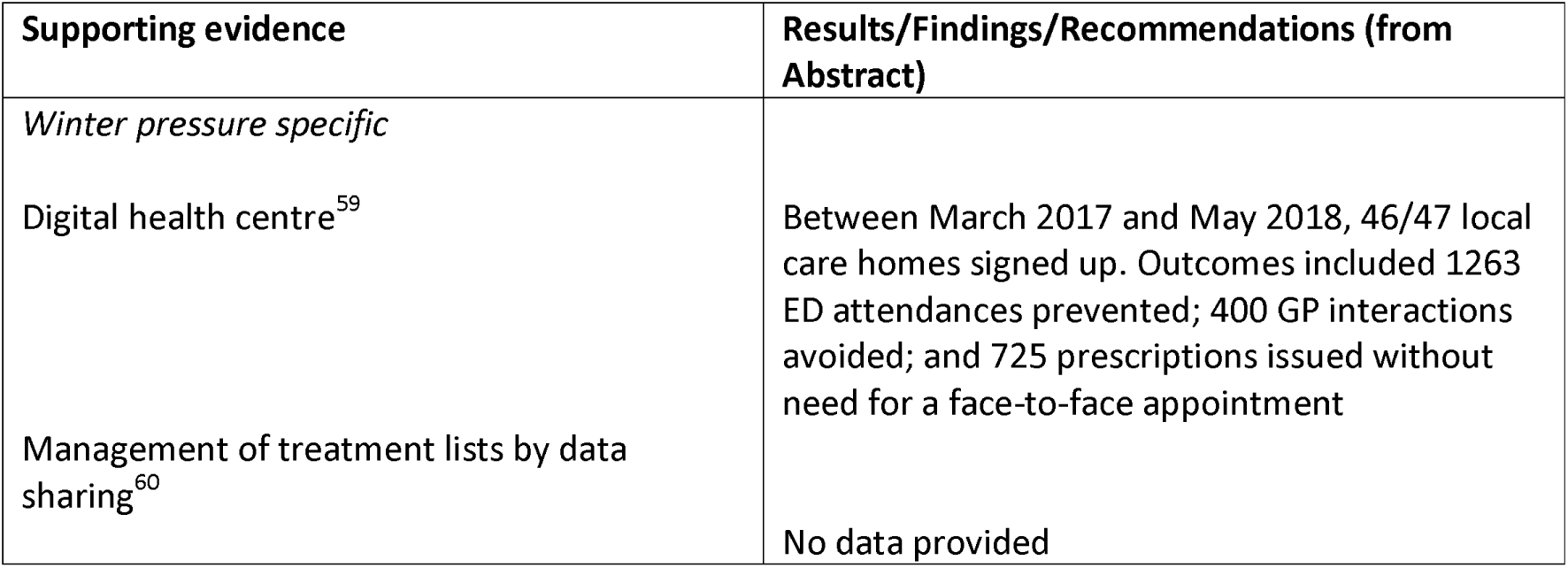
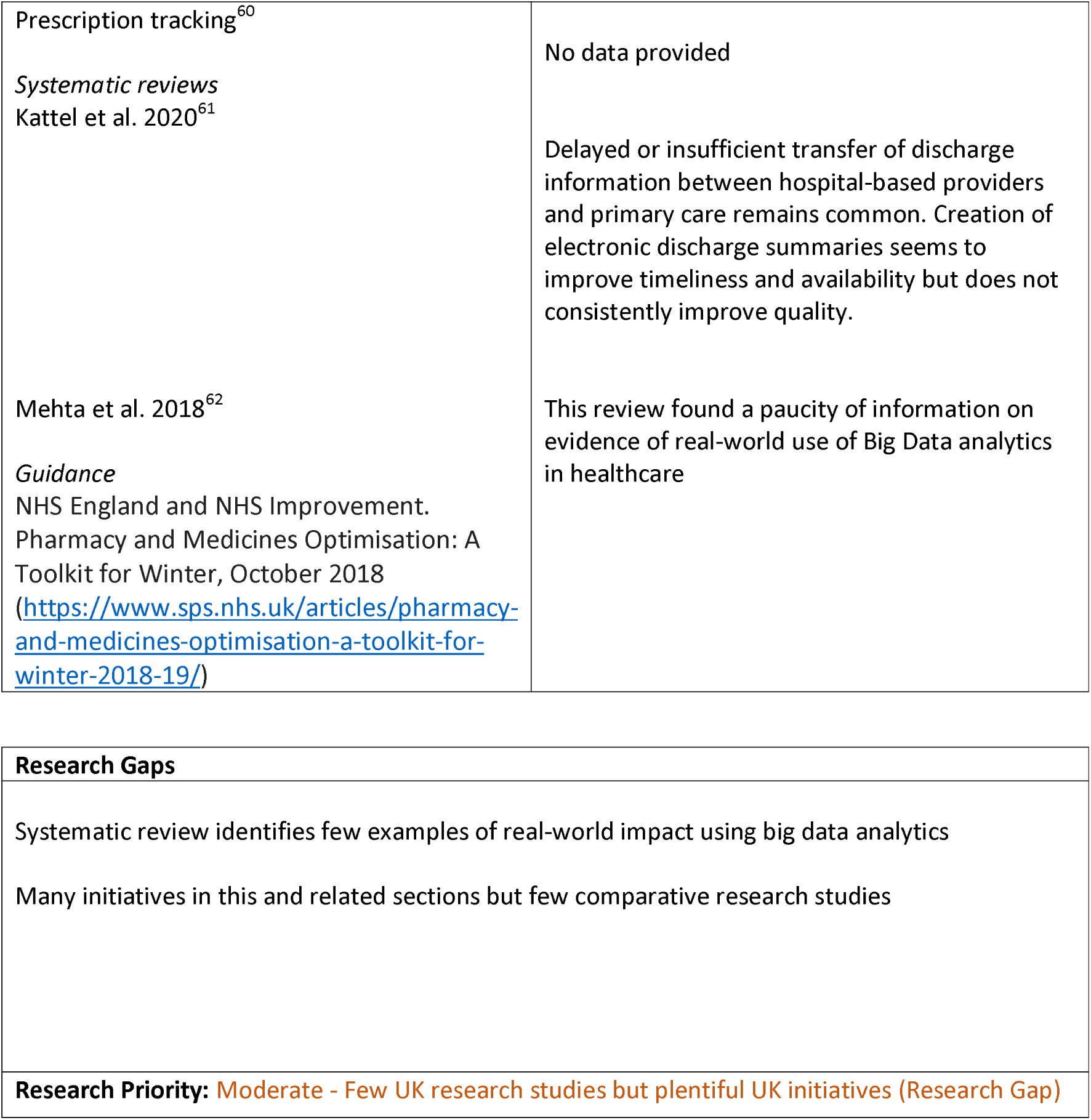
Digital and data: Interventions and supporting evidence.

##### S – Governance

**Table 22.** Governance: Definitions and Rationales.

| Taxonomy heading | Brief description/definition | Mechanism for minimising winter pressures |
| --- | --- | --- |
| S – Governance | NHS England defines clinical governance as ‘a system through which NHS organisations are accountable for continuously improving the quality of their services and safeguarding high standards of care by creating an environment in which excellence in clinical care will flourish’. It ‘involves monitoring systems and processes to provide assurance of patient safety and quality of care across the organisation’.<br>(https://www.england.nhs.uk/mat-transformation/matrons-handbook/governance-patient-safety-and-quality/) | Monitoring systems help to identify rising pressure on services and governance seeks to ensure that appropriate processes are followed, with named individuals held accountable and feedback (i.e. audit and administrative data) on outcomes. |
| Example interventions: | Any changes to governance systems, normally at the hospital level |  |

**Table 23.** Governance: Interventions and Supporting Evidence.

| Supporting evidence | Results/Findings/Recommendations (from Abstract) |
| --- | --- |
| *Winter pressure specific*<br>*Case studies/evaluations*<br>*Systematic reviews*<br>Zonneveld et al. 2018<sup>63</sup><br>Values of integrated care | No supporting evidence identified<br>This review identified 23 underlying values that may drive behaviour, decision-making, collaboration and governance processes in integrated care networks. The most frequently identified values were ‘collaborative’, ‘co-ordinated’, ‘transparent’, ‘empowering’, ‘comprehensive’, ‘co-produced’ and ‘shared responsibility and accountability’. Authors suggest that values could serve as governance principles for the alignment between |
| Nicholson et al. 2013 (BMC Health Serv Res. 2013 Dec 20;13:528. doi: 10.1186/1472-6963-13-528.)<br>Governance framework for integrated primary/secondary care<br>*Guidance* | organisations in integrated care initiatives and partnerships, for example by influencing decision-making processes or leadership behaviour.<br>This review identified ten elements needed for integrated primary/secondary health care governance across a regional setting including: joint planning; integrated information communication technology; change management; shared clinical priorities; incentives; population focus; measurement – using data as a quality improvement tool; continuing professional development supporting joint working; patient/community engagement; and, innovation. All included studies (n = 21) evaluated the process of integrated governance and service delivery structures, rather than the effectiveness of services. Authors concluded that the elements listed above provide a focus for setting up integration initiatives, which need to be flexible for adapting to local conditions and settings. |

| **Research Gaps** |
| --- |
| We identified no UK research studies or initiatives related to governance. It is possible that the nature of the topic requires a more detailed and focused search to identify any relevant evidence. |
| **Research Priority:** High - Few UK initiatives (Implementation and Research Gap) |

##### S - Managed Care Approaches

**Table 24.** Managed Care: Definitions and Rationales.

| Taxonomy heading | Brief description/definition | Mechanism for minimising winter pressures |
| --- | --- | --- |
| S – Managed care approaches | Managed care involves purchasers of health care (e.g. insurance companies) using their bargaining power to control prices and influence how health and care services are provided.<br>Managed care is primarily a feature of insurance-funded health systems (particularly the USA) and evidence of its use to address winter pressures is unlikely to be relevant to the UK setting. | The rationale for managed care approaches is based on the role of financial incentives in improving quality of care, e.g. financial penalties for readmissions would be expected to encourage providers to ensure that patients were discharged when medically fit and adequately supported after discharge. |

##### S –Policies

**Table 25.** Policies: Definitions and Rationales.

| Taxonomy heading | Brief description/definition | Mechanism for minimising winter pressures |
| --- | --- | --- |
| S – Policies | Policies represent recommended or mandated actions and may be developed at a variety of levels, e.g. national (devolved in UK); regional; local (e.g. local government, integrated care systems/boards); and institutional. See also CSB - Protocols and Guidelines | Policies can be developed in advance of expected winter pressures and/or enacted rapidly in response to changing circumstances. Their source may confer authority and they may be associated with monitoring and evaluation of implementation/effectiveness |
| Example interventions:<br>Patient FIRST; NHS England winter resilience plans | Patient FIRST is a CQC policy for emergency departments in the wake of the COVID-19 pandemic.<br>NHS England outlines required/recommended actions for Integrated Care Boards, NHS Trusts, GP practices and other organisations | Increased efficiency and safety in the ED incorporating learning from the pandemic<br>Prompts organisations to consider readiness for winter pressures and take appropriate actions |

**Table 26.**
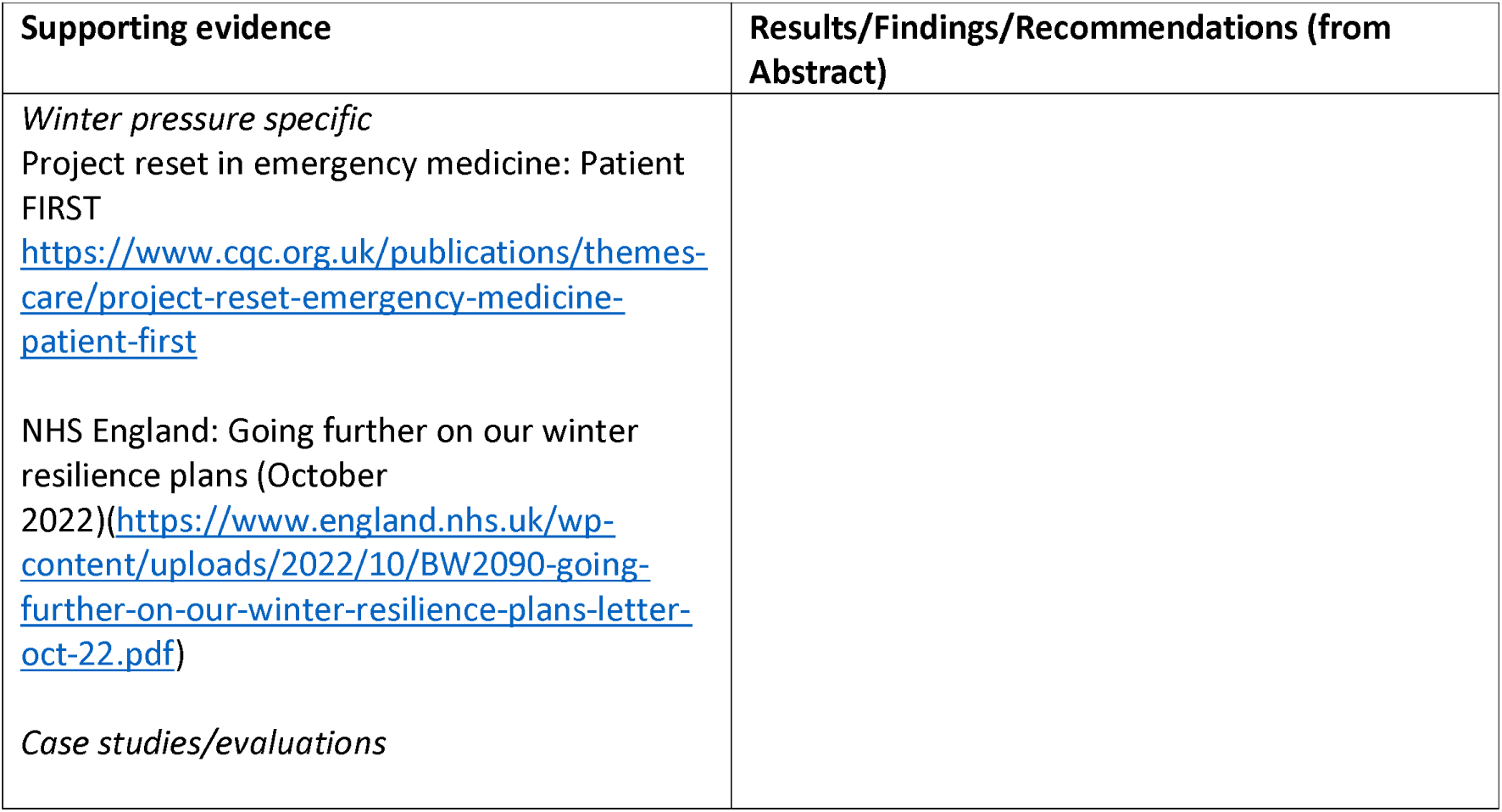
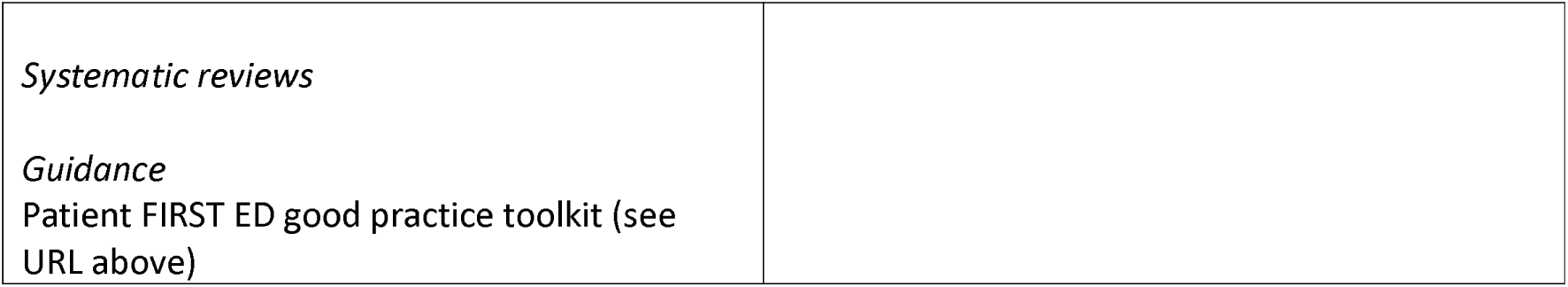
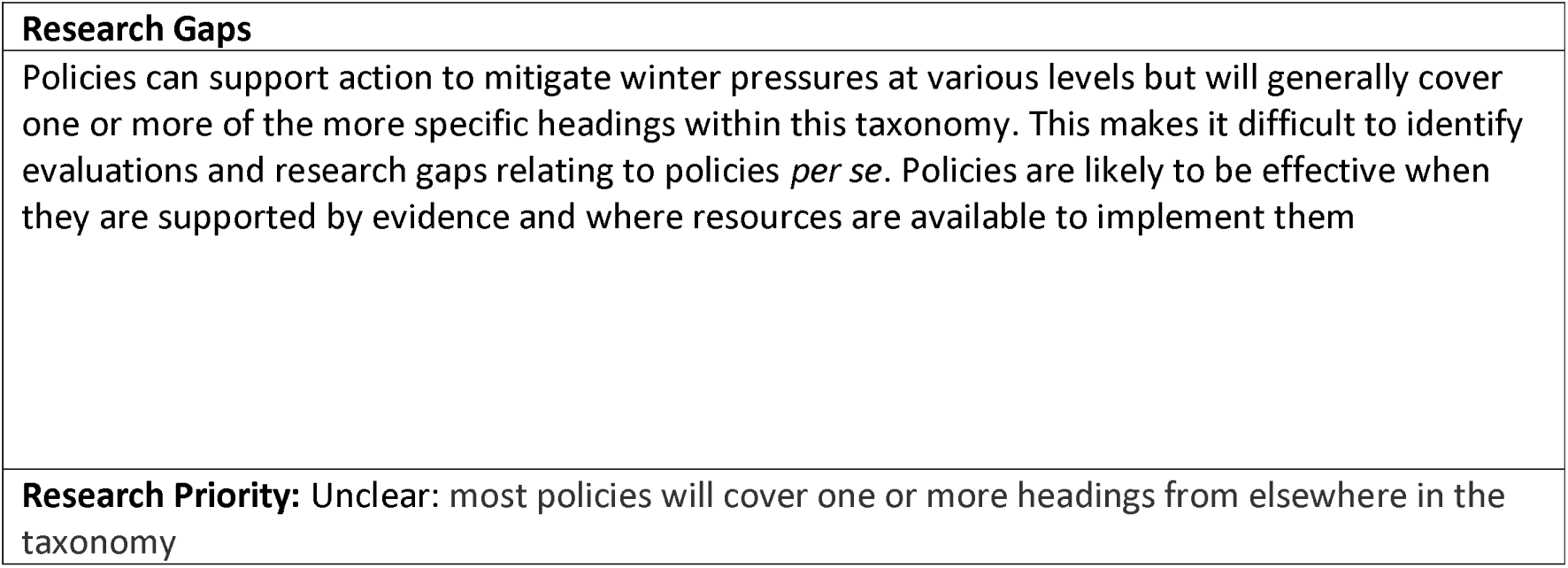
Policies: Interventions and Supporting Evidence.

| Supporting evidence | Results/Findings/Recommendations (from Abstract) |
| --- | --- |
| *Winter pressure specific*<br>Project reset in emergency medicine: Patient FIRST<br>https://www.cqc.org.uk/publications/themes-care/project-reset-emergency-medicine-patient-first<br>NHS England: Going further on our winter resilience plans (October 2022)(https://www.england.nhs.uk/wp-content/uploads/2022/10/BW2090-going-further-on-our-winter-resilience-plans-letter-oct-22.pdf)<br>*Case studies/evaluations* |  |
| *Systematic reviews*<br>*Guidance*<br>Patient FIRST ED good practice toolkit (see URL above) |  |

|  |
| --- |
| **Research Gaps** |
| Policies can support action to mitigate winter pressures at various levels but will generally cover one or more of the more specific headings within this taxonomy. This makes it difficult to identify evaluations and research gaps relating to policies *per se*. Policies are likely to be effective when they are supported by evidence and where resources are available to implement them |
| **Research Priority:** Unclear: most policies will cover one or more headings from elsewhere in the taxonomy |

##### S - Seven Day Services

**Table 27.** Seven day services: Definitions and Rationales.

| Taxonomy heading | Brief description/definition | Mechanism for minimising winter pressures |
| --- | --- | --- |
| S – Seven Day Services<br>Seven Day Services (7DS) | Provide high quality safe care every day of the week from 8am-8pm to improve patient flow through the system and across the interface between health and social care. | Minimise delays to patient flow through the system and across the interface between health and social care at the weekends to help manage increased healthcare usage during the winter months. |

**Table 28.**
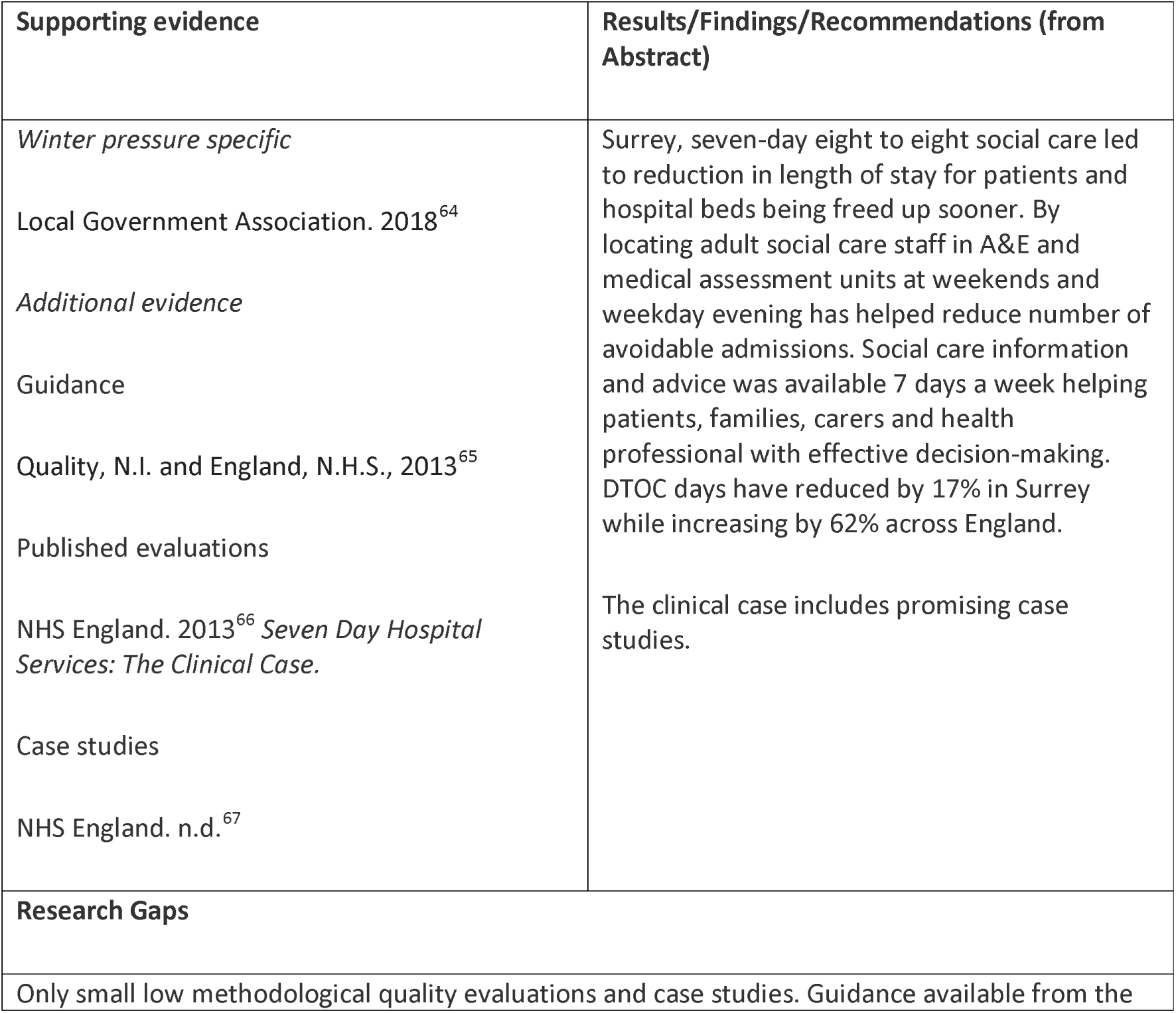
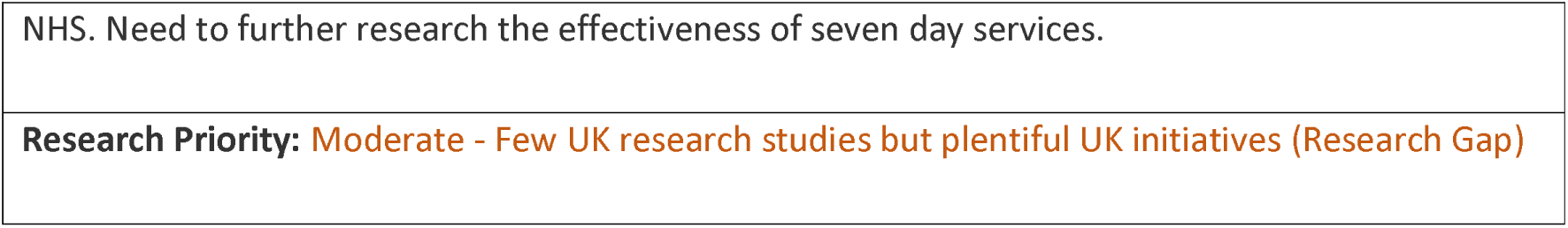
Seven day services: Interventions and Supporting Evidence.

##### S - Prioritisation and Triage

**Table 29.** Prioritisation and triage: Definitions and Rationales.

| Taxonomy heading | Brief description/definition | Mechanism for minimising winter pressures |
| --- | --- | --- |
| S – Prioritisation and triage | Prioritisation and triage (assigning patients to care based on the urgency of their needs) can take place at different levels of the healthcare system. In addition to triage in urgent and emergency care settings, times and/or sites can be allocated to more or less urgent procedures. | Can help to avoid hospital admission when safe and appropriate. Reducing variation where possible for routine procedures may represent a more efficient use of resources at times of high demand. Closely related to 'Patient flow' (e.g. green/red day system linked to SAFER patient flow bundle) |
| Example interventions: Green/red days; 'Hot' (trauma) and 'cold' (elective) sites for orthopaedics; rapid assessment areas | Examples of separation by time (red days indicate when a patient is awaiting an intervention); degree of urgency; and prioritisation in the ED | See above |

**Table 30.**
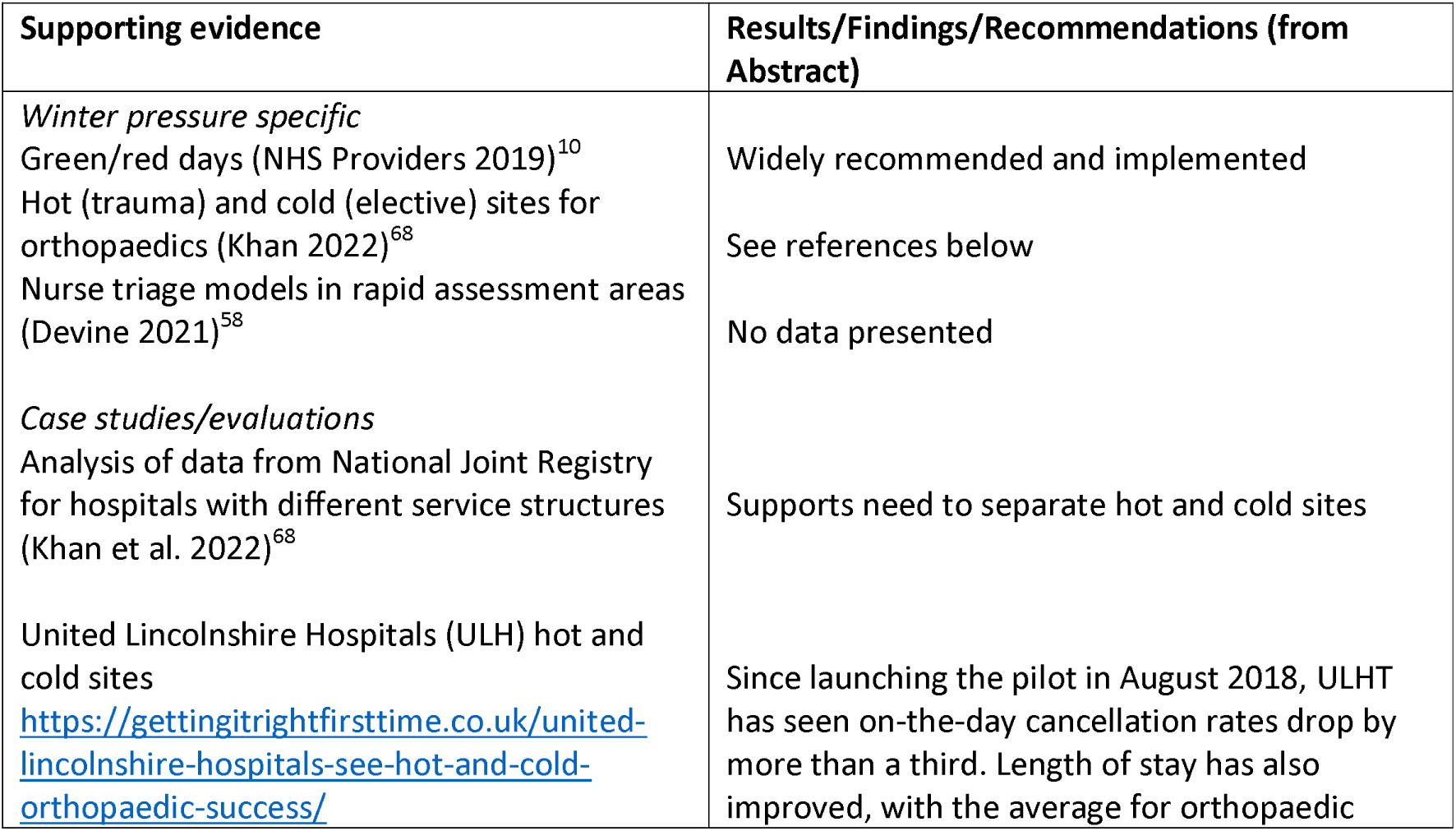
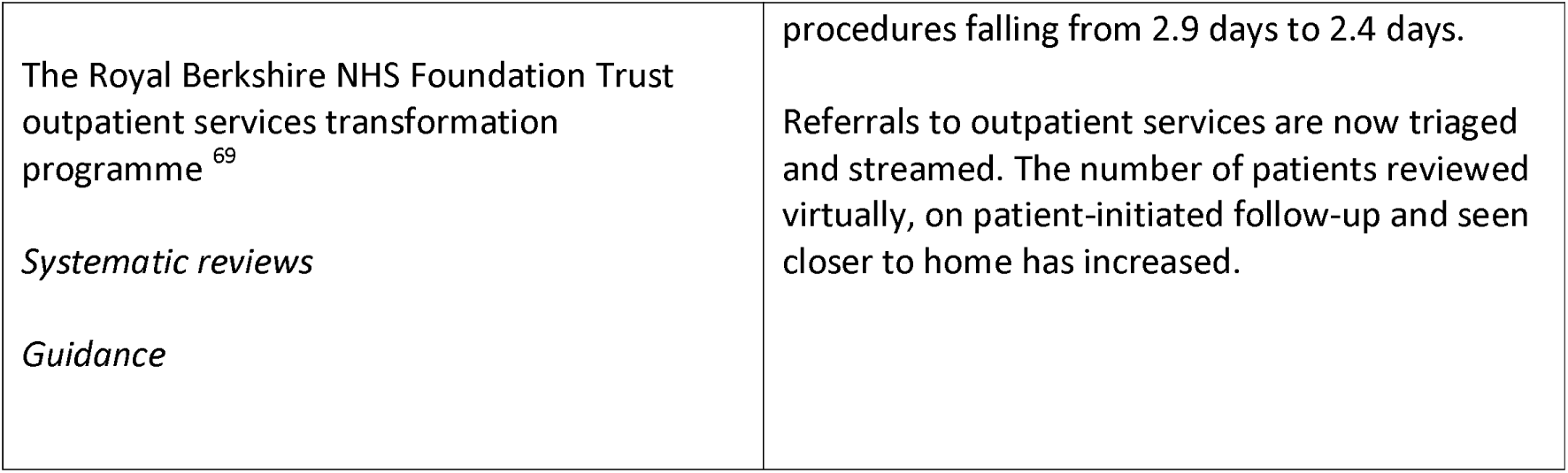
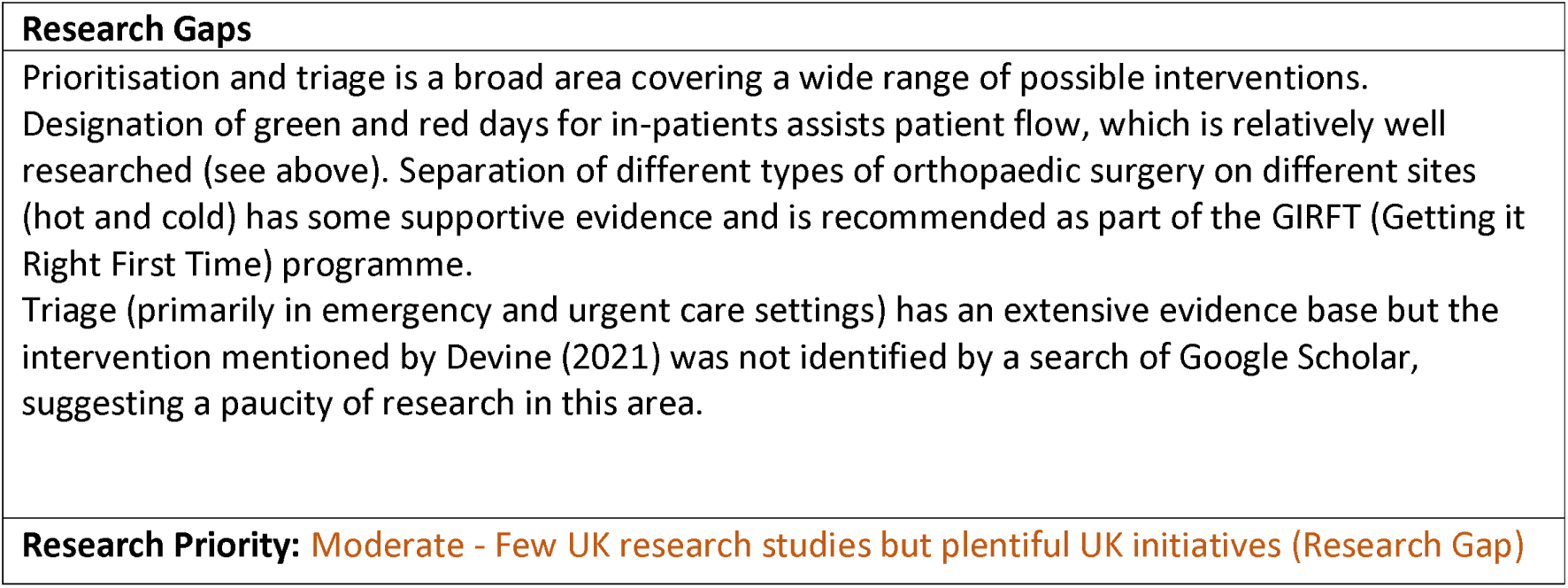
Prioritisation and triage: Interventions and Supporting Evidence.

##### S – Staff redeployment

**Table 31.** Staff redeployment: Definitions and Rationales.

| Taxonomy heading | Brief description/definition | Mechanism for minimising winter pressures |
| --- | --- | --- |
| **S – Staff Redeployment** | Redeployment of staff to improve patient flow through the health and social care system. | Help with patient flow through health and social care services, useful in winter when healthcare usage increases. |
| Examples of initiatives: Early Senior Review/Pharmacist on paediatric ward/Physiotherapists in ED/Respiratory nurse specialist in ED/Surgeons focus on day cases/Surgeons could support acute wards/ Sharing staff between trusts |  |  |

**Table 32.** Staff redeployment: Interventions and Supporting Evidence.

| Supporting evidence | Results/Findings/Recommendations (from Abstract) |
| --- | --- |
| *Winter pressure specific*<br>Care Quality Commission. 2022<sup>7</sup><br>Ferguson, F. 2020<sup>70</sup> (conference abstract)<br>Rickards, E. et al. 2021<sup>71</sup> (conference abstract)<br>Scriven, N., 2018<sup>72</sup><br>Wilkinson, E. 2018<sup>60</sup><br>*Additional evidence* | The recent systematic review on the impact of pharmacist interventions in the ED<sup>73</sup> found good evidence for improved quality use of medicines when pharmacists are included in the ED care teams. The number of medication errors reduced by a mean of 0.33 per patient. Patients had more complete and accurate medication histories, increased appropriateness of prescribed medications by 58% and time-critical medications initiation was quicker. However, no clear impact was observed on length of time spent in the ED or in hospital, and in-hospital mortality similar findings to the 2015 review<sup>74</sup>.<br>The feature by Wilkinson on how pharmacists are being reallocated to offer ward-based prescribing and seven-day pharmacy services to relieve winter pressures includes examples of relevant initiatives<sup>60</sup>. Luton and Dunstable University Hospital has The pharmacy prescription tracker, |
| *Guidance* | part of the 'control centre' suite of tools and once the trust hits a specific level of pressure, a member of the senior pharmacy team attends the bed meetings that are held four times per day. The carefully managed system allows the pharmacy team to be actively aware throughout the day of wider hospital pressures and able to meet anticipated changes in demand. |
| *Published evaluations* |  |
| *Systematic reviews* | A recent systematic review reviewed the evidence on the training and redeployment of healthcare workers to ICUs during the pandemic<sup>75</sup>. The review found that medical students experienced anxiety around treating critically ill patients and intubation procedures, they was also miscommunication with other healthcare workers and challenges from the tension between textbook medicine and complex social situation. The review findings could be helpful in informing future practice for redeployment and training in ICU settings for future pandemics and similar approaches could be introduced to mitigate the challenges of workforce shortage crisis affecting many countries and to mitigate winter pressures in UK. |
| Atey TM, Peterson GM, Salahudeen MS, et al. 2022<sup>73</sup> |  |
| Hohl CM, Wickham ME, Sobolev B, et al. 2015<sup>74</sup> |  |
| San Juan, N.V., Clark, S.E., Camilleri, M., Jeans, J.P., Monkhouse, A., Chisnall, G. and Vindrola-Padros, C., 2022<sup>75</sup>. |  |
| *Case studies* | A conference abstract details how emergency department physiotherapist practitioner (EDPP) posts were introduced to 3 EDs in Scotland. Across the 3 sites (28/01/19 to 07/04/19) the EDPP managed 1045 patients, freeing up doctors and nurses to see other more medically complex patients<sup>70</sup>.<br>Another conference abstract details the introduction a respiratory nurse specialist within the ED to provide care for patients presenting with a respiratory condition<sup>71</sup>. The nurse specialist prevented 240 admissions during a 5-month period and enabled discharges from ED to all community respiratory services. This initiative improved connections with community respiratory teams and standardisation of care for shared services.<br>Examples of initiatives related to Winter pressures<sup>7</sup>: Northamptonshire - shared strategy for workforce planning into the winter months was in place. Originally developed in response to the pandemic the workforce cell looked at national and local staffing drives as well as staff skills to see where |
|  | useful to move staff and where upskilling of staff could be beneficial. West Yorkshire and Harrogate, Suffolk and North East Essex, and Cornwall and the Isles of Scilly - passporting of staff between NHS trusts. The NHS competencies passport enabled staff to move between providers within a system more easily with the aim of reducing gaps in staff and improving patient care. |
| **Research Gaps** |  |
| Systematic reviews exist for certain staff redeployment initiatives, the use of pharmacists in ED and redeployment of staff to ICU. However, areas remain to research gaps where only UK initiatives exist. |  |
| **Research Priority:** Low - Good quality Systematic Review(s) and plentiful UK research studies (No Gap)<br>Moderate - Few UK research studies but plentiful UK initiatives (Research Gap) |  |

##### S – Volunteers

**Table 33.** Volunteers: Definitions and Rationales.

| **Taxonomy heading** | **Brief description/definition** | **Mechanism for minimising winter pressures** |
| --- | --- | --- |
| **S – Volunteers** | Use of volunteers to improve patient flow through the health and social care system. | Help with patient flow through health and social care services, useful in winter when healthcare usage increases. |

**Table 34.**
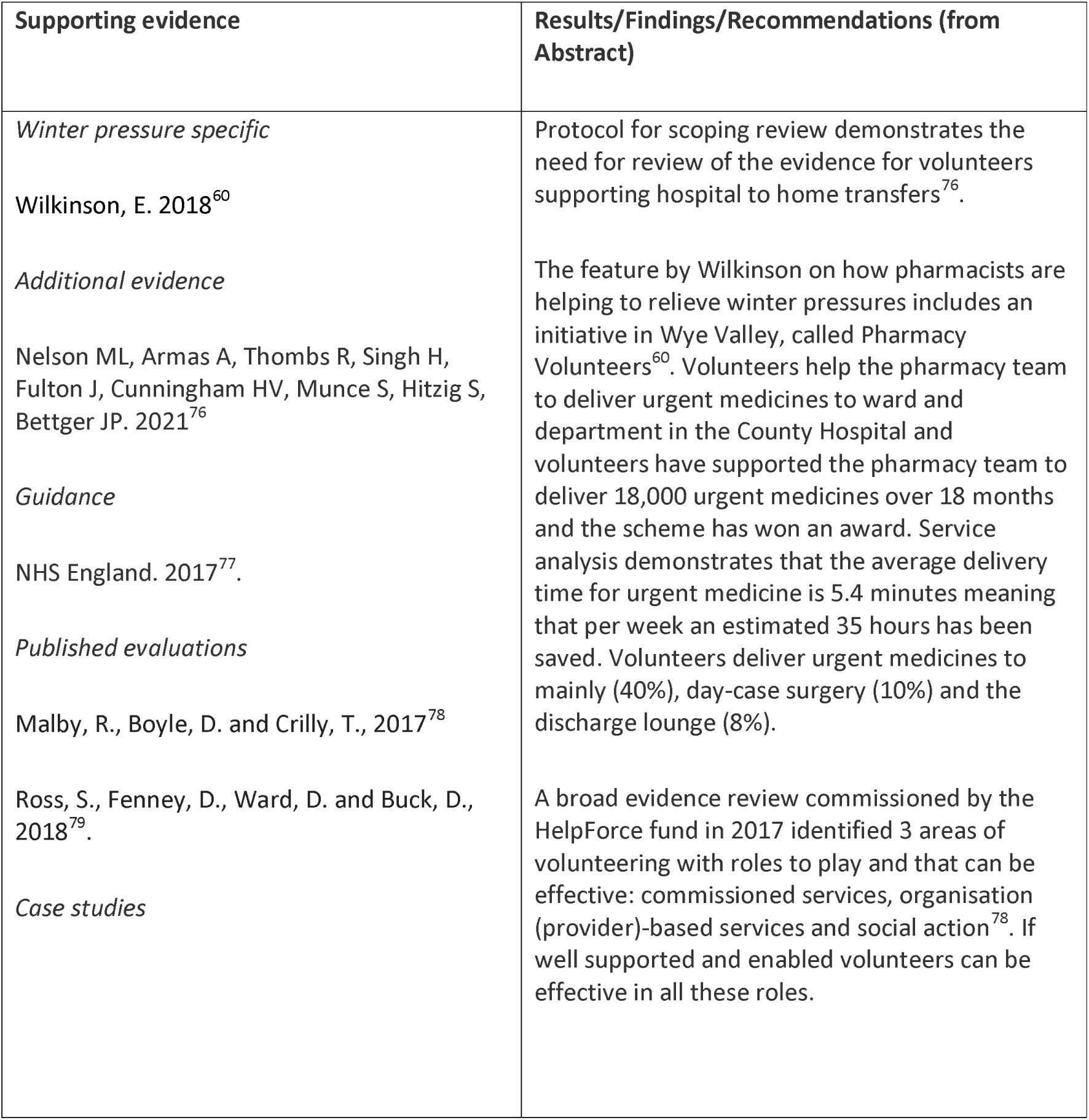
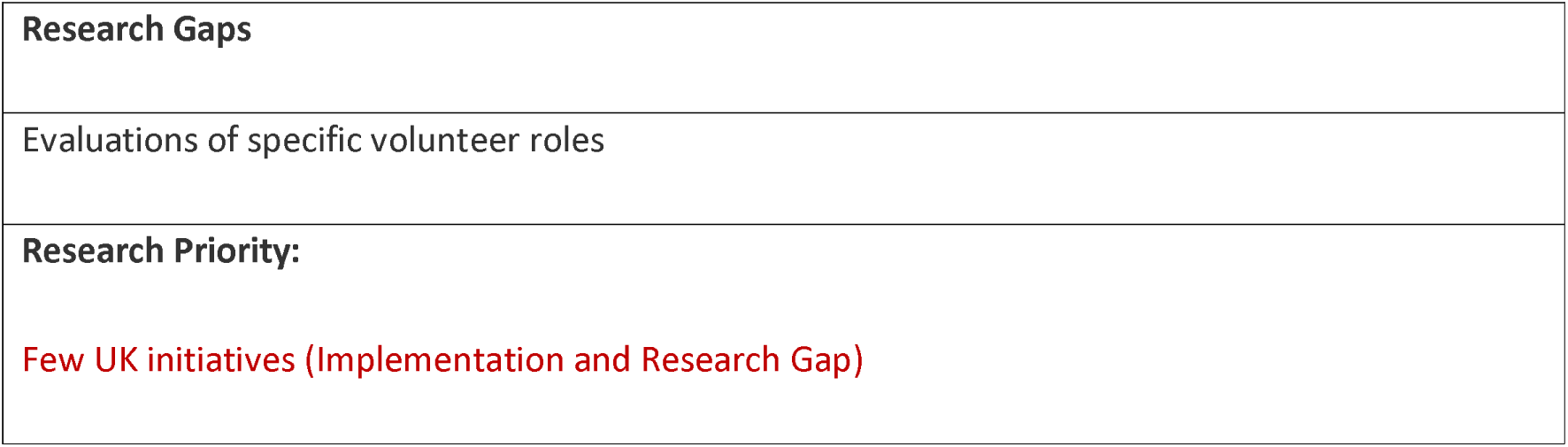
Volunteers: Interventions and Supporting Evidence.

| **Supporting evidence** | **Results/Findings/Recommendations (from Abstract)** |
| --- | --- |
| *Winter pressure specific*<br>Wilkinson, E. 2018<sup>60</sup><br>*Additional evidence*<br>Nelson ML, Armas A, Thombs R, Singh H, Fulton J, Cunningham HV, Munce S, Hitzig S, Bettger JP. 2021<sup>76</sup><br>*Guidance*<br>NHS England. 2017<sup>77</sup>.<br>*Published evaluations*<br>Malby, R., Boyle, D. and Crilly, T., 2017<sup>78</sup><br>Ross, S., Fenney, D., Ward, D. and Buck, D., 2018<sup>79</sup>.<br>*Case studies* | Protocol for scoping review demonstrates the need for review of the evidence for volunteers supporting hospital to home transfers<sup>76</sup>.<br>The feature by Wilkinson on how pharmacists are helping to relieve winter pressures includes an initiative in Wye Valley, called Pharmacy Volunteers<sup>60</sup>. Volunteers help the pharmacy team to deliver urgent medicines to ward and department in the County Hospital and volunteers have supported the pharmacy team to deliver 18,000 urgent medicines over 18 months and the scheme has won an award. Service analysis demonstrates that the average delivery time for urgent medicine is 5.4 minutes meaning that per week an estimated 35 hours has been saved. Volunteers deliver urgent medicines to mainly (40%), day-case surgery (10%) and the discharge lounge (8%).<br>A broad evidence review commissioned by the HelpForce fund in 2017 identified 3 areas of volunteering with roles to play and that can be effective: commissioned services, organisation (provider)-based services and social action<sup>78</sup>. If well supported and enabled volunteers can be effective in all these roles. |

|  |
| --- |
| **Research Gaps** |
| Evaluations of specific volunteer roles |
| **Research Priority:**<br>**Few UK initiatives (Implementation and Research Gap)** |

In summary, evidence support for initiatives in this group varied widely. The winter pressure search identified UK initiatives to support improved teamwork, together with associated guidance (Table 19). Various initiatives have applied data sharing and digital technology, but we found a relative lack of research studies. Actions at the organisational level, e.g. involving changes in governance or formulation and implementation of policy, play an important part in responding to winter pressures but we found this was not reflected in research. Further research studies are needed in the area of seven day services although promising case studies do exist. Staff redeployment has achieved successes within the pharmacy profession. Further research into staff redeployment would be useful. Volunteers have been used within the NHS in many different areas but there needs to be further evaluation of these initiatives in relation to winter pressures.

### Changing Staff Behaviour

Findings for taxonomy headings classified as changing staff behaviour are summarised in Tables 35 to 44. Detailed information can be found in the tables. The taxonomy headings were clinical audit, education of staff, protocols/guidelines, quality improvement programmes and quality management systems.

#### Changing Staff Behaviour – Cross-cutting

All of the headings were classified as cross-cutting.

##### CSB – Clinical Audit

**Table 35.** Clinical Audit: Definitions and Rationales.

| Taxonomy heading | Brief description/definition | Mechanism for minimising winter pressures |
| --- | --- | --- |
| CSB - Clinical Audit | Clinical audit is a method to find out if healthcare delivery meets relevant standards and it allows patients and care providers to understand what their service is doing well and potential areas for improvement. | A clinic audit could be used to assess different healthcare policies and procedures for example compliance with discharge policy which could they suggested areas for improvement which could help to improve patient flow. |

**Table 36.** Clinical Audit: Interventions and Supporting Evidence.

| Supporting evidence | Results/Findings/Recommendations (from Abstract) |
| --- | --- |
| *Winter pressure specific*<br>Clinical audit has been used for benchmarking to mitigate winter pressures. The Winter Society for Acute Medicine Benchmarking Audit (SAMBA) provides a comparison of performance within acute medicine against clinical quality indicators during winter.<br>*Additional evidence*<br>Guidance<br>Healthcare Quality Improvement Partnership. | No evidence of the use of clinical audit to mitigate winter pressures.<br>Recent guidance from the Healthcare Quality Improvement Partnership on documenting local clinical audit<sup>80</sup>.<br>A case study of the use of clinical audit to investigate compliance with discharge policy at SaTH. The initial audit in 2014 evaluated compliance with the discharge policy and suggested changes to address identified problems<sup>81</sup>. In 2015 re-audit sought to determine if recommendations from 2014 audit had been implemented, compliance with the 2015 discharge policy and any changes to improve the |
| 2020<sup>80</sup><br>Published evaluations<br>Case studies<br>Shrewsbury and Telford Hospital (SaTH). 2014<sup>81</sup><br>Shrewsbury and Telford Hospital (SaTH). 2015<sup>82</sup><br>Shrewsbury and Telford Hospital (SaTH). 2019<sup>83</sup> | discharge experience for patients<sup>82</sup>. In 2019, after changes from the previous audit had had time to be embedded in practice, a re-audit checked if recommendations from previous audits had been implemented<sup>83</sup>. The 2019 audit found that patients were kept waiting for the same reasons in the discharge lounge, that transferral to the discharge lounge only happened for a small number of patients. |
| **Research Gaps** |  |
| Clinical audit could be used at a local level as a methodology to investigate the effectiveness of initiatives to mitigate winter pressures. Clinical audits typically target local and national priorities, areas of clinical variability and adverse events and so are well aligned to the needs of exploring winter pressures. However, a tension exists between clinical demand and resources required for audit making most candidate approaches likely to be retrospective. The Winter Society for Acute Medicine Benchmarking Audit (SAMBA) indicates the potential for national approaches relying on uniform national datasets. The recently announced Winter Improvement Collaborative (see separate entry) also relies on audit and benchmarking principles in targeting a narrow and specific range of indicators. |  |
| **Research Priority:** Few UK initiatives (Implementation and Research Gap) |  |

##### CSB – Education of Staff

**Table 37.** Education of Staff: Definitions and Rationales.

| **Taxonomy heading** | **Brief description/definition** | **Mechanism for minimising winter pressures** |
| --- | --- | --- |
| CSB - Education of Staff | Education of staff | Staff education is important to ensure that they can fulfil their roles and provide safe effective care making it an important part of new initiatives. |
|  | Staff education about patient flow | Good patient flow in an acute hospital helps ensure patients experience minimal delays throughout their hospital journey. Effective processes, staff buy-in and staff education are needed for good flow. |

**Table 38.**
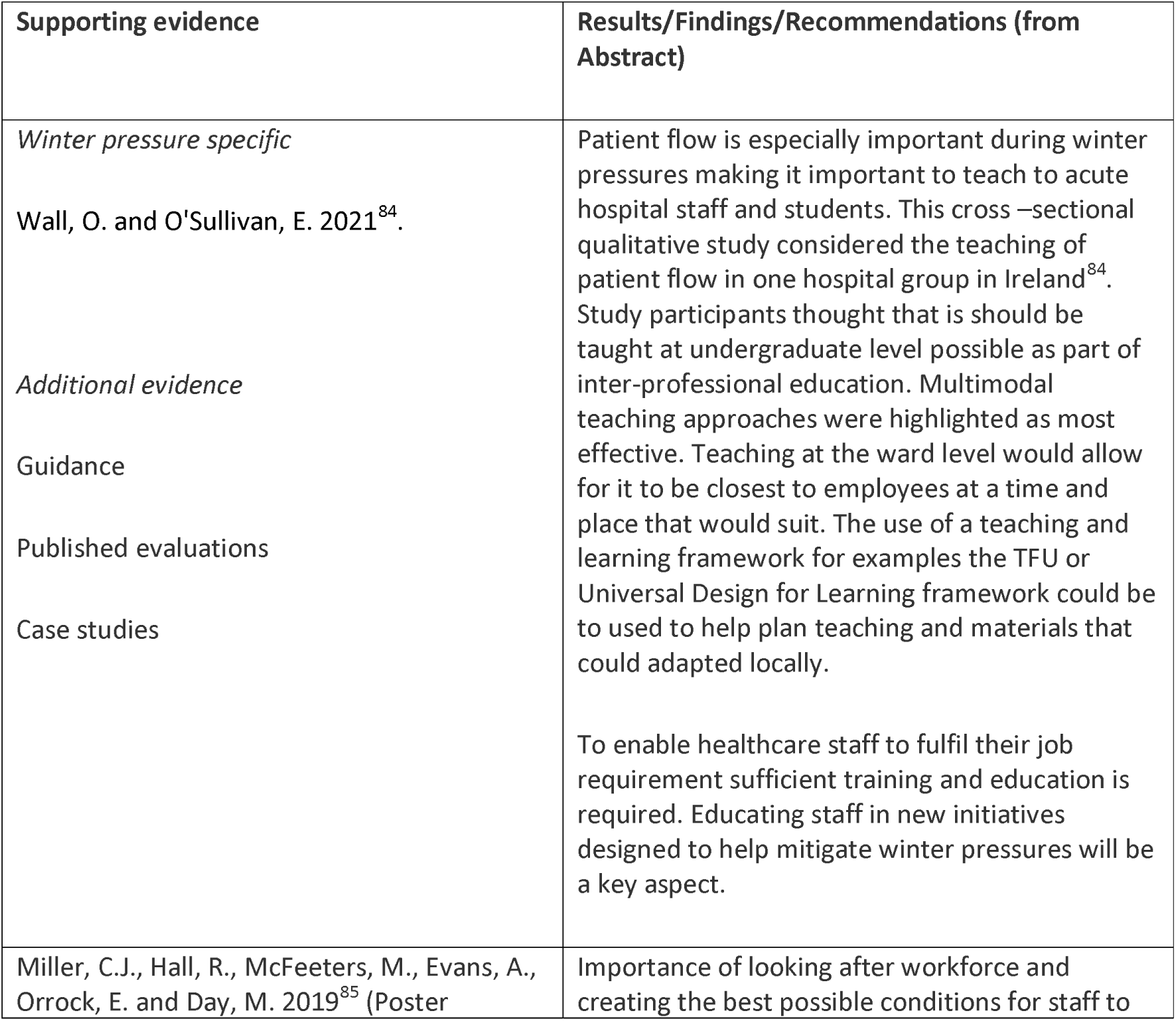
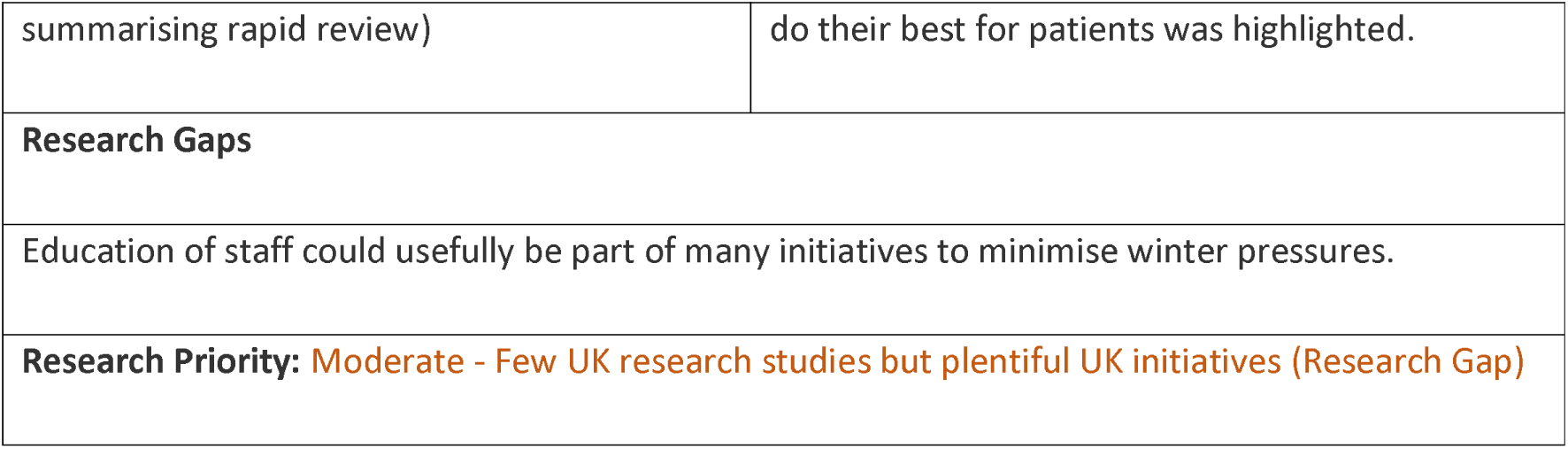
Education of Staff: Interventions and Supporting Evidence.

##### CSB – Protocols/Guidelines

**Table 39.**
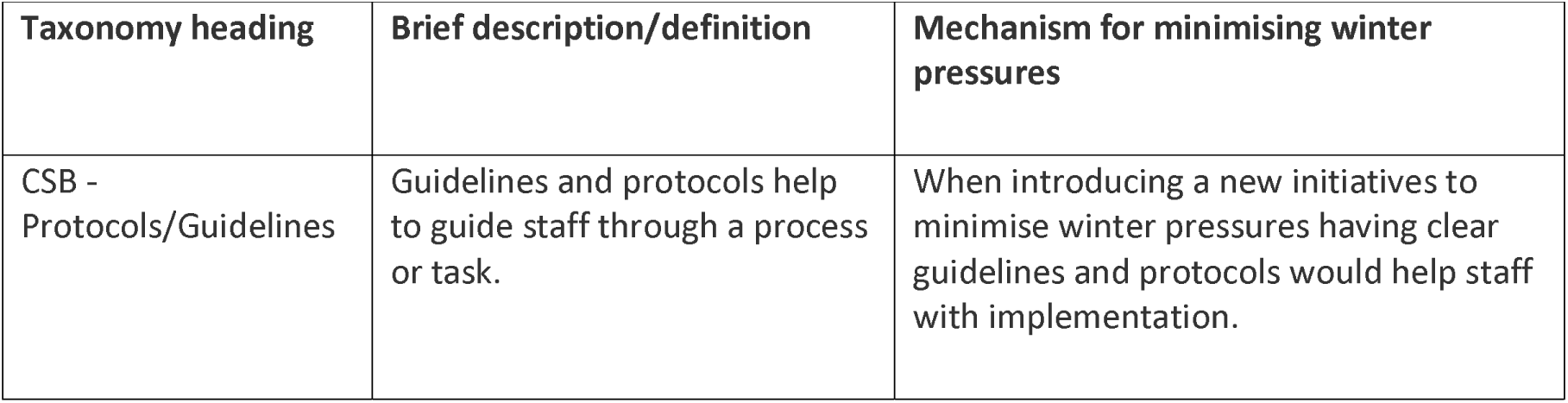
Protocols/Guidelines: Definitions and Rationales.

| Taxonomy heading | Brief description/definition | Mechanism for minimising winter pressures |
| --- | --- | --- |
| CSB - Protocols/Guidelines | Guidelines and protocols help to guide staff through a process or task. | When introducing a new initiatives to minimise winter pressures having clear guidelines and protocols would help staff with implementation. |

**Table 40.** Protocols/Guidelines: Interventions and Supporting Evidence.

| Supporting evidence | Results/Findings/Recommendations (from Abstract) |
| --- | --- |
| *Winter pressure specific*<br>*Additional evidence*<br>Systematic review<br>Unnewehr, M., Schaaf, B., Marev, R., Fitch, J. and Friederichs, H., 2015<sup>86</sup><br>Guidance<br>Published evaluations<br>Case studies | No evidence for the use of protocols/ guidelines to mitigate winter pressures.<br>A systematic review from 2015 aimed to consider how the quality of discharge summaries could be improved by (1) intervention, (2) reviews and guidelines of regulatory bodies; and (3) other practical recommendations<sup>86</sup>. Guidelines were helpful for providing the key components of discharge summaries. |
| **Research Gaps** |  |
| Protocols/guidelines are important when introducing new initiatives and thus could be an important part of new initiatives to mitigate winter pressures. |  |
| **Research Priority:** Few UK initiatives (Implementation and Research Gap) |  |

##### CSB – Quality Improvement Programmes

**Table 41.** Quality Improvement Programmes: Definitions and Rationales.

| **Taxonomy heading** | **Brief description/definition** | **Mechanism for minimising winter pressures** |
| --- | --- | --- |
| CSB - Quality Improvement Programmes | A systematic approach to improve quality in a specific area. The programme involves the people delivering the service in introducing initiative to improve their service. | Quality improvement programmes are useful for implementing new initiatives and thus could be useful when introducing initiative to mitigate winter pressures. |
|  | Healthcare quality improvement programmes have generated large numbers of healthcare quality improvement initiatives. |  |

**Table 42.**
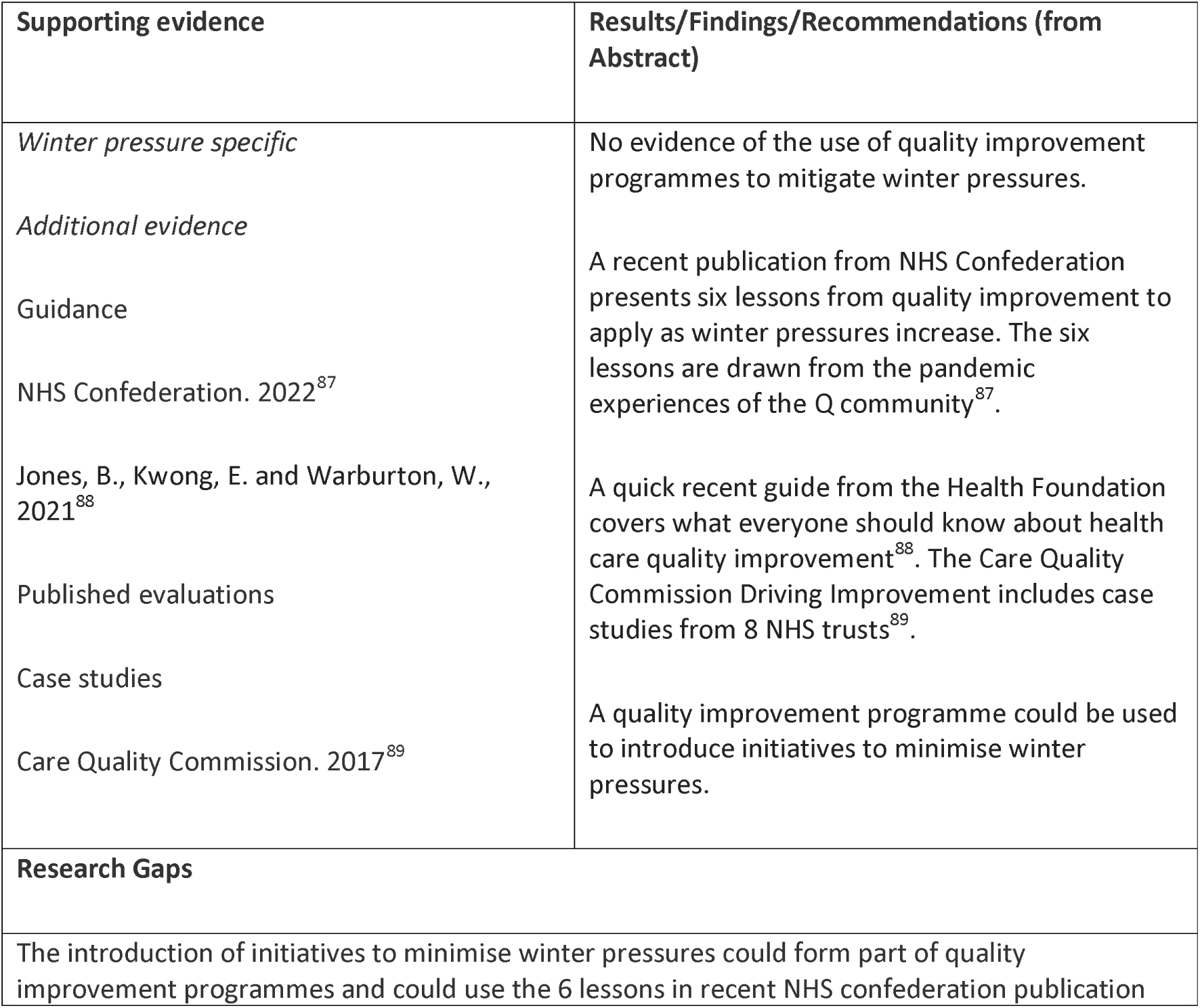
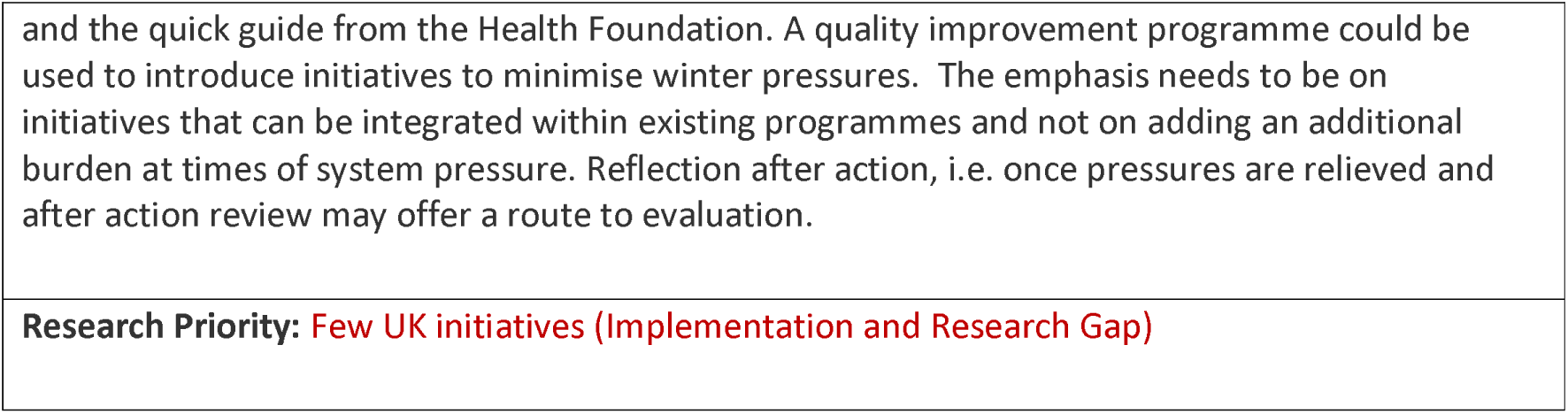
Quality Improvement Programmes: Interventions and Supporting Evidence.

##### CSB – Quality Management Systems

**Table 43.** Quality Management Systems: Definitions and Rationales.

| Taxonomy heading | Brief description/definition | Mechanism for minimising winter pressures |
| --- | --- | --- |
| CSB - Quality Management Systems | Quality management systems support the delivery of reliable and high quality care. | Quality management systems can help services to continue to deliver reliable and high quality care while experiencing winter pressures. |

**Table 44.**
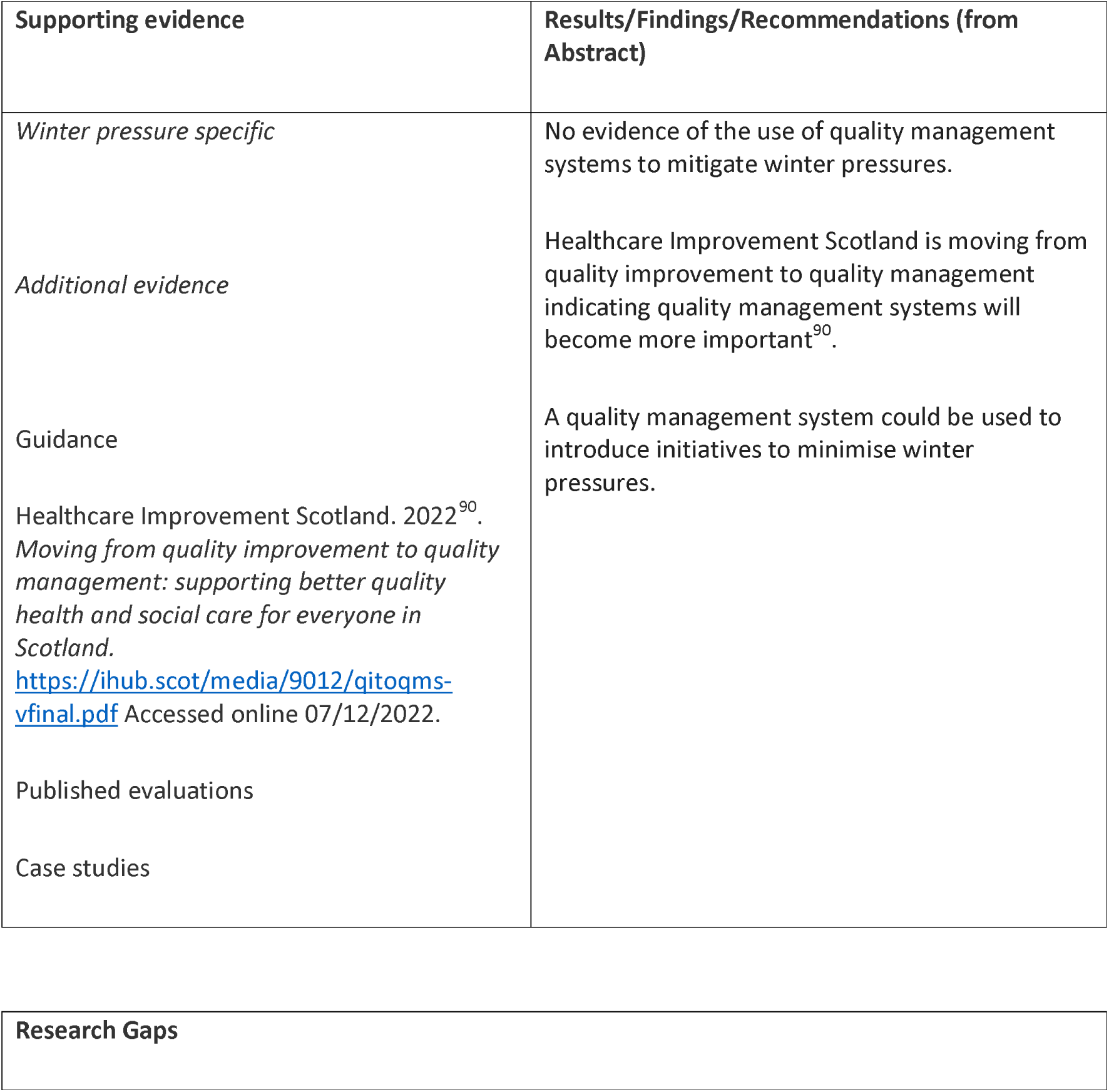
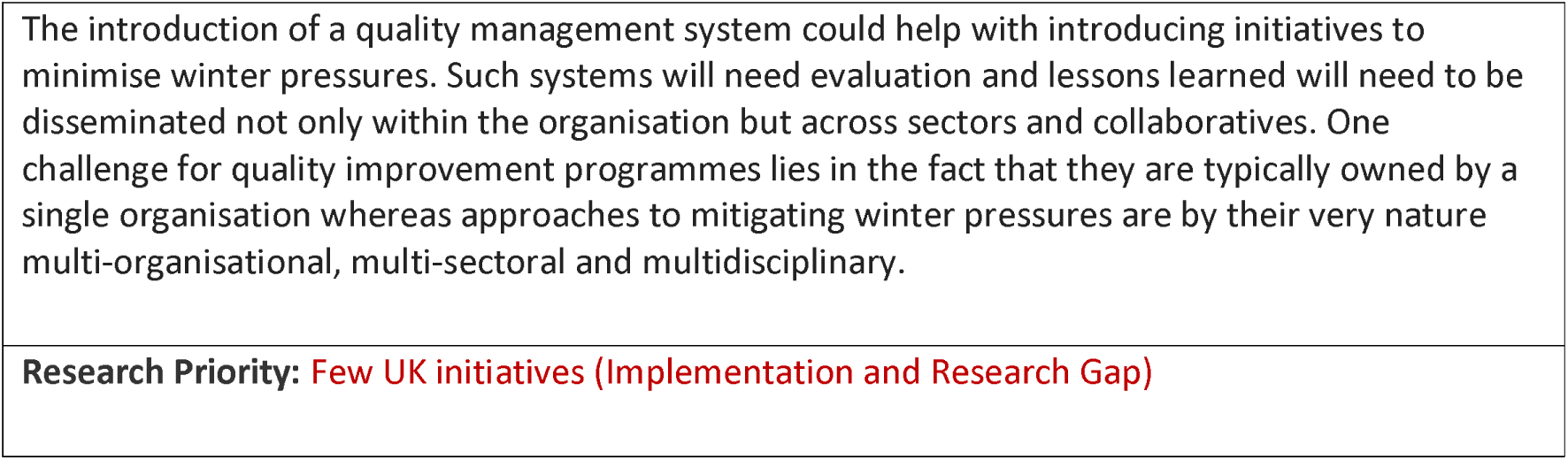
Quality Management Systems: Interventions and Supporting Evidence.

Within this taxonomy section, we identified limited initiatives or research studies. Changing staff behaviour can take time and initiatives to minimise winter pressures may need to be implemented rapidly without the time for thorough evaluation or to measure all potential outcomes. Initiatives could potentially be too short to make changes to staff behaviour and they might not measure staff behaviour as an outcome. Further research studies could plan evaluations that consider a variety of outcomes including changes in staff behaviour, although it can be difficult to measure.

### Changing Community Provision

In the first version of the taxonomy, as employed during the initial winter pressures mapping process, all Changing Community Provision (CCP) entries were subsumed under a single overarching heading. Subsequently, as we sought evidence on the specific intervention types, the review team decided to differentiate community provision within three subcategories. We believe that this is the first, and important, step towards starting to think how different types of intervention attempt to respond to winter pressures in advance of a realist focus on mechanisms. The three subcategories are therefore labelled as CCP – Hospital Avoidance, CCP – Alternate Delivery Site, and CCP – Facilitated Discharge. Each is considered, together with specific interventions under each subcategory in the tables below.

#### Changing Community Provision - Hospital Avoidance

Findings for taxonomy headings classified as changing community provision are summarised in Tables 45 to 52. Detailed information can be found in the tables. The taxonomy headings were rapid response/see and treat, single point response and step up facilities.

**Table 45.** Hospital Avoidance: Definitions and Rationales.

| **Taxonomy heading** | **Brief description/definition** | **Mechanism for minimising winter pressures** |
| --- | --- | --- |
| CCP – Hospital Avoidance | Provides active treatment by health care professionals outside hospital for a condition that otherwise would require admission. | Offers treatment outside hospital as alternative to acute hospital in-patient admission |
| Example interventions: 'Community Connect' (Somerset) | Works alongside 'Home First' discharge model using mix of Voluntary, Community and Social Enterprise and micro-provider provision to provide rapid response away from hospital setting. Drew upon robust community infrastructure including village agents (people who link their local communities to services/information they need and at times deliver it themselves) county-wide. | Harnesses community resilience to allow people to receive care closer to their home, support admission avoidance and link to discharge to assess model (home with therapy support and reablement beds). |
| Hospital avoidance response team | Provided to either prevent an avoidable A&E attendance or admission, or speed up discharge from secondary care. Facilitates supported discharge | 72-hour period gives other domiciliary or reablement services opportunity to commence later in the pathway supporting the clinical |
|  | and provides up to 72 hours of care and support to resettle a person at home offering 'bridging the gap service' for 72-hour period offering telecare unit, enabling access to responders 24/7 – assurance of support through the night if required and wellbeing service assessment with onward referral as appropriate. | assessment service to avoid hospital admission and/or attendance at A&E. |
| Example interventions:<br>**Hospital avoidance response team**<br>Local Government Managing transfers of care resource<br>https://www.local.gov.uk/case-studies/lincolnshire-independent-living-partnership-hospital-avoidance-response-team | Service delivered by Lincolnshire Independent Living Partnership which takes referrals from secondary care discharge hubs, A&E in-reach teams, the ambulance service, primary care and community health providers. | Provides appropriate support and referral bypassing formal presentation to hospital services |
| Example interventions:<br>Talking Cafes (Somerset) | Village agents run talking cafes (face to face or online) to offer a point of contact and reassurance. Village agent service works with local authority and GPs to support the vulnerable, setting up food and equipment networks, and harnessing support from within communities. | Increase community resilience by offering – local events where people can get information, advice, support, or even company |

**Table 46.**
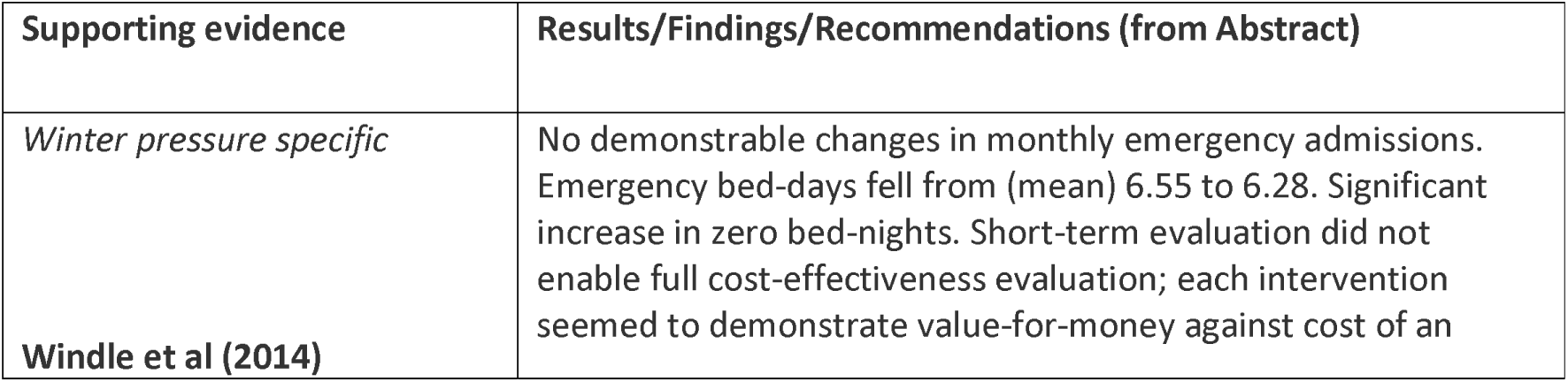
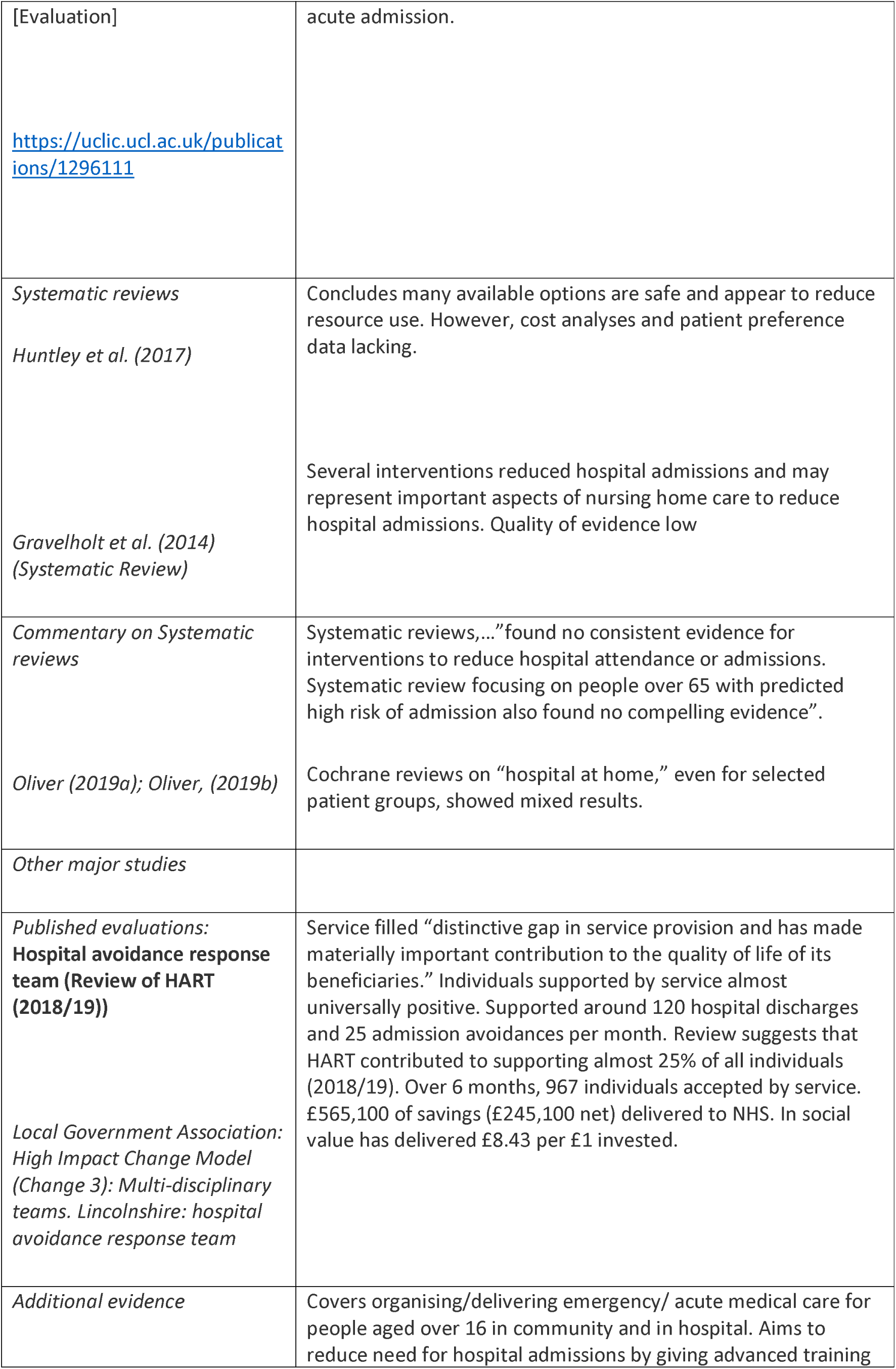
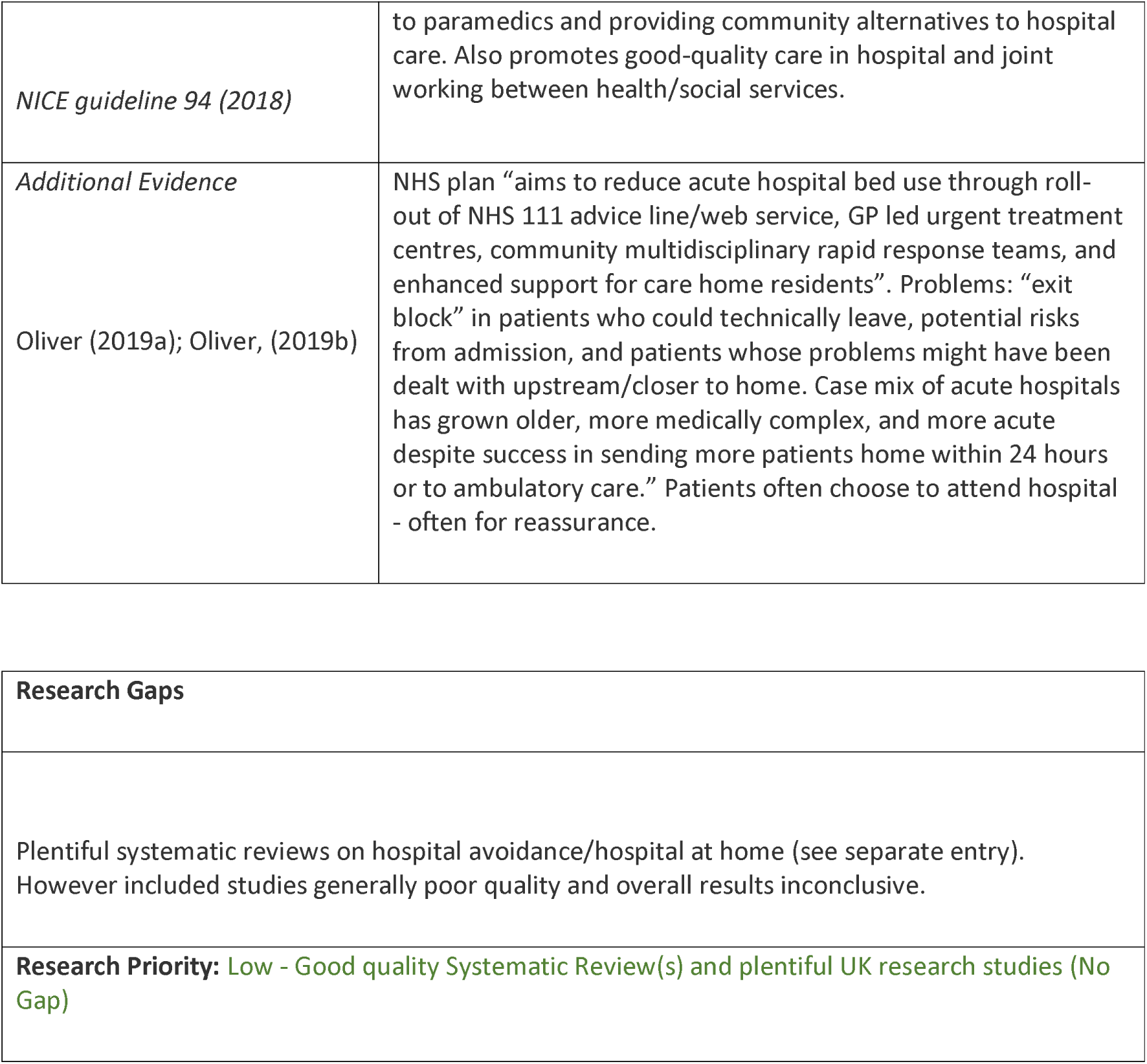
Hospital Avoidance: Interventions and Supporting Evidence.

##### CCP - Rapid Response/See and Treat

**Table 47.** Rapid Response/See and Treat: Definitions and Rationales.

| **Taxonomy heading** | **Brief description/definition** | **Mechanism for minimising winter pressures** |
| --- | --- | --- |
| CCP - Rapid Response/See and Treat | Aim to support user by providing urgent response in their own home or other location either to prevent admission or to facilitate discharge from the acute hospital sector. Usually led by nurse/allied health professional; cover diverse interventions including administration of intravenous therapies, peg tube and catheter replacement, crisis psychiatric care and provide enhanced care to palliative care patients (NICE Guideline 94). | Aims to treat potential hospital admissions in situ thereby reducing admission and potential utilisation of hospital services and beds. |
| **Example interventions: Falls Specialist Response Car** | Provided by Ambulance Service under call sign K466, staffed with Community Treatment team (CTT) nurse and paramedic. LAS Control Centre identify patient on criteria including the patient being over 65. Service operates seven days a week from 07:00-19:00. | Aims to keep 20 patients at home per week to reduce attendance, admissions and ambulance conveyances to Emergency Department. |
| **Example interventions: Community based falls response service** | Seek to improve response times and enable ambulance services to prioritise higher acuity patients. Service handles level one and two falls (no known illness or injury, or minor injury/illness) for adults over 18 in people's own homes or in care homes. All people who fall and are unable to get up receive appropriate clinical assessment to determine level of response. Coordinated by appropriate level clinicians, e.g. advanced practitioners within ambulance emergency control room, with clinical follow-up to assess fall reason and risk assess where appropriate. | Part of Winter Resilience Plans. Aims to reduce pressures on general practice and social care, and reduce admissions to hospital. |
| **Example interventions: Community urgent** | Collective name for services that improve quality/capacity of care for people through delivery of urgent, | Seeks to avoid demands on hospital care by providing a community based service that |
| **care schemes** | crisis response care within two-hours and/or reablement care responses within two-days. Some providers offer a single, integrated service that covers all these types of care from crisis response to reablement. This is preferred service delivery model. | targets urgent needs. |
| **Example interventions: Hull Fall First** | Falls pick up/wellbeing response service within Hull. Involves: Hull CCG, Humberside Fire & Rescue, Yorkshire Ambulance Service, Hull City Council and City Health Care Partnership (current community services provider). Team of firefighters selected to receive clinical training from healthcare experts i.e. paramedics, occupational therapists and physiotherapists. Team also receives higher level safeguarding training and safety awareness. | Aims to ensure an appropriate level of clinical response according to causes and severity of falls ensuring only most critical cases are transported to hospital. |
| Rapid response vehicle with wireless point-of-care testing (Oxford)<br>(Novak et al, 2022). | Rapid response vehicle, staffed by specialist paramedic and equipped with wireless point-of-care testing platform to transmit results to hospital electronic patient record (EPR) systems, dispatched to frail, non-injured patients. Results obtained on scene and transmitted to Oxford University Hospitals EPR system. Patients discussed over telephone with a senior acute medical assessment physician at Trust for advice/decision support and to coordinate referral to other services. | Relieves pressure on physical attendance and increases availability of senior clinician |

**Table 48.**
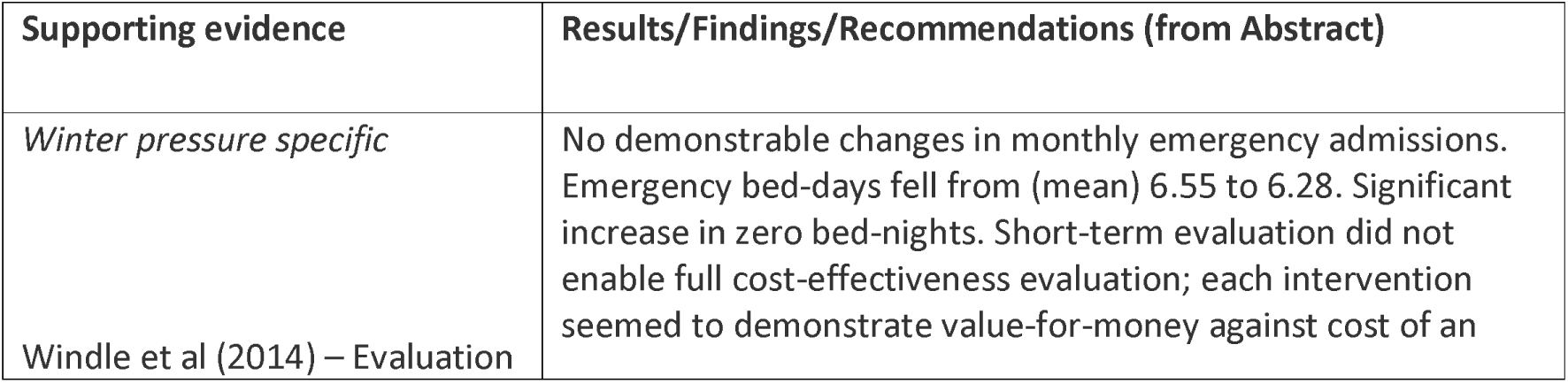
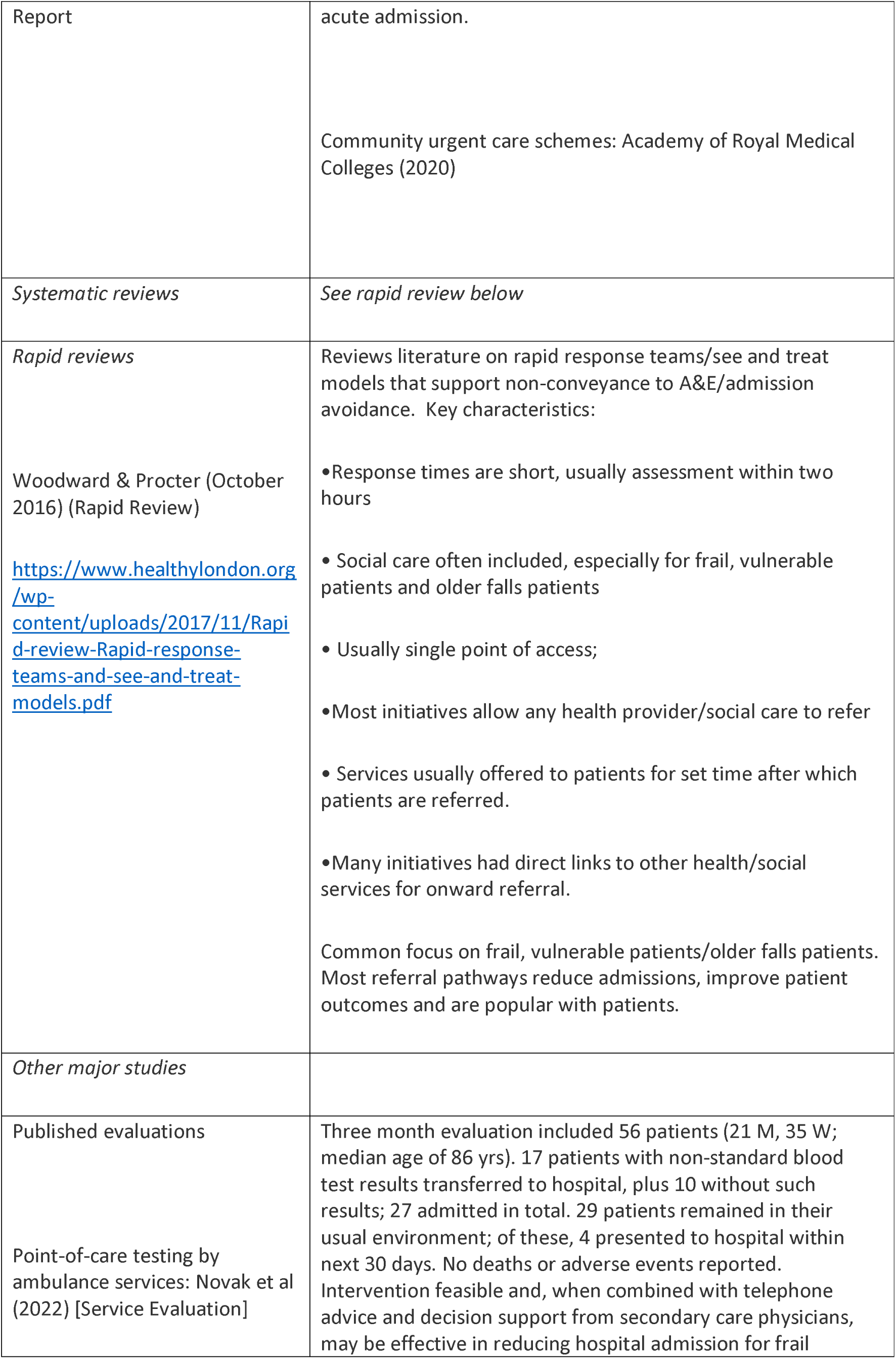
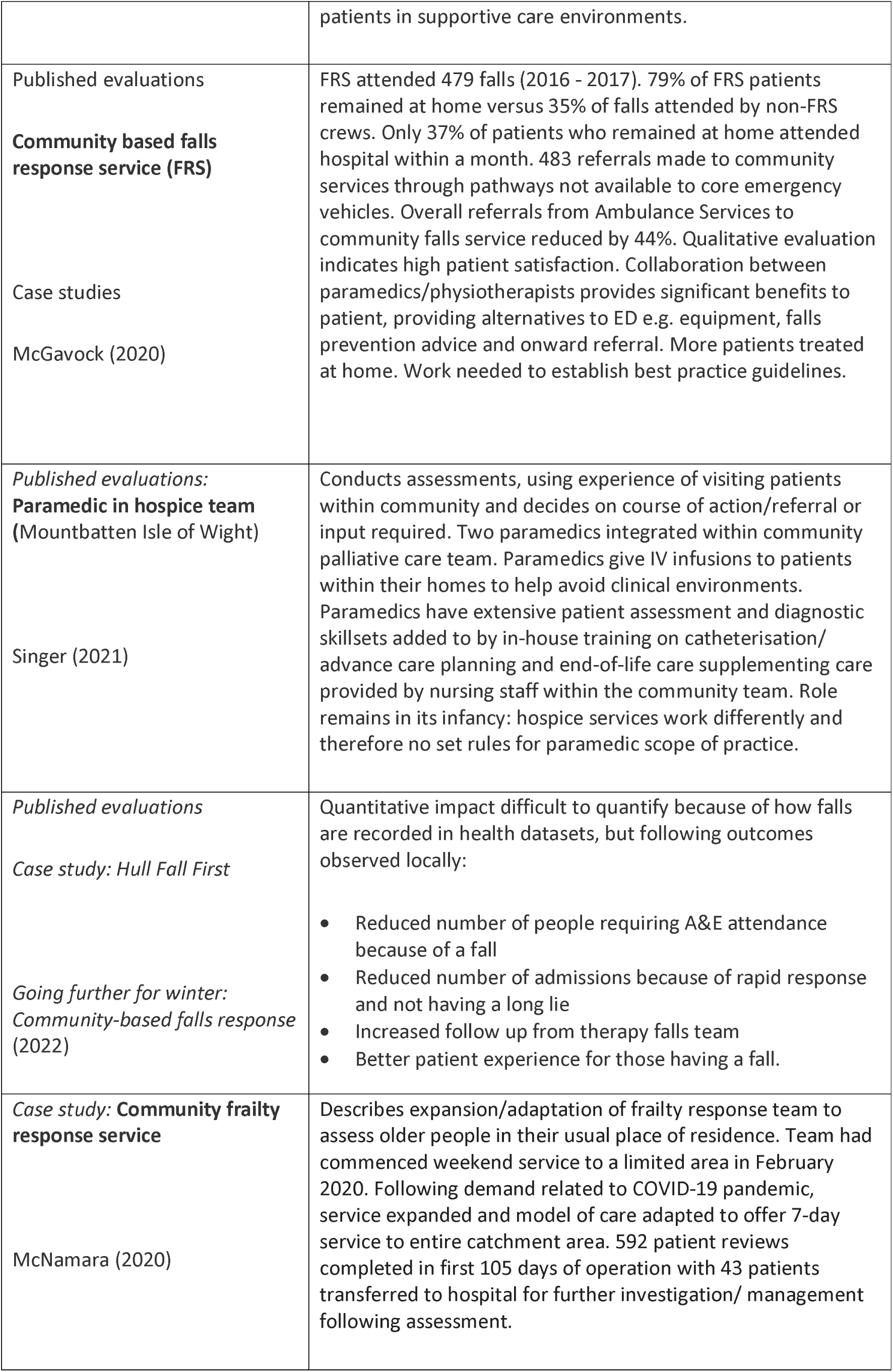
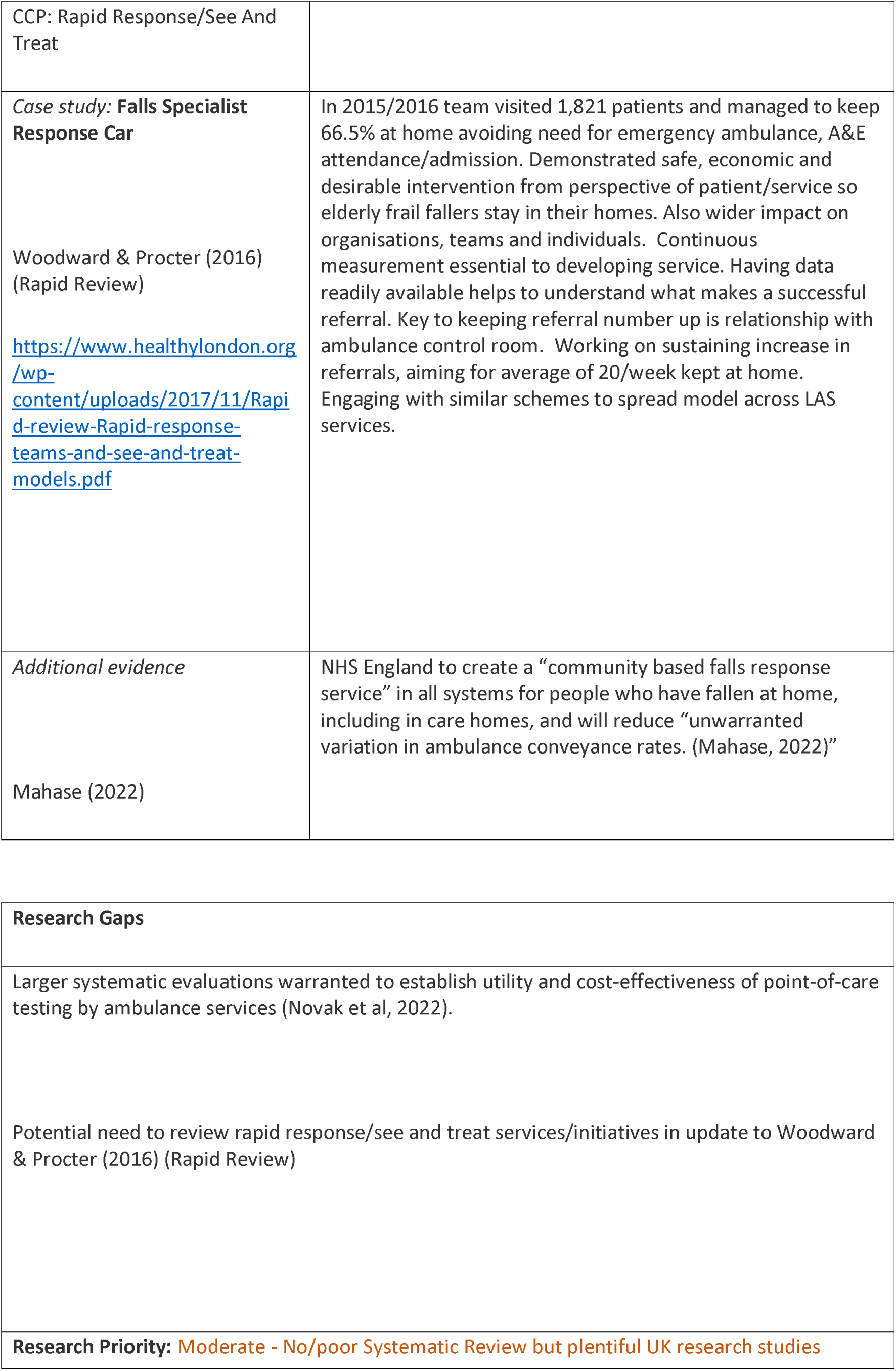
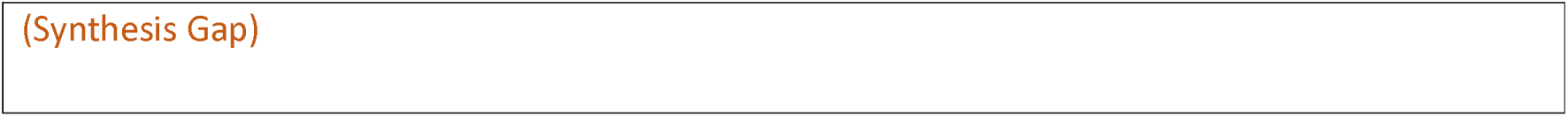
Rapid Response/See and Treat: Interventions and Supporting Evidence.

| Supporting evidence | Results/Findings/Recommendations (from Abstract) |
| --- | --- |
| Winter pressure specific<br>Windle et al (2014) – Evaluation | No demonstrable changes in monthly emergency admissions. Emergency bed-days fell from (mean) 6.55 to 6.28. Significant increase in zero bed-nights. Short-term evaluation did not enable full cost-effectiveness evaluation; each intervention seemed to demonstrate value-for-money against cost of an |
| Report | acute admission.<br>Community urgent care schemes: Academy of Royal Medical Colleges (2020) |
| *Systematic reviews* | *See rapid review below* |
| *Rapid reviews*<br>Woodward & Procter (October 2016) (Rapid Review)<br>https://www.healthylondon.org/wp-content/uploads/2017/11/Rapid-review-Rapid-response-teams-and-see-and-treat-models.pdf | Reviews literature on rapid response teams/see and treat models that support non-conveyance to A&E/admission avoidance. Key characteristics:<br>• Response times are short, usually assessment within two hours<br>• Social care often included, especially for frail, vulnerable patients and older falls patients<br>• Usually single point of access;<br>• Most initiatives allow any health provider/social care to refer<br>• Services usually offered to patients for set time after which patients are referred.<br>• Many initiatives had direct links to other health/social services for onward referral.<br>Common focus on frail, vulnerable patients/older falls patients. Most referral pathways reduce admissions, improve patient outcomes and are popular with patients. |
| *Other major studies* |  |
| Published evaluations<br>Point-of-care testing by ambulance services: Novak et al (2022) [Service Evaluation] | Three month evaluation included 56 patients (21 M, 35 W; median age of 86 yrs). 17 patients with non-standard blood test results transferred to hospital, plus 10 without such results; 27 admitted in total. 29 patients remained in their usual environment; of these, 4 presented to hospital within next 30 days. No deaths or adverse events reported. Intervention feasible and, when combined with telephone advice and decision support from secondary care physicians, may be effective in reducing hospital admission for frail |
|  | patients in supportive care environments. |
| Published evaluations<br>**Community based falls response service (FRS)**<br>Case studies<br>McGavock (2020) | FRS attended 479 falls (2016 - 2017). 79% of FRS patients remained at home versus 35% of falls attended by non-FRS crews. Only 37% of patients who remained at home attended hospital within a month. 483 referrals made to community services through pathways not available to core emergency vehicles. Overall referrals from Ambulance Services to community falls service reduced by 44%. Qualitative evaluation indicates high patient satisfaction. Collaboration between paramedics/physiotherapists provides significant benefits to patient, providing alternatives to ED e.g. equipment, falls prevention advice and onward referral. More patients treated at home. Work needed to establish best practice guidelines. |
| *Published evaluations:*<br>**Paramedic in hospice team**<br>(Mountbatten Isle of Wight)<br>Singer (2021) | Conducts assessments, using experience of visiting patients within community and decides on course of action/referral or input required. Two paramedics integrated within community palliative care team. Paramedics give IV infusions to patients within their homes to help avoid clinical environments. Paramedics have extensive patient assessment and diagnostic skillsets added to by in-house training on catheterisation/advance care planning and end-of-life care supplementing care provided by nursing staff within the community team. Role remains in its infancy: hospice services work differently and therefore no set rules for paramedic scope of practice. |
| *Published evaluations*<br>*Case study: Hull Fall First*<br>*Going further for winter: Community-based falls response*<br>(2022) | Quantitative impact difficult to quantify because of how falls are recorded in health datasets, but following outcomes observed locally:<br>• Reduced number of people requiring A&E attendance because of a fall<br>• Reduced number of admissions because of rapid response and not having a long lie<br>• Increased follow up from therapy falls team<br>• Better patient experience for those having a fall. |
| *Case study: Community frailty response service*<br>McNamara (2020) | Describes expansion/adaptation of frailty response team to assess older people in their usual place of residence. Team had commenced weekend service to a limited area in February 2020. Following demand related to COVID-19 pandemic, service expanded and model of care adapted to offer 7-day service to entire catchment area. 592 patient reviews completed in first 105 days of operation with 43 patients transferred to hospital for further investigation/ management following assessment. |
| CCP: Rapid Response/See And Treat |  |
| **Case study: Falls Specialist Response Car**<br>Woodward & Procter (2016)<br>(Rapid Review)<br>https://www.healthylondon.org/wp-content/uploads/2017/11/Rapid-review-Rapid-response-teams-and-see-and-treat-models.pdf | In 2015/2016 team visited 1,821 patients and managed to keep 66.5% at home avoiding need for emergency ambulance, A&E attendance/admission. Demonstrated safe, economic and desirable intervention from perspective of patient/service so elderly frail fallers stay in their homes. Also wider impact on organisations, teams and individuals. Continuous measurement essential to developing service. Having data readily available helps to understand what makes a successful referral. Key to keeping referral number up is relationship with ambulance control room. Working on sustaining increase in referrals, aiming for average of 20/week kept at home. Engaging with similar schemes to spread model across LAS services. |
| *Additional evidence*<br>Mahase (2022) | NHS England to create a “community based falls response service” in all systems for people who have fallen at home, including in care homes, and will reduce “unwarranted variation in ambulance conveyance rates. (Mahase, 2022)” |

|  |
| --- |
| **Research Gaps** |
| Larger systematic evaluations warranted to establish utility and cost-effectiveness of point-of-care testing by ambulance services (Novak et al, 2022).<br>Potential need to review rapid response/see and treat services/initiatives in update to Woodward & Procter (2016) (Rapid Review) |
| **Research Priority:** Moderate - No/poor Systematic Review but plentiful UK research studies |

(Synthesis Gap)

##### CCP - Single Point Response

**Table 49.** Single Point Response: Definitions and Rationales.

| **Taxonomy heading** | **Brief description/definition** | **Mechanism for minimising winter pressures** |
| --- | --- | --- |
| CCP - Single Point Response | Coordination centres that seek to provide a single point of access to health and/or social care services and therefore speed up interactions with services. | Improved coordination of discharge planning and speedier discharge |
| Example interventions:<br>Glasgow City HSCP Complex Needs Service | Operates single point of access to vulnerable, transient, and high-risk population, providing diverse medical, health and social care interventions for individuals with multiple and complex health and social care needs. | Uses assertive outreach model to ensure appropriate assessment and level of response is in place relative to level of individual risk with aim of actively engaging “hard to reach” individuals with a focus on women and young people. |
| Example interventions:<br>Home, But Not Alone<br>Care Quality Commission. (2021). Collaboration in urgent and emergency care.<sup>7</sup> | Suffolk/North East Essex phone line open 9am - 5pm, 7 days/week, to help on topics including food, medication, loneliness and isolation. Provided practical support to vulnerable people, particularly regarding medicines, food supplies and transport to health and care services, and for other urgent/non-urgent needs. Health/council services created single point of access – food, prescriptions, transport or support, and all linked into medical services. | Offers single point of access with co-ordination of needs and faster response. |
| Example interventions: Hull Single Point of Access (Charles, 2018) | All referrals for CHCP services through single point of access, open to professionals, service users and carers. Referrals processed by trained support workers who triage according to urgency/complexity, decide what assessments are needed, and link them into services as required. Separate single point of access for social care services. | Coordinated response with prioritisation according to need. |
| Example interventions:<br>Open House | Open House community café to provide support for carers and others needing support. | Improved carer resilience and less dependence upon emergency medical services. |
| Example interventions:<br>Single point of access for social care | Offers first point of contact for all social care services | Improved coordination and easier navigation of social care services |

**Table 50.**
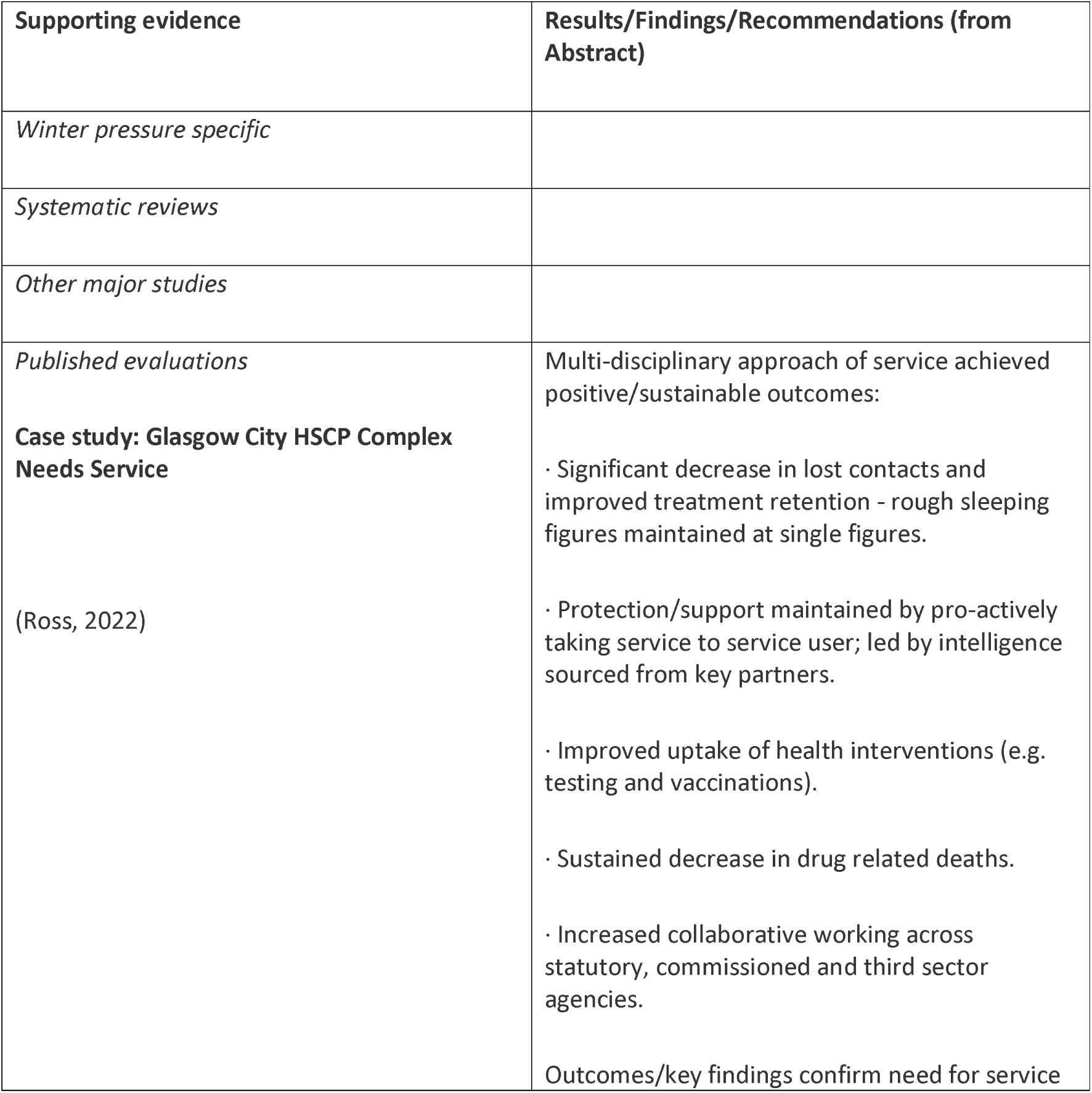
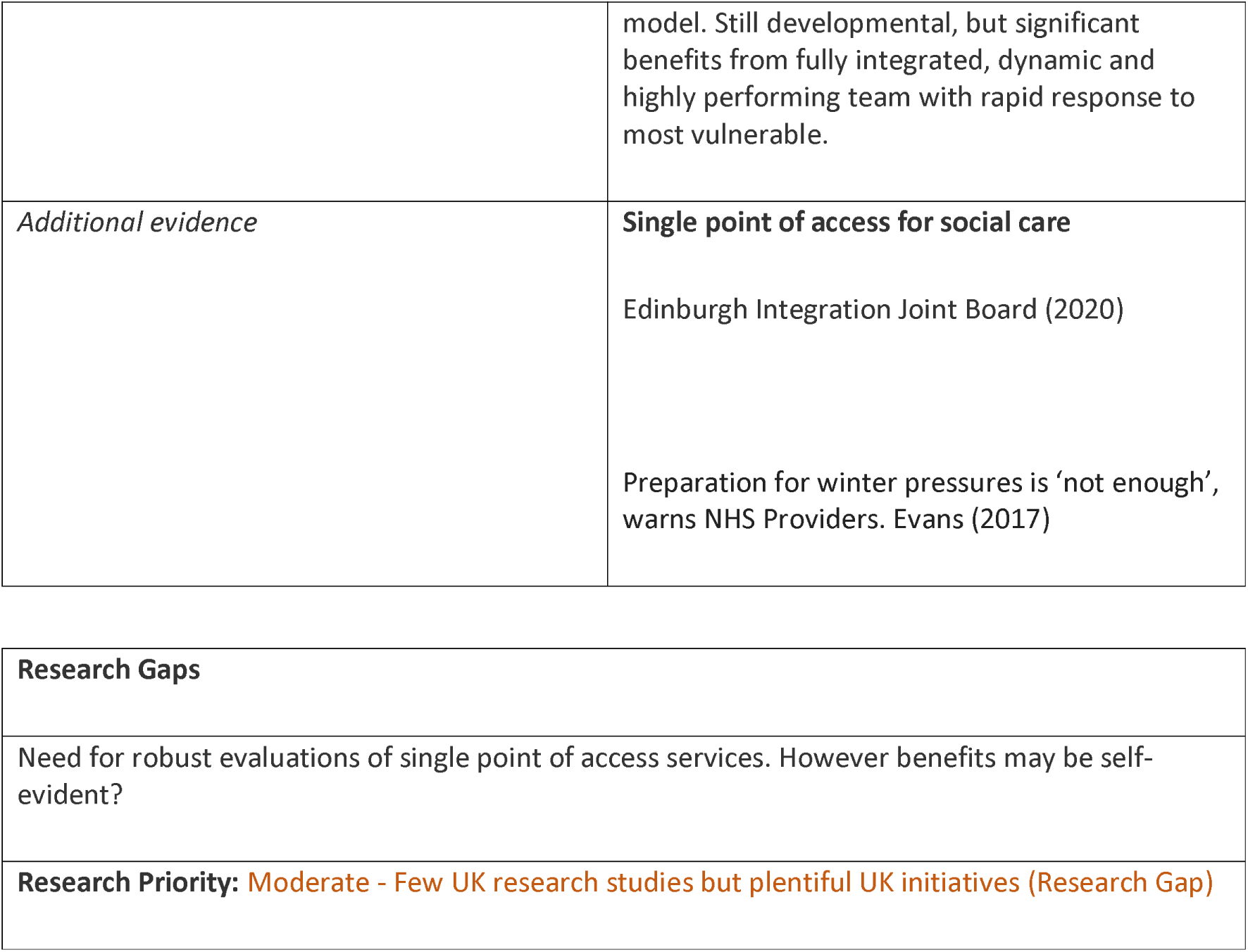
Single Point of Response: Interventions and Supporting Evidence.

| Supporting evidence | Results/Findings/Recommendations (from Abstract) |
| --- | --- |
| *Winter pressure specific* |  |
| *Systematic reviews* |  |
| *Other major studies* |  |
| *Published evaluations*<br>**Case study: Glasgow City HSCP Complex Needs Service**<br>(Ross, 2022) | Multi-disciplinary approach of service achieved positive/sustainable outcomes:<br>· Significant decrease in lost contacts and improved treatment retention - rough sleeping figures maintained at single figures.<br>· Protection/support maintained by pro-actively taking service to service user; led by intelligence sourced from key partners.<br>· Improved uptake of health interventions (e.g. testing and vaccinations).<br>· Sustained decrease in drug related deaths.<br>· Increased collaborative working across statutory, commissioned and third sector agencies.<br>Outcomes/key findings confirm need for service |
|  | model. Still developmental, but significant benefits from fully integrated, dynamic and highly performing team with rapid response to most vulnerable. |
| *Additional evidence* | **Single point of access for social care**<br>Edinburgh Integration Joint Board (2020)<br>Preparation for winter pressures is 'not enough', warns NHS Providers. Evans (2017) |

|  |
| --- |
| **Research Gaps** |
| Need for robust evaluations of single point of access services. However benefits may be self-evident? |
| **Research Priority:** Moderate - Few UK research studies but plentiful UK initiatives (Research Gap) |

##### CCP - Step Up Facilities

**Table 51.** Step Up Facilities: Definitions and Rationales.

| Taxonomy heading | Brief description/definition | Mechanism for minimising winter pressures |
| --- | --- | --- |
| CCP - Step Up Facilities | Typically provide care in a setting other than acute hospital. Time limited in nature, schemes aim to prevent admission to hospital, only advancing to hospital in cases of observed deterioration. Step up schemes can target similar patients to rapid response. However, normally users require more intensive care than could be provided in their own home. | Investment in ‘step-up’ community rehabilitation beds helps to relieve pressure on NHS beds and to flex capacity at times of especially high demand. Acute trusts need to plan effectively for sufficient acute beds as well as access to community beds to improve patient flow. |
| Example interventions:<br>**Step-up facilities based in primary care**<br>“Bridging the Care Gap” | Closely aligned to step-down facilities, services that step-up from primary care, prior to acute care admission: “Two innovative-minded GPs have set up pharmacy, dentist and training centre within the practice...so that people don’t have to travel everywhere to do something...” Focus on creating step-up facility on surgery land and using this to improve care for elderly. Housing recognised as a principal factor that leads to delayed discharges: “Resources in housing sector could be better used to support health and wellbeing.” | Enlarging the primary care offer to encompass step-up facilities means thinking more widely about environments in which people live and how partnership working with different agencies and sectors can support this. Innovative approaches to housing/health assist with workforce issues, e.g. including key-worker accommodation in housing development plans for healthier environments, and live-in carers and support workers. |

**Table 52.**
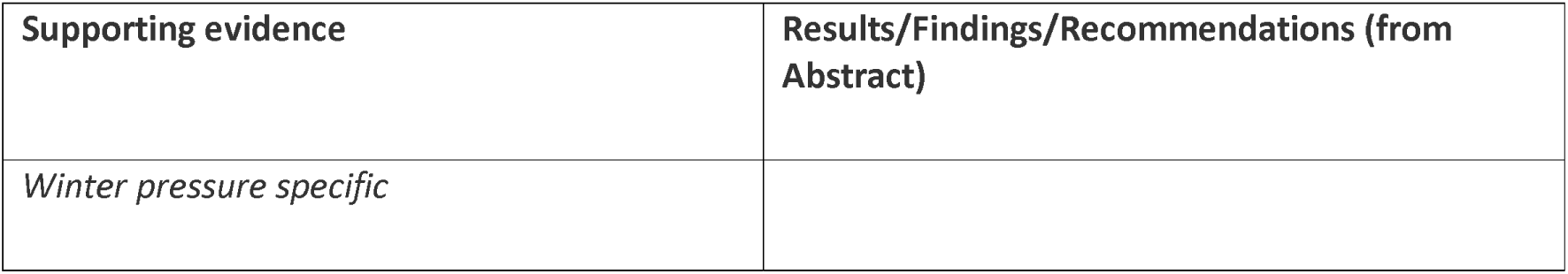
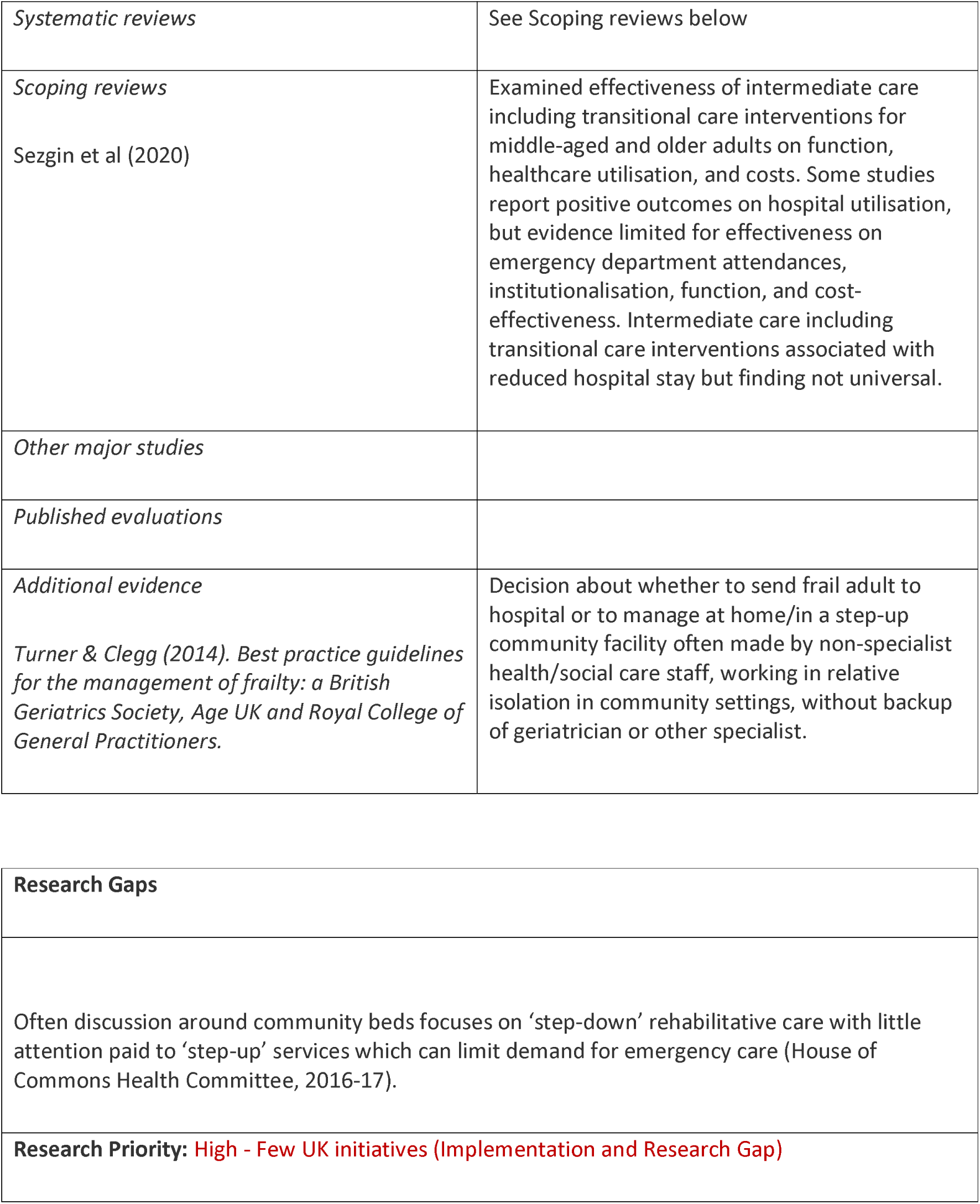
Step Up Facilities: Interventions and Supporting Evidence.

| Supporting evidence | Results/Findings/Recommendations (from Abstract) |
| --- | --- |
| Winter pressure specific |  |
| *Systematic reviews* | See Scoping reviews below |
| *Scoping reviews*<br>Sezgin et al (2020) | Examined effectiveness of intermediate care including transitional care interventions for middle-aged and older adults on function, healthcare utilisation, and costs. Some studies report positive outcomes on hospital utilisation, but evidence limited for effectiveness on emergency department attendances, institutionalisation, function, and cost-effectiveness. Intermediate care including transitional care interventions associated with reduced hospital stay but finding not universal. |
| *Other major studies* |  |
| *Published evaluations* |  |
| *Additional evidence*<br>Turner & Clegg (2014). *Best practice guidelines for the management of frailty: a British Geriatrics Society, Age UK and Royal College of General Practitioners.* | Decision about whether to send frail adult to hospital or to manage at home/in a step-up community facility often made by non-specialist health/social care staff, working in relative isolation in community settings, without backup of geriatrician or other specialist. |

|  |
| --- |
| **Research Gaps** |
| Often discussion around community beds focuses on ‘step-down’ rehabilitative care with little attention paid to ‘step-up’ services which can limit demand for emergency care (House of Commons Health Committee, 2016-17). |
| **Research Priority: High - Few UK initiatives (Implementation and Research Gap)** |

#### Changing Community Provision – Alternate Delivery Site

Findings are presented in detail in Tables 53 to 62 below and associated summary paragraphs.

##### CCP - Care Homes

**Table 53.**
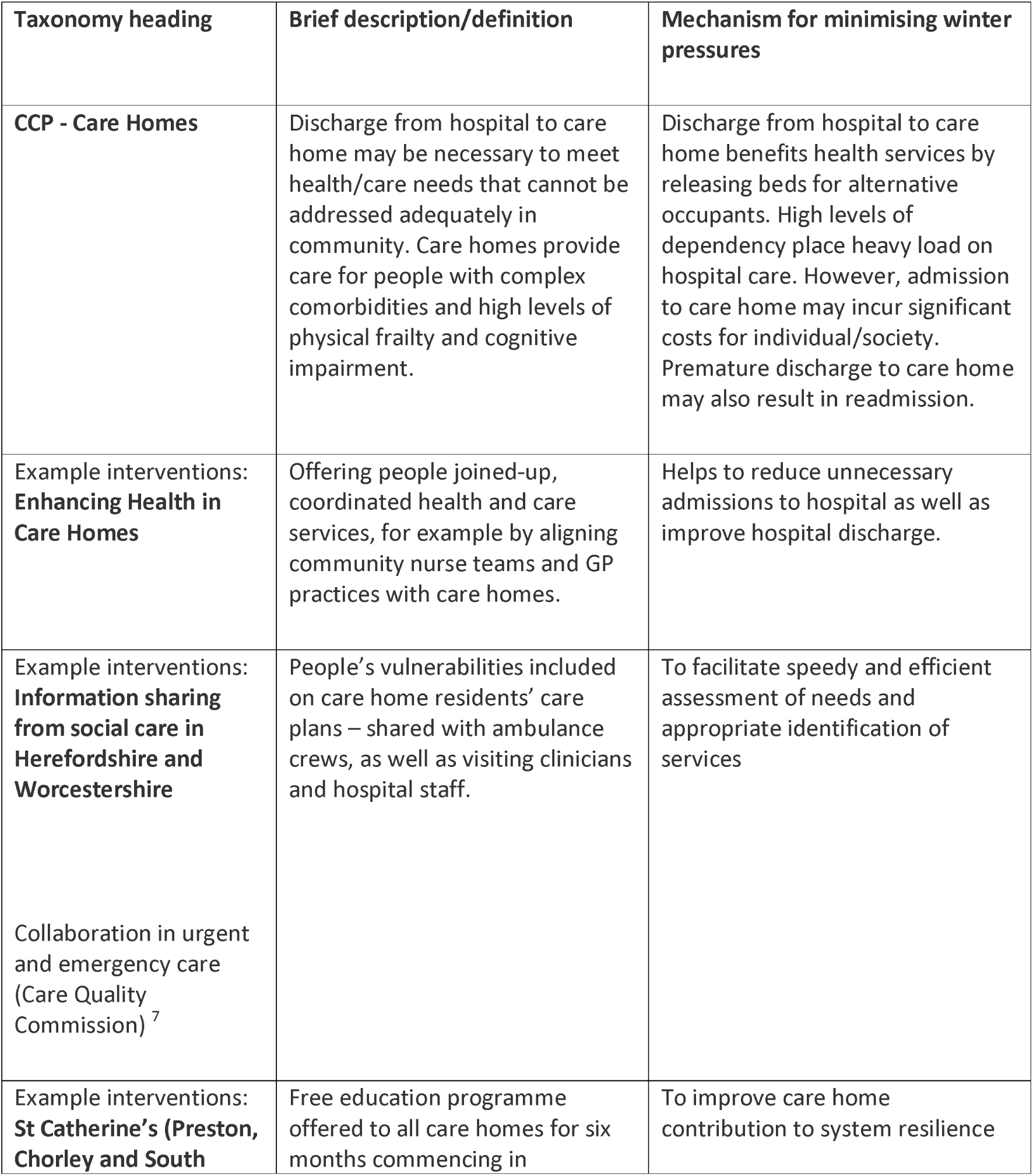
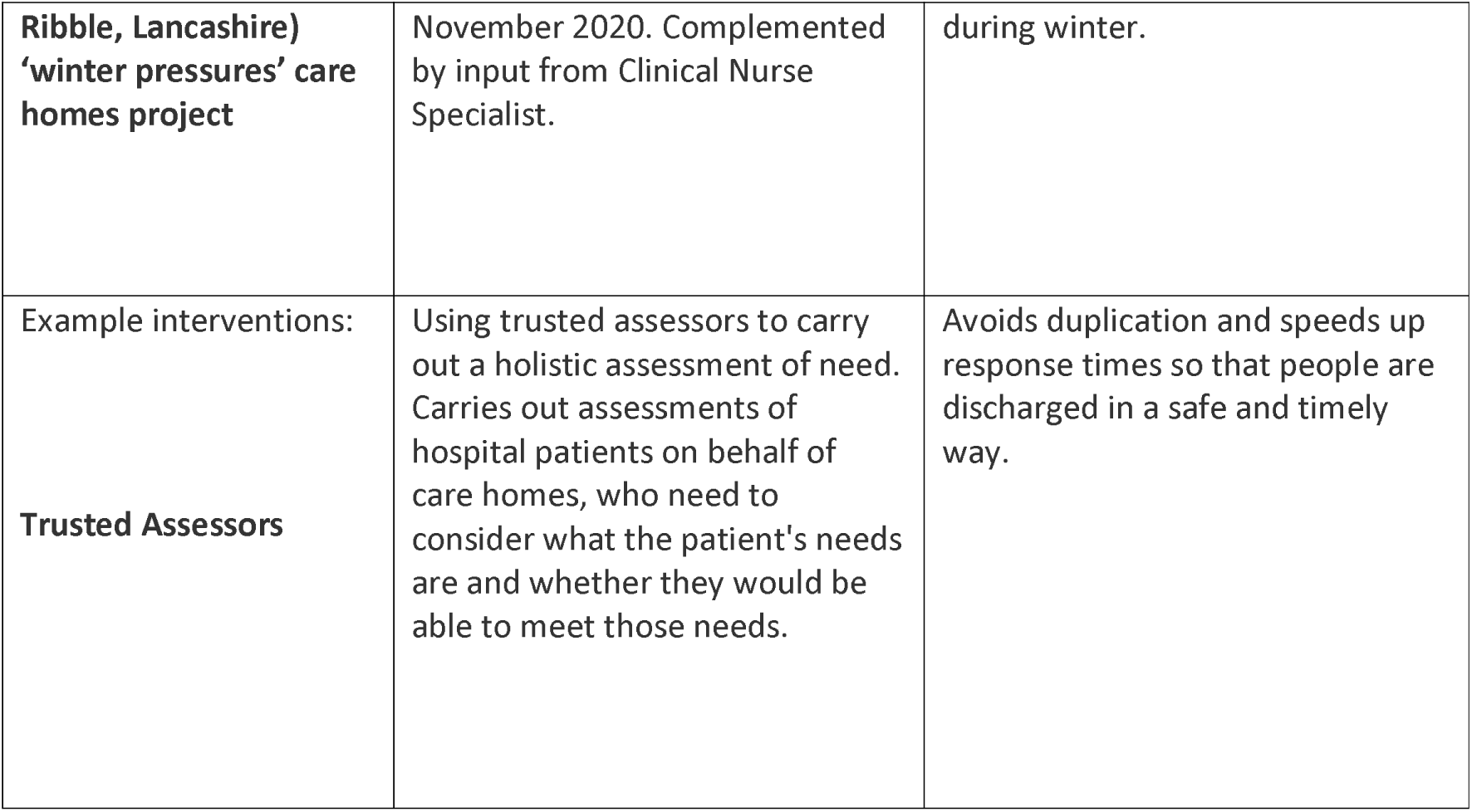
Care Homes: Definitions and Rationales.

**Table 54.**
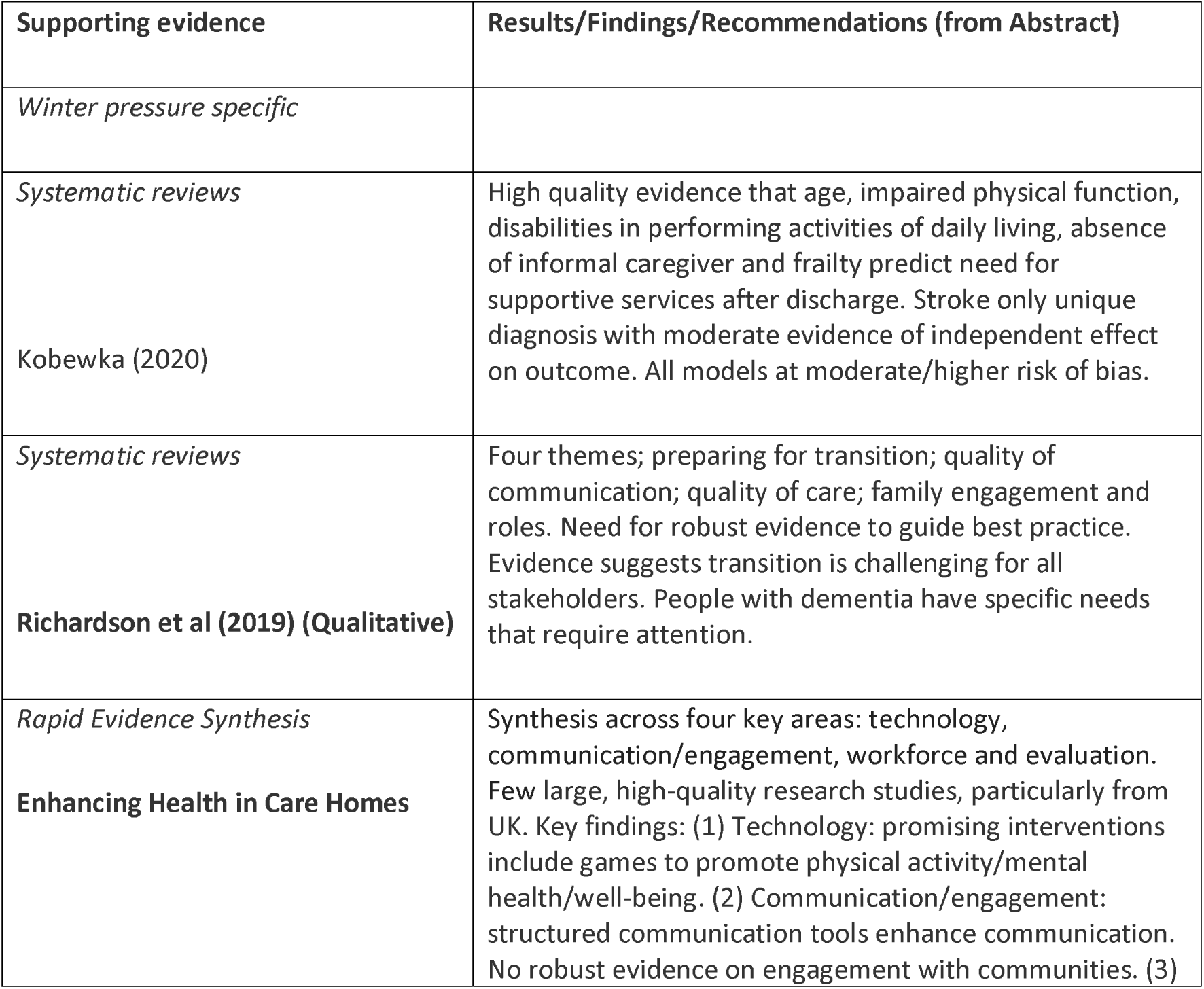
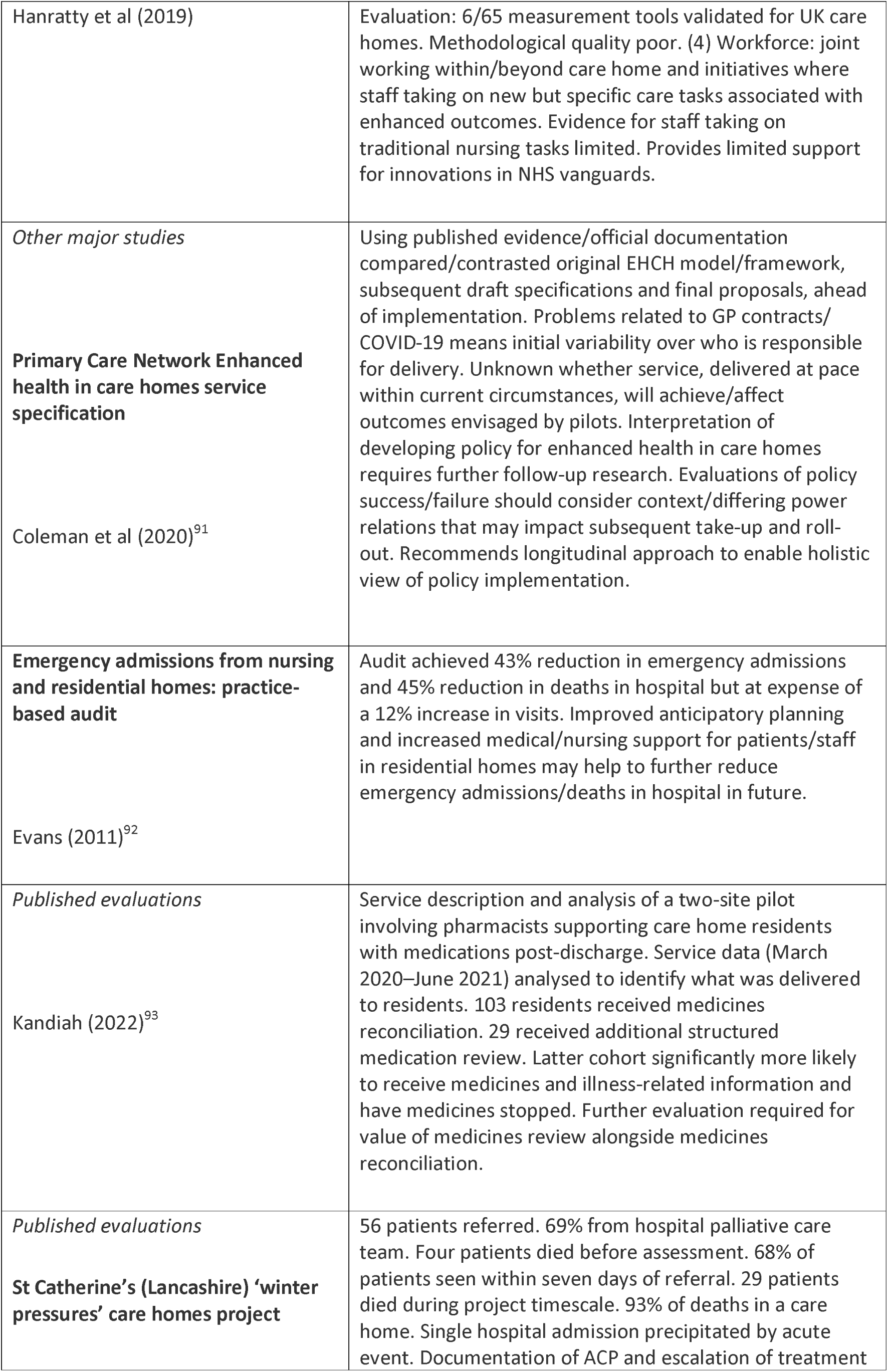
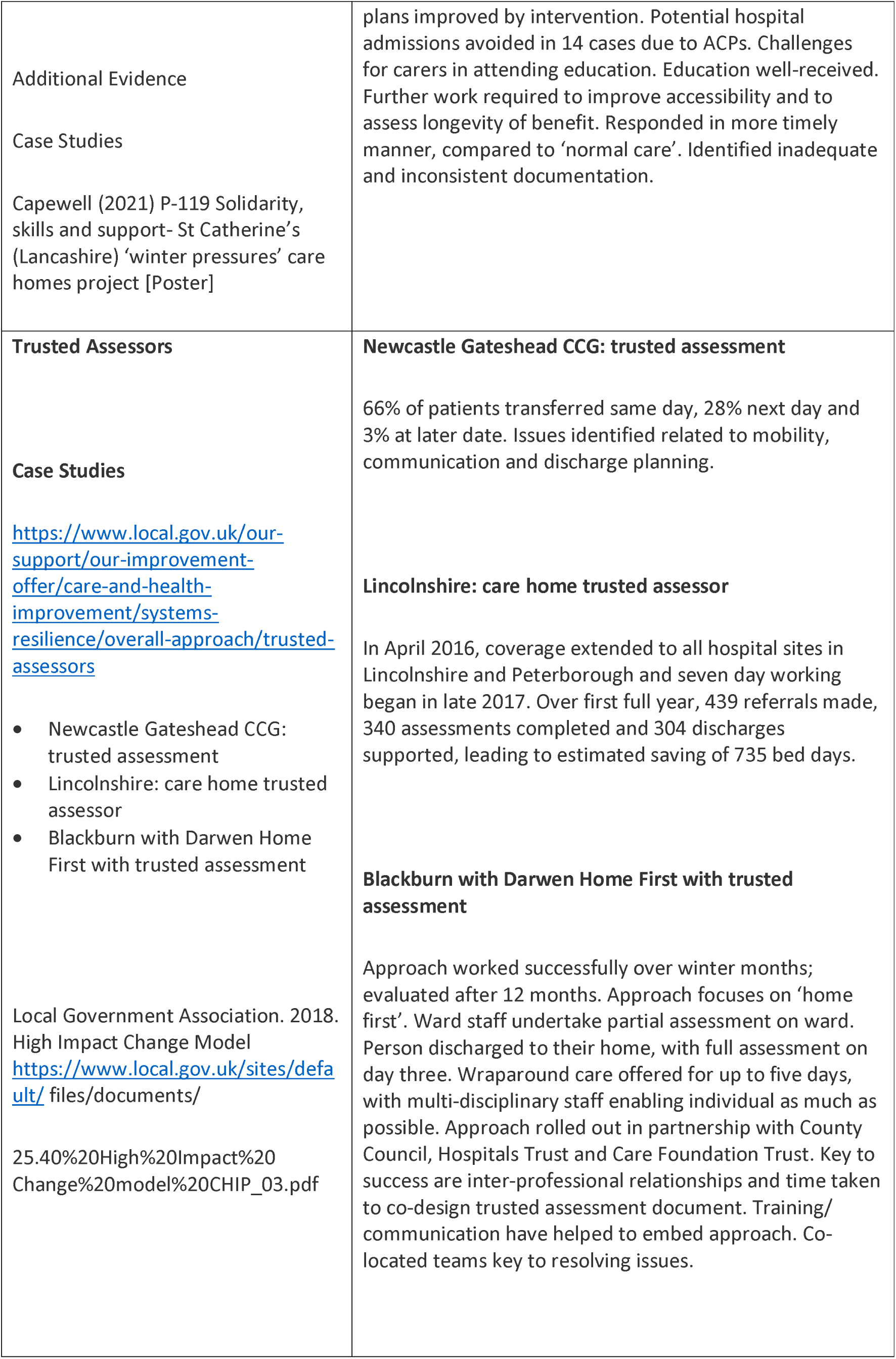
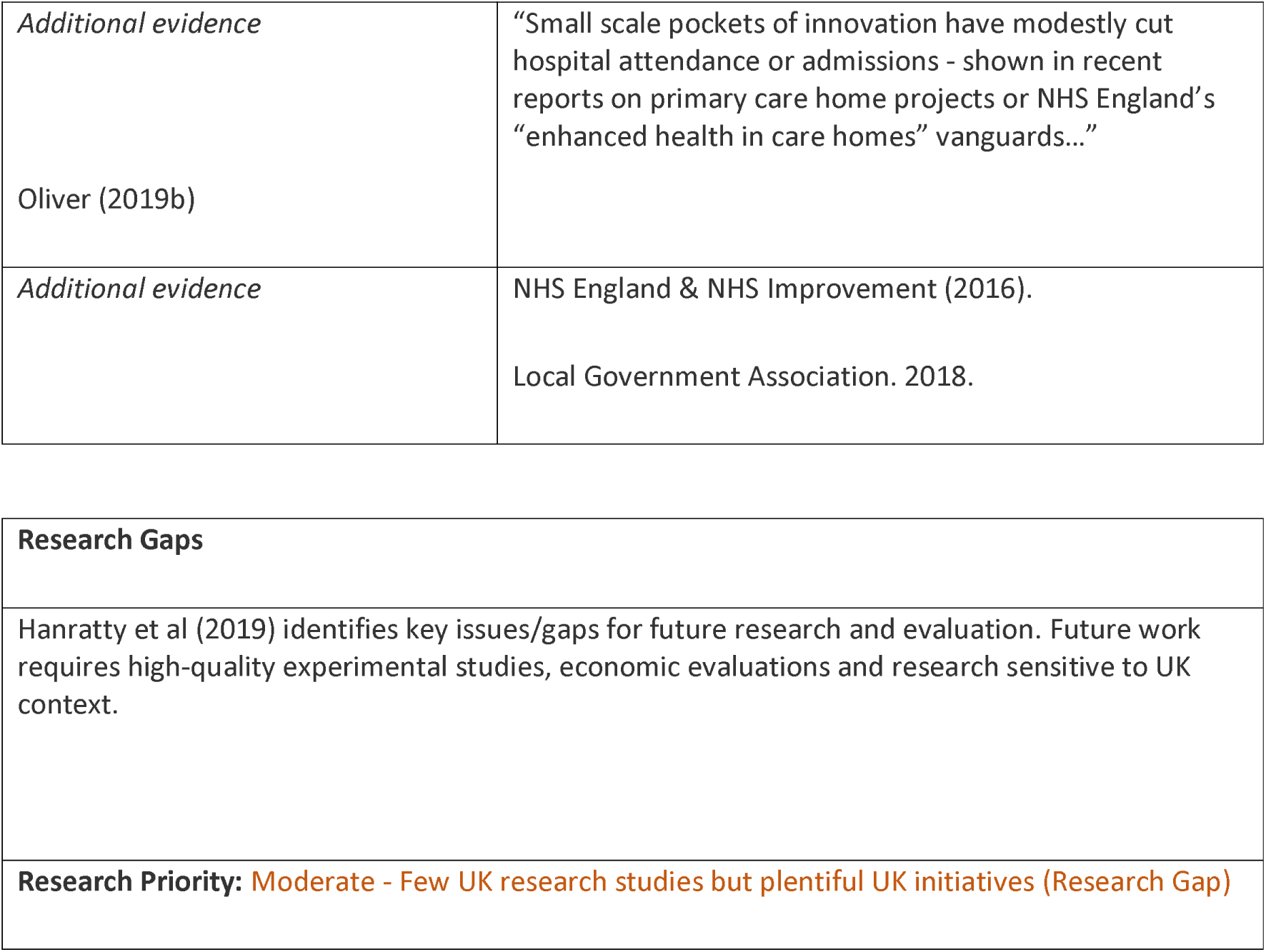
Care Homes: Interventions and Supporting Evidence.

| **Supporting evidence** | **Results/Findings/Recommendations (from Abstract)** |
| --- | --- |
| *Winter pressure specific* |  |
| *Systematic reviews*<br>Kobewka (2020) | High quality evidence that age, impaired physical function, disabilities in performing activities of daily living, absence of informal caregiver and frailty predict need for supportive services after discharge. Stroke only unique diagnosis with moderate evidence of independent effect on outcome. All models at moderate/higher risk of bias. |
| *Systematic reviews*<br>**Richardson et al (2019) (Qualitative)** | Four themes; preparing for transition; quality of communication; quality of care; family engagement and roles. Need for robust evidence to guide best practice. Evidence suggests transition is challenging for all stakeholders. People with dementia have specific needs that require attention. |
| *Rapid Evidence Synthesis*<br>**Enhancing Health in Care Homes** | Synthesis across four key areas: technology, communication/engagement, workforce and evaluation. Few large, high-quality research studies, particularly from UK. Key findings: (1) Technology: promising interventions include games to promote physical activity/mental health/well-being. (2) Communication/engagement: structured communication tools enhance communication. No robust evidence on engagement with communities. (3) |
| Hanratty et al (2019) | Evaluation: 6/65 measurement tools validated for UK care homes. Methodological quality poor. (4) Workforce: joint working within/beyond care home and initiatives where staff taking on new but specific care tasks associated with enhanced outcomes. Evidence for staff taking on traditional nursing tasks limited. Provides limited support for innovations in NHS vanguards. |
| *Other major studies*<br>**Primary Care Network Enhanced health in care homes service specification**<br>Coleman et al (2020)<sup>91</sup> | Using published evidence/official documentation compared/contrasted original EHCH model/framework, subsequent draft specifications and final proposals, ahead of implementation. Problems related to GP contracts/ COVID-19 means initial variability over who is responsible for delivery. Unknown whether service, delivered at pace within current circumstances, will achieve/affect outcomes envisaged by pilots. Interpretation of developing policy for enhanced health in care homes requires further follow-up research. Evaluations of policy success/failure should consider context/differing power relations that may impact subsequent take-up and roll-out. Recommends longitudinal approach to enable holistic view of policy implementation. |
| **Emergency admissions from nursing and residential homes: practice-based audit**<br>Evans (2011)<sup>92</sup> | Audit achieved 43% reduction in emergency admissions and 45% reduction in deaths in hospital but at expense of a 12% increase in visits. Improved anticipatory planning and increased medical/nursing support for patients/staff in residential homes may help to further reduce emergency admissions/deaths in hospital in future. |
| *Published evaluations*<br>Kandiah (2022)<sup>93</sup> | Service description and analysis of a two-site pilot involving pharmacists supporting care home residents with medications post-discharge. Service data (March 2020–June 2021) analysed to identify what was delivered to residents. 103 residents received medicines reconciliation. 29 received additional structured medication review. Latter cohort significantly more likely to receive medicines and illness-related information and have medicines stopped. Further evaluation required for value of medicines review alongside medicines reconciliation. |
| *Published evaluations*<br>**St Catherine's (Lancashire) 'winter pressures' care homes project** | 56 patients referred. 69% from hospital palliative care team. Four patients died before assessment. 68% of patients seen within seven days of referral. 29 patients died during project timescale. 93% of deaths in a care home. Single hospital admission precipitated by acute event. Documentation of ACP and escalation of treatment |
| Additional Evidence<br>Case Studies<br>Capewell (2021) P-119 Solidarity, skills and support- St Catherine's (Lancashire) 'winter pressures' care homes project [Poster] | plans improved by intervention. Potential hospital admissions avoided in 14 cases due to ACPs. Challenges for carers in attending education. Education well-received. Further work required to improve accessibility and to assess longevity of benefit. Responded in more timely manner, compared to 'normal care'. Identified inadequate and inconsistent documentation. |
| **Trusted Assessors**<br>**Case Studies**<br>https://www.local.gov.uk/our-support/our-improvement-offer/care-and-health-improvement/systems-resilience/overall-approach/trusted-assessors<br>• Newcastle Gateshead CCG: trusted assessment<br>• Lincolnshire: care home trusted assessor<br>• Blackburn with Darwen Home First with trusted assessment<br>Local Government Association. 2018. High Impact Change Model<br>https://www.local.gov.uk/sites/default/files/documents/25.40%20High%20Impact%20Change%20model%20CHIP_03.pdf | **Newcastle Gateshead CCG: trusted assessment**<br>66% of patients transferred same day, 28% next day and 3% at later date. Issues identified related to mobility, communication and discharge planning.<br>**Lincolnshire: care home trusted assessor**<br>In April 2016, coverage extended to all hospital sites in Lincolnshire and Peterborough and seven day working began in late 2017. Over first full year, 439 referrals made, 340 assessments completed and 304 discharges supported, leading to estimated saving of 735 bed days.<br>**Blackburn with Darwen Home First with trusted assessment**<br>Approach worked successfully over winter months; evaluated after 12 months. Approach focuses on 'home first'. Ward staff undertake partial assessment on ward. Person discharged to their home, with full assessment on day three. Wraparound care offered for up to five days, with multi-disciplinary staff enabling individual as much as possible. Approach rolled out in partnership with County Council, Hospitals Trust and Care Foundation Trust. Key to success are inter-professional relationships and time taken to co-design trusted assessment document. Training/communication have helped to embed approach. Co-located teams key to resolving issues. |
| *Additional evidence*<br>Oliver (2019b) | “Small scale pockets of innovation have modestly cut hospital attendance or admissions - shown in recent reports on primary care home projects or NHS England’s “enhanced health in care homes” vanguards...” |
| *Additional evidence* | NHS England & NHS Improvement (2016).<br>Local Government Association. 2018. |

|  |
| --- |
| **Research Gaps** |
| Hanratty et al (2019) identifies key issues/gaps for future research and evaluation. Future work requires high-quality experimental studies, economic evaluations and research sensitive to UK context. |
| **Research Priority:** Moderate - Few UK research studies but plentiful UK initiatives (Research Gap) |

##### CCP - Community Teams

**Table 55.** Community Teams: Definitions and Rationales.

| Taxonomy heading | Brief description/definition | Mechanism for minimising winter pressures |
| --- | --- | --- |
| CCP - Community Teams | Provide holistic care to patients, carers and families within community setting. Multi-disciplinary teams (e.g. Team Leader, Clinical Leads, Nurses, Physiotherapists, Occupational therapists, Care Co-ordinators, Support Workers, Administrators). Support housebound or those whose condition requires nursing or therapy at home. Also provide clinician-led clinics in community. | Offers alternative venue to some hospital care. |
| Example interventions:<br>**Multidisciplinary community teams** | Coordinated discharge planning based on joint assessment processes and protocols and on shared/agreed responsibilities. | Promotes effective discharge and positive outcomes for patients |

**Table 56.**
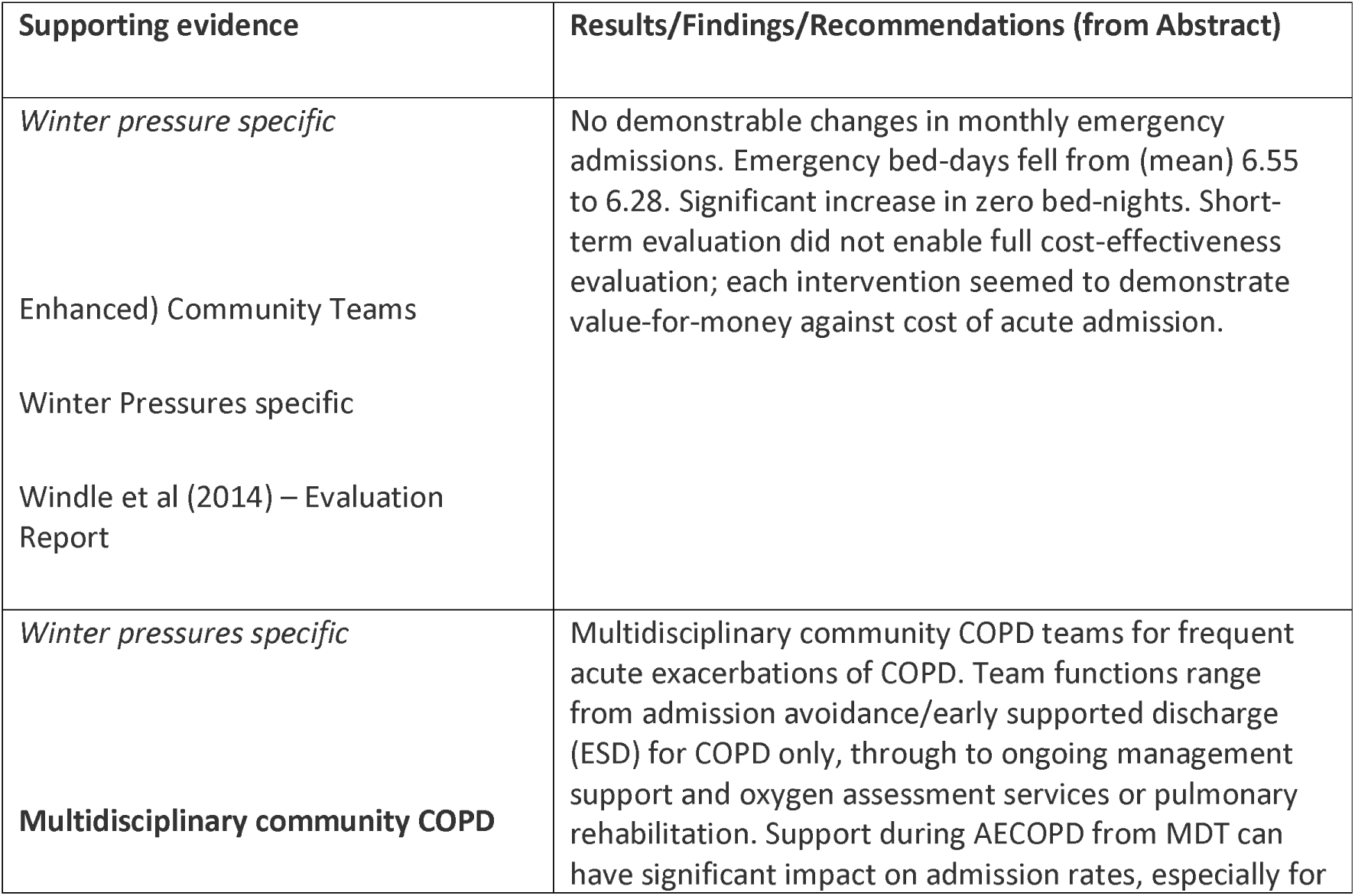
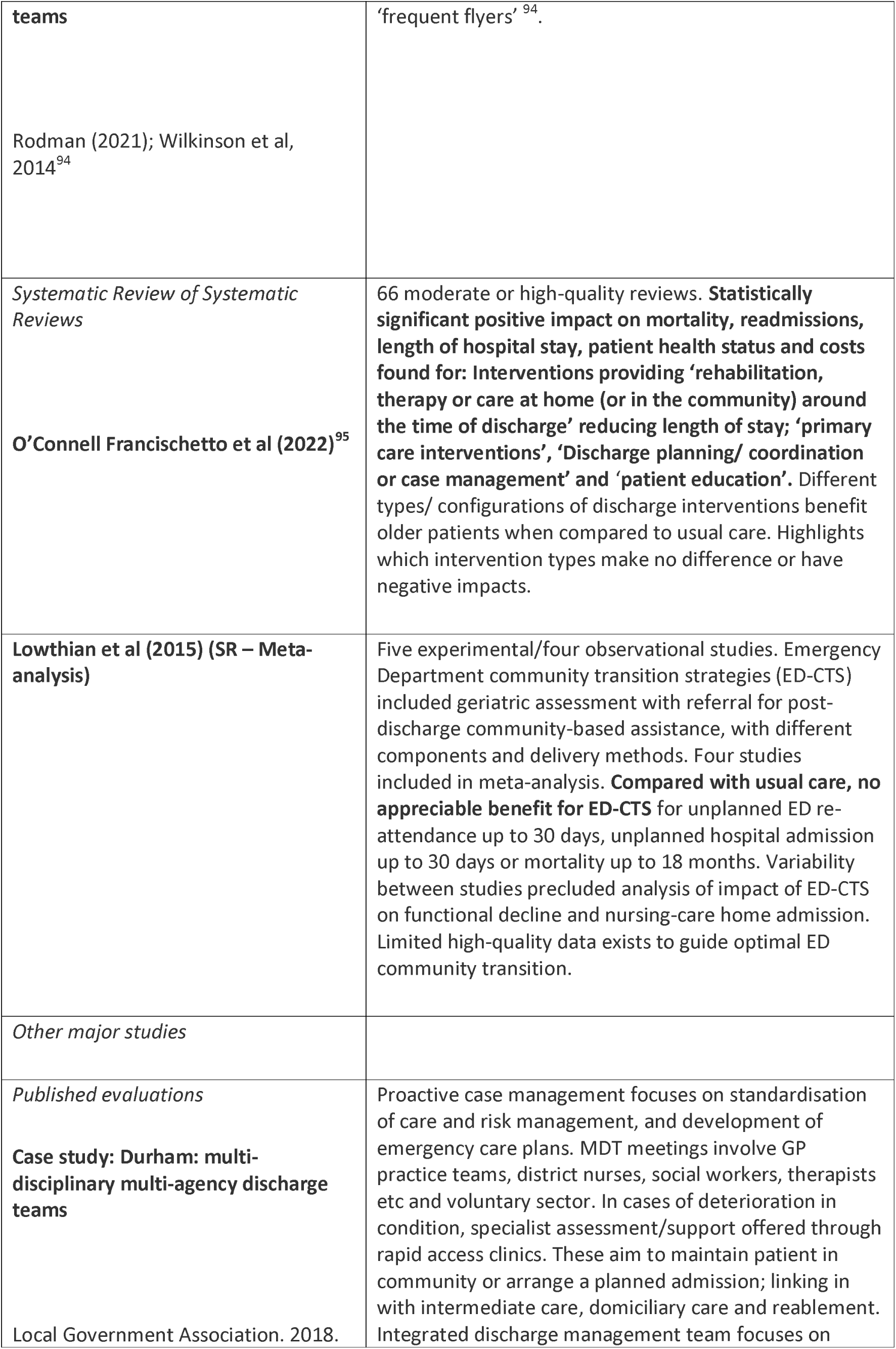
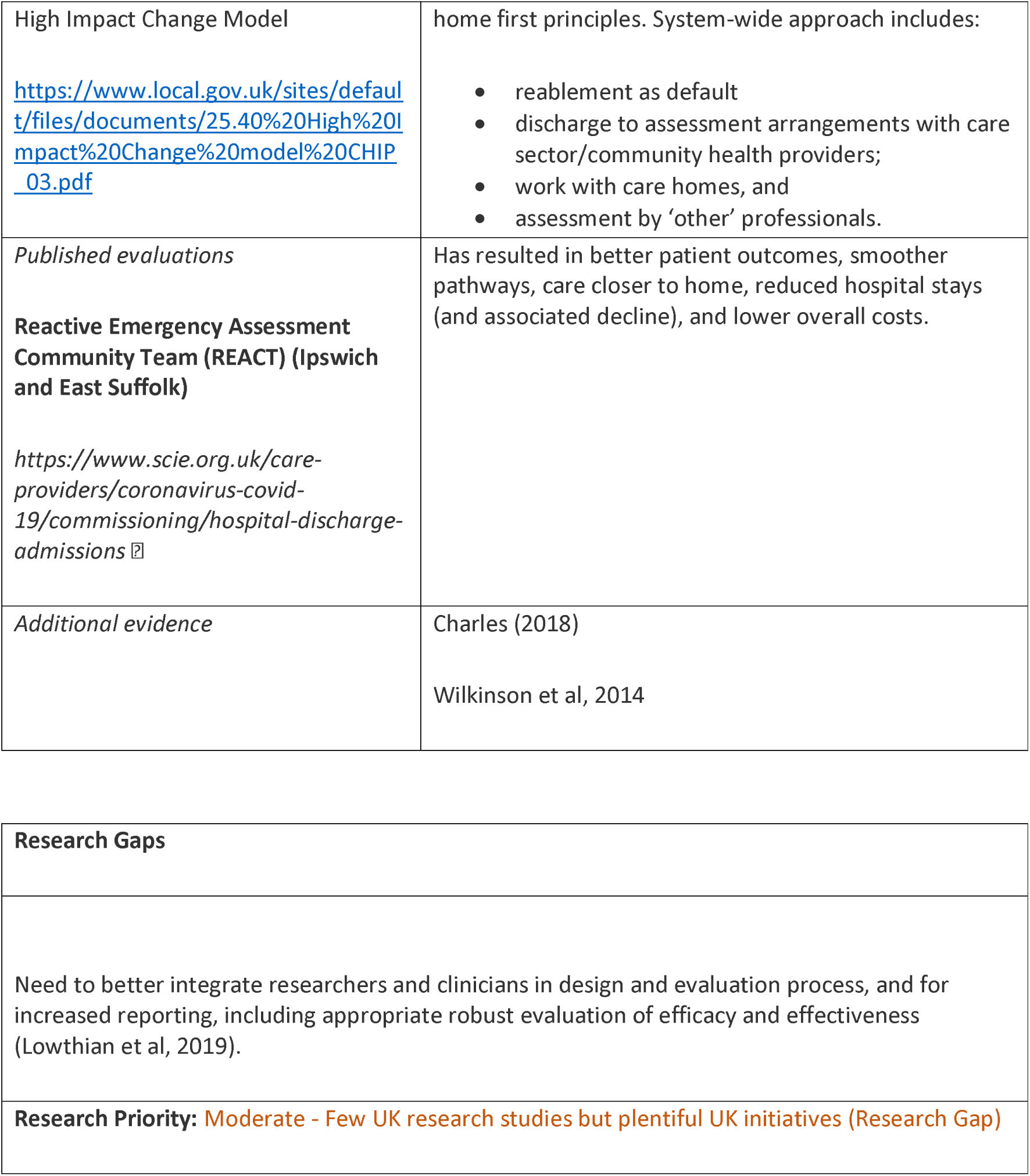
Community Teams: Interventions and Supporting Evidence.

| Supporting evidence | Results/Findings/Recommendations (from Abstract) |
| --- | --- |
| *Winter pressure specific*<br>Enhanced) Community Teams<br>Winter Pressures specific<br>Windle et al (2014) – Evaluation Report | No demonstrable changes in monthly emergency admissions. Emergency bed-days fell from (mean) 6.55 to 6.28. Significant increase in zero bed-nights. Short-term evaluation did not enable full cost-effectiveness evaluation; each intervention seemed to demonstrate value-for-money against cost of acute admission. |
| *Winter pressures specific*<br>**Multidisciplinary community COPD** | Multidisciplinary community COPD teams for frequent acute exacerbations of COPD. Team functions range from admission avoidance/early supported discharge (ESD) for COPD only, through to ongoing management support and oxygen assessment services or pulmonary rehabilitation. Support during AECOPD from MDT can have significant impact on admission rates, especially for |
| **teams**<br>Rodman (2021); Wilkinson et al, 2014<sup>94</sup> | ‘frequent flyers’<sup>94</sup>. |
| *Systematic Review of Systematic Reviews*<br>**O’Connell Francischetto et al (2022)<sup>95</sup>** | 66 moderate or high-quality reviews. **Statistically significant positive impact on mortality, readmissions, length of hospital stay, patient health status and costs found for: Interventions providing ‘rehabilitation, therapy or care at home (or in the community) around the time of discharge’ reducing length of stay; ‘primary care interventions’, ‘Discharge planning/ coordination or case management’ and ‘patient education’.** Different types/ configurations of discharge interventions benefit older patients when compared to usual care. Highlights which intervention types make no difference or have negative impacts. |
| **Lowthian et al (2015) (SR – Meta-analysis)** | Five experimental/four observational studies. Emergency Department community transition strategies (ED-CTS) included geriatric assessment with referral for post-discharge community-based assistance, with different components and delivery methods. Four studies included in meta-analysis. **Compared with usual care, no appreciable benefit for ED-CTS** for unplanned ED re-attendance up to 30 days, unplanned hospital admission up to 30 days or mortality up to 18 months. Variability between studies precluded analysis of impact of ED-CTS on functional decline and nursing-care home admission. Limited high-quality data exists to guide optimal ED community transition. |
| *Other major studies* |  |
| *Published evaluations*<br>**Case study: Durham: multi-disciplinary multi-agency discharge teams**<br>Local Government Association. 2018. | Proactive case management focuses on standardisation of care and risk management, and development of emergency care plans. MDT meetings involve GP practice teams, district nurses, social workers, therapists etc and voluntary sector. In cases of deterioration in condition, specialist assessment/support offered through rapid access clinics. These aim to maintain patient in community or arrange a planned admission; linking in with intermediate care, domiciliary care and reablement. Integrated discharge management team focuses on |
| High Impact Change Model<br>https://www.local.gov.uk/sites/default/files/documents/25.40%20High%20Impact%20Change%20model%20CHIP_03.pdf | home first principles. System-wide approach includes:<br>• reablement as default<br>• discharge to assessment arrangements with care sector/community health providers;<br>• work with care homes, and<br>• assessment by 'other' professionals. |
| *Published evaluations*<br>**Reactive Emergency Assessment Community Team (REACT) (Ipswich and East Suffolk)**<br>https://www.scie.org.uk/care-providers/coronavirus-covid-19/commissioning/hospital-discharge-admissions | Has resulted in better patient outcomes, smoother pathways, care closer to home, reduced hospital stays (and associated decline), and lower overall costs. |
| *Additional evidence* | Charles (2018)<br>Wilkinson et al, 2014 |

|  |
| --- |
| **Research Gaps** |
| Need to better integrate researchers and clinicians in design and evaluation process, and for increased reporting, including appropriate robust evaluation of efficacy and effectiveness (Lowthian et al, 2019). |
| **Research Priority:** Moderate - Few UK research studies but plentiful UK initiatives (Research Gap) |

##### CCP - Home Care

**Table 57.** Home Care: Definitions and Rationales.

| Taxonomy heading | Brief description/definition | Mechanism for minimising winter pressures |
| --- | --- | --- |
| CCP - Home Care | Health care or supportive care provided by a professional caregiver in individual home where patient/client is living, as opposed to care in group accommodations (e.g. clinics/nursing home). Also known as domiciliary care, social care or in-home care. Comprises diverse activities, especially paramedical aid by nurses and assistance in daily living for ill, disabled or elderly people. | Offers alternative venue to some hospital care. Clients receiving home health care may incur lower costs, receive equal to better care, and have increased satisfaction in contrast to other settings |
| Example interventions:<br>**Multidisciplinary community teams** | Coordinated discharge planning based on joint assessment processes and protocols and on shared and agreed responsibilities. | Promotes effective discharge and positive outcomes for patients |
| Ambulance Service Home Visits<br>Collaboration in Urgent and Emergency Care (2022) <sup>7</sup>; Halliwell (2022) | GPs at Northamptonshire practice had regular calls with ambulance service and worked together on patient cases to avoid unnecessary ED attendance. Ambulance service carried out visits to people's homes for GP practice, in liaison with GPs, helping people to avoid going into hospital. | Potential to reduce hospital admissions, ambulance calls and ED self-referrals. |
| Example interventions:<br>**Dolphin Home from Hospital service** | Aims to provide short-term post-discharge care at a more intensive level than normally provided by professionals such as District Nursing. Generally delivered in user's own home and led by nursing staff, sometimes with input from medical/allied health professionals (NICE Guideline 94). | Transfers care from hospital setting to home setting |
| Example interventions:<br>**Follow-up via** | Early assessment identifies patients for discharge with increased outpatient support, with follow-up | Transfers ongoing follow up and monitoring from hospital setting to |
| **home visits** | via home visits, telephone consultation or remote monitoring. | home setting |
| Example interventions:<br>**Home Visiting Services** | Visits (undertaken by paramedics or qualified advanced nurse practitioners) scheduled through the day (rather than saving them for lunch & evening slots) | Potential to reduce hospital admissions, ambulance calls and ED self-referrals. |
| Example interventions:<br>Hospital to a Healthier Home (H2HH) | Aims to support vulnerable older people by making their homes safe, warm and more accessible. | Aims for older people to be safely and more quickly discharged from hospitals to their homes and to prevent them being re-admitted. |

**Table 58.** Home Care: Interventions and Supporting Evidence.

| **Supporting evidence** | **Results/Findings/Recommendations (from Abstract)** |
| --- | --- |
| *Winter pressure specific* | None specified |
| *Systematic reviews*<br>*O'Connell Francischetto et al (2022)<sup>95</sup>*<br>**(SR of Systematic Reviews)** | Statistically significant positive impact on mortality, readmissions, length of hospital stay, patient health status and costs found for: Interventions providing 'rehabilitation, therapy or care at home (or in the community) around the time of discharge' reducing length of stay; 'primary care interventions', 'Discharge planning/ coordination or case management' and 'patient education'. |
| *Systematic reviews*<br>**Home-based intervention (HBI) (education and support) for heart failure vs. usual care**<br>Malik et al (2019)<br>(Meta-analysis) | 17 articles (3214 patients). HBI associated with reduced readmission rates and mortality (22 and 16% respectively; $p < 0.05$). Benefit of HBI increases significantly with longer follow-up. HBI in form of education and support significantly reduces readmission rates and improves survival of HF patients. HBI to be considered in discharge planning of HF patients. |
| *Systematic reviews* | 71 articles. Most interventions improved satisfaction but interventions variably reduced use. Interventions |
| **Hamline et al (2018)**<br>**(Systematic review)**<br>**[Paediatric population]** | focused on follow-up care, discharge planning, teach back-based parental education, and contingency planning associated with reduced use across patient groups. Bundled care coordination/family engagement interventions associated with lower use in patients with chronic illnesses and neonates. Variability limits findings/reduced generalizability. Highlights utility of paediatric discharge bundle in reducing health care use. |
| *Systematic reviews*<br>**Allen et al. (2017)**<br>**[Qualitative Meta-synthesis]** | Users' experience of discharge and transitional care is social process of 'negotiation and navigation of independence (older people/ carers), or dependence (health providers).' Users engaged in negotiation and navigation through interrogative strategies of questioning, discussion, information provision, information seeking, assessment, and translation. Themes reflected care integration that facilitated/ lack of care integration that constrained, users' experiences of negotiation and navigation of independence/dependence. |
| **Van Spall et al. (2017) [Systematic review and network meta-analysis]** | Nurse home visits/disease management clinics decrease all-cause mortality after hospitalization for HF. Along with nurse case management, they reduce all-cause readmissions. No significant difference in comparative effectiveness. Services reduce healthcare system costs to varying degrees. |
| *Systematic reviews*<br>**Feltner et al. (2014)**<br>**[Systematic review and meta-analysis]** | Home Visit programmes and MDC-Heart Failure clinics reduced all-cause readmission and mortality. These interventions should receive greatest consideration by systems or providers seeking to implement transitional care interventions for persons with Heart Failure |
| *Other major studies* |  |
| *Published evaluations*<br>**Dolphin Home from Hospital service** | Most cases create savings in health professional time, even disregarding hospital bed costs. Cost of care at home excluded from evaluation together with potentially high cost of clients entering long-term residential care when unable to return home. |
| *(Mackintosh, 2016)* |  |
| **Case studies**<br>**Hospital to a Healthier Home (H2HH)**<br>Intervention group: Care & Repair Cymru (C & RC) service, home advice and modification service vs. control group who do not.<br>**Published evaluations**<br>Hollinghurst (2020)<br>Wolton (2018)<br>Care & Repair<br>https://www.careandrepair.org.uk/ | Intervention group had increased risk of care home admission at 1-, 3- and 5-years for those classified as fit and mildly frail, but reduced risk of care home admission for moderately and severely frail individuals. C & RC service helped to prevent care home admissions for moderately/severely frail individuals. Wolton (2018) used mixed methods to examine interactions between Caseworkers across Care & Repair Cymru, their professional links and relationships with clients. Context in which Caseworkers operate (agency location/issues encountered when delivering services) influences how they link with others. |
| *Published evaluations*<br>**@home team St Thomas's or King's College Hospital**<br>**(Pickstone & Lee, 2019)** | 1084 patients referred to @home team in a 3-month period with 755 (72%) referrals. 387 local ED attendances prevented (298 from King's College Hospital and 89 from St Thomas's Hospital ED). Over same time period, King's College Hospital had 71,688 ED attendances; St Thomas's ED had 48,030 attendances. Although @home team reduces small number of ED attendances each month (1 in 300), number not enough to make significant impact on average performance against 4-h target at local EDs alone. |
| *Additional evidence*<br>**Follow-up home visits**<br>*Karlsson et al (2019)* | Follow-up home visits by nurses can relieve patient anxiety, as patients are often unsure of next steps for medication/care. According to nurses, visits created trust in the nurse-patient relationship and ensured patient safety. Follow-up visits soon after discharge from hospital should become routine nursing, especially for older people with multi-morbidity. |
| *Additional evidence* | "Small scale pockets of innovation have modestly cut hospital attendance or admissions - shown in recent reports on primary care home projects" |
| Oliver (2019b) |  |

| Research Gaps |
| --- |
| In paediatric population, coordinating follow-up, discharge planning, teach back-based parental education, and contingency planning are potential foci for future efforts to improve hospital-to-home transitions (Hamline et al (2018)).<br>Understanding, defining and measuring the components of homecare and homecare outcomes. Requires research to: (i) identify and define the components, or active ingredients, of homecare associated with user outcomes and experiences; and (ii) identify and agree on outcomes which evaluative research should include (i.e. core outcomes set)<br>O'Rourke & Beresford (2022) have identified comprehensive research priorities for home care including:<br>• Mapping & understanding the homecare population, providers and workforce.<br>• Public sector funding - cost-effectiveness of greater public investment in homecare. Homecare compared to other care options - whether type of care (e.g. homecare vs. residential care vs. supported care) affects outcomes and experiences. Includes preferences for place of care<br>• Timely engagement with homecare. Includes whether there are reliable predictors (or early warning signs) of the need for social care, the effectiveness and cost-effectiveness of 'pre-emptive' or early use of homecare and how to shift public understanding of homecare<br>• Comparing different homecare models in terms of outcomes, resource use/costs, process and experience.<br>• Housing and homecare: role of housing factors (e.g. physical layout, location and<br>• Technology: supporting the delivery of care through digitisation of care records and its impacts on the quality and safety of care.<br>**Research Priority:** Moderate - Few UK research studies but plentiful UK initiatives (Research Gap) |

##### CCP – Hospital At Home

**Table 59.** Hospital At Home: Definitions and Rationales.

| Taxonomy heading | Brief description/definition | Mechanism for minimising winter pressures |
| --- | --- | --- |
| CCP – Hospital At Home | Provides intensive, hospital-level care for acute conditions that normally require an acute hospital bed, in patient's home for short episode through multidisciplinary healthcare teams. | Offers alternative location for hospital-level care by managing those who would otherwise present to hospital safely in the community |
| Example interventions:<br>**Hospital At Home** | Patients with chronic disease sent home to recover as part of a hospital at home programme. Signs of deteriorating health picked up after patient discharge from hospital. Connected real-time monitoring means less time spent on in-person visits and can help to improve clinical documentation. | Offers alternative to hospital admission, by managing those who would otherwise remain in hospital safely in the community |

**Table 60.**
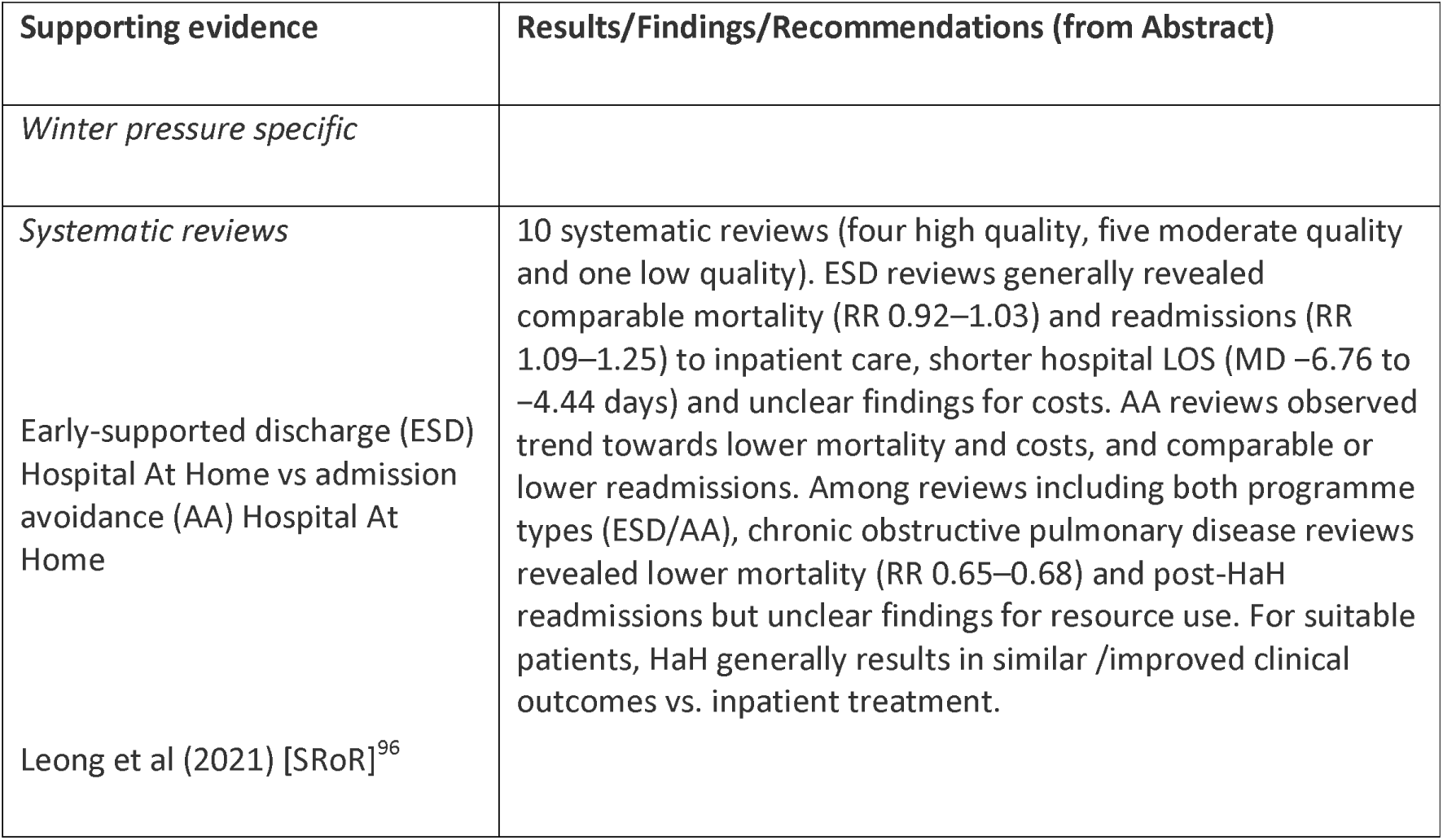
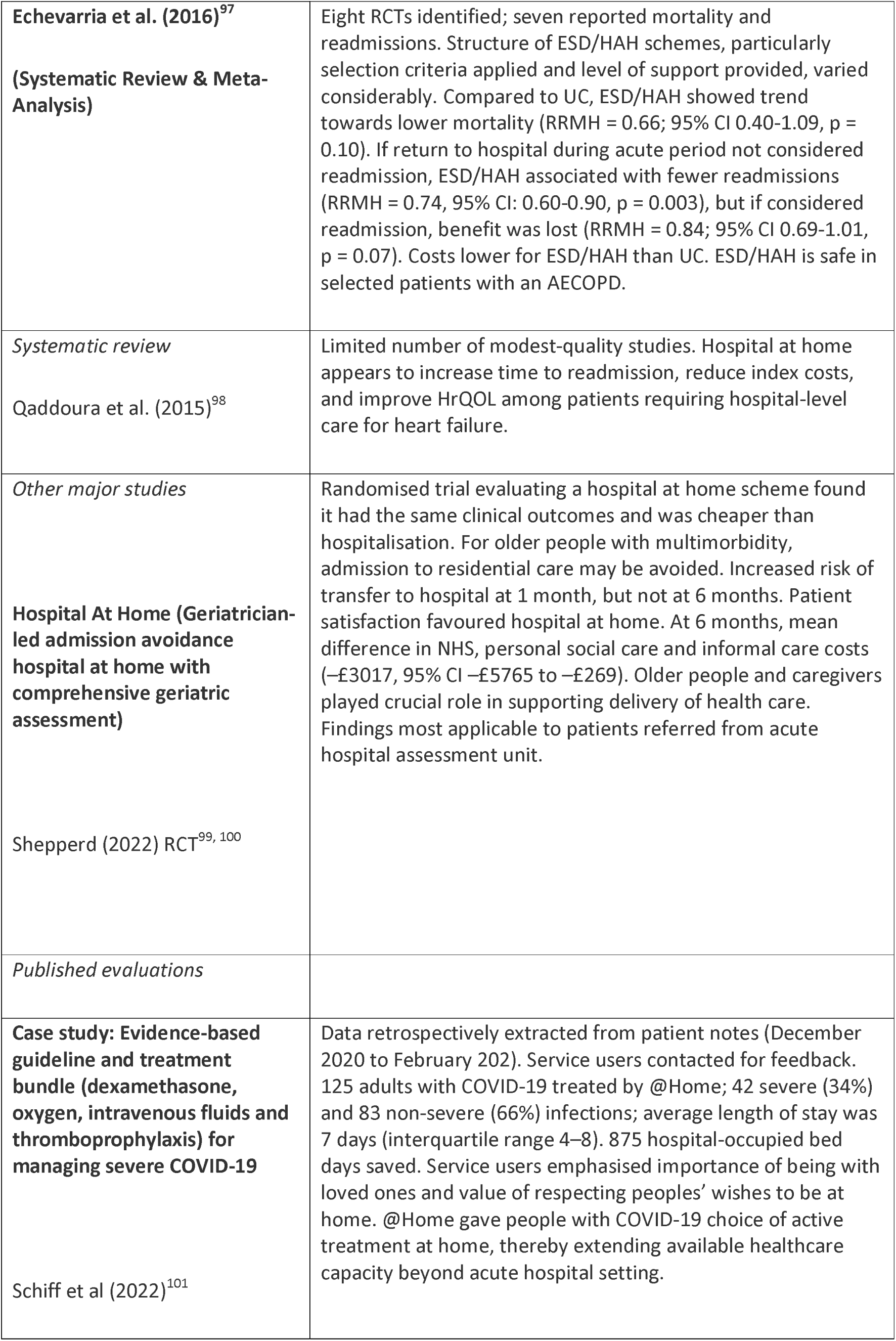
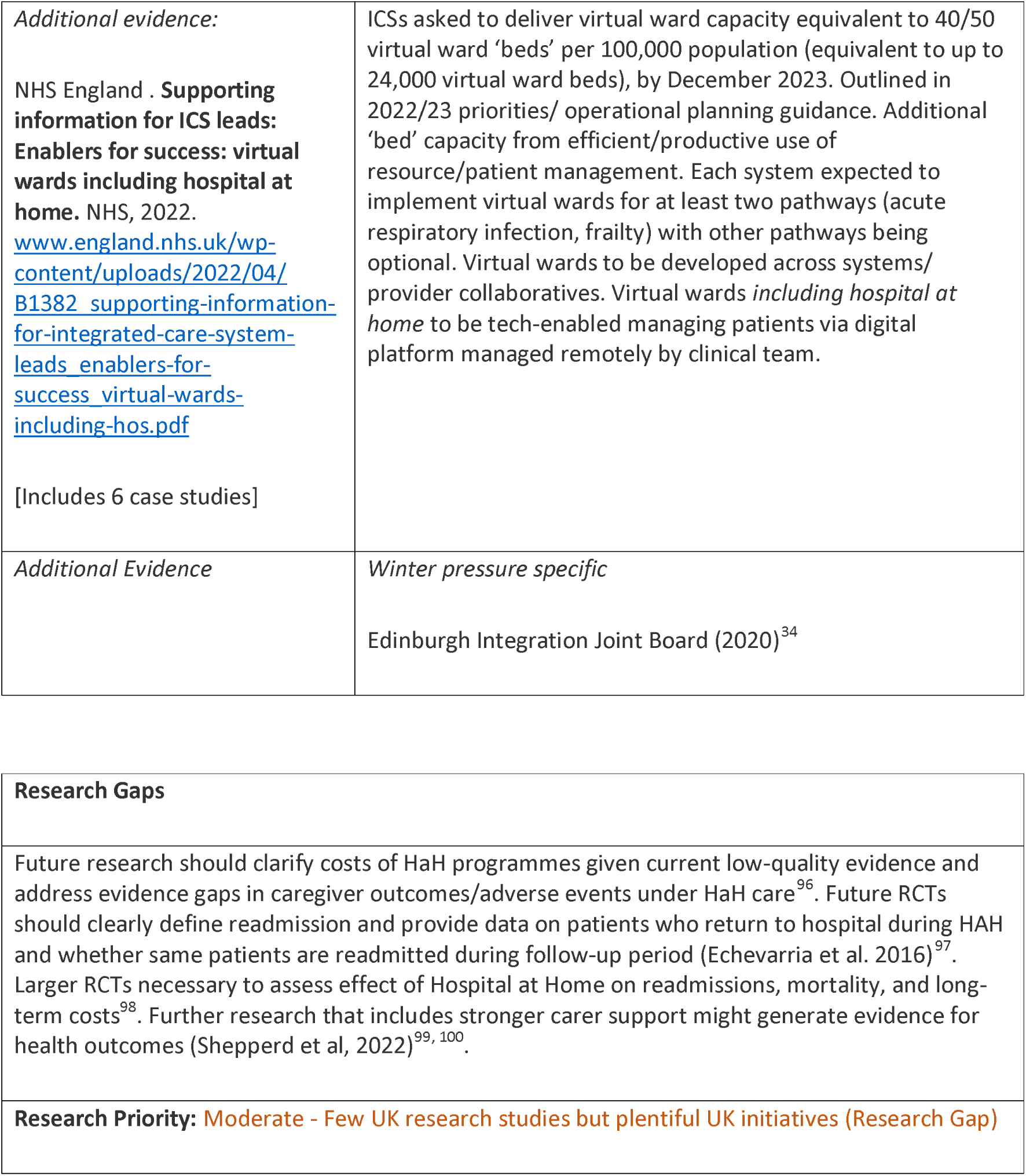
Hospital At Home: Interventions and Supporting Evidence.

##### CCP – Telecare

**Table 61.** Telecare: Definitions and Rationales.

| Taxonomy heading | Brief description/definition | Mechanism for minimising winter pressures |
| --- | --- | --- |
| CCP – Telecare | Telecare services use personal alarm and other monitored devices such as fall detectors or home sensors to call the monitoring centre in an emergency. Telecare offer alternative to moving into a care home or having a live-in carer. | Transfer demands on physical premises and resources to remote delivery. Also may substitute for home visits. |
| Example interventions:<br>**Remote consultations** | Appointment takes place between a patient and a clinician over the telephone or using video, as opposed to face-to-face. | Aims to reduce pressure on secondary care staff – allows for distributed service across multiple sites. |
| Example interventions:<br>**Paediatric symptom app**<br>Care Quality Commission. (2021). Collaboration in urgent and emergency care.<sup>7</sup><br>CCP – Telecare | In Cornwall and Isles of Scilly, parents encouraged to download a paediatric app – parents and carers could input symptoms and be advised whether to provide care at home or seek medical attention. Newsletters developed to inform families. | Reduces numbers of patients presenting to emergency department by targeting and prioritising symptoms |
| Example interventions:<br>**Remote consultations**<br>(Herefordshire and Worcestershire)<br>Care Quality Commission. (2021). Collaboration in urgent | Care homes provided with computer tablets to enable remote consultations with frail/vulnerable residents - allowed virtual ward rounds from primary care and timely access to clinical advice. Care home staff supported with training. 30 care homes included in social media pilot to enable greater connectivity with family and friends. | Allows more timely access to clinical advice and allows ongoing monitoring of frail and vulnerable patients. |
| and emergency care.<sup>7</sup> |  |  |
| Example interventions:<br>**Remote monitoring** | Allows clinical teams to keep track of patients with chronic conditions safely and in the comfort of their own home. Part of wider plan to improve digital health services for people with long term conditions. | Aims to reduce pressure on hospital services and improve outcomes by detecting and addressing signs of deteriorating health earlier among recently discharged and chronically ill patients. |

**Table 62.**
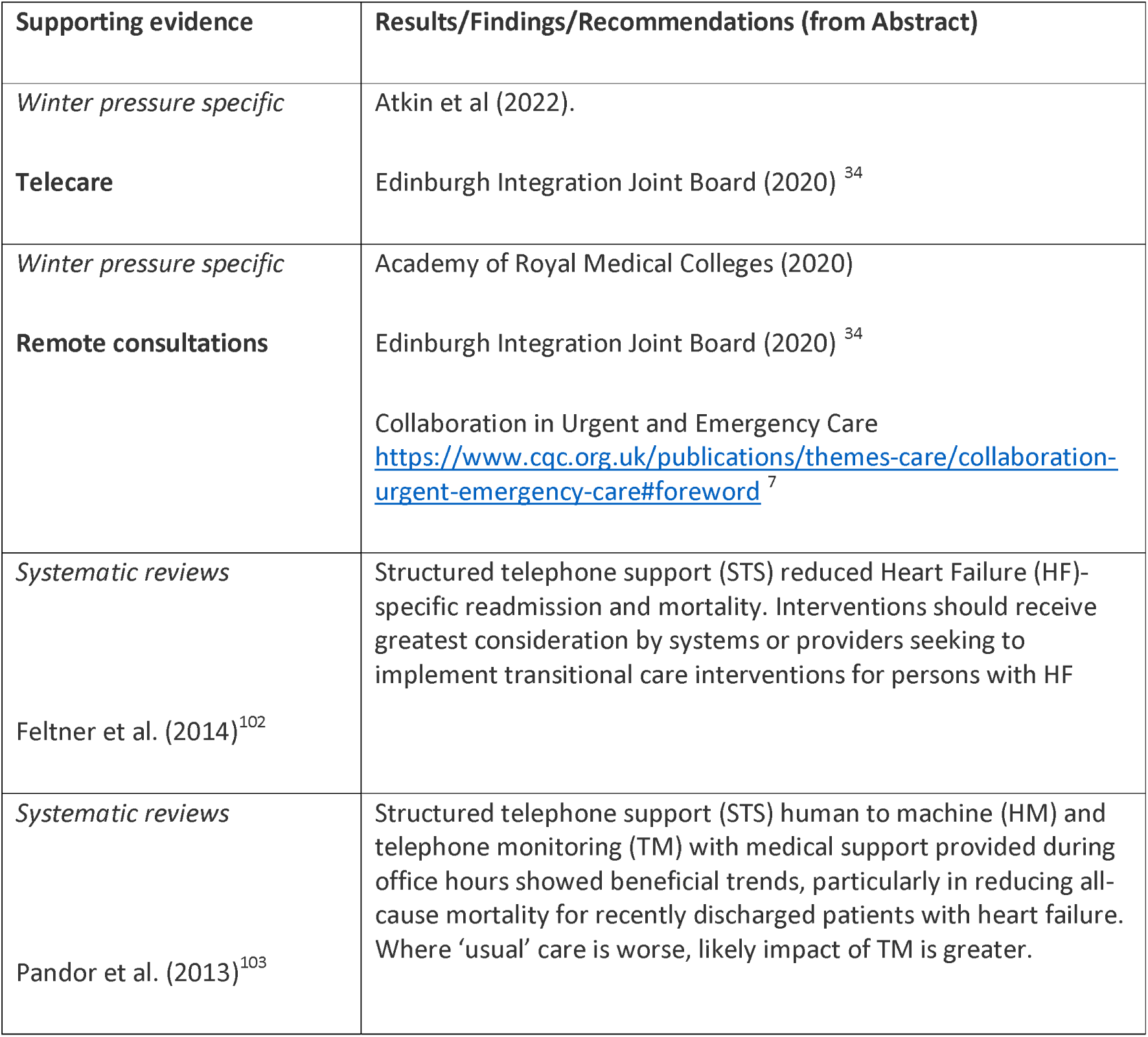
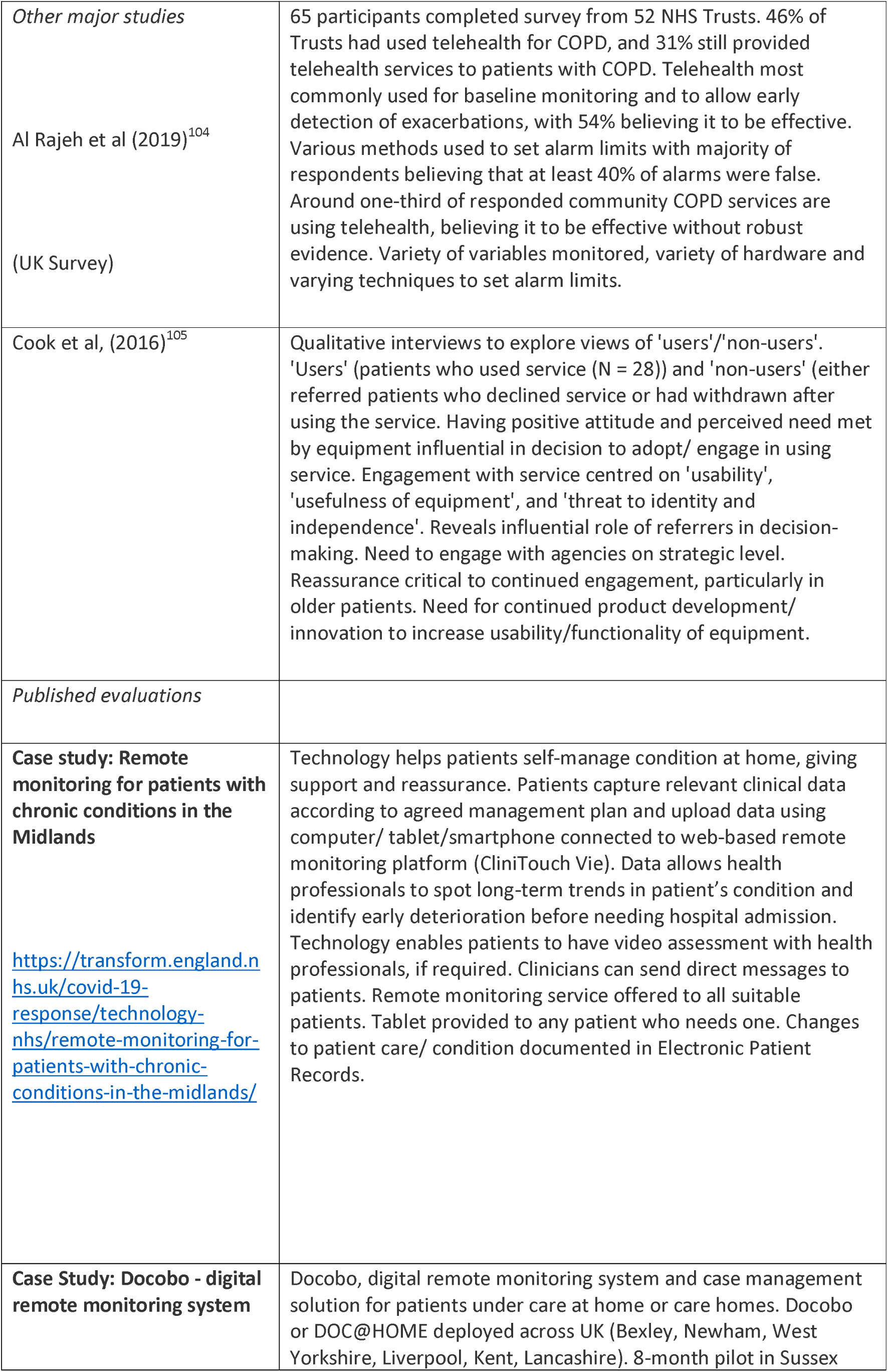
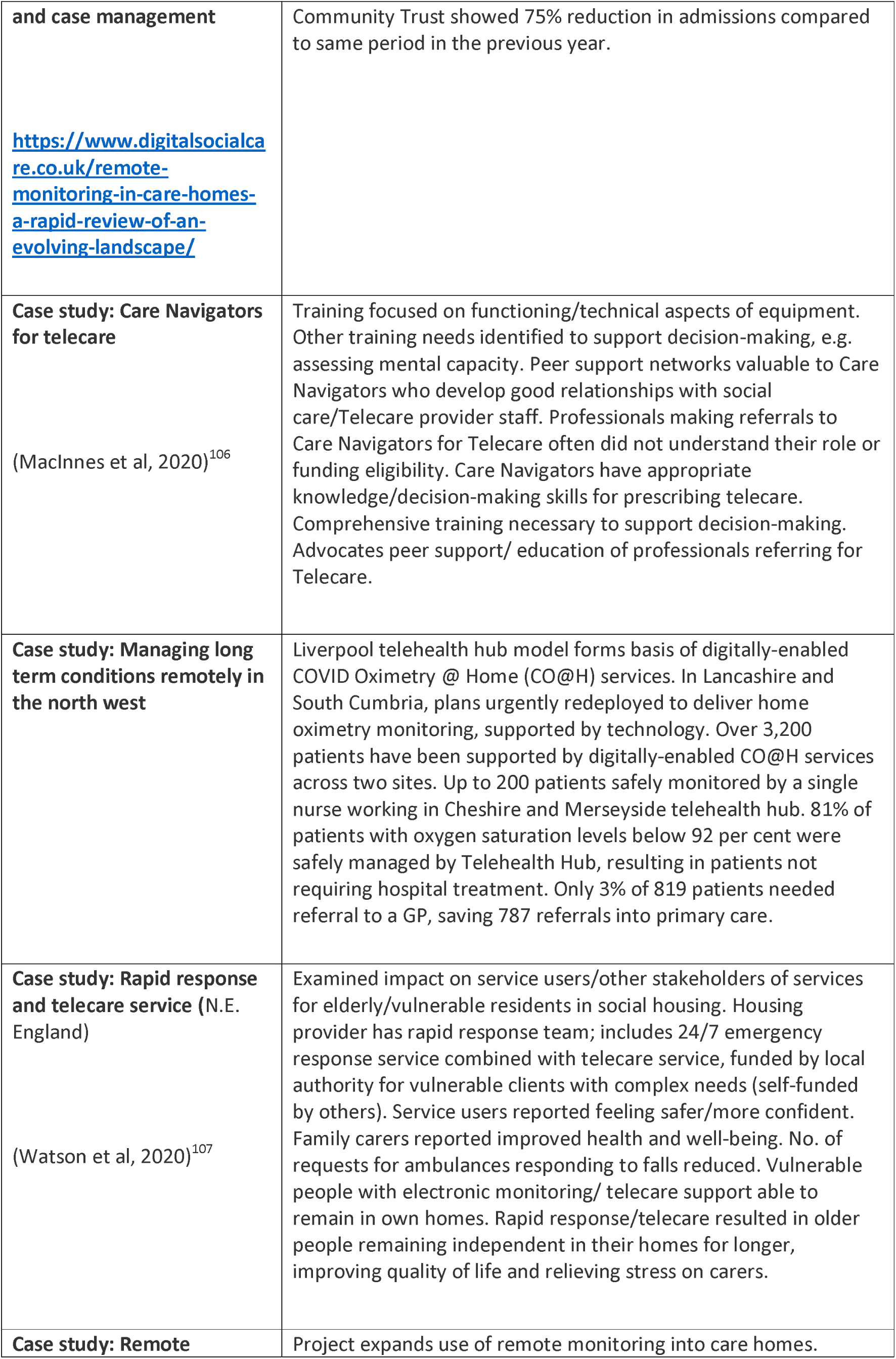
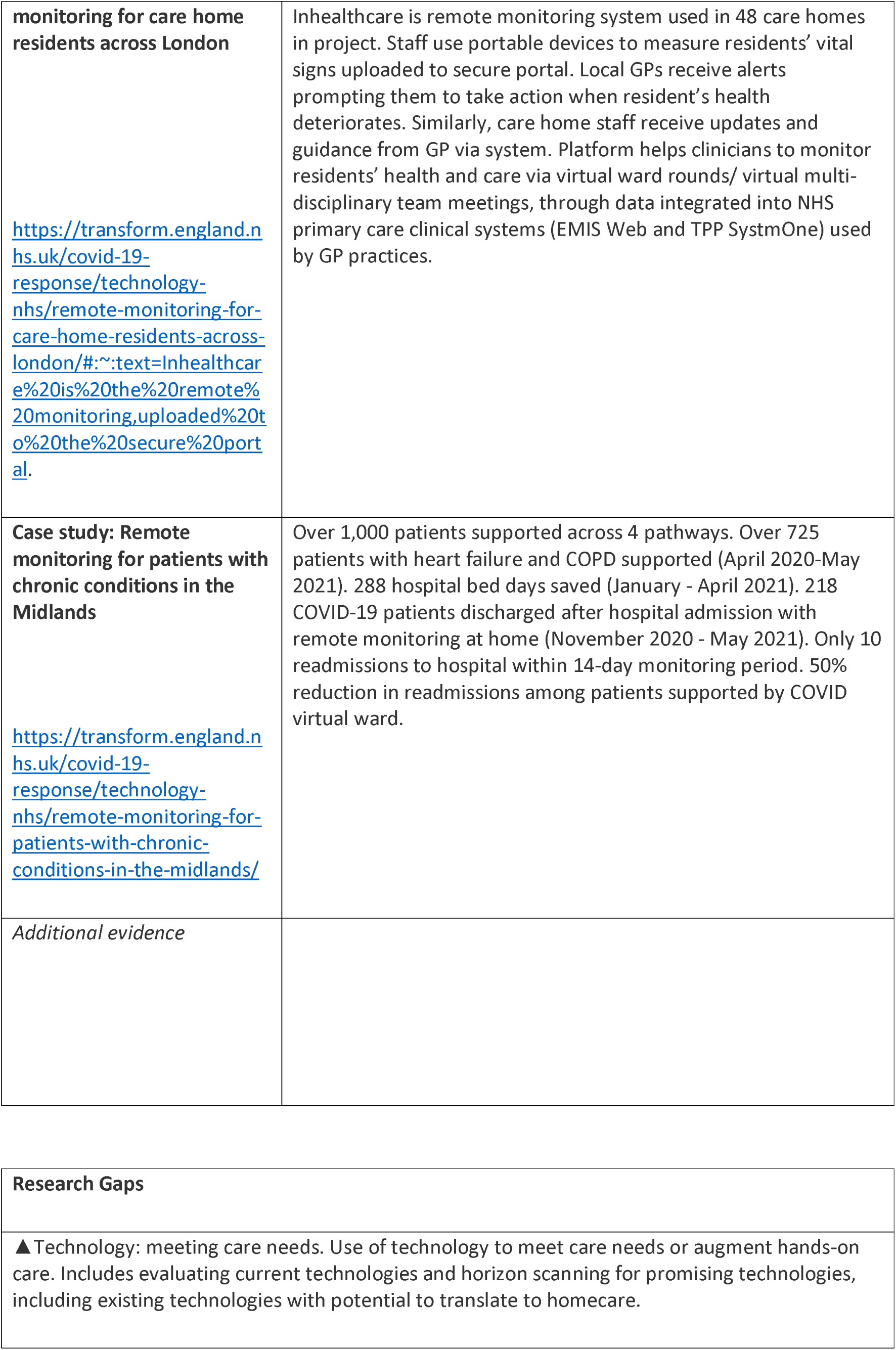
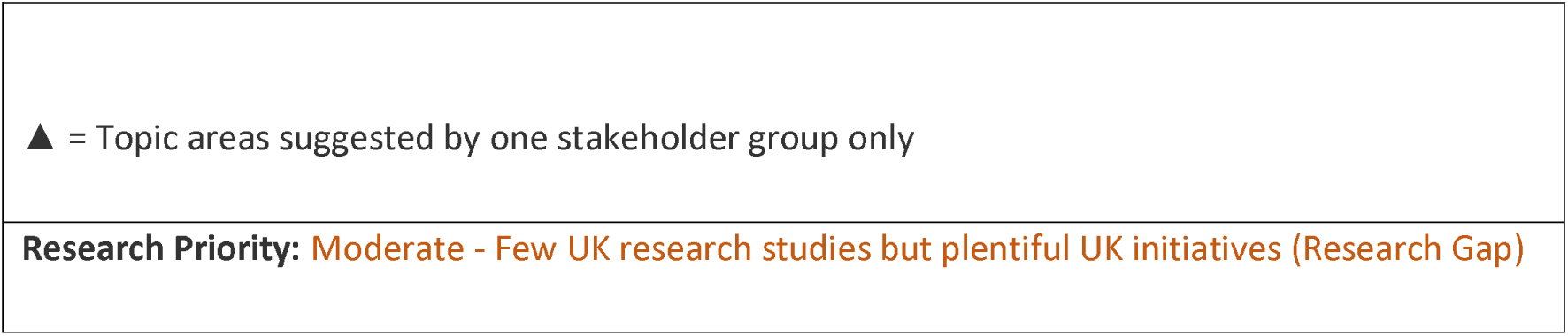
Telecare: Interventions and Supporting Evidence.

#### Changing Community Provision – Facilitated Discharge

Findings are presented in detail in Tables 63 to 81 below and associated summary paragraphs.

##### CCP - Rehabilitation, Recovery and Reablement

**Table 63.** Rehabilitation, Recovery and Reablement: Definitions and Rationales.

| **Taxonomy heading** | **Brief description/definition** | **Mechanism for minimising winter pressures** |
| --- | --- | --- |
| CCP - Rehabilitation, Recovery and Reablement | Usually provide time limited therapy for patients who require on-going physiotherapy or occupational therapy to regain maximum independence. Users of rehabilitation schemes have often sustained a fracture and may have undergone surgery. Rehabilitation schemes delivered by multi-disciplinary team, often led by physiotherapists/occupational therapists. 'Doing with' model vs. home care which is a 'doing for' model. Reablement services help people to retain or regain their skills and confidence so they can learn to manage again after a period of illness. Service usually provided in person's own home by team of social care professionals. | Minimises unnecessary delays in hospital discharge. Not specific to winter but frequently mentioned in winter planning documents. Reablement generally designed to help people learn or relearn skills necessary for daily living which may have been lost through deterioration in health and/or increased support needs. Central focus on regaining physical ability and active reassessment. |
| Example interventions: **Front line Occupational therapists**<br>https://www.scie.org.uk/care-providers/coronavirus-covid-19/commissioning/hospital-discharge-admissions | Occupational therapists (OTs) moved to frontline calls to provide/advise on reablement, home equipment and adaptations so people did not lose skills and independence. |  |
| Example interventions: **Advanced Clinical Practitioner Roles** (Hove, 2022)<sup>108</sup> | Registered practitioner with expert knowledge base, complex decision-making skills and clinical competencies for expanded autonomous scope of practice. Characteristics shaped by |  |
|  | context in which individual practices. |  |
| Example interventions:<br>**Reablement Assistants** | Reablement Assistants work within the community, usually in peoples' homes, to promote independence and choice for people by helping them to work towards their reablement goals as set out in their support plan. Role involves working closely with carers and others (like a Team leader and OT's) and meeting person's needs. | Reablement supports people to do things for themselves to try to prevent unnecessary admission to hospital or residential care. |
| Example interventions:<br>**Reablement Coordinators** | Delivers outward facing service to coordinate services for people accessing Community Reablement. Works within support team, receiving referrals into the service via electronic systems, planning and coordinating resources to facilitate delivery of services. | Reablement supports people to do things for themselves. |
| Example interventions:<br>**Intermediate care (IC) beds - rehabilitation and reablement** | Involves temporary stay in a care home, community hospital, or standalone IC facility. Patient receives support similar to home-based IC to help reach their goals. The sooner bedbased IC starts, ideally within two days of referral, the better the chance of success. | IC that aims to avoid unnecessary hospital admission involves rehabilitation to help patients continue to live at home when facing increasing difficulty with daily life due to illness or disability, or prevent a premature, permanent move into residential care. |

**Table 64.**
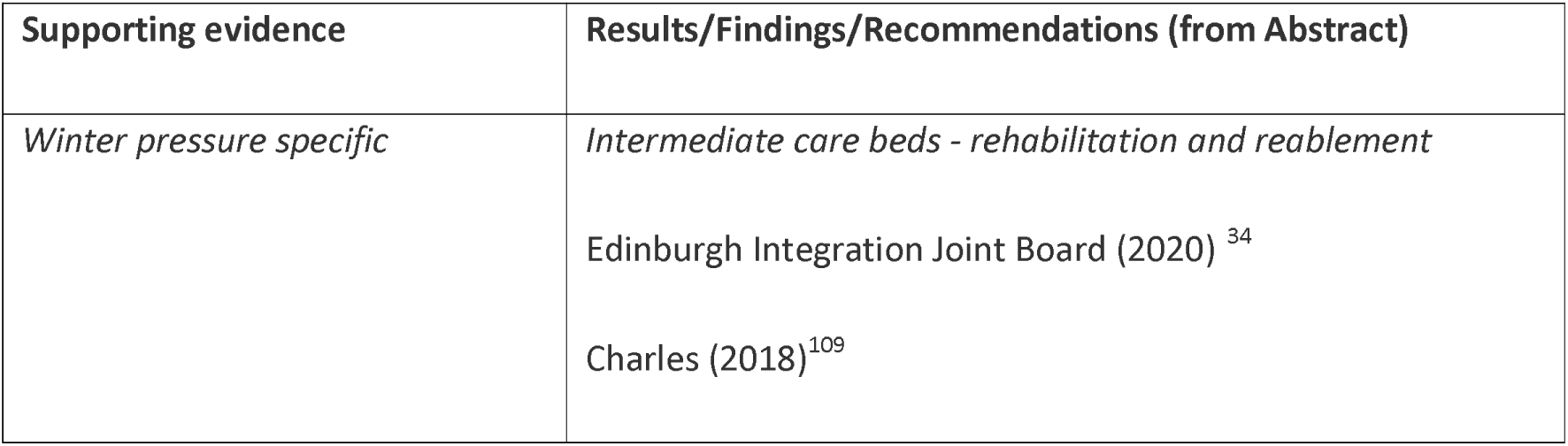
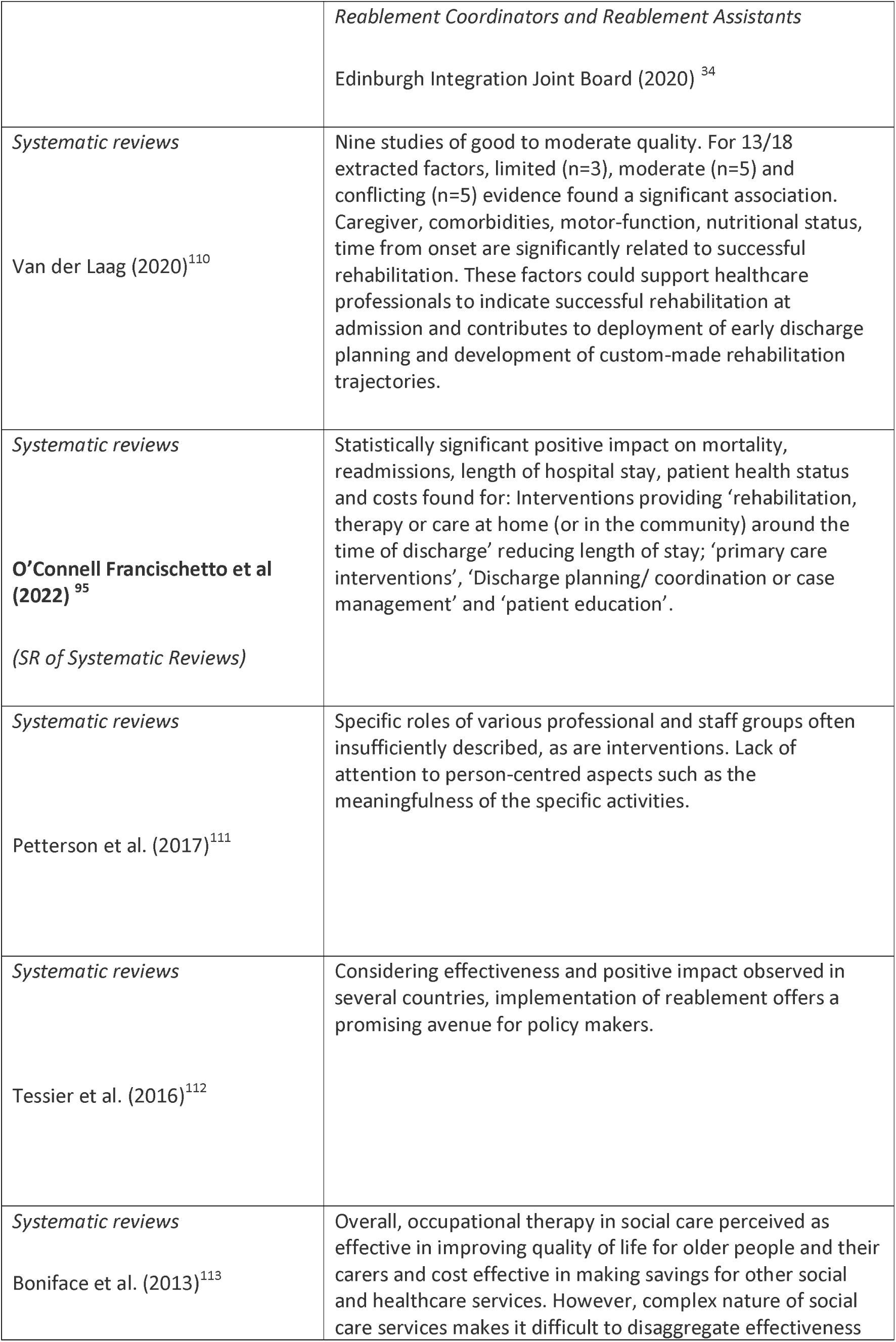
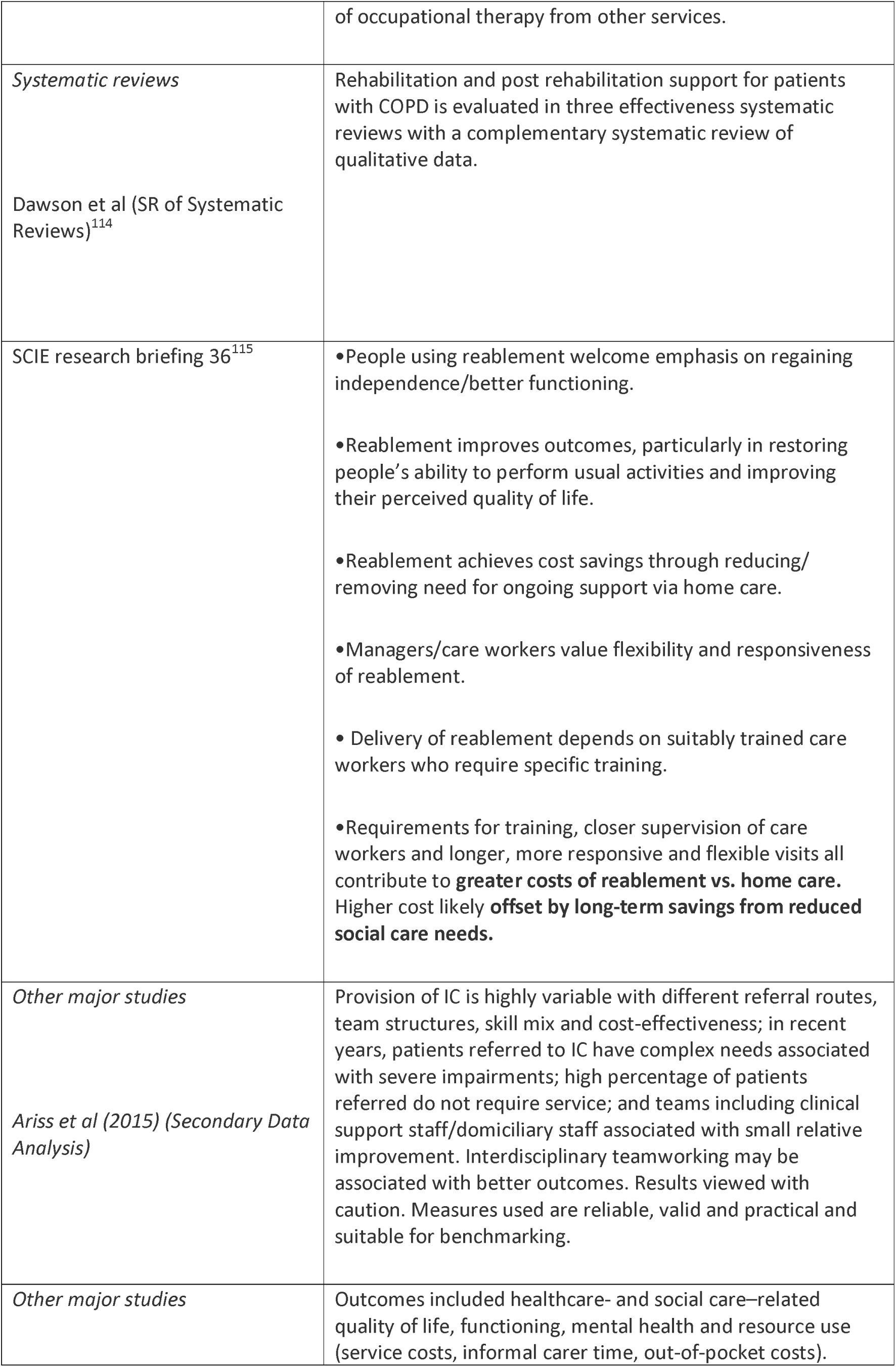
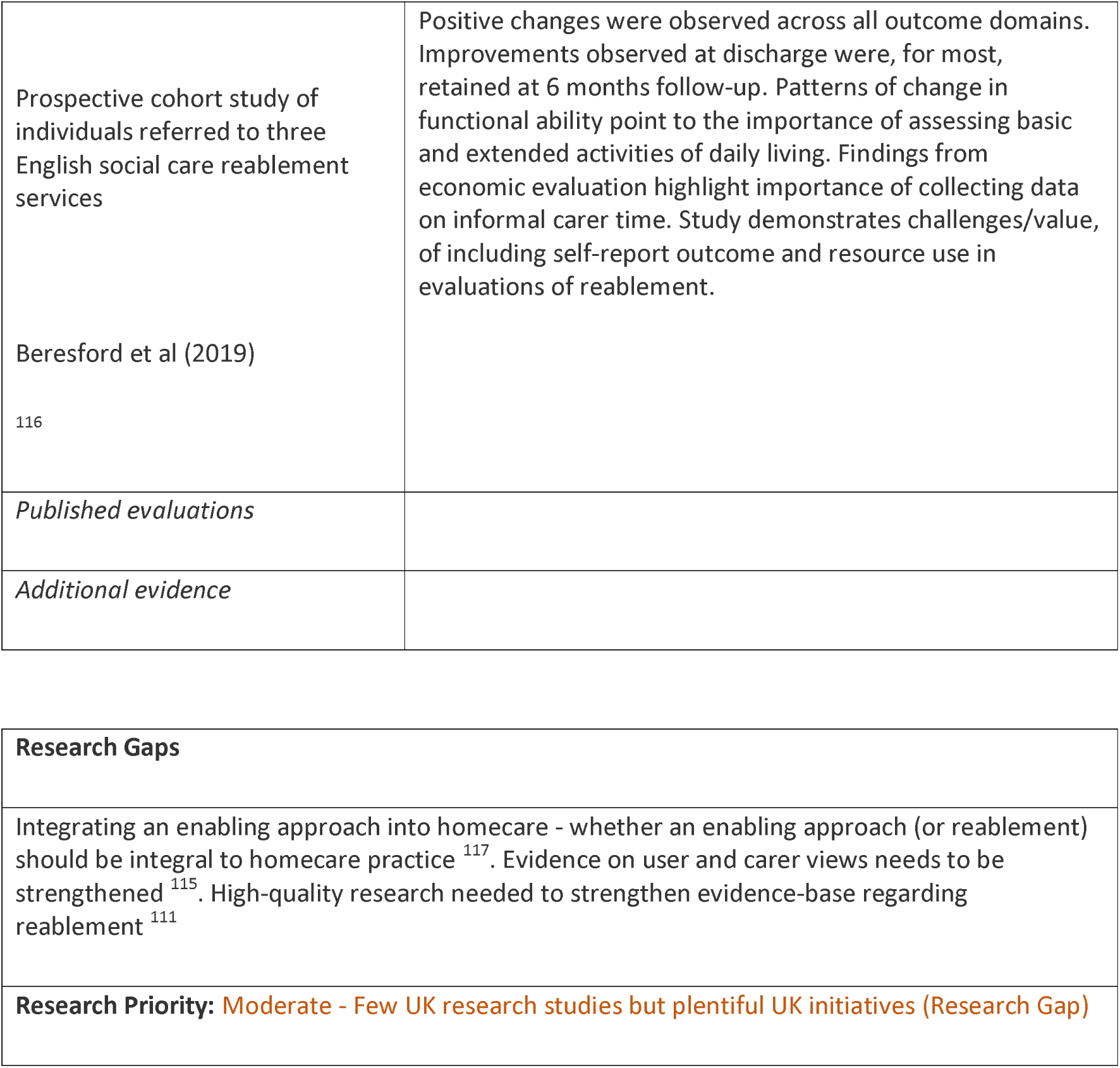
Rehabilitation, Recovery and Reablement: Interventions and Supporting Evidence.

| Supporting evidence | Results/Findings/Recommendations (from Abstract) |
| --- | --- |
| Winter pressure specific | *Intermediate care beds - rehabilitation and reablement*<br>Edinburgh Integration Joint Board (2020) <sup>34</sup><br>Charles (2018)<sup>109</sup> |
|  | *Reablement Coordinators and Reablement Assistants*<br>Edinburgh Integration Joint Board (2020) <sup>34</sup> |
| *Systematic reviews*<br>Van der Laag (2020)<sup>110</sup> | Nine studies of good to moderate quality. For 13/18 extracted factors, limited (n=3), moderate (n=5) and conflicting (n=5) evidence found a significant association. Caregiver, comorbidities, motor-function, nutritional status, time from onset are significantly related to successful rehabilitation. These factors could support healthcare professionals to indicate successful rehabilitation at admission and contributes to deployment of early discharge planning and development of custom-made rehabilitation trajectories. |
| *Systematic reviews*<br>**O'Connell Francischetto et al (2022)** <sup>95</sup><br>*(SR of Systematic Reviews)* | Statistically significant positive impact on mortality, readmissions, length of hospital stay, patient health status and costs found for: Interventions providing 'rehabilitation, therapy or care at home (or in the community) around the time of discharge' reducing length of stay; 'primary care interventions', 'Discharge planning/ coordination or case management' and 'patient education'. |
| *Systematic reviews*<br>Petterson et al. (2017)<sup>111</sup> | Specific roles of various professional and staff groups often insufficiently described, as are interventions. Lack of attention to person-centred aspects such as the meaningfulness of the specific activities. |
| *Systematic reviews*<br>Tessier et al. (2016)<sup>112</sup> | Considering effectiveness and positive impact observed in several countries, implementation of reablement offers a promising avenue for policy makers. |
| *Systematic reviews*<br>Boniface et al. (2013)<sup>113</sup> | Overall, occupational therapy in social care perceived as effective in improving quality of life for older people and their carers and cost effective in making savings for other social and healthcare services. However, complex nature of social care services makes it difficult to disaggregate effectiveness |
|  | of occupational therapy from other services. |
| *Systematic reviews*<br>Dawson et al (SR of Systematic Reviews)<sup>114</sup> | Rehabilitation and post rehabilitation support for patients with COPD is evaluated in three effectiveness systematic reviews with a complementary systematic review of qualitative data. |
| SCIE research briefing 36<sup>115</sup> | •People using reablement welcome emphasis on regaining independence/better functioning.<br>•Reablement improves outcomes, particularly in restoring people's ability to perform usual activities and improving their perceived quality of life.<br>•Reablement achieves cost savings through reducing/removing need for ongoing support via home care.<br>•Managers/care workers value flexibility and responsiveness of reablement.<br>• Delivery of reablement depends on suitably trained care workers who require specific training.<br>•Requirements for training, closer supervision of care workers and longer, more responsive and flexible visits all contribute to **greater costs of reablement vs. home care**. Higher cost likely **offset by long-term savings from reduced social care needs**. |
| *Other major studies*<br>Ariss et al (2015) (Secondary Data Analysis) | Provision of IC is highly variable with different referral routes, team structures, skill mix and cost-effectiveness; in recent years, patients referred to IC have complex needs associated with severe impairments; high percentage of patients referred do not require service; and teams including clinical support staff/domiciliary staff associated with small relative improvement. Interdisciplinary teamworking may be associated with better outcomes. Results viewed with caution. Measures used are reliable, valid and practical and suitable for benchmarking. |
| *Other major studies* | Outcomes included healthcare- and social care–related quality of life, functioning, mental health and resource use (service costs, informal carer time, out-of-pocket costs). |
| Prospective cohort study of individuals referred to three English social care reablement services<br>Beresford et al (2019)<br>116 | Positive changes were observed across all outcome domains. Improvements observed at discharge were, for most, retained at 6 months follow-up. Patterns of change in functional ability point to the importance of assessing basic and extended activities of daily living. Findings from economic evaluation highlight importance of collecting data on informal carer time. Study demonstrates challenges/value, of including self-report outcome and resource use in evaluations of reablement. |
| *Published evaluations* |  |
| *Additional evidence* |  |

|  |
| --- |
| **Research Gaps** |
| Integrating an enabling approach into homecare - whether an enabling approach (or reablement) should be integral to homecare practice <sup>117</sup>. Evidence on user and carer views needs to be strengthened <sup>115</sup>. High-quality research needed to strengthen evidence-base regarding reablement <sup>111</sup> |
| **Research Priority:** Moderate - Few UK research studies but plentiful UK initiatives (Research Gap) |

##### CCP - Step Down Beds

**Table 65.** Step Down Beds: Definitions and Rationales.

| Taxonomy heading | Brief description/definition | Mechanism for minimising winter pressures |
| --- | --- | --- |
| CCP - Step Down Beds | Aim to provide community bed for patients following acute inpatient episode, not yet ready to go home, needing period of recuperation/ recovery. Includes people requiring residential long-term placement who need further recovery period before being able to engage in active rehabilitation; they may need additional rehabilitation therapy before being able to return home: or may be yet to complete Continuing Healthcare or Social Care Assessments for their long-term care needs prior to discharge home or long-term placement. | Helps to relieve pressure on NHS beds and to flex capacity at times of especially high demand. Acute trusts need to plan effectively for sufficient acute beds as well as access to community beds to improve patient flow. Minimises unnecessary delays in hospital discharge. Not specific to winter but frequently mentioned in winter planning documents |
| Example interventions: **Step-down beds for discharge from stroke unit** | Provide high level of support for patients who have undergone a cerebrovascular accident (CVA) and to provide high level rehabilitation, to assist in gaining increased independence. Stroke schemes delivered by multi-disciplinary team (medical, nursing, allied health and social work, with assistance of generic rehabilitation assistants (covering physiotherapy and occupational therapy)). Schemes often longer-term than other schemes | Usually provide care in alternative setting to acute hospital. |
| **Step Down Facilities (Care and Reablement Centre)**<br>https://www.scie.org.uk/care-providers/<br>coronavirus-covid-19/commissioning/hospital-discharge- | Buckinghamshire Council made up to 240 beds available as care and reablement centre within just three weeks, staffed by care workers and a team of volunteers. | Supports people leaving hospital but unable to return home, as well as those who live at home who may need temporary short-term support. |
| admissions |  |  |
| Example interventions:<br>**‘Discharge to assess’ facility (See main section)**<br>https://www.scie.org.uk/care-providers/coronavirus-covid-19/commissioning/hospital-discharge-admissions | Reading Borough Council paid for use of hotel rooms at a local Holiday Inn. Others stayed in the hotel to protect shielding relatives, or self-isolated with COVID-19 symptoms. | Operated ‘discharge to assess’ facility, allowing time to arrange care packages and equipment for people due to go home. |
| **‘Discharge to assess’ facility (See main section)**<br>https://www.scie.org.uk/care-providers/coronavirus-covid-19/commissioning/hospital-discharge-admissions<br>and<br>https://www.theguardian.com/society/2020/may/03/care-room-with-a-view-uk-hotels-offer-respite-to-non-covid-patients | Hammersmith & Fulham made similar arrangements to Reading Borough Council (above) with Novotel but stood down when capacity not needed. Astute commissioning enabled good ongoing relationships with business whilst avoiding expensive contractual commitments. | See above |

**Table 66.**
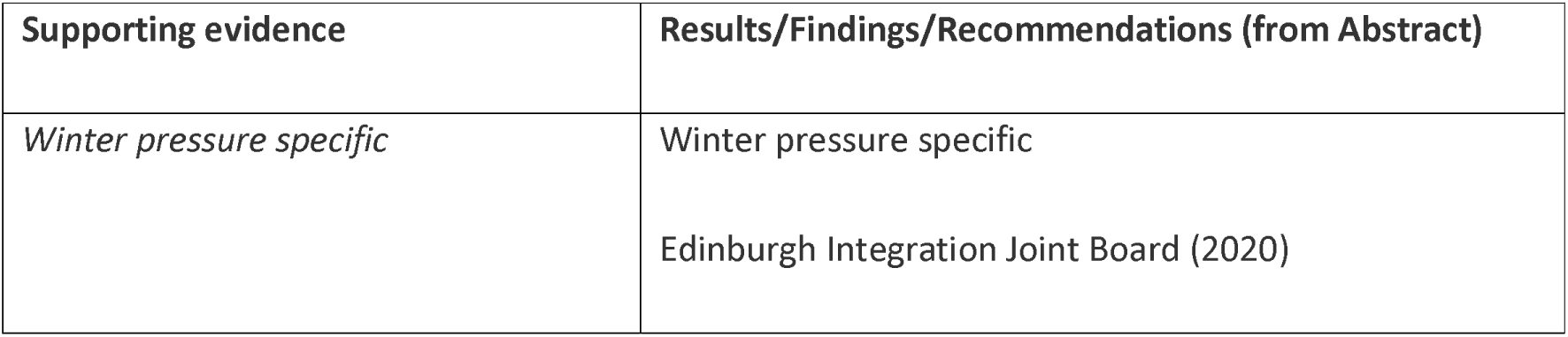
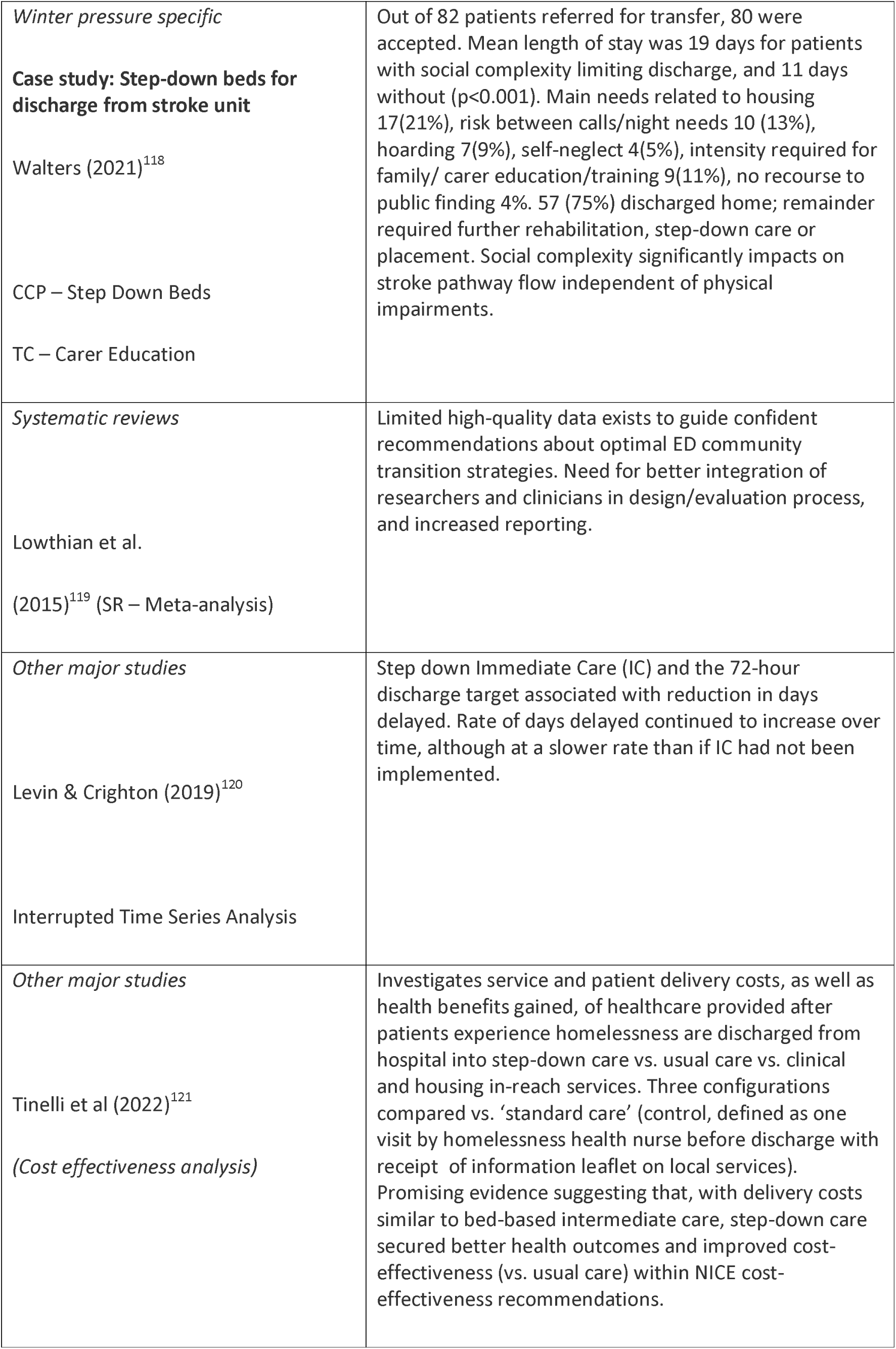
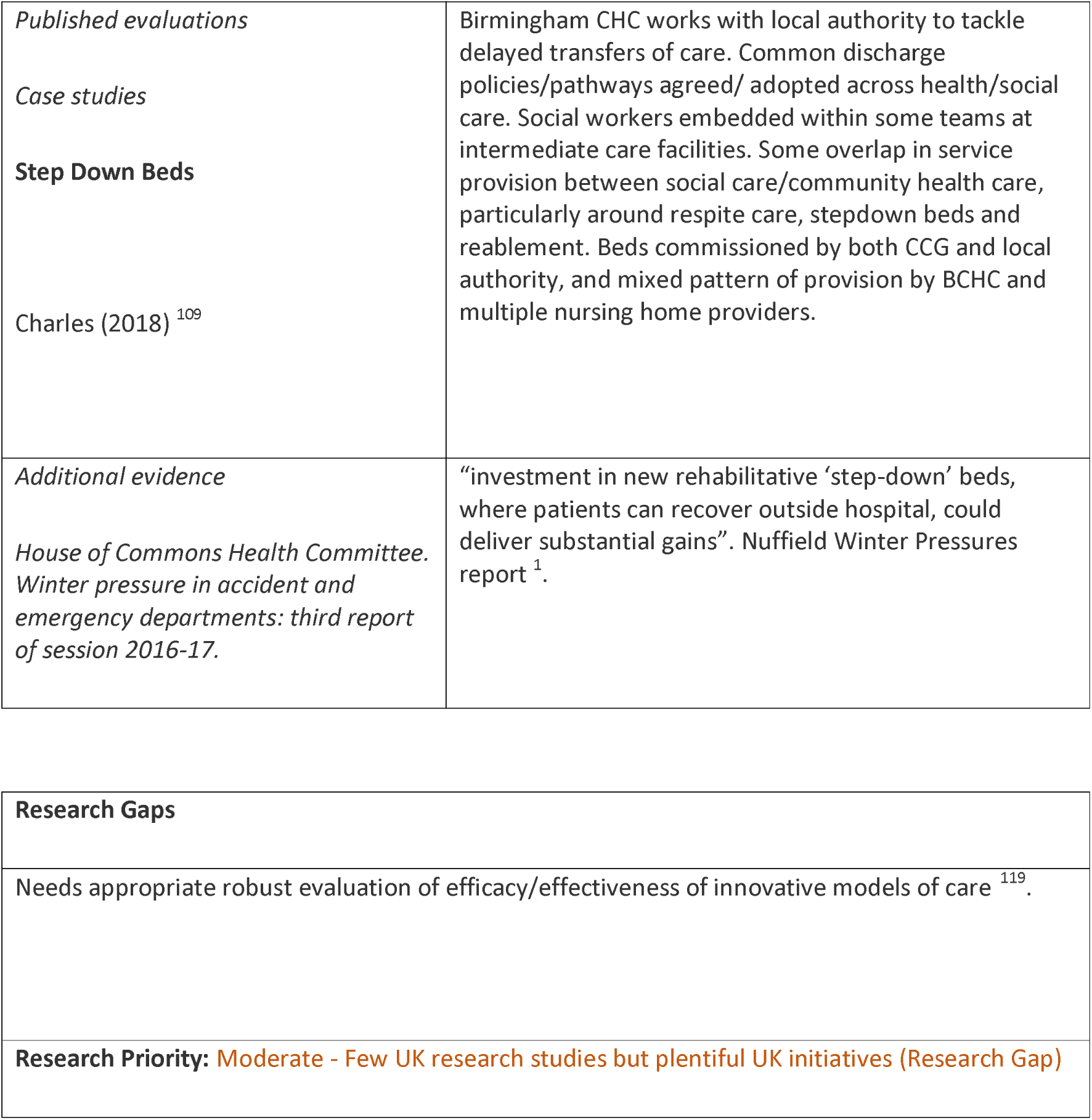
Step Down Beds: Interventions and Supporting Evidence.

##### CCP – Virtual Hospitals/Virtual Wards

**Table 67.**
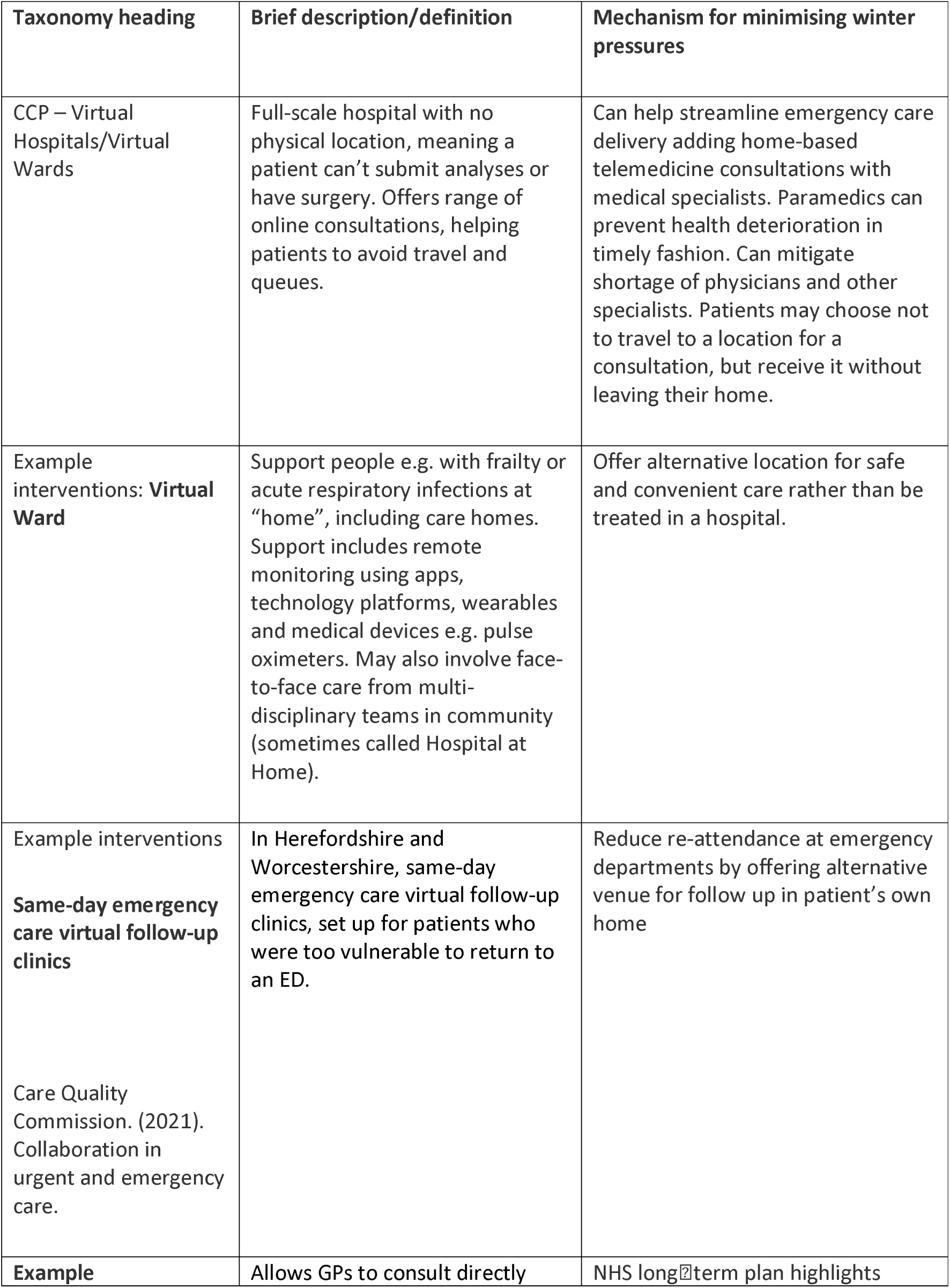
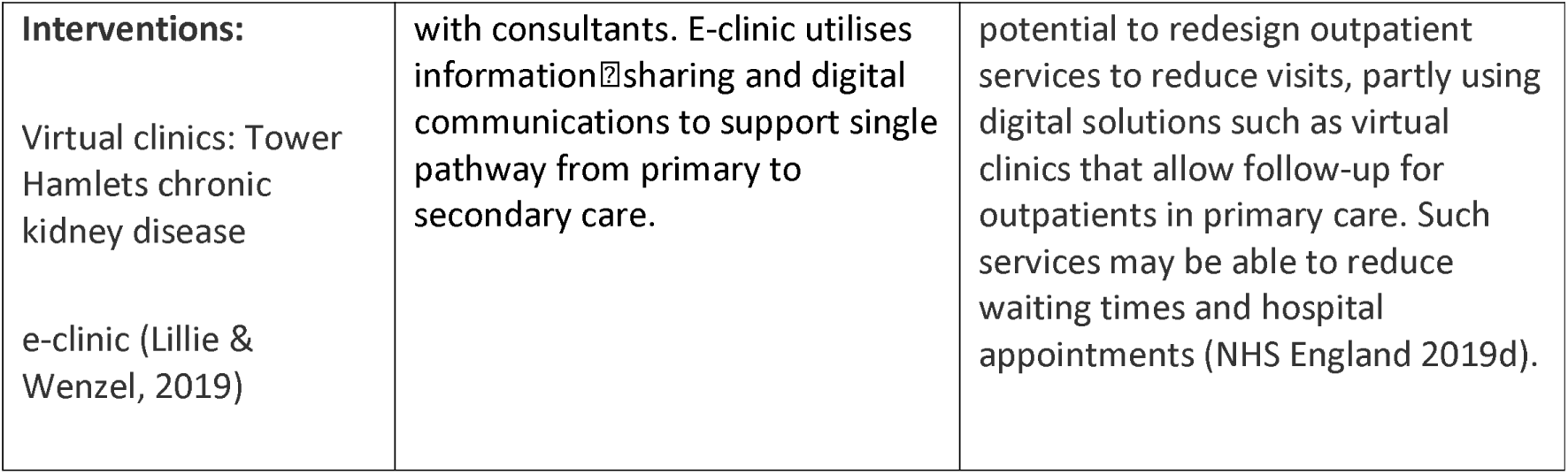
Virtual Hospitals/Virtual Wards: Definitions and Rationales.

| Taxonomy heading | Brief description/definition | Mechanism for minimising winter pressures |
| --- | --- | --- |
| CCP – Virtual Hospitals/Virtual Wards | Full-scale hospital with no physical location, meaning a patient can't submit analyses or have surgery. Offers range of online consultations, helping patients to avoid travel and queues. | Can help streamline emergency care delivery adding home-based telemedicine consultations with medical specialists. Paramedics can prevent health deterioration in timely fashion. Can mitigate shortage of physicians and other specialists. Patients may choose not to travel to a location for a consultation, but receive it without leaving their home. |
| Example interventions: **Virtual Ward** | Support people e.g. with frailty or acute respiratory infections at "home", including care homes. Support includes remote monitoring using apps, technology platforms, wearables and medical devices e.g. pulse oximeters. May also involve face-to-face care from multi-disciplinary teams in community (sometimes called Hospital at Home). | Offer alternative location for safe and convenient care rather than be treated in a hospital. |
| Example interventions<br>**Same-day emergency care virtual follow-up clinics**<br>Care Quality Commission. (2021). Collaboration in urgent and emergency care. | In Herefordshire and Worcestershire, same-day emergency care virtual follow-up clinics, set up for patients who were too vulnerable to return to an ED. | Reduce re-attendance at emergency departments by offering alternative venue for follow up in patient's own home |
| **Example** | Allows GPs to consult directly | NHS long-term plan highlights |
| **Interventions:**<br>Virtual clinics: Tower Hamlets chronic kidney disease<br>e-clinic (Lillie & Wenzel, 2019) | with consultants. E-clinic utilises information sharing and digital communications to support single pathway from primary to secondary care. | potential to redesign outpatient services to reduce visits, partly using digital solutions such as virtual clinics that allow follow-up for outpatients in primary care. Such services may be able to reduce waiting times and hospital appointments (NHS England 2019d). |

**Table 68.**
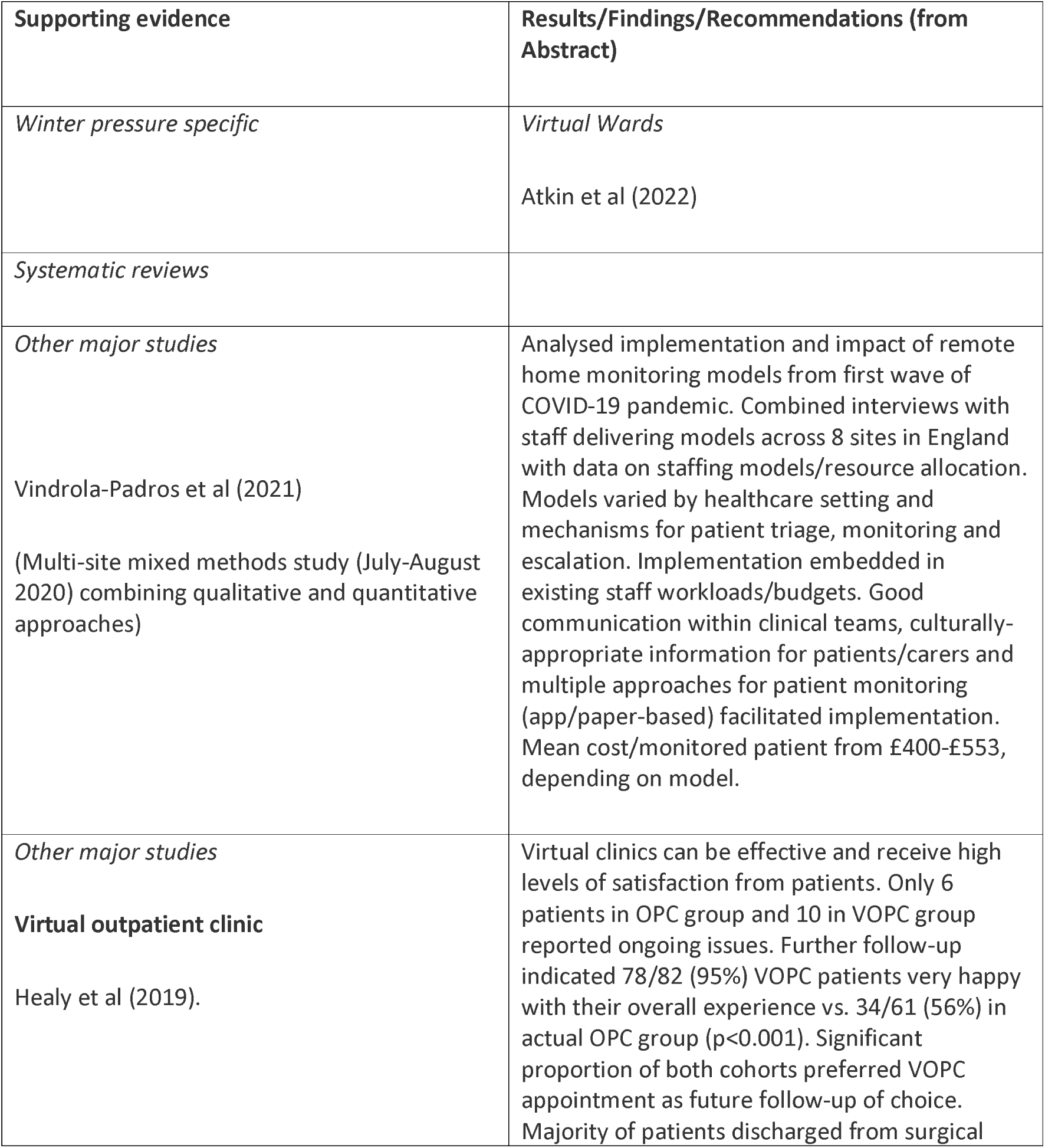
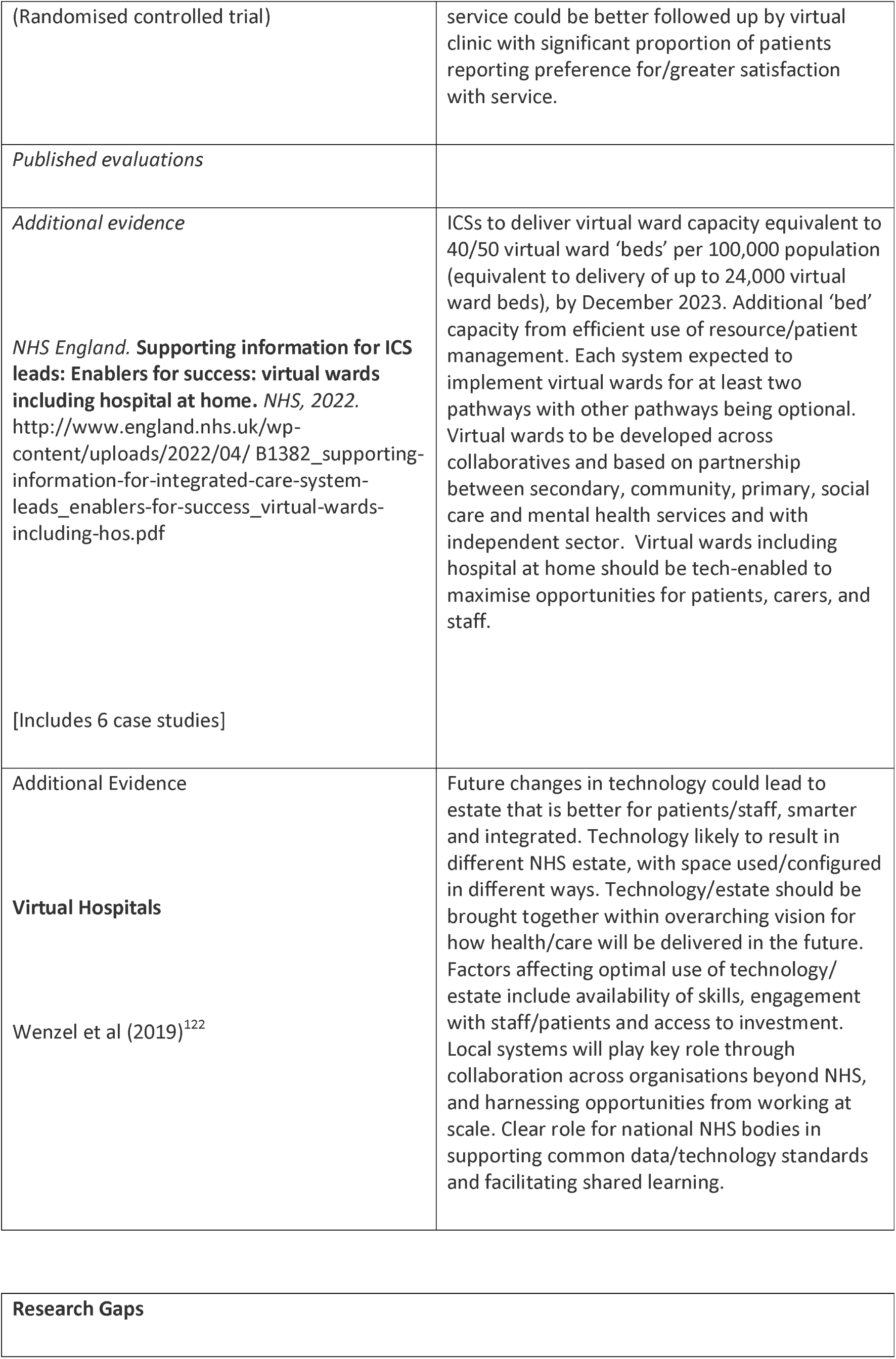
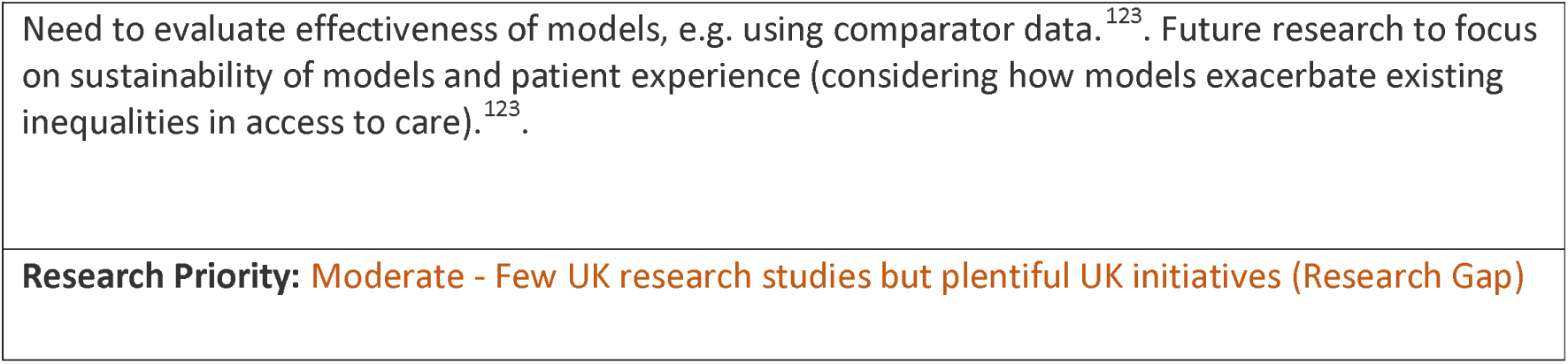
Virtual Hospitals/Virtual Wards: Interventions and Supporting Evidence.

##### CCP - Other Agencies

**Table 69.**
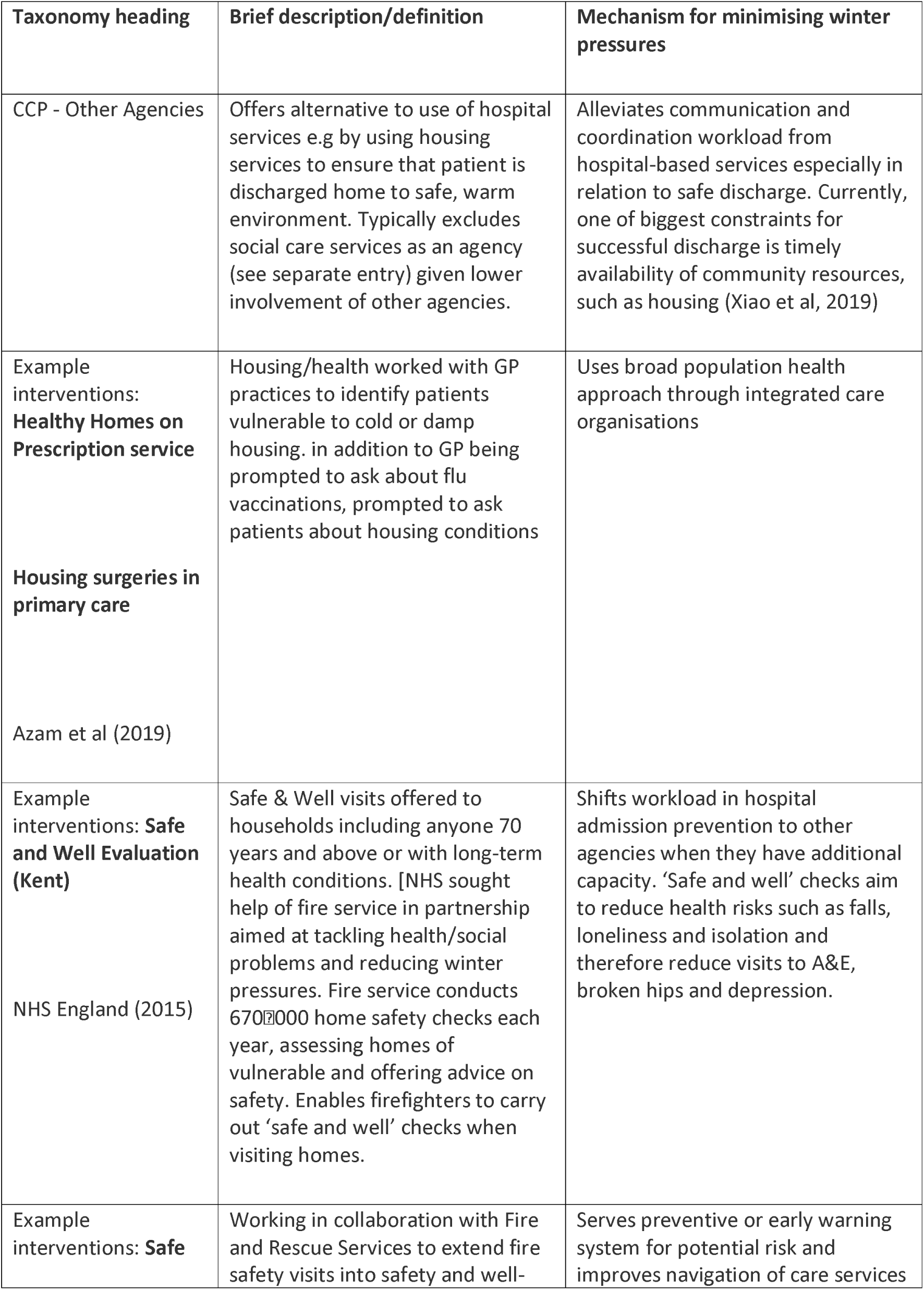
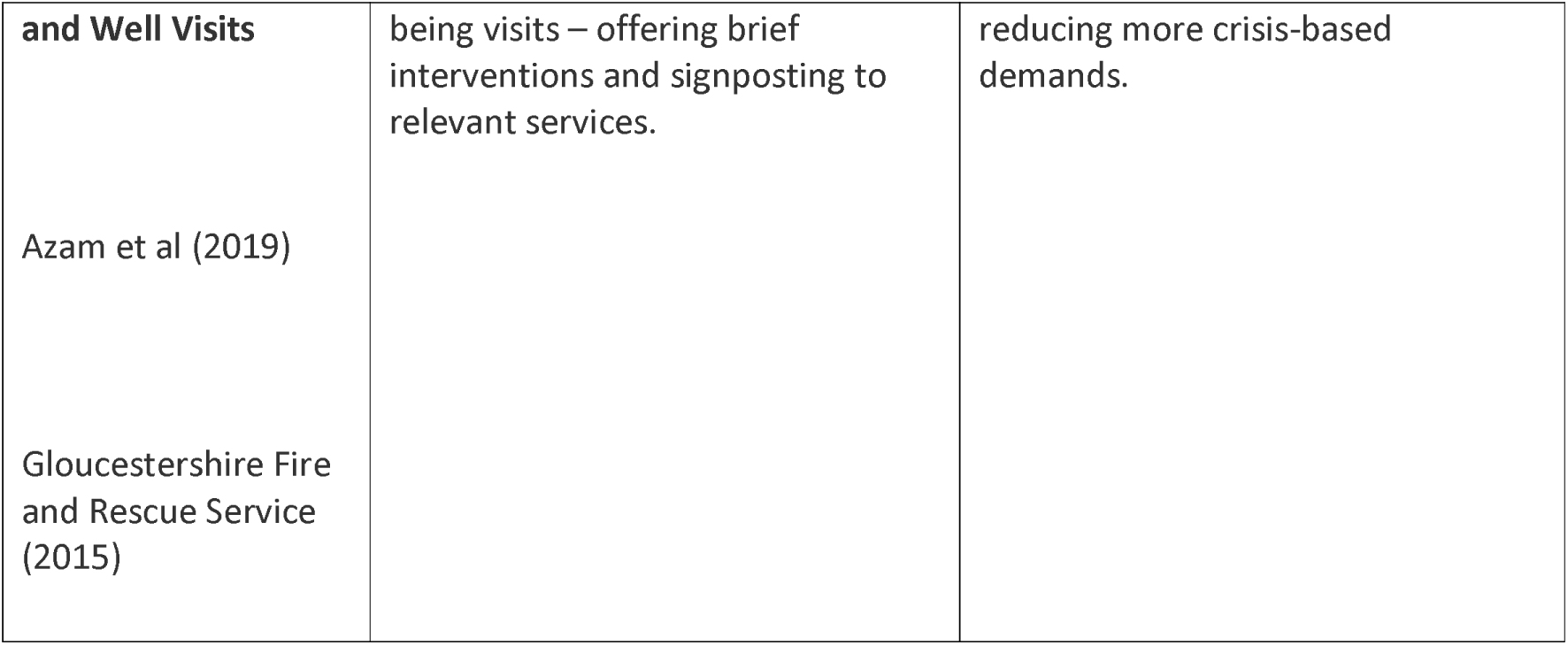
Other Agencies: Definitions and Rationales.

**Table 70.**
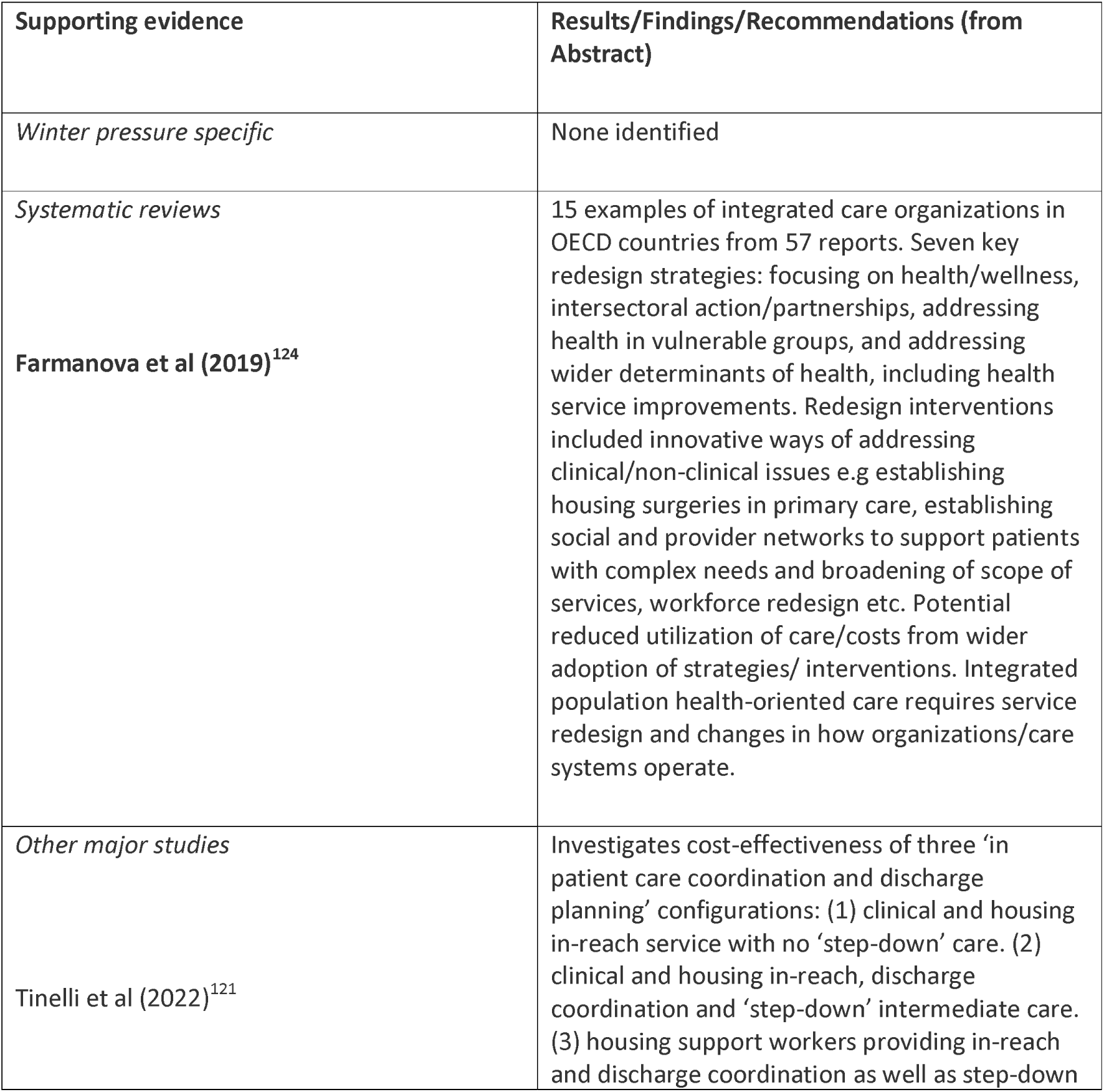
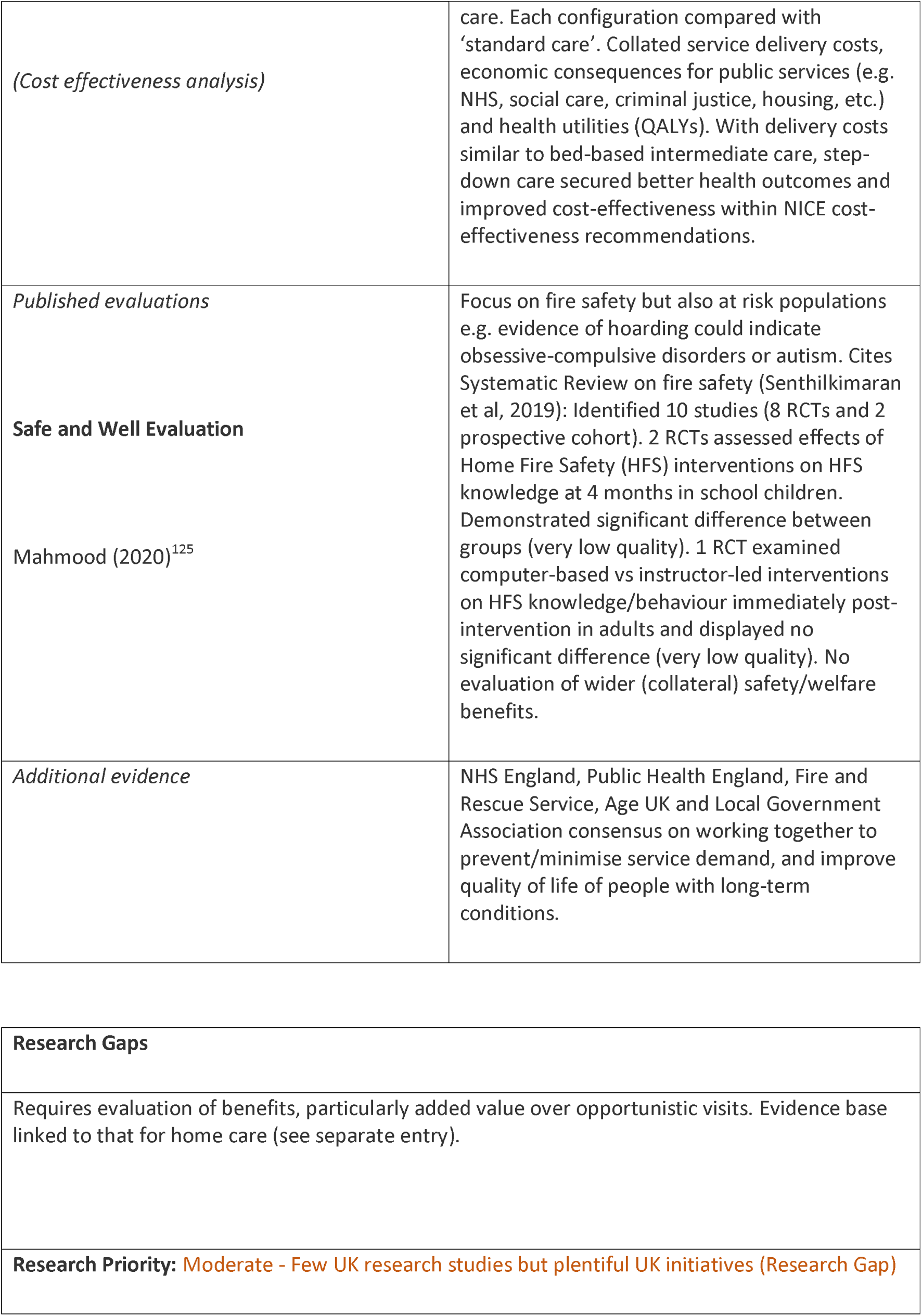
Other Agencies: Interventions and Supporting Evidence.

| Supporting evidence | Results/Findings/Recommendations (from Abstract) |
| --- | --- |
| *Winter pressure specific* | None identified |
| *Systematic reviews*<br>**Farmanova et al (2019)<sup>124</sup>** | 15 examples of integrated care organizations in OECD countries from 57 reports. Seven key redesign strategies: focusing on health/wellness, intersectoral action/partnerships, addressing health in vulnerable groups, and addressing wider determinants of health, including health service improvements. Redesign interventions included innovative ways of addressing clinical/non-clinical issues e.g establishing housing surgeries in primary care, establishing social and provider networks to support patients with complex needs and broadening of scope of services, workforce redesign etc. Potential reduced utilization of care/costs from wider adoption of strategies/ interventions. Integrated population health-oriented care requires service redesign and changes in how organizations/care systems operate. |
| *Other major studies*<br>**Tinelli et al (2022)<sup>121</sup>** | Investigates cost-effectiveness of three ‘in patient care coordination and discharge planning’ configurations: (1) clinical and housing in-reach service with no ‘step-down’ care. (2) clinical and housing in-reach, discharge coordination and ‘step-down’ intermediate care. (3) housing support workers providing in-reach and discharge coordination as well as step-down |
| *(Cost effectiveness analysis)* | care. Each configuration compared with 'standard care'. Collated service delivery costs, economic consequences for public services (e.g. NHS, social care, criminal justice, housing, etc.) and health utilities (QALYs). With delivery costs similar to bed-based intermediate care, step-down care secured better health outcomes and improved cost-effectiveness within NICE cost-effectiveness recommendations. |
| *Published evaluations*<br>**Safe and Well Evaluation**<br>Mahmood (2020)<sup>125</sup> | Focus on fire safety but also at risk populations e.g. evidence of hoarding could indicate obsessive-compulsive disorders or autism. Cites Systematic Review on fire safety (Senthilkimaran et al, 2019): Identified 10 studies (8 RCTs and 2 prospective cohort). 2 RCTs assessed effects of Home Fire Safety (HFS) interventions on HFS knowledge at 4 months in school children. Demonstrated significant difference between groups (very low quality). 1 RCT examined computer-based vs instructor-led interventions on HFS knowledge/behaviour immediately post-intervention in adults and displayed no significant difference (very low quality). No evaluation of wider (collateral) safety/welfare benefits. |
| *Additional evidence* | NHS England, Public Health England, Fire and Rescue Service, Age UK and Local Government Association consensus on working together to prevent/minimise service demand, and improve quality of life of people with long-term conditions. |

|  |
| --- |
| **Research Gaps** |
| Requires evaluation of benefits, particularly added value over opportunistic visits. Evidence base linked to that for home care (see separate entry). |
| **Research Priority:** Moderate - Few UK research studies but plentiful UK initiatives (Research Gap) |

##### CCP - Private Sector

**Table 71.** Private Sector: Definitions and Rationales.

| **Taxonomy heading** | **Brief description/definition** | **Mechanism for minimising winter pressures** |
| --- | --- | --- |
| CCP - Private Sector | Offers alternative response to use of hospital services by using commercial services. Typically involves use of alternative premises and workforce. Although sometimes an expensive option it has been shown to be attractive when compared with hospital utilisation. | Expedites hospital discharge and thereby releases hospital beds for other demand. Often in context of step down services. |
| Example interventions:<br>**Technological solutions**<br>Jones (2018)<sup>8</sup> | Potential technological solutions include rapid diagnostics, genetically-tailored medicine, AI-assisted analysis, and new models of remote/self-administered care. Highlights challenge of adopting innovation across current care system. | Offers alternative venues, including home care, to acute NHS services and alternatives to NHS facilities. Also suggests potential for improved efficiencies. |

**Table 72.**
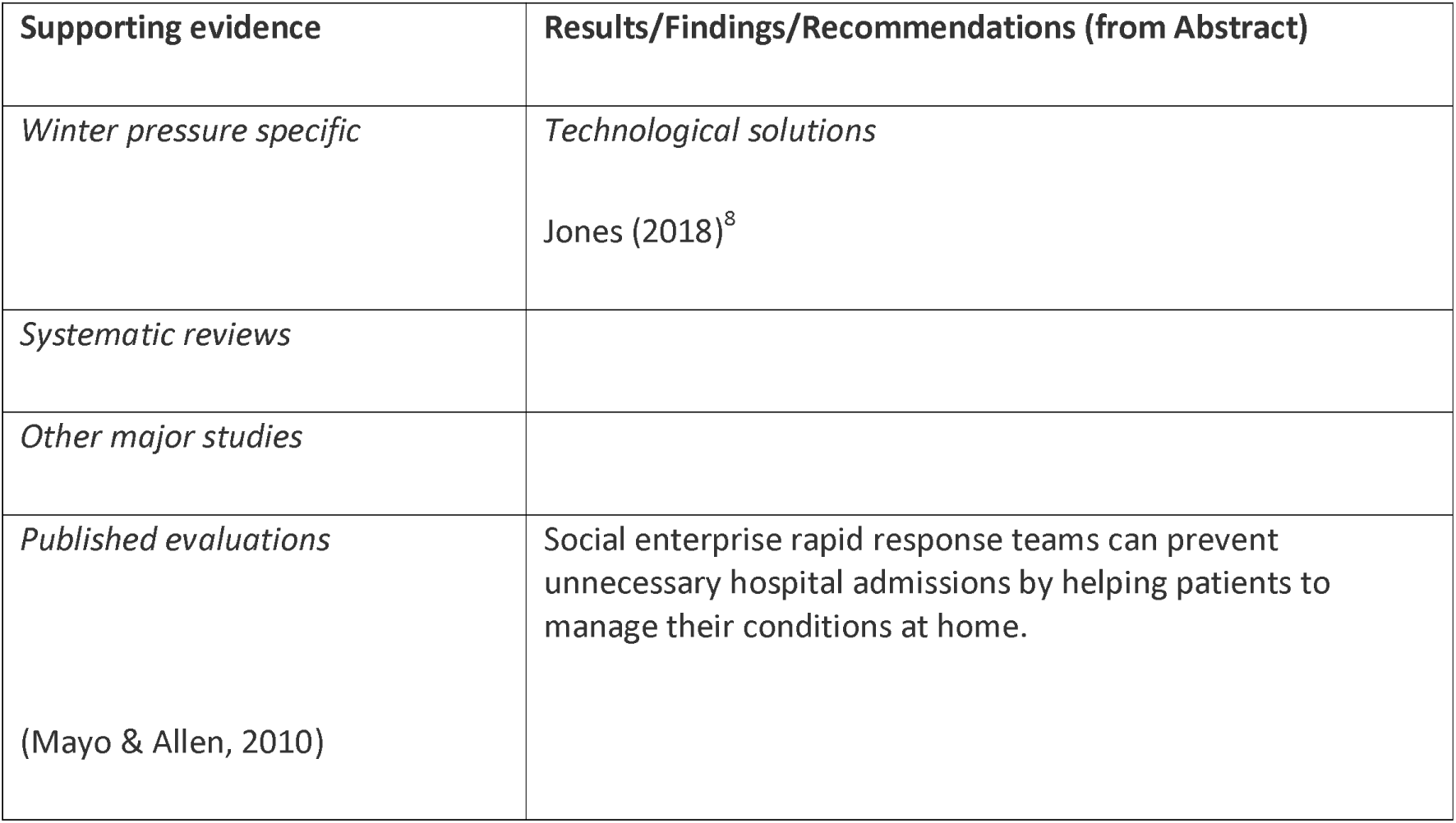
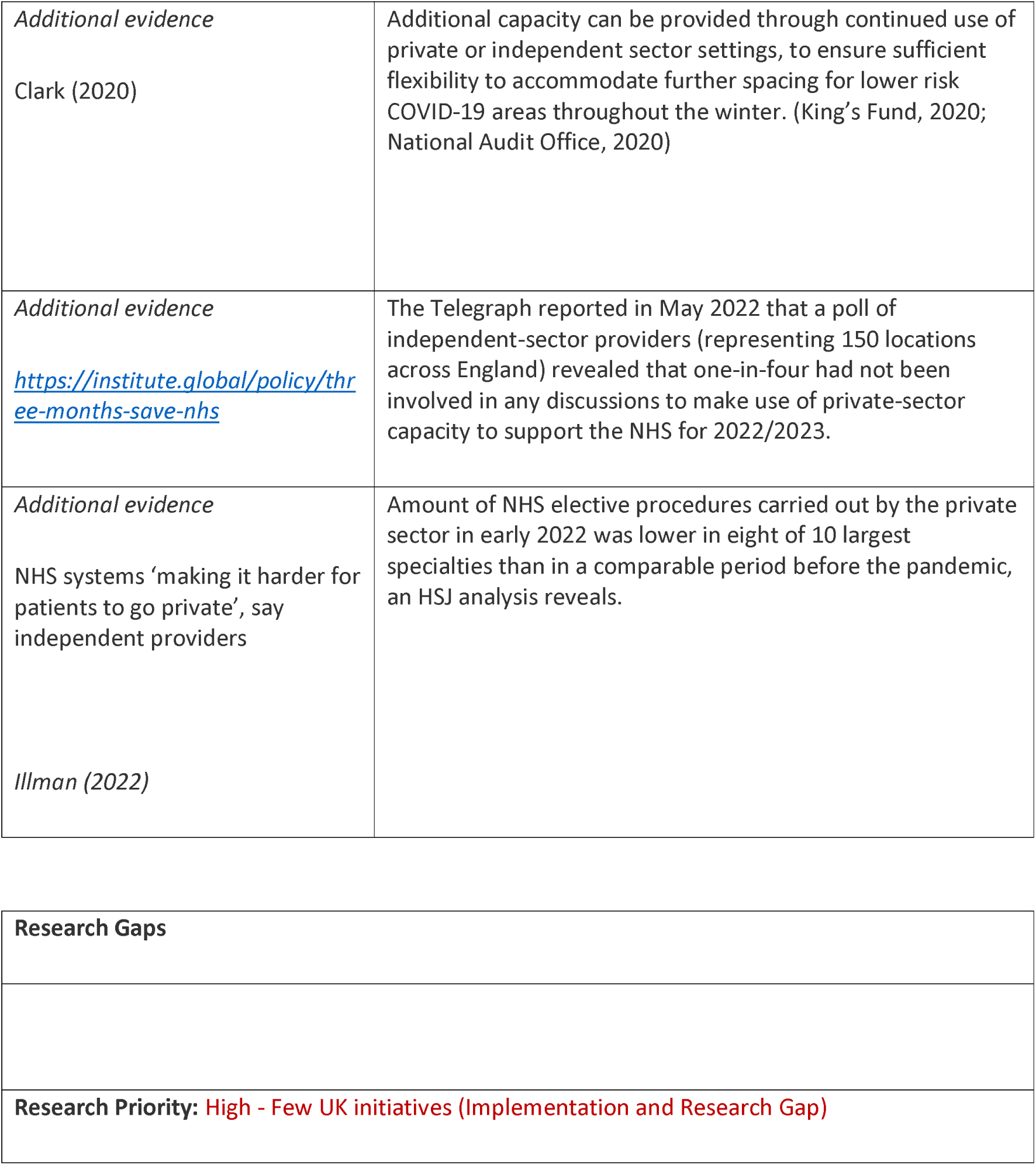
Private Sector: Interventions and Supporting Evidence.

| **Supporting evidence** | **Results/Findings/Recommendations (from Abstract)** |
| --- | --- |
| *Winter pressure specific* | *Technological solutions*<br>Jones (2018)<sup>8</sup> |
| *Systematic reviews* |  |
| *Other major studies* |  |
| *Published evaluations*<br>(Mayo & Allen, 2010) | Social enterprise rapid response teams can prevent unnecessary hospital admissions by helping patients to manage their conditions at home. |
| *Additional evidence*<br>Clark (2020) | Additional capacity can be provided through continued use of private or independent sector settings, to ensure sufficient flexibility to accommodate further spacing for lower risk COVID-19 areas throughout the winter. (King's Fund, 2020; National Audit Office, 2020) |
| *Additional evidence*<br>https://institute.global/policy/three-months-save-nhs | The Telegraph reported in May 2022 that a poll of independent-sector providers (representing 150 locations across England) revealed that one-in-four had not been involved in any discussions to make use of private-sector capacity to support the NHS for 2022/2023. |
| *Additional evidence*<br>NHS systems 'making it harder for patients to go private', say independent providers<br>*Illman (2022)* | Amount of NHS elective procedures carried out by the private sector in early 2022 was lower in eight of 10 largest specialties than in a comparable period before the pandemic, an HSJ analysis reveals. |

|  |
| --- |
| **Research Gaps** |
| **Research Priority: High - Few UK initiatives (Implementation and Research Gap)** |

##### CCP - Social Care

**Table 73.**
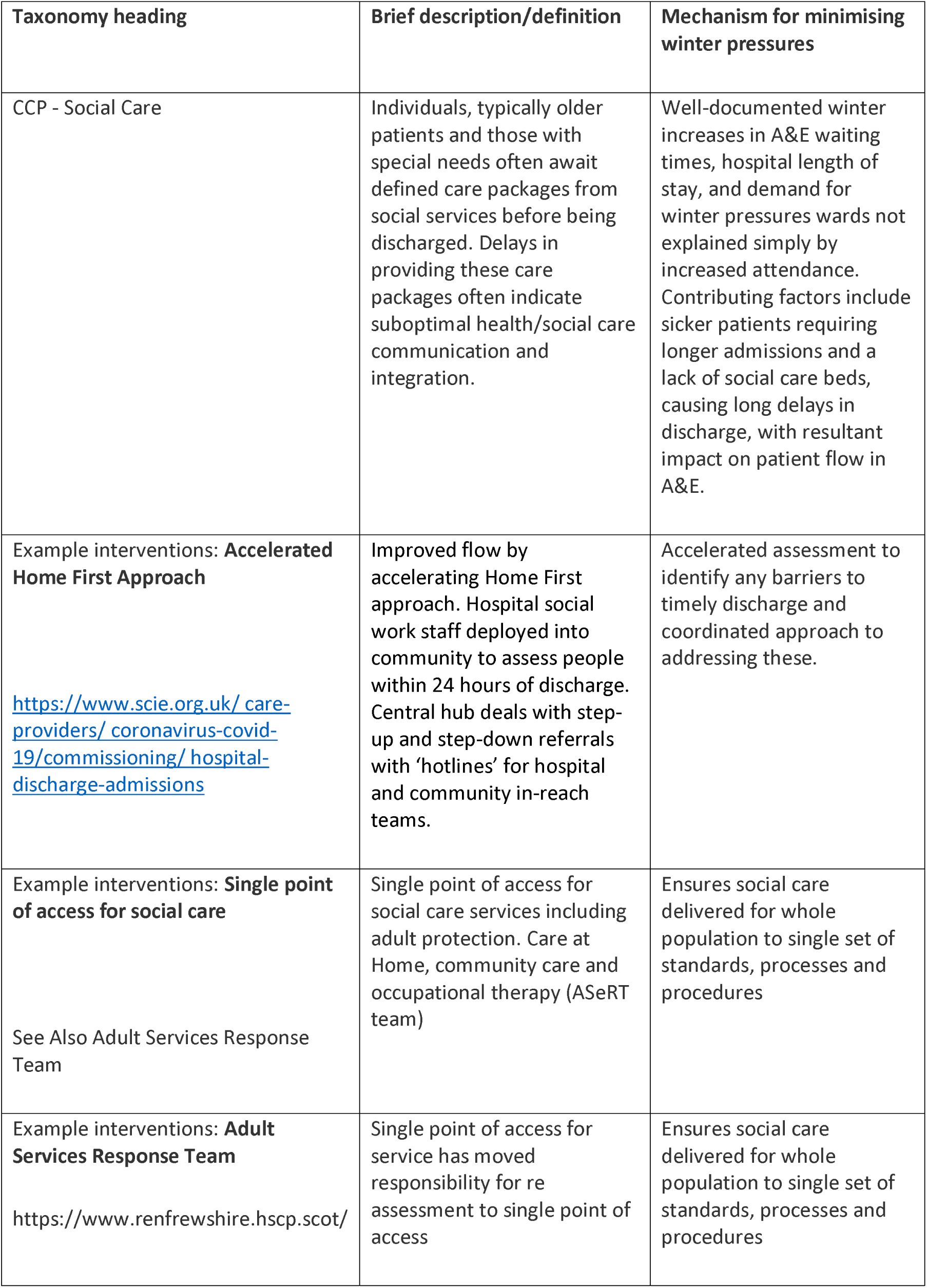
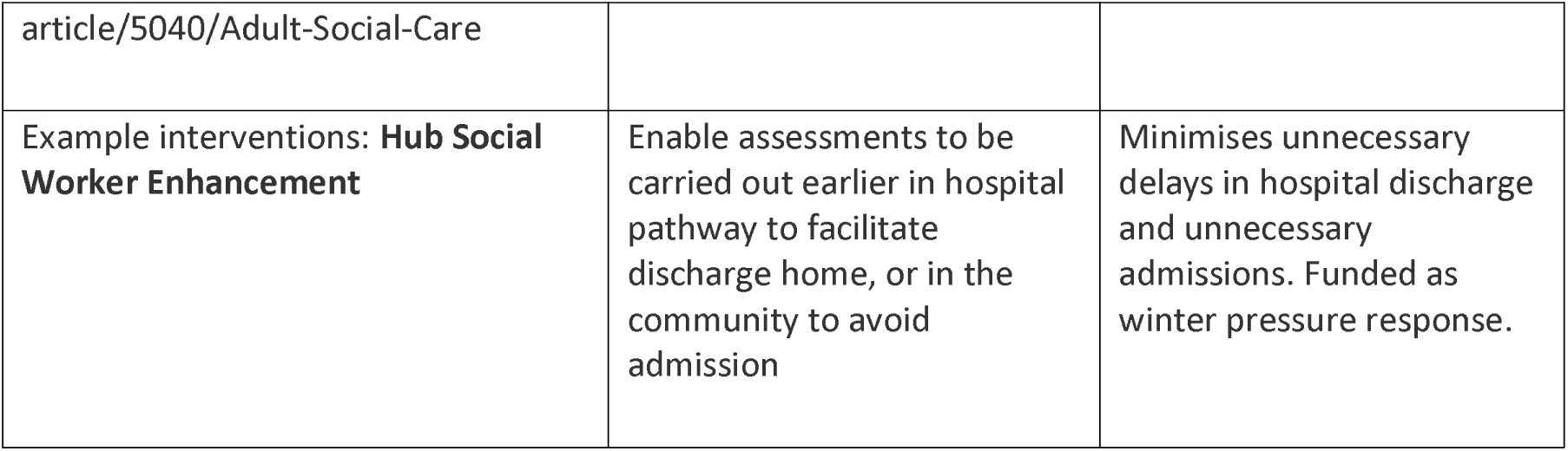
Social Care: Definitions and Rationales.

**Table 74.**
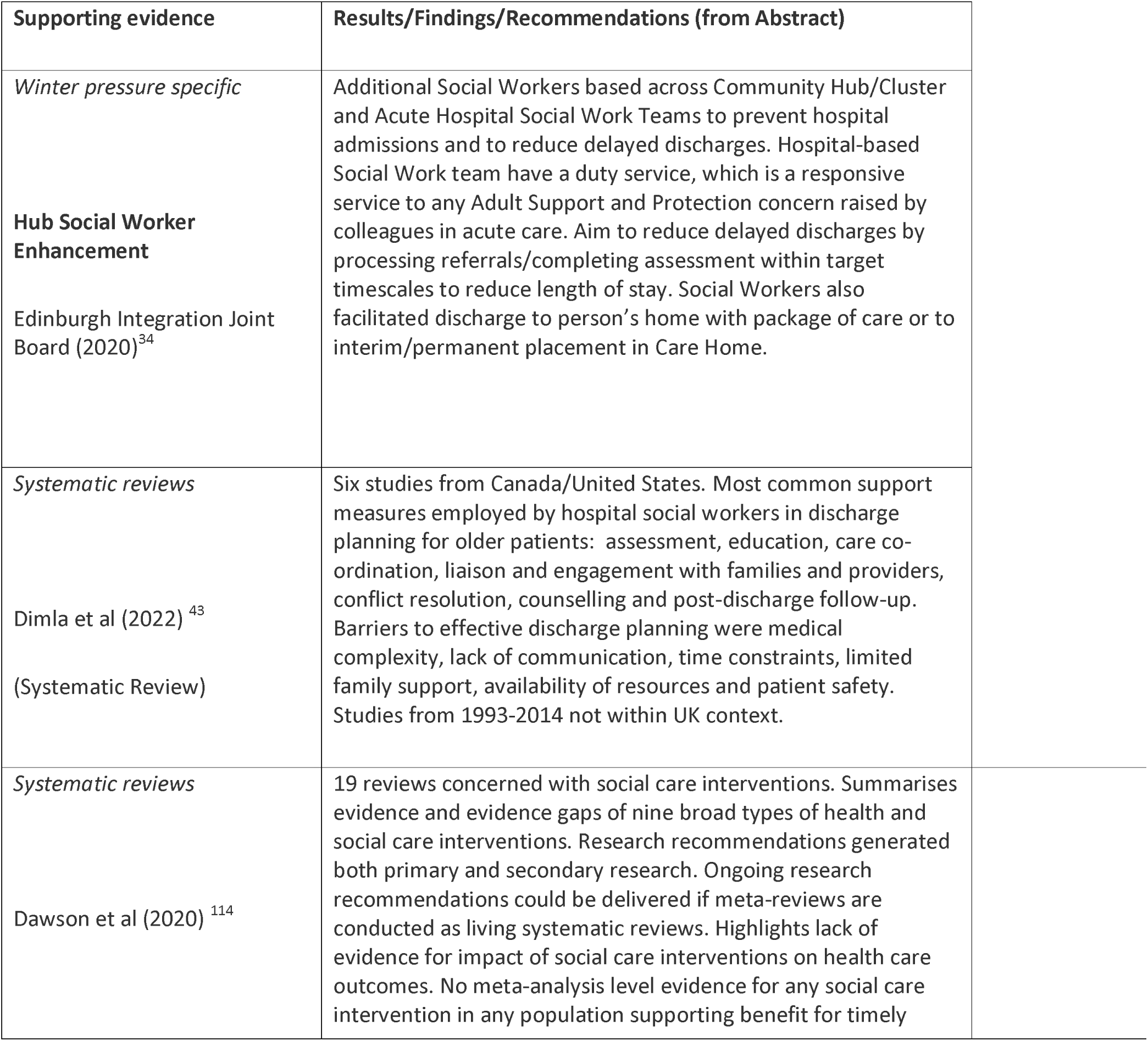
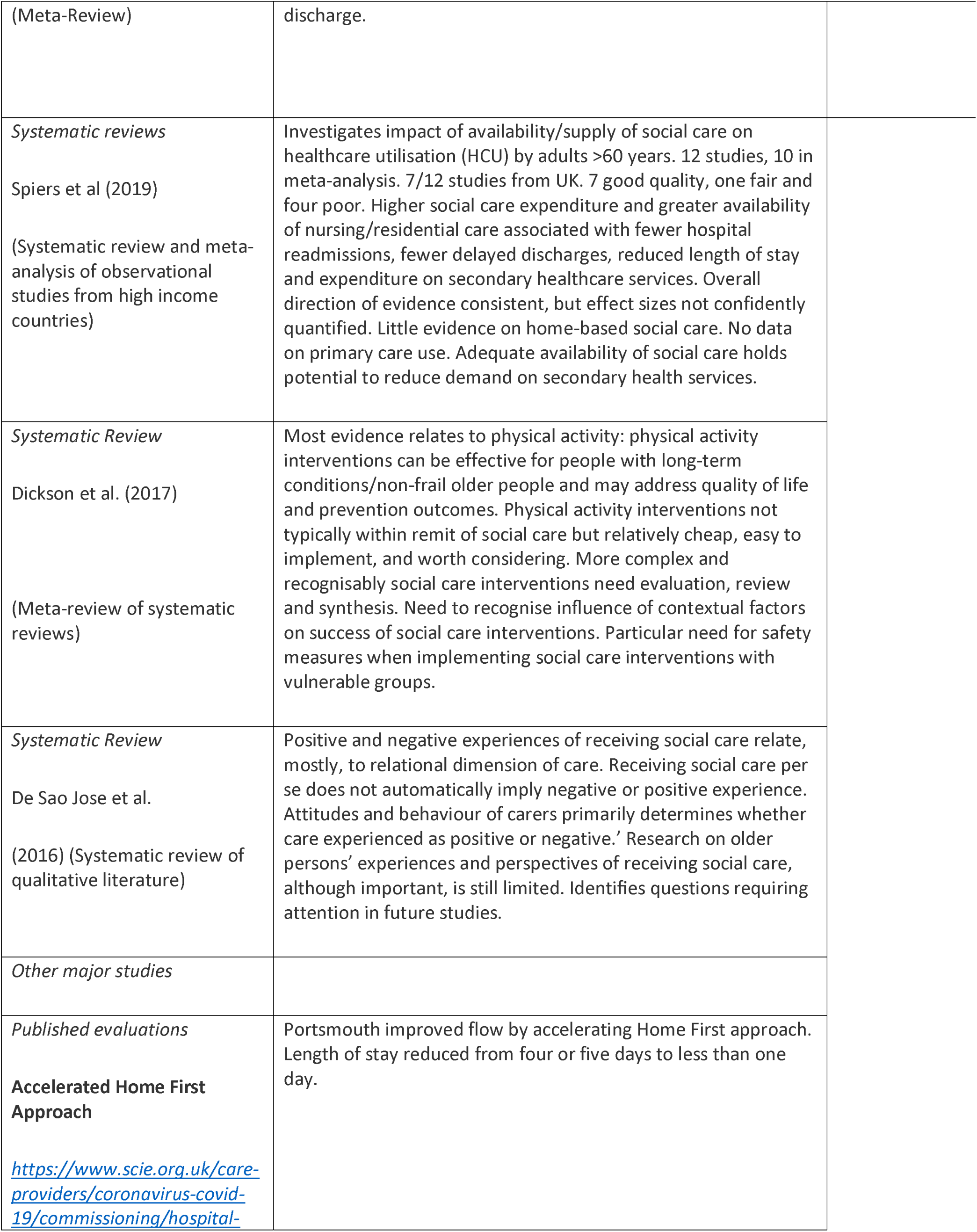
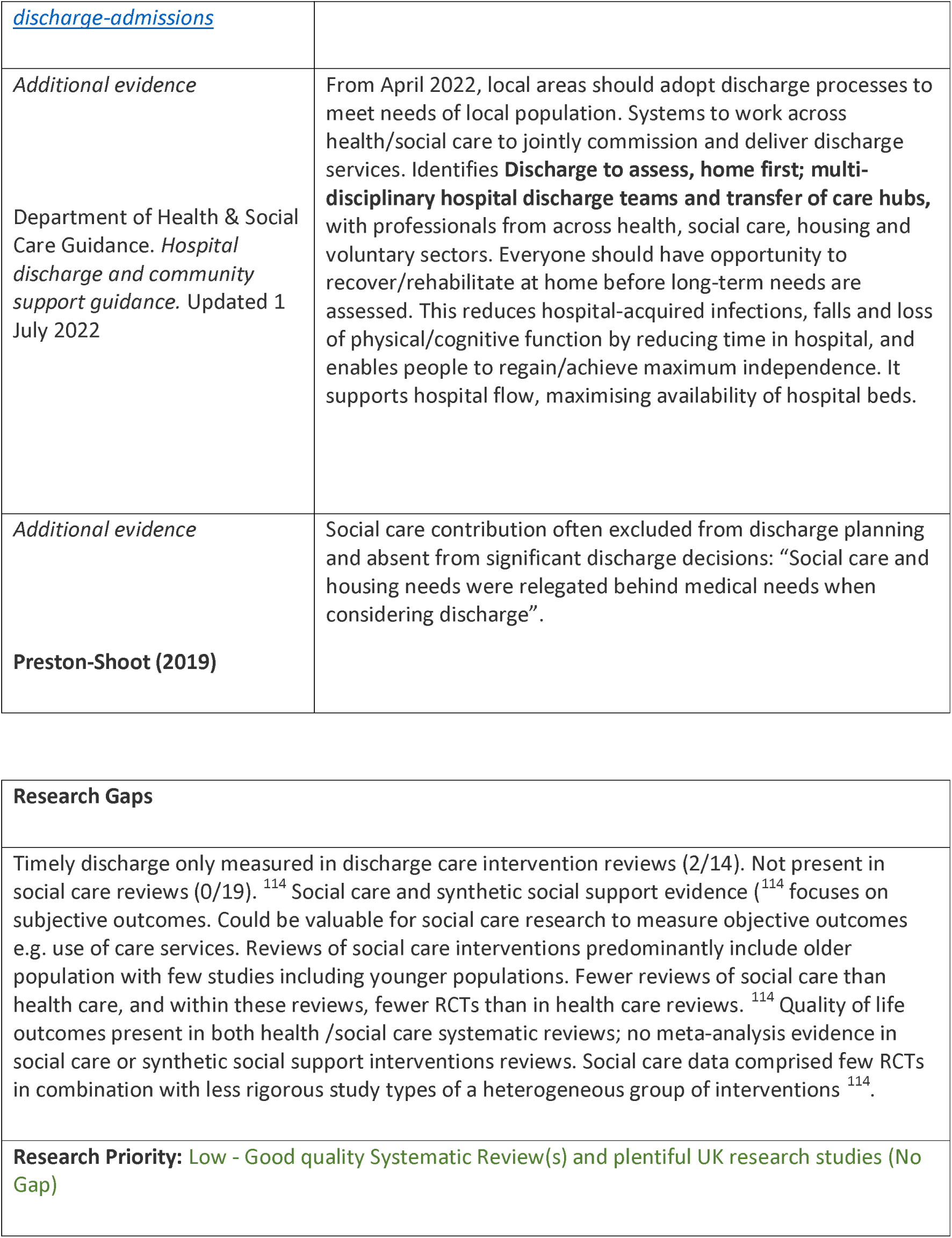
Social Care: Interventions and Supporting Evidence.

| Supporting evidence | Results/Findings/Recommendations (from Abstract) |
| --- | --- |
| *Winter pressure specific*<br>**Hub Social Worker Enhancement**<br>Edinburgh Integration Joint Board (2020)<sup>34</sup> | Additional Social Workers based across Community Hub/Cluster and Acute Hospital Social Work Teams to prevent hospital admissions and to reduce delayed discharges. Hospital-based Social Work team have a duty service, which is a responsive service to any Adult Support and Protection concern raised by colleagues in acute care. Aim to reduce delayed discharges by processing referrals/completing assessment within target timescales to reduce length of stay. Social Workers also facilitated discharge to person's home with package of care or to interim/permanent placement in Care Home. |
| *Systematic reviews*<br>Dimla et al (2022)<sup>43</sup><br>(Systematic Review) | Six studies from Canada/United States. Most common support measures employed by hospital social workers in discharge planning for older patients: assessment, education, care co-ordination, liaison and engagement with families and providers, conflict resolution, counselling and post-discharge follow-up. Barriers to effective discharge planning were medical complexity, lack of communication, time constraints, limited family support, availability of resources and patient safety. Studies from 1993-2014 not within UK context. |
| *Systematic reviews*<br>Dawson et al (2020)<sup>114</sup> | 19 reviews concerned with social care interventions. Summarises evidence and evidence gaps of nine broad types of health and social care interventions. Research recommendations generated both primary and secondary research. Ongoing research recommendations could be delivered if meta-reviews are conducted as living systematic reviews. Highlights lack of evidence for impact of social care interventions on health care outcomes. No meta-analysis level evidence for any social care intervention in any population supporting benefit for timely |
| (Meta-Review) | discharge. |
| *Systematic reviews*<br>Spiers et al (2019)<br>(Systematic review and meta-analysis of observational studies from high income countries) | Investigates impact of availability/supply of social care on healthcare utilisation (HCU) by adults >60 years. 12 studies, 10 in meta-analysis. 7/12 studies from UK. 7 good quality, one fair and four poor. Higher social care expenditure and greater availability of nursing/residential care associated with fewer hospital readmissions, fewer delayed discharges, reduced length of stay and expenditure on secondary healthcare services. Overall direction of evidence consistent, but effect sizes not confidently quantified. Little evidence on home-based social care. No data on primary care use. Adequate availability of social care holds potential to reduce demand on secondary health services. |
| *Systematic Review*<br>Dickson et al. (2017)<br>(Meta-review of systematic reviews) | Most evidence relates to physical activity: physical activity interventions can be effective for people with long-term conditions/non-frail older people and may address quality of life and prevention outcomes. Physical activity interventions not typically within remit of social care but relatively cheap, easy to implement, and worth considering. More complex and recognisably social care interventions need evaluation, review and synthesis. Need to recognise influence of contextual factors on success of social care interventions. Particular need for safety measures when implementing social care interventions with vulnerable groups. |
| *Systematic Review*<br>De Sao Jose et al.<br>(2016) (Systematic review of qualitative literature) | Positive and negative experiences of receiving social care relate, mostly, to relational dimension of care. Receiving social care per se does not automatically imply negative or positive experience. Attitudes and behaviour of carers primarily determines whether care experienced as positive or negative.’ Research on older persons’ experiences and perspectives of receiving social care, although important, is still limited. Identifies questions requiring attention in future studies. |
| *Other major studies* |  |
| *Published evaluations*<br>**Accelerated Home First Approach**<br>https://www.scie.org.uk/care-providers/coronavirus-covid-19/commissioning/hospital- | Portsmouth improved flow by accelerating Home First approach. Length of stay reduced from four or five days to less than one day. |
| *discharge-admissions* |  |
| *Additional evidence*<br>Department of Health & Social Care Guidance. *Hospital discharge and community support guidance*. Updated 1 July 2022 | From April 2022, local areas should adopt discharge processes to meet needs of local population. Systems to work across health/social care to jointly commission and deliver discharge services. Identifies **Discharge to assess, home first; multi-disciplinary hospital discharge teams and transfer of care hubs**, with professionals from across health, social care, housing and voluntary sectors. Everyone should have opportunity to recover/rehabilitate at home before long-term needs are assessed. This reduces hospital-acquired infections, falls and loss of physical/cognitive function by reducing time in hospital, and enables people to regain/achieve maximum independence. It supports hospital flow, maximising availability of hospital beds. |
| *Additional evidence*<br>**Preston-Shoot (2019)** | Social care contribution often excluded from discharge planning and absent from significant discharge decisions: "Social care and housing needs were relegated behind medical needs when considering discharge". |

|  |
| --- |
| **Research Gaps** |
| Timely discharge only measured in discharge care intervention reviews (2/14). Not present in social care reviews (0/19).<sup>114</sup> Social care and synthetic social support evidence<sup>114</sup> focuses on subjective outcomes. Could be valuable for social care research to measure objective outcomes e.g. use of care services. Reviews of social care interventions predominantly include older population with few studies including younger populations. Fewer reviews of social care than health care, and within these reviews, fewer RCTs than in health care reviews.<sup>114</sup> Quality of life outcomes present in both health /social care systematic reviews; no meta-analysis evidence in social care or synthetic social support interventions reviews. Social care data comprised few RCTs in combination with less rigorous study types of a heterogeneous group of interventions<sup>114</sup>. |
| **Research Priority:** Low - Good quality Systematic Review(s) and plentiful UK research studies (No Gap) |

##### CCP - Voluntary Services

**Table 75.**
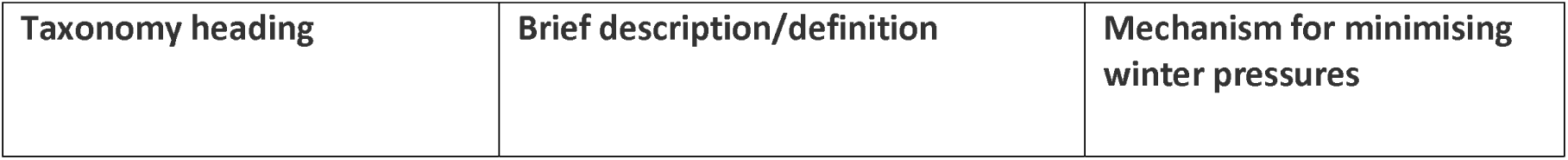
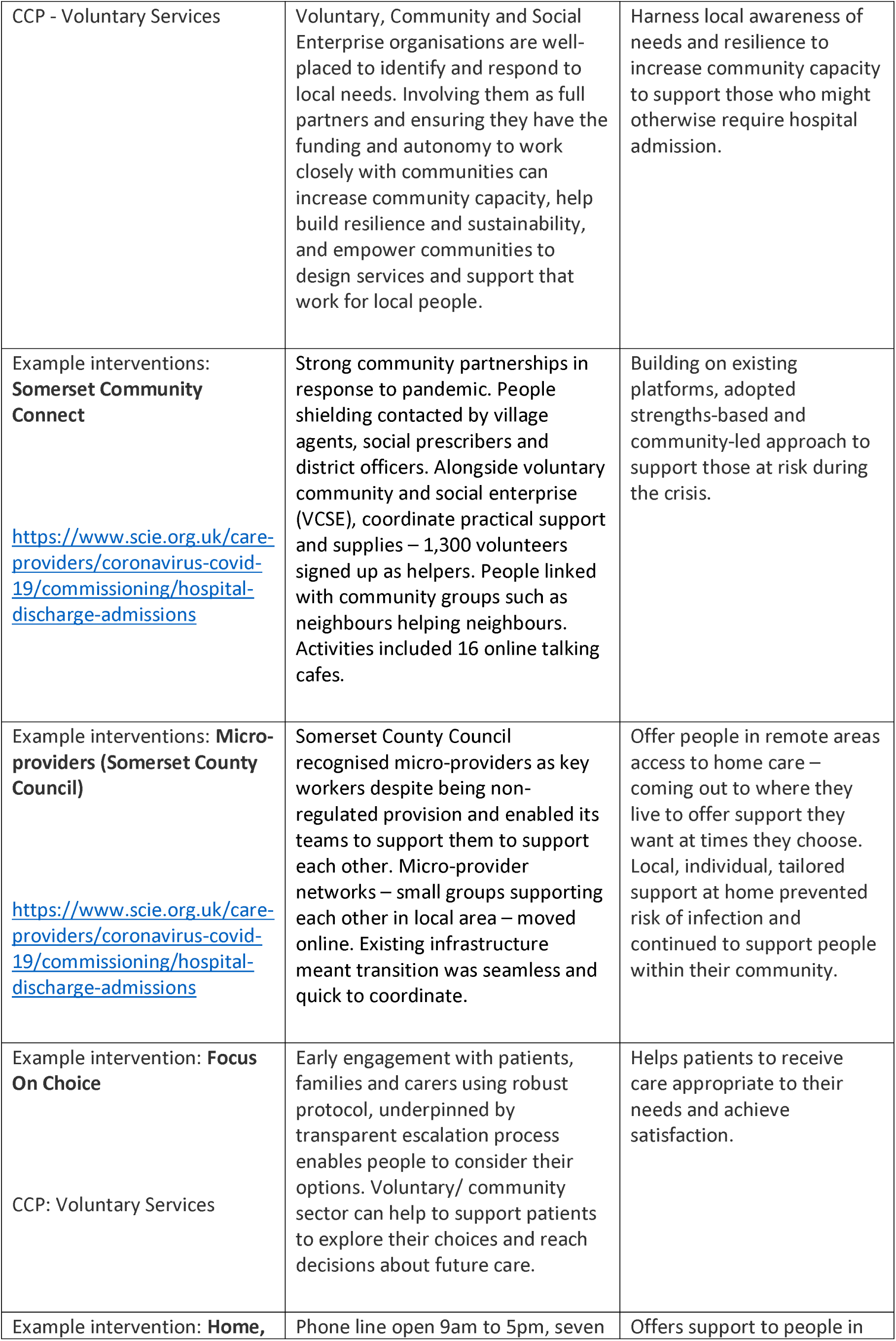
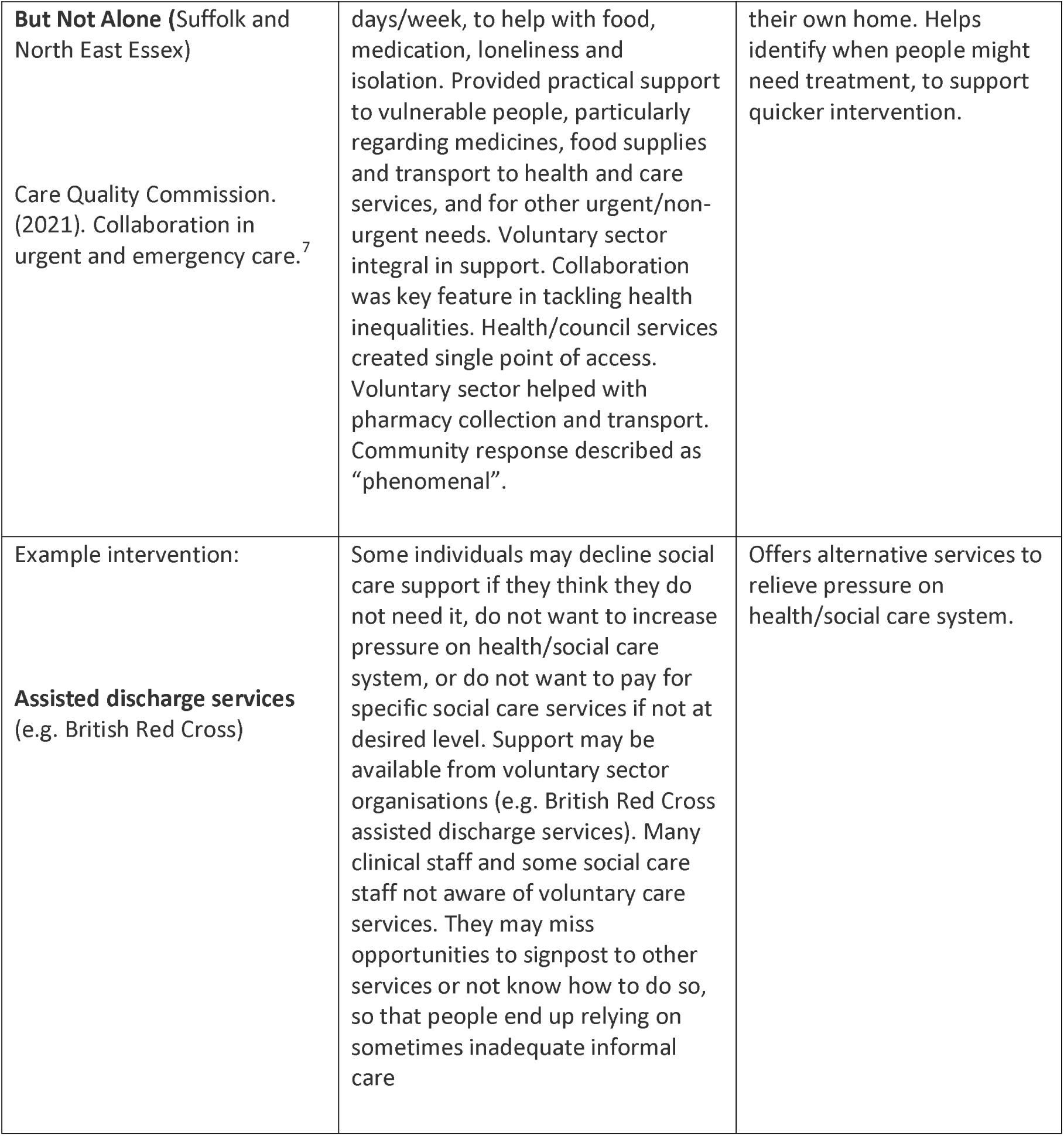
Voluntary Services: Definitions and Rationales.

| **Taxonomy heading** | **Brief description/definition** | **Mechanism for minimising winter pressures** |
| --- | --- | --- |
| CCP - Voluntary Services | Voluntary, Community and Social Enterprise organisations are well-placed to identify and respond to local needs. Involving them as full partners and ensuring they have the funding and autonomy to work closely with communities can increase community capacity, help build resilience and sustainability, and empower communities to design services and support that work for local people. | Harness local awareness of needs and resilience to increase community capacity to support those who might otherwise require hospital admission. |
| Example interventions: **Somerset Community Connect**<br>https://www.scie.org.uk/care-providers/coronavirus-covid-19/commissioning/hospital-discharge-admissions | Strong community partnerships in response to pandemic. People shielding contacted by village agents, social prescribers and district officers. Alongside voluntary community and social enterprise (VCSE), coordinate practical support and supplies – 1,300 volunteers signed up as helpers. People linked with community groups such as neighbours helping neighbours. Activities included 16 online talking cafes. | Building on existing platforms, adopted strengths-based and community-led approach to support those at risk during the crisis. |
| Example interventions: **Micro-providers (Somerset County Council)**<br>https://www.scie.org.uk/care-providers/coronavirus-covid-19/commissioning/hospital-discharge-admissions | Somerset County Council recognised micro-providers as key workers despite being non-regulated provision and enabled its teams to support them to support each other. Micro-provider networks – small groups supporting each other in local area – moved online. Existing infrastructure meant transition was seamless and quick to coordinate. | Offer people in remote areas access to home care – coming out to where they live to offer support they want at times they choose. Local, individual, tailored support at home prevented risk of infection and continued to support people within their community. |
| Example intervention: **Focus On Choice**<br>CCP: Voluntary Services | Early engagement with patients, families and carers using robust protocol, underpinned by transparent escalation process enables people to consider their options. Voluntary/ community sector can help to support patients to explore their choices and reach decisions about future care. | Helps patients to receive care appropriate to their needs and achieve satisfaction. |
| Example intervention: **Home,** | Phone line open 9am to 5pm, seven | Offers support to people in |
| **But Not Alone** (Suffolk and North East Essex)<br>Care Quality Commission. (2021). Collaboration in urgent and emergency care.<sup>7</sup> | days/week, to help with food, medication, loneliness and isolation. Provided practical support to vulnerable people, particularly regarding medicines, food supplies and transport to health and care services, and for other urgent/non-urgent needs. Voluntary sector integral in support. Collaboration was key feature in tackling health inequalities. Health/council services created single point of access. Voluntary sector helped with pharmacy collection and transport. Community response described as “phenomenal”. | their own home. Helps identify when people might need treatment, to support quicker intervention. |
| Example intervention:<br>**Assisted discharge services** (e.g. British Red Cross) | Some individuals may decline social care support if they think they do not need it, do not want to increase pressure on health/social care system, or do not want to pay for specific social care services if not at desired level. Support may be available from voluntary sector organisations (e.g. British Red Cross assisted discharge services). Many clinical staff and some social care staff not aware of voluntary care services. They may miss opportunities to signpost to other services or not know how to do so, so that people end up relying on sometimes inadequate informal care | Offers alternative services to relieve pressure on health/social care system. |

**Table 76.**
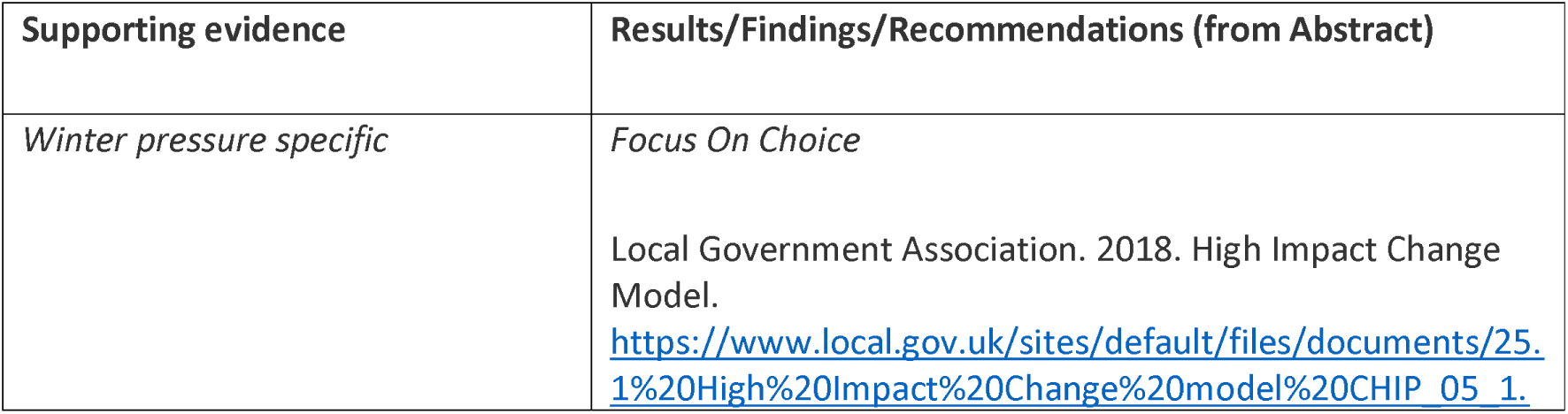
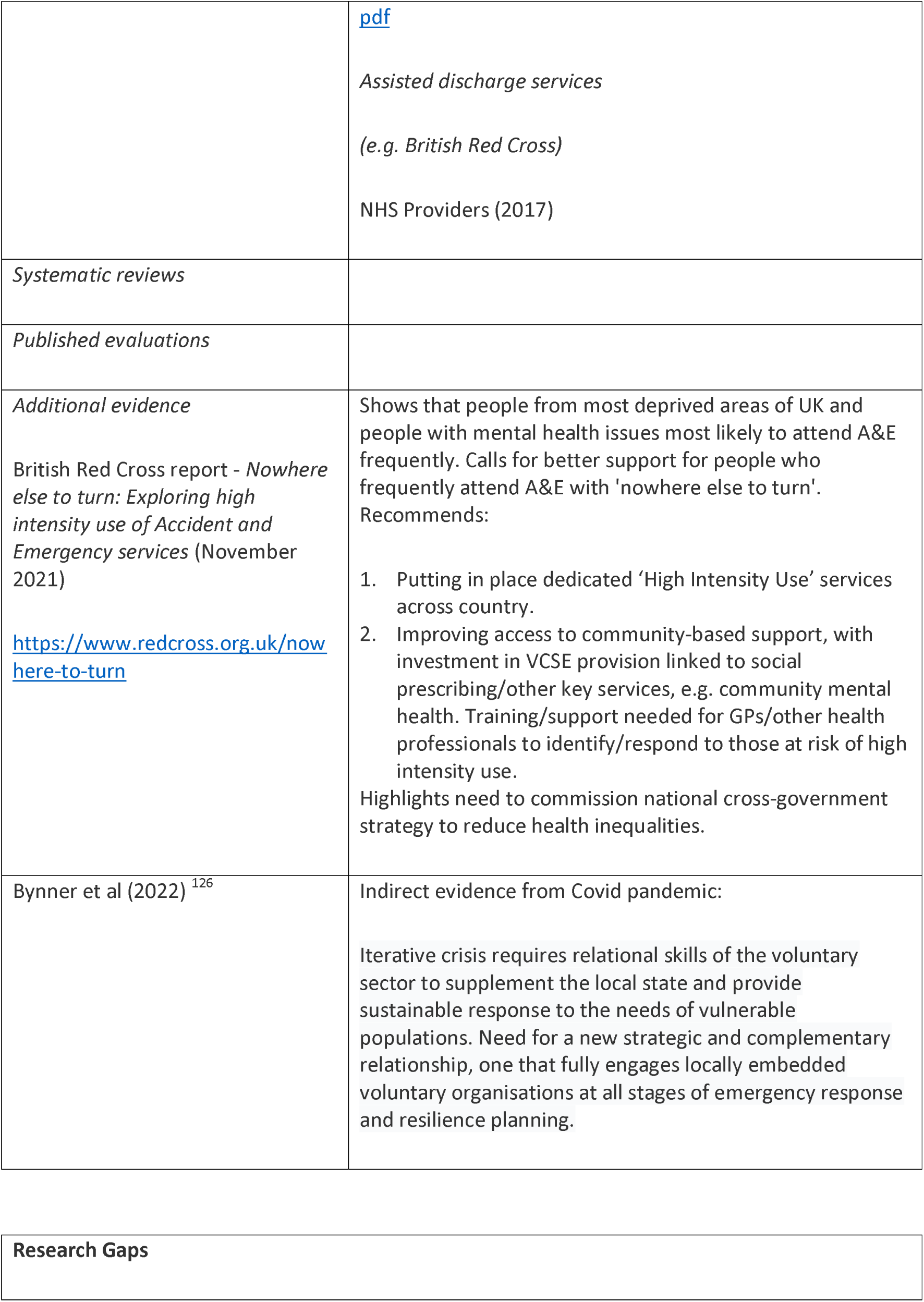
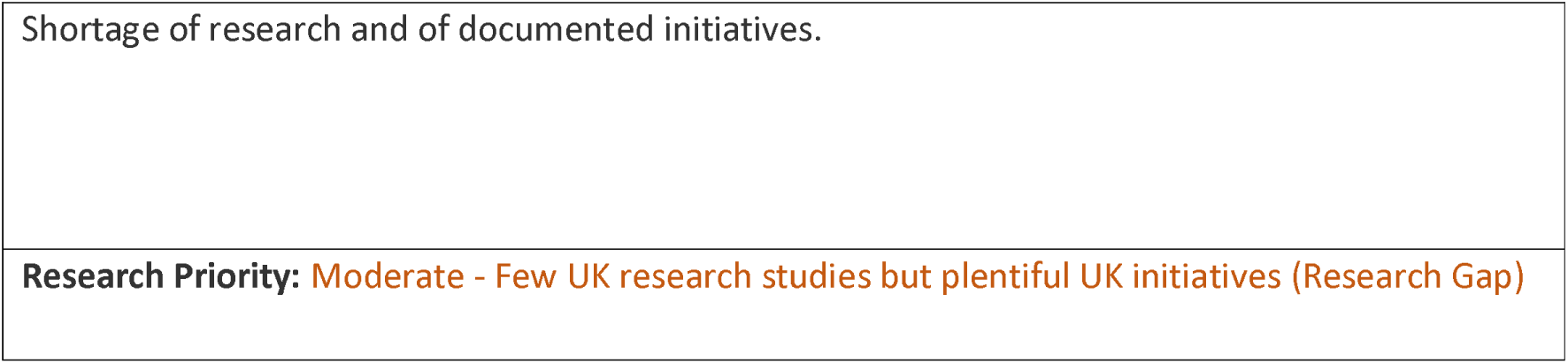
Voluntary Services: Interventions and Supporting Evidence.

Within the Changing Community Provision taxonomy section, comprised of three subsections and numerous interventions, we identified variable numbers of initiatives or research studies. Some areas such as hospital at home are well explored. Other approaches such as virtual wards are already stimulating their own initiatives and research agenda, motivated in part by significant growth in these remote initiatives during the pandemic. Other sectors such as the voluntary agencies and private sector are less frequently reported and even less frequently researched. Initiatives reported in this section are reported and evaluated in the short-term with little consideration of longer-term sustainment or rigorous evaluation. The Community sector has always been a “poor relation” compared with the acute sector and has neither the resources nor infrastructure to tackle the complete research agenda required by such a whole system wicked problem. Further research studies could seek to undertake cross-cutting research along clinical pathways (e.g. primary care- acute care-social care) but this may require an integration of research strategies and research funding to mirror the integration of care taking place in the health sector itself.

### Integrated Care

findings for the Integrated Care (IC) are presented in detail in Tables 77 to 82 below and associated summary paragraphs. Probably even more than any other section of the taxonomy we found considerable overlap both between integrated care interventions and other interventions. As mentioned above a focus on “delivery site” elsewhere in the taxonomy has meant that the section on integrated care tends to focus on initiatives that are distinctively branded as “integrated”. Furthermore, this particular category within the taxonomy, a specific focus identified during the commissioning of the review, is distinctive in that it operates across multiple levels; from integration at an organisation level, through shared pathways of care to front-line integration of health, social care and voluntary services staff within individual teams. As a consequence, coverage of this category is notably heterogeneous. As a combined front for research activity it would particularly benefit from exploration of mechanisms to establish whether these operate at multiple levels as a feature of integration or whether they operate exclusively within a single level or strata of the organisation.

**Table 77.** Integrated Care: Definitions and Rationales.

| Taxonomy heading | Brief description/definition | Mechanism for minimising winter pressures |
| --- | --- | --- |
| IC - Integrated Care | Involves partnerships of organisations that come together to plan and deliver joined up health/care services, and improve lives of people who live/work in their area. | Improved communication between health and care services to speed up admission/discharge and to effect hospital avoidance. |
| Example interventions:<br>**Comprehensive systems approach (Somerset)**<br>https://www.scie.org.uk/care-providers/coronavirus-covid-19/commissioning/hospital-discharge-admissions | **Comprehensive systems approach** links hospital discharge and avoidance with Community Connect to maximise local support and connections. | Outcomes through rapid response enhanced by locality neighbourhood approach and innovative micro-providers delivering flexible support. |
| Example intervention:<br>**Developing capacity and demand model for out of hospital care**<br>https://www.scie.org.uk/care-providers/coronavirus-covid-19/commissioning/hospital-discharge-admissions<br>https://www.local.gov.uk/publications/developing-capacity-and-demand-model-out-hospital-care | Shares learning from seven health/care communities (July 2020-June 2021). Challenges (i) bringing together health and social care data to understand demand and pressures on commissioned services, (ii) availability and supply of domiciliary care. Workshops organised to explore challenges and inform local action plans. | Demonstrated that better-staffed community hospitals can help get older people back to their own homes; and support for care homes by therapists and nurses can increase percentage of older people then returning home. |
| Example interventions:<br>**High Impact User Group (HIU)**<br>**(Bristol Royal Infirmary (BRI))**<br>Care Quality Commission | Multidisciplinary group works with ED frequent/ “high impact” attenders, e.g. violence and aggression. HIU aim to manage top 100 frequent attenders. Team of ED matron, consultant, psychiatry liaison nurse, homeless health team, drug/alcohol nurse, and primary care, with input from police, ambulance and other specialties. | Creates individualised support plans and behavioural contracts and signposts people to other services to support them. |
| **Hull Integrated Care Centre**<br>(Charles, 2018) <sup>109</sup><br>https://operabeds.com/blogs/news/hull-integrated-care-centre | Facility to support integrated out-of-hospital care across health, social care, social housing, voluntary sector and other organisations. Hub initially to focus on supporting frail individuals identified through screening in primary care. | Uses integrated single response model to facilitate appropriate navigation of health/care system. |
| Example intervention:<br>**Integrated Care Across Northamptonshire**<br>Care Quality Commission. (2021). Collaboration in urgent and emergency care. <sup>7</sup><br>https://www.cqc.org.uk/publications/themes-care/collaboration-urgent-emergency-care#adult-social-care | Programme of transformation work driven by Northamptonshire Health and Care Partnership to improve quality of care and achieve best possible health/wellbeing outcomes for older people across the county. | Three key areas make up iCAN programme:<br>**Community Resilience:** fully supported to live independently within community as older person.<br>**Frailty Escalation and Front Door:** assessed swiftly and treated effectively so older person remains independent.<br>**Flow and Grip:** fully aware of when they will leave hospital and available support once they are home. |
| Example interventions:<br>**Integrated primary and secondary care pharmacy team**<br>NHS Improvement and NHS England (2018)<sup>127</sup>; Wilkinson (2018)<sup>60</sup>. | In 2018, NHS England published guidance on steps hospitals should take to use pharmacists to tackle winter pressures. Recommendations included putting pharmacy professionals on hospital wards to manage medicines and help discharge patients; placing pharmacists in emergency departments to prescribe medicines before patients even reach wards; and extending hours of pharmacy services to support hospital teams. | Facilitating speed of discharge by removing barriers in making medicines available prior to discharge. |
| Example interventions:<br>**Trusted Assessors** | Local health economies use 'trusted assessor models':<br>• University Hospital Coventry and Warwickshire NHS Trust's | Enables one person to be the single point of contact for multiple teams to co-ordinate and make appropriate discharge suggestions to suit |
| (Charles et al, 2018) <sup>109</sup> | Integrated Discharge Team are trusted assessors for all residential and housing with care units in Coventry. IDT can also restart packages of social care, if no change in people's needs.<br>• Medway Community Healthcare staff can restart packages of care if care requirements unchanged. | each patient's needs. |

**Table 78.**
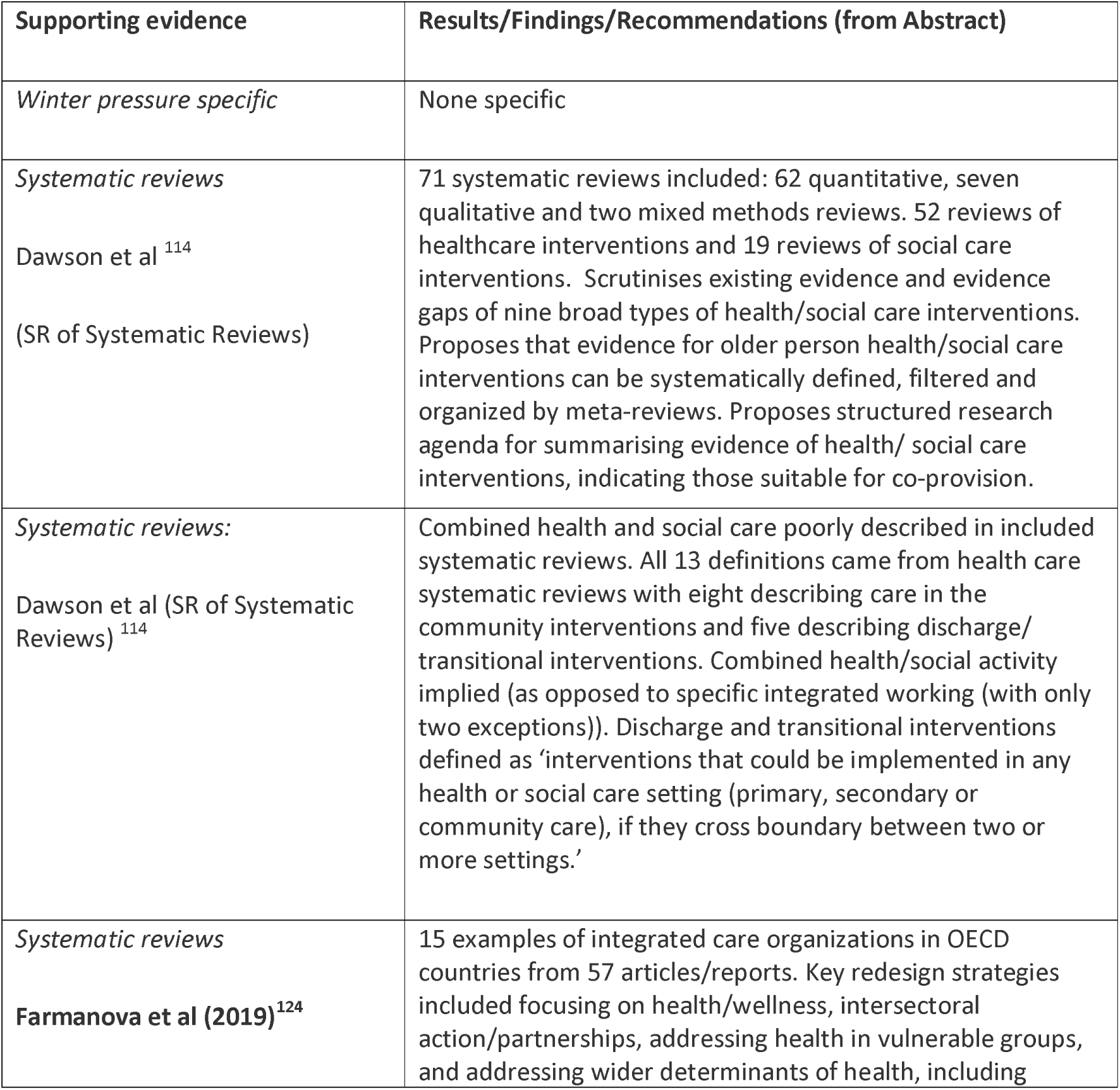
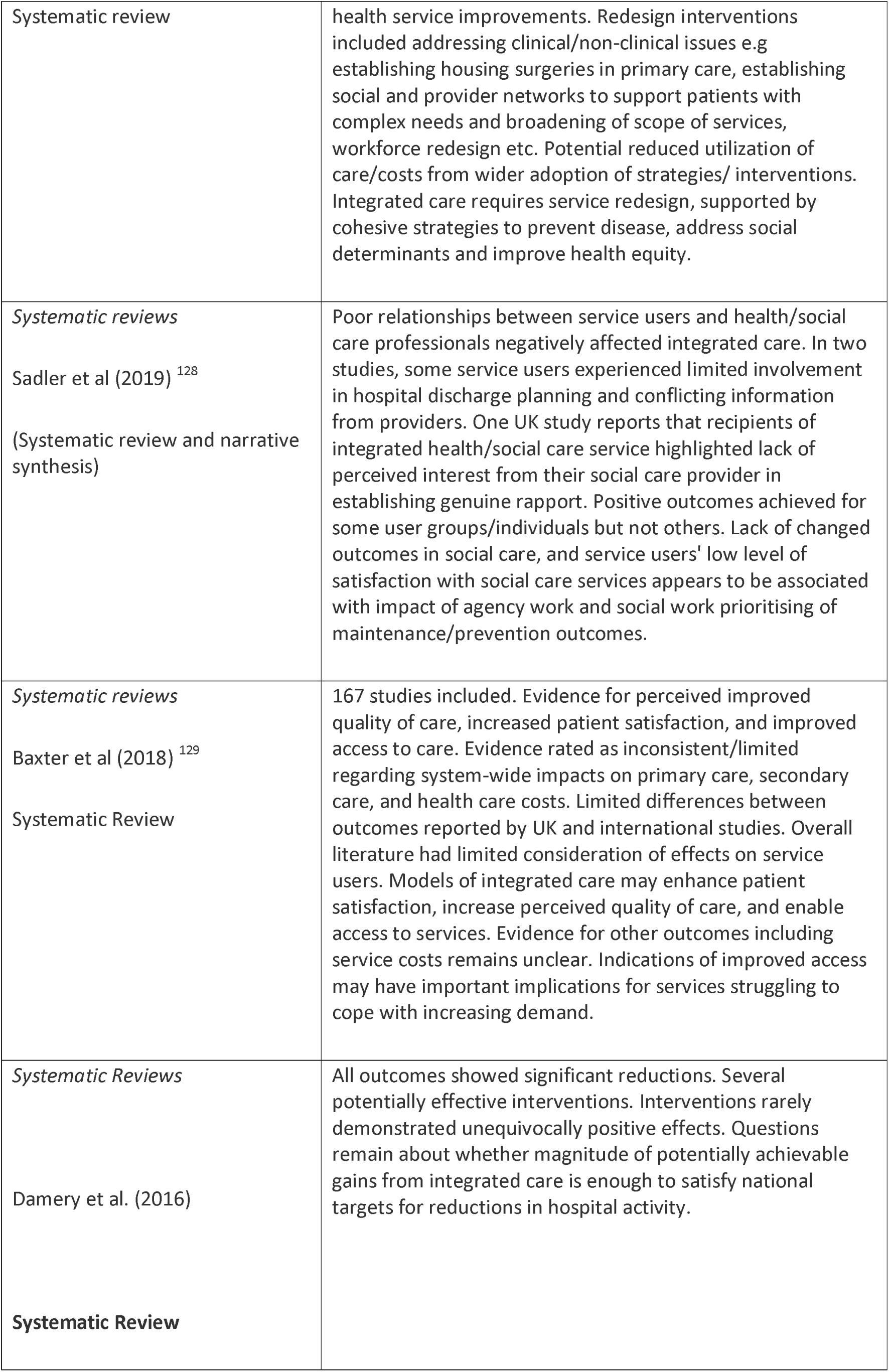
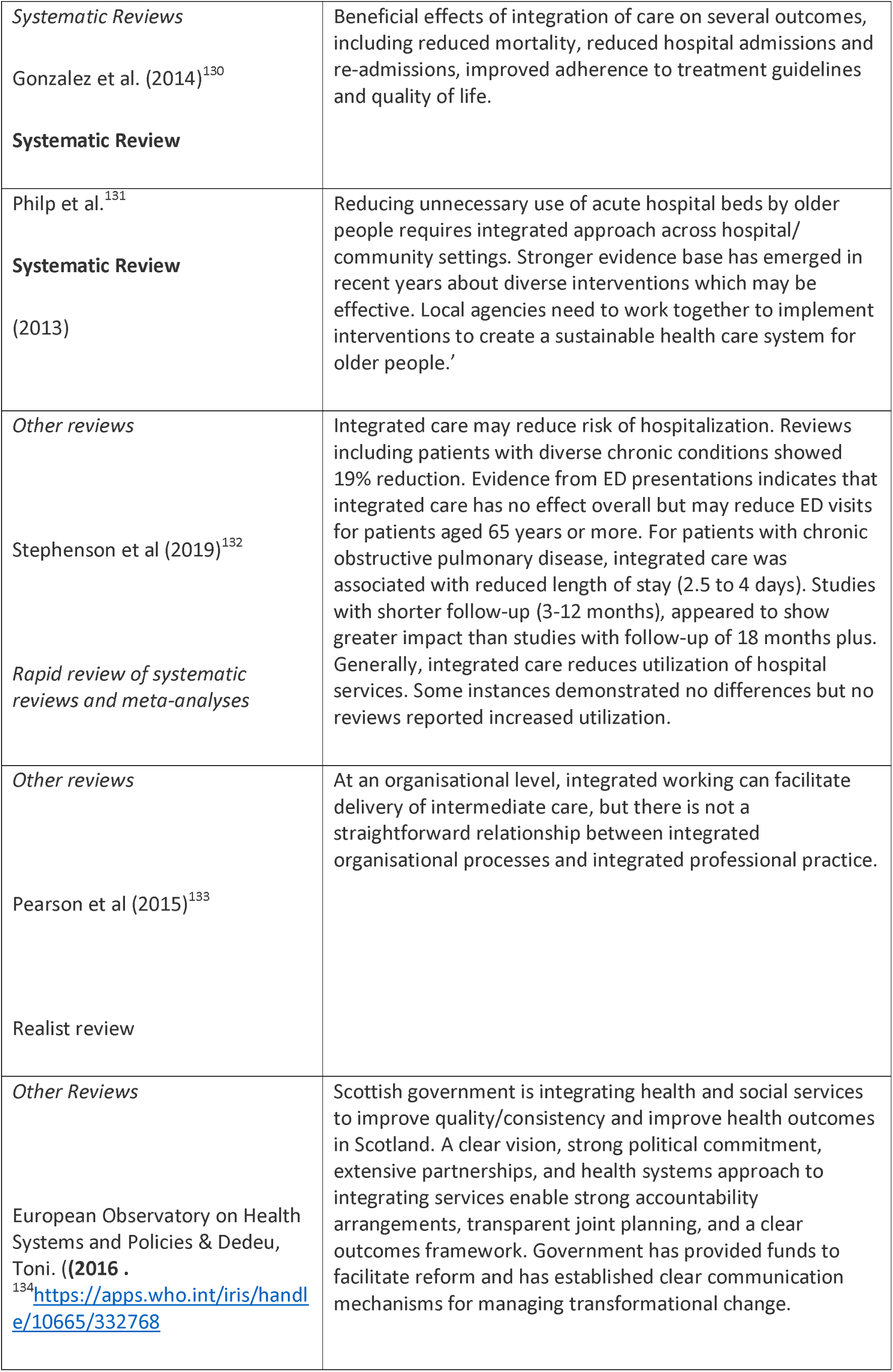
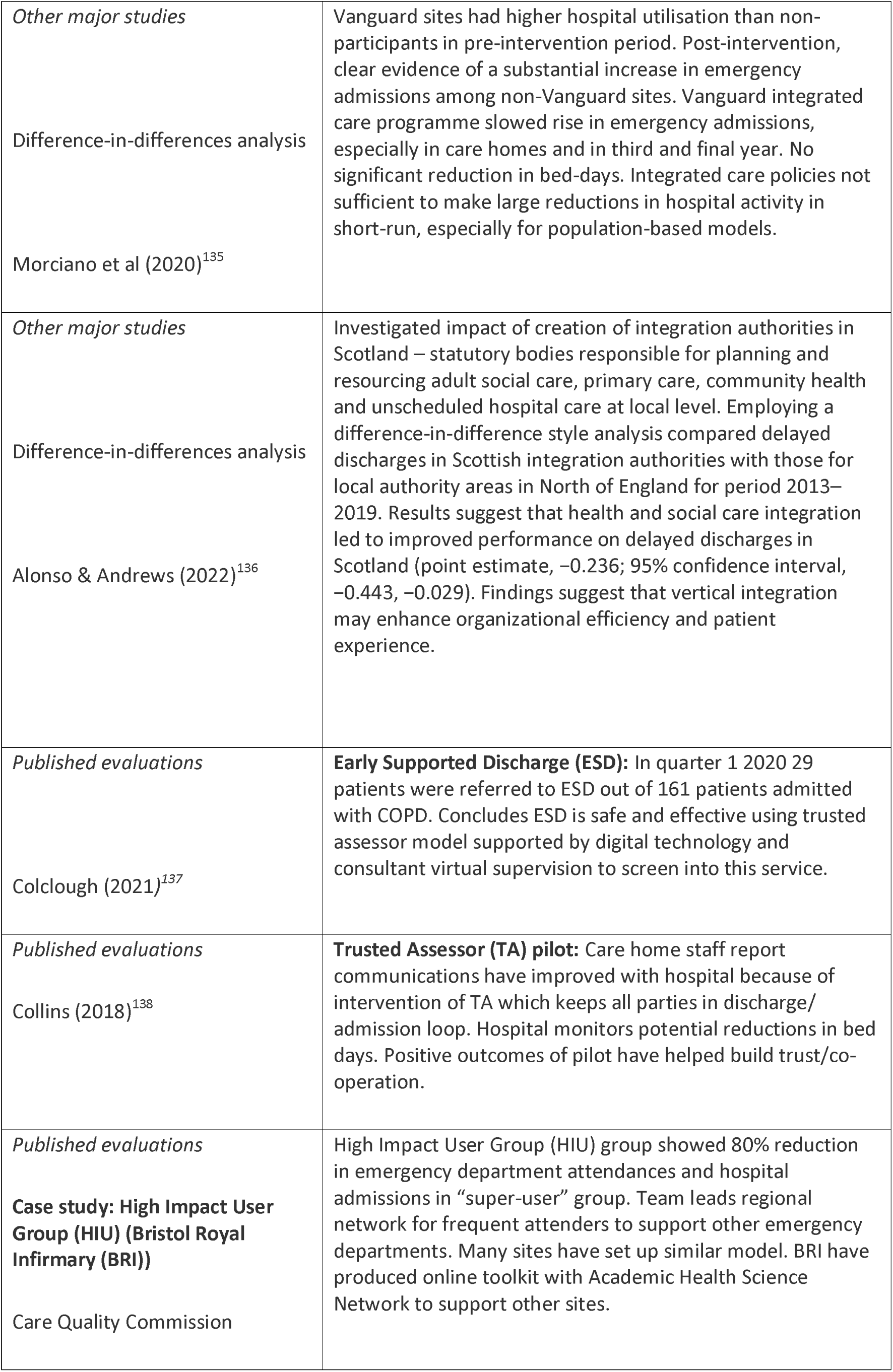
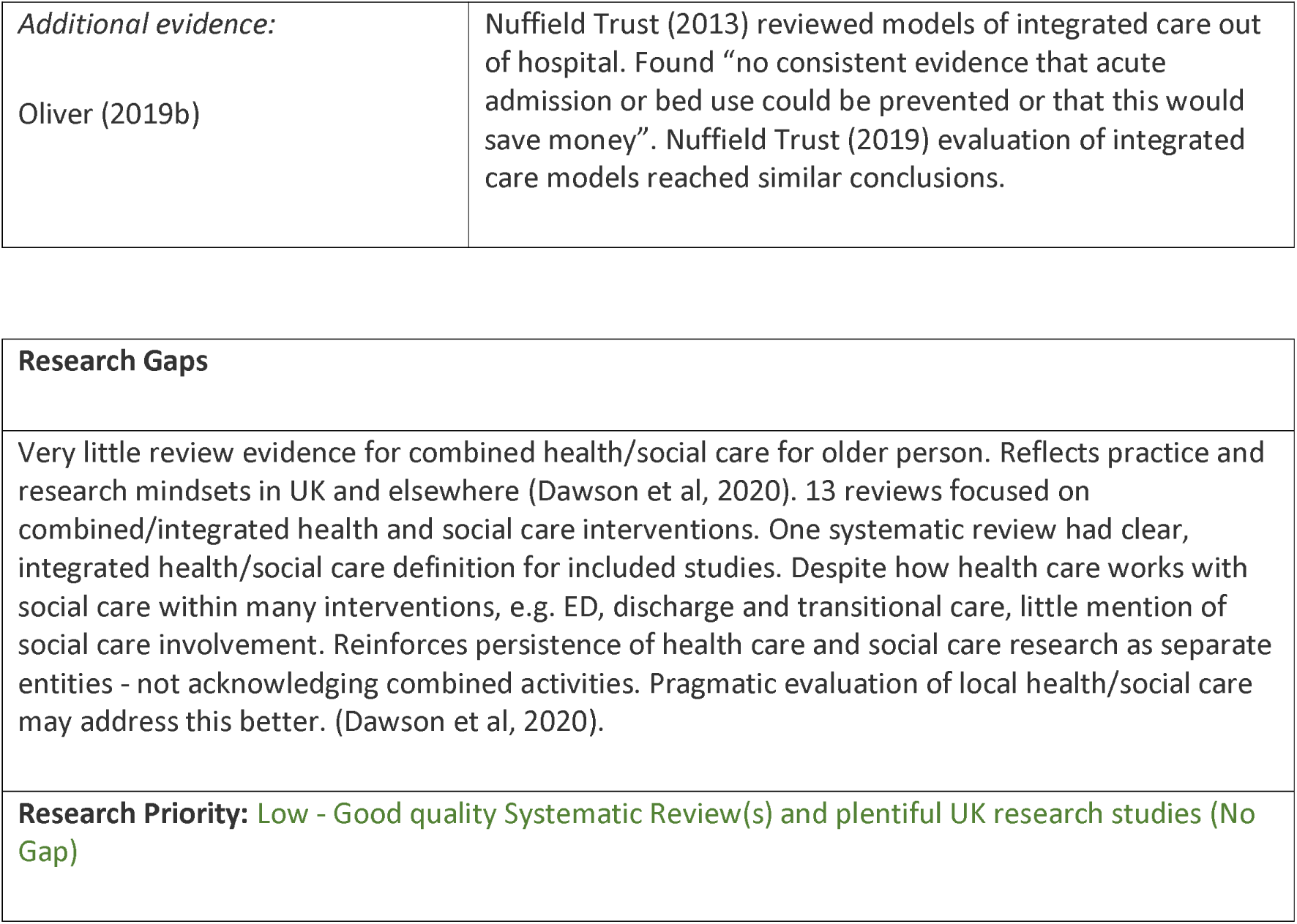
Integrated Care: Interventions and Supporting Evidence.

| Supporting evidence | Results/Findings/Recommendations (from Abstract) |
| --- | --- |
| *Winter pressure specific* | None specific |
| *Systematic reviews*<br>Dawson et al <sup>114</sup><br>(SR of Systematic Reviews) | 71 systematic reviews included: 62 quantitative, seven qualitative and two mixed methods reviews. 52 reviews of healthcare interventions and 19 reviews of social care interventions. Scrutinises existing evidence and evidence gaps of nine broad types of health/social care interventions. Proposes that evidence for older person health/social care interventions can be systematically defined, filtered and organized by meta-reviews. Proposes structured research agenda for summarising evidence of health/ social care interventions, indicating those suitable for co-provision. |
| *Systematic reviews:*<br>Dawson et al (SR of Systematic Reviews) <sup>114</sup> | Combined health and social care poorly described in included systematic reviews. All 13 definitions came from health care systematic reviews with eight describing care in the community interventions and five describing discharge/ transitional interventions. Combined health/social activity implied (as opposed to specific integrated working (with only two exceptions)). Discharge and transitional interventions defined as 'interventions that could be implemented in any health or social care setting (primary, secondary or community care), if they cross boundary between two or more settings.' |
| *Systematic reviews*<br>Farmanova et al (2019) <sup>124</sup> | 15 examples of integrated care organizations in OECD countries from 57 articles/reports. Key redesign strategies included focusing on health/wellness, intersectoral action/partnerships, addressing health in vulnerable groups, and addressing wider determinants of health, including |
| Systematic review | health service improvements. Redesign interventions included addressing clinical/non-clinical issues e.g establishing housing surgeries in primary care, establishing social and provider networks to support patients with complex needs and broadening of scope of services, workforce redesign etc. Potential reduced utilization of care/costs from wider adoption of strategies/ interventions. Integrated care requires service redesign, supported by cohesive strategies to prevent disease, address social determinants and improve health equity. |
| *Systematic reviews*<br>Sadler et al (2019) <sup>128</sup><br>(Systematic review and narrative synthesis) | Poor relationships between service users and health/social care professionals negatively affected integrated care. In two studies, some service users experienced limited involvement in hospital discharge planning and conflicting information from providers. One UK study reports that recipients of integrated health/social care service highlighted lack of perceived interest from their social care provider in establishing genuine rapport. Positive outcomes achieved for some user groups/individuals but not others. Lack of changed outcomes in social care, and service users' low level of satisfaction with social care services appears to be associated with impact of agency work and social work prioritising of maintenance/prevention outcomes. |
| *Systematic reviews*<br>Baxter et al (2018) <sup>129</sup><br>Systematic Review | 167 studies included. Evidence for perceived improved quality of care, increased patient satisfaction, and improved access to care. Evidence rated as inconsistent/limited regarding system-wide impacts on primary care, secondary care, and health care costs. Limited differences between outcomes reported by UK and international studies. Overall literature had limited consideration of effects on service users. Models of integrated care may enhance patient satisfaction, increase perceived quality of care, and enable access to services. Evidence for other outcomes including service costs remains unclear. Indications of improved access may have important implications for services struggling to cope with increasing demand. |
| *Systematic Reviews*<br>Damery et al. (2016)<br>Systematic Review | All outcomes showed significant reductions. Several potentially effective interventions. Interventions rarely demonstrated unequivocally positive effects. Questions remain about whether magnitude of potentially achievable gains from integrated care is enough to satisfy national targets for reductions in hospital activity. |
| *Systematic Reviews*<br>Gonzalez et al. (2014)<sup>130</sup><br>**Systematic Review** | Beneficial effects of integration of care on several outcomes, including reduced mortality, reduced hospital admissions and re-admissions, improved adherence to treatment guidelines and quality of life. |
| Philp et al.<sup>131</sup><br>**Systematic Review**<br>(2013) | Reducing unnecessary use of acute hospital beds by older people requires integrated approach across hospital/ community settings. Stronger evidence base has emerged in recent years about diverse interventions which may be effective. Local agencies need to work together to implement interventions to create a sustainable health care system for older people.' |
| *Other reviews*<br>Stephenson et al (2019)<sup>132</sup><br>*Rapid review of systematic reviews and meta-analyses* | Integrated care may reduce risk of hospitalization. Reviews including patients with diverse chronic conditions showed 19% reduction. Evidence from ED presentations indicates that integrated care has no effect overall but may reduce ED visits for patients aged 65 years or more. For patients with chronic obstructive pulmonary disease, integrated care was associated with reduced length of stay (2.5 to 4 days). Studies with shorter follow-up (3-12 months), appeared to show greater impact than studies with follow-up of 18 months plus. Generally, integrated care reduces utilization of hospital services. Some instances demonstrated no differences but no reviews reported increased utilization. |
| *Other reviews*<br>Pearson et al (2015)<sup>133</sup><br>Realist review | At an organisational level, integrated working can facilitate delivery of intermediate care, but there is not a straightforward relationship between integrated organisational processes and integrated professional practice. |
| *Other Reviews*<br>European Observatory on Health Systems and Policies & Dedeu, Toni. ((2016 .<br><sup>134</sup>https://apps.who.int/iris/handle/10665/332768 | Scottish government is integrating health and social services to improve quality/consistency and improve health outcomes in Scotland. A clear vision, strong political commitment, extensive partnerships, and health systems approach to integrating services enable strong accountability arrangements, transparent joint planning, and a clear outcomes framework. Government has provided funds to facilitate reform and has established clear communication mechanisms for managing transformational change. |
| *Other major studies*<br>Difference-in-differences analysis<br>Morciano et al (2020)<sup>135</sup> | Vanguard sites had higher hospital utilisation than non-participants in pre-intervention period. Post-intervention, clear evidence of a substantial increase in emergency admissions among non-Vanguard sites. Vanguard integrated care programme slowed rise in emergency admissions, especially in care homes and in third and final year. No significant reduction in bed-days. Integrated care policies not sufficient to make large reductions in hospital activity in short-run, especially for population-based models. |
| *Other major studies*<br>Difference-in-differences analysis<br>Alonso & Andrews (2022)<sup>136</sup> | Investigated impact of creation of integration authorities in Scotland – statutory bodies responsible for planning and resourcing adult social care, primary care, community health and unscheduled hospital care at local level. Employing a difference-in-difference style analysis compared delayed discharges in Scottish integration authorities with those for local authority areas in North of England for period 2013–2019. Results suggest that health and social care integration led to improved performance on delayed discharges in Scotland (point estimate, –0.236; 95% confidence interval, –0.443, –0.029). Findings suggest that vertical integration may enhance organizational efficiency and patient experience. |
| *Published evaluations*<br>Colclough (2021)<sup>137</sup> | **Early Supported Discharge (ESD):** In quarter 1 2020 29 patients were referred to ESD out of 161 patients admitted with COPD. Concludes ESD is safe and effective using trusted assessor model supported by digital technology and consultant virtual supervision to screen into this service. |
| *Published evaluations*<br>Collins (2018)<sup>138</sup> | **Trusted Assessor (TA) pilot:** Care home staff report communications have improved with hospital because of intervention of TA which keeps all parties in discharge/admission loop. Hospital monitors potential reductions in bed days. Positive outcomes of pilot have helped build trust/co-operation. |
| *Published evaluations*<br>**Case study: High Impact User Group (HIU) (Bristol Royal Infirmary (BRI))**<br>Care Quality Commission | High Impact User Group (HIU) group showed 80% reduction in emergency department attendances and hospital admissions in “super-user” group. Team leads regional network for frequent attenders to support other emergency departments. Many sites have set up similar model. BRI have produced online toolkit with Academic Health Science Network to support other sites. |
| *Additional evidence:*<br>Oliver (2019b) | Nuffield Trust (2013) reviewed models of integrated care out of hospital. Found “no consistent evidence that acute admission or bed use could be prevented or that this would save money”. Nuffield Trust (2019) evaluation of integrated care models reached similar conclusions. |

|  |
| --- |
| **Research Gaps** |
| Very little review evidence for combined health/social care for older person. Reflects practice and research mindsets in UK and elsewhere (Dawson et al, 2020). 13 reviews focused on combined/integrated health and social care interventions. One systematic review had clear, integrated health/social care definition for included studies. Despite how health care works with social care within many interventions, e.g. ED, discharge and transitional care, little mention of social care involvement. Reinforces persistence of health care and social care research as separate entities - not acknowledging combined activities. Pragmatic evaluation of local health/social care may address this better. (Dawson et al, 2020). |
| **Research Priority:** Low - Good quality Systematic Review(s) and plentiful UK research studies (No Gap) |

#### IC- Winter Improvement Collaborative

**Table 79.**
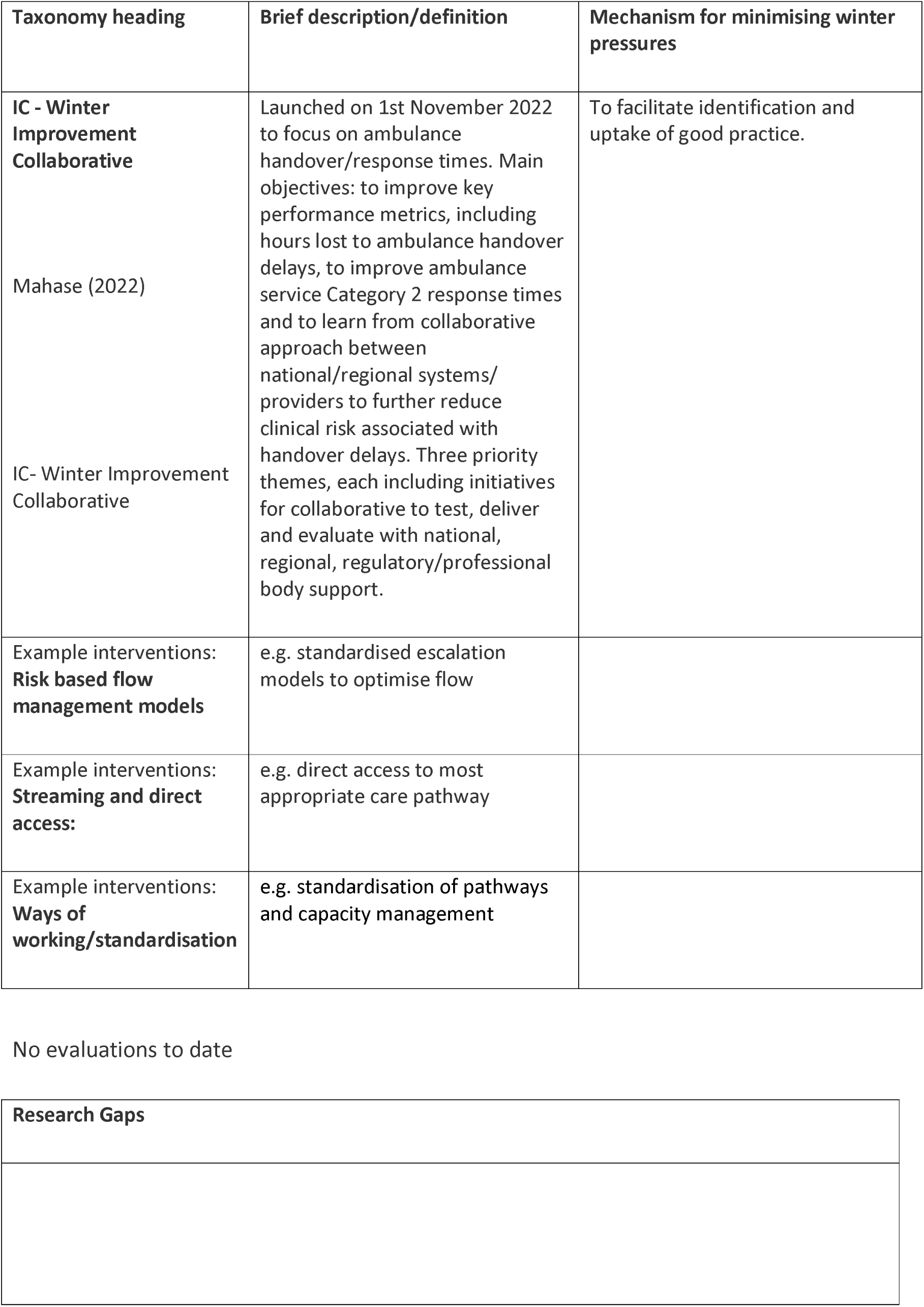
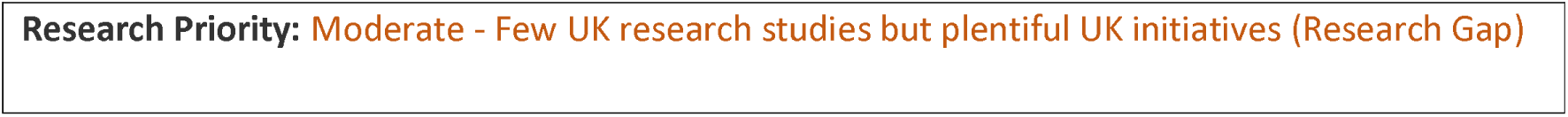
Winter Improvement Collaborative: Definitions and Rationales.

#### IC - Integrated Care Discharge “Huddles”

**Table 80.** IC - Integrated Care Discharge “Huddles”: Definitions and Rationales.

| Taxonomy heading | Brief description/definition | Mechanism for minimising winter pressures |
| --- | --- | --- |
| IC- Discharge Huddle | Hospital wide, multidisciplinary team based, patient discharge meeting often including social care staff and other agencies | To improve flow and discharge planning through multidisciplinary communication and problem solving. |
| Example interventions:<br>**Multidisciplinary Team Discharge "Huddle"**<br>Randles et al (2019)<sup>139</sup> | Introduces hospital-wide multidisciplinary team based huddles (HUDDLE) in a city centre hospital to improve flow and discharge planning. | Rather than negative view of 'bed blockers', allows pro-active approach to management and re-enablement of patients with complex care and discharge needs. Early identification of patients with complex needs allows greater focus on appropriate planning earlier in patient journey. |
| Example interventions:<br>**West Lothian Health and Social Care Partnership Integrated Daily Discharge Flow Huddle**<br>Wilson (2022)<sup>140</sup> | Introduces 'Integrated Discharge Hub' approach to hospital discharge planning where acute care staff work with social care professionals and local carers centre to solve community placement problems for patients awaiting placements. | Seeks to improve patient flow through health and social care system, reduce delayed discharges and improve patient journey. |

**Table 81.**
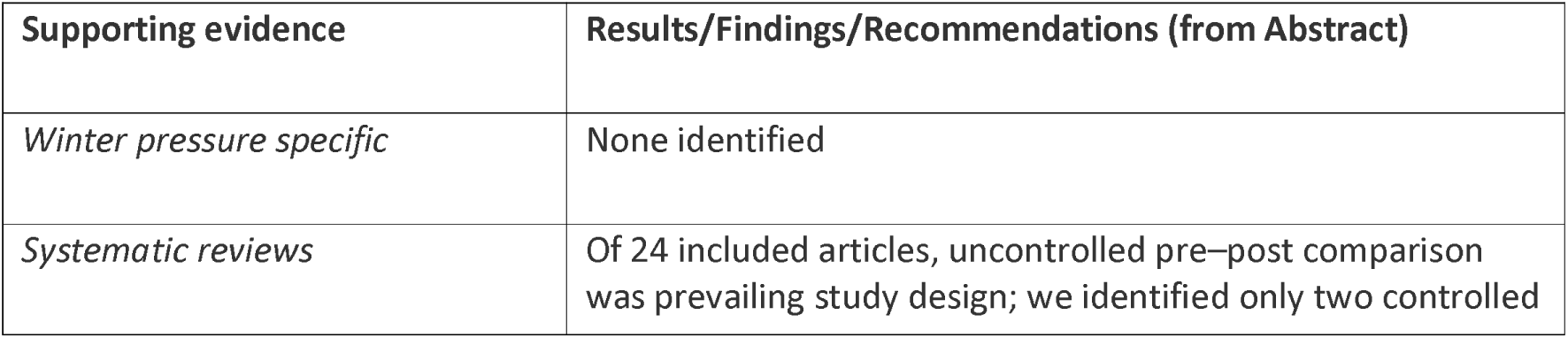
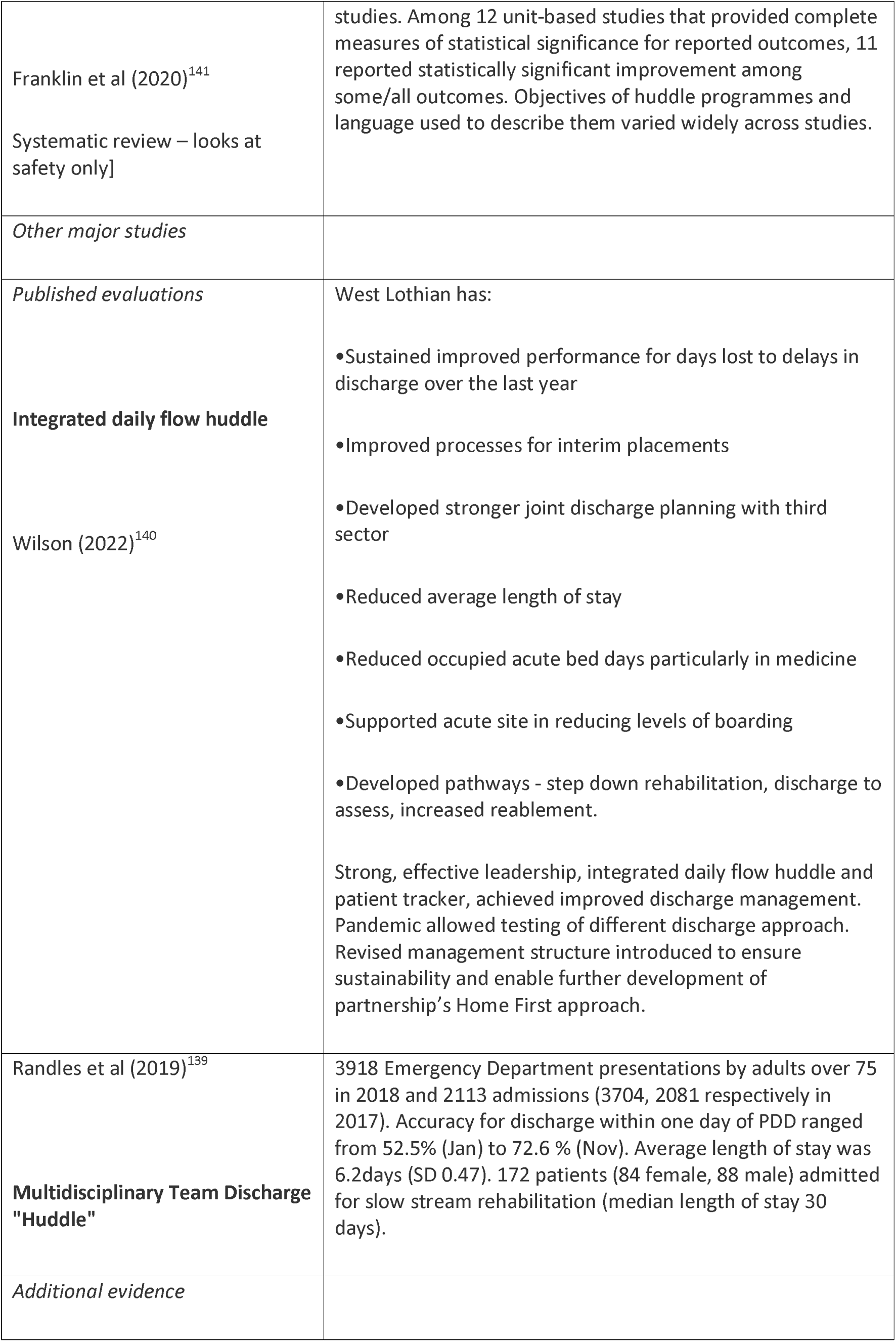
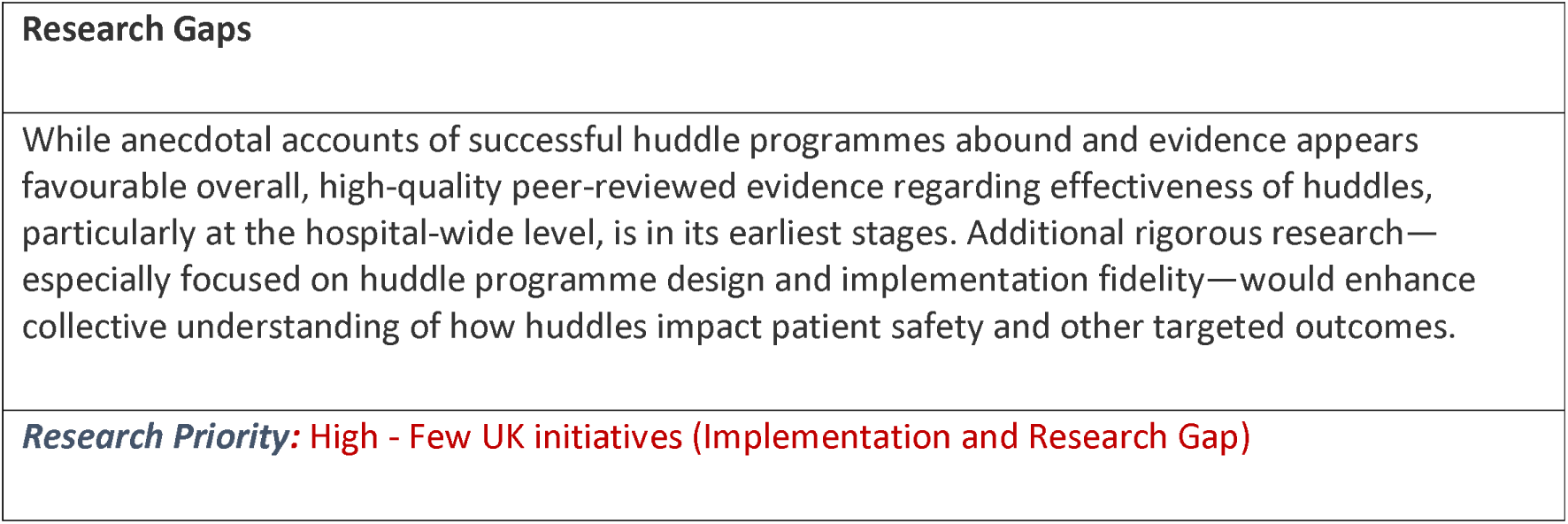
IC - Integrated Care Discharge “Huddles”: Interventions and Supporting Evidence.

| Supporting evidence | Results/Findings/Recommendations (from Abstract) |
| --- | --- |
| *Winter pressure specific* | None identified |
| *Systematic reviews* | Of 24 included articles, uncontrolled pre–post comparison was prevailing study design; we identified only two controlled |
| Franklin et al (2020)<sup>141</sup><br>Systematic review – looks at safety only] | studies. Among 12 unit-based studies that provided complete measures of statistical significance for reported outcomes, 11 reported statistically significant improvement among some/all outcomes. Objectives of huddle programmes and language used to describe them varied widely across studies. |
| *Other major studies* |  |
| *Published evaluations*<br>**Integrated daily flow huddle**<br>Wilson (2022)<sup>140</sup> | West Lothian has:<br>•Sustained improved performance for days lost to delays in discharge over the last year<br>•Improved processes for interim placements<br>•Developed stronger joint discharge planning with third sector<br>•Reduced average length of stay<br>•Reduced occupied acute bed days particularly in medicine<br>•Supported acute site in reducing levels of boarding<br>•Developed pathways - step down rehabilitation, discharge to assess, increased reablement.<br>Strong, effective leadership, integrated daily flow huddle and patient tracker, achieved improved discharge management. Pandemic allowed testing of different discharge approach. Revised management structure introduced to ensure sustainability and enable further development of partnership's Home First approach. |
| Randles et al (2019)<sup>139</sup><br>**Multidisciplinary Team Discharge "Huddle"** | 3918 Emergency Department presentations by adults over 75 in 2018 and 2113 admissions (3704, 2081 respectively in 2017). Accuracy for discharge within one day of PDD ranged from 52.5% (Jan) to 72.6 % (Nov). Average length of stay was 6.2days (SD 0.47). 172 patients (84 female, 88 male) admitted for slow stream rehabilitation (median length of stay 30 days). |
| *Additional evidence* |  |

|  |
| --- |
| **Research Gaps** |
| While anecdotal accounts of successful huddle programmes abound and evidence appears favourable overall, high-quality peer-reviewed evidence regarding effectiveness of huddles, particularly at the hospital-wide level, is in its earliest stages. Additional rigorous research—especially focused on huddle programme design and implementation fidelity—would enhance collective understanding of how huddles impact patient safety and other targeted outcomes. |
| **Research Priority:** High - Few UK initiatives (Implementation and Research Gap) |

Within this taxonomy section, we identified numerous but varied approaches to integration and, as mentioned elsewhere a focus on interventions that are formally labelled as “integrated care” provides a potentially misleading picture of a landscape where integration is present to so many different degrees and at different levels. Indeed the sheer heterogeneity of initiatives makes it challenging to take stock of current activity.

Integrated care organisations constitute one of the development areas for both health and care services and for research and it seems that this research agenda will little supplementary stimulation. However a particular focus on the contribution to hospital avoidance and prevention of delayed discharge would potentially harness the benefits of being able to track pathways throughout integrated care organisations rather than stop at the artificial borders between sectors as may previously have been the case for more siloed research. Further research studies could therefore potentially capitalise on the new models of care to develop a more contemporary and forward-looking research agenda.

### Targeting carers (includes TC - Carer Education; TC - Carer Preparation; TC - Carer Respite and TC - Carer Support)

Findings from the Targeting Carers (TC) initiatives are presented in detail in Tables 82 to 83 below and associated summary paragraphs. Four overall approaches were classified as Targeting Carers; Carer Education; Carer Preparation; Carer Respite and Carer Support. As with other sections of the taxonomy we found considerable overlap both within carer interventions and between other interventions and carer provision.

**Table 82.** Targeting carers: Definitions and Rationales.

| **Taxonomy heading** | **Brief description/definition** | **Mechanism for minimising winter pressures** |
| --- | --- | --- |
| TC - Carer Education | Carer education, typically during periods of low demand when the carer can attend without compromising their care provision, helps a carer to build up their own capacity and resources. Attendance at carer education opportunities may require short-term respite (see below) with the expectation that this will offset longer unmanaged periods of demand that occur at times of high demand. | Carer education increases knowledge and resources of individual carers and their capacity to anticipate and respond to crises. By handling individual demand using informal care resources the carer offers an alternative to unplanned presentation to acute services and/or admissions. |
| TC - Carer Preparation | Carer preparation helps to anticipate certain types of need and demand thereby increasing the carer's capacity to manage such individual demands as and when they arise. | Carer preparation increases capacity of informal care to anticipate and respond to potential individual demands, preventing some crises and handling other demands using informal care resources as an alternative to unplanned presentation to acute services and/or admissions. |
| TC - Carer Respite | Regular demands of caring can lead to physical and emotional exhaustion. Carer respite offers the opportunity for the carer to take a break from their caring responsibility. Carer respite may involve multiple different options e.g. an hour each week, an occasional day, a week or two for a holiday, or a combination of all of these. | Carer respite reduces occurrence of crises leading to unplanned demands upon acute services. Increases capacity of informal care to manage individual demand over an extended period and uses respite resources to handle demands as an alternative to unplanned presentation to acute services and/or admissions. |
| TC - Carer | Carers often share similar needs and can act as self help resource to one | Carer support increases capacity of carer and thus of informal care to |
| Support | another. Providing opportunities to meet and support one another increases individual and collective carer resources and the capacity to manage demands. Attendance at carer support opportunities may require short-term respite (see above) | manage individual demand thereby providing an alternative to unplanned presentation to acute services and/or admissions. |

**Table 83.**
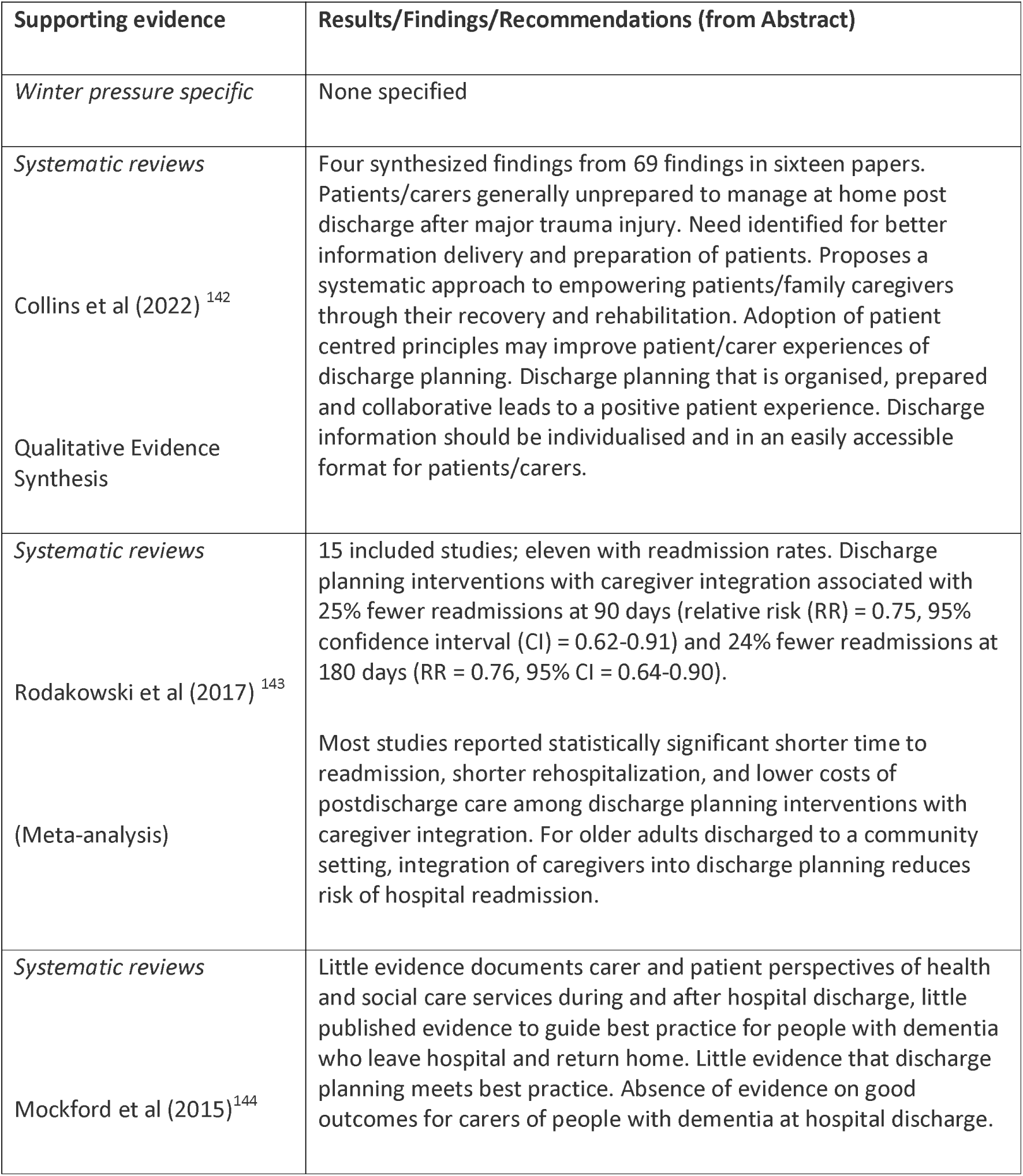
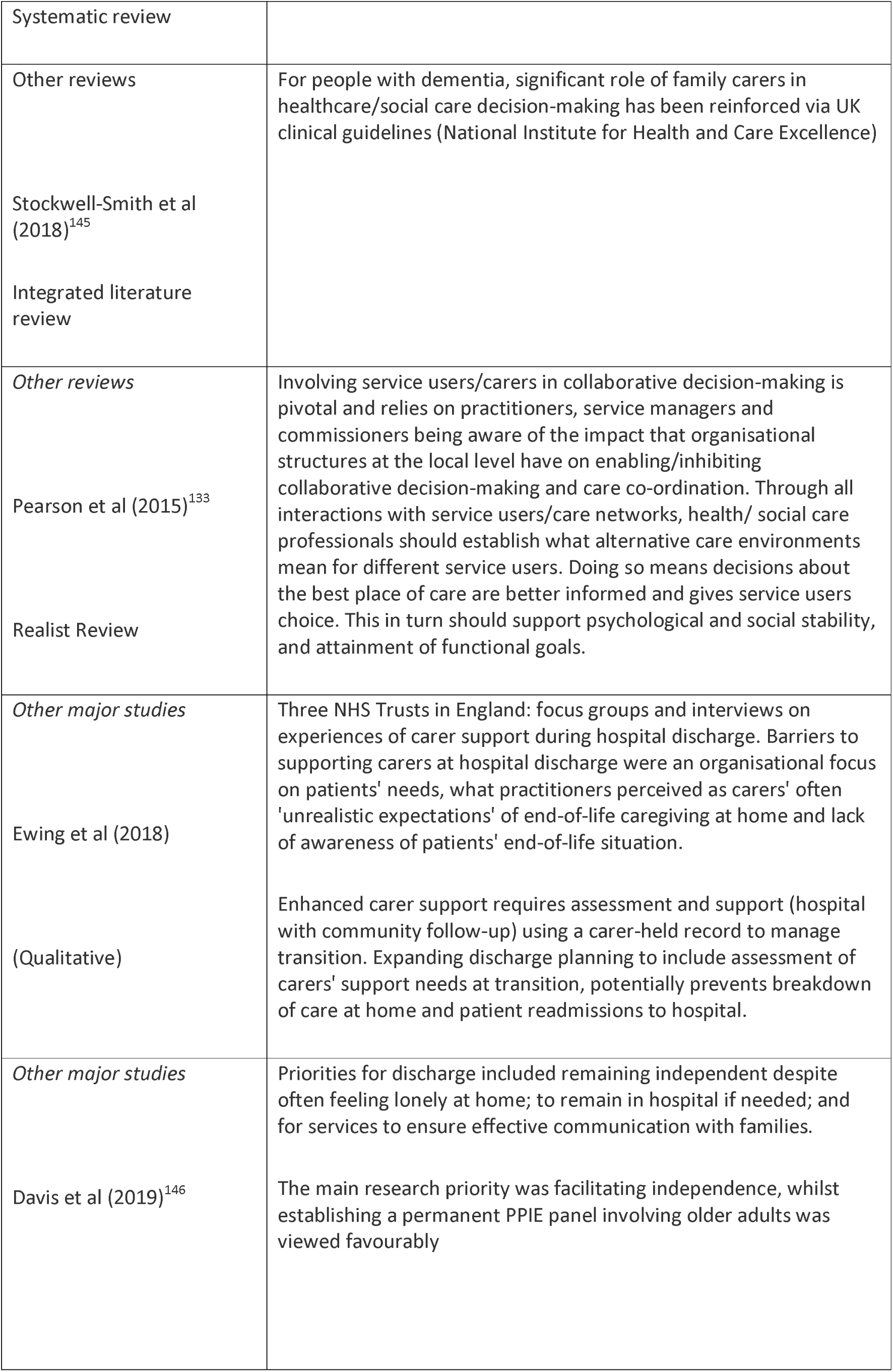
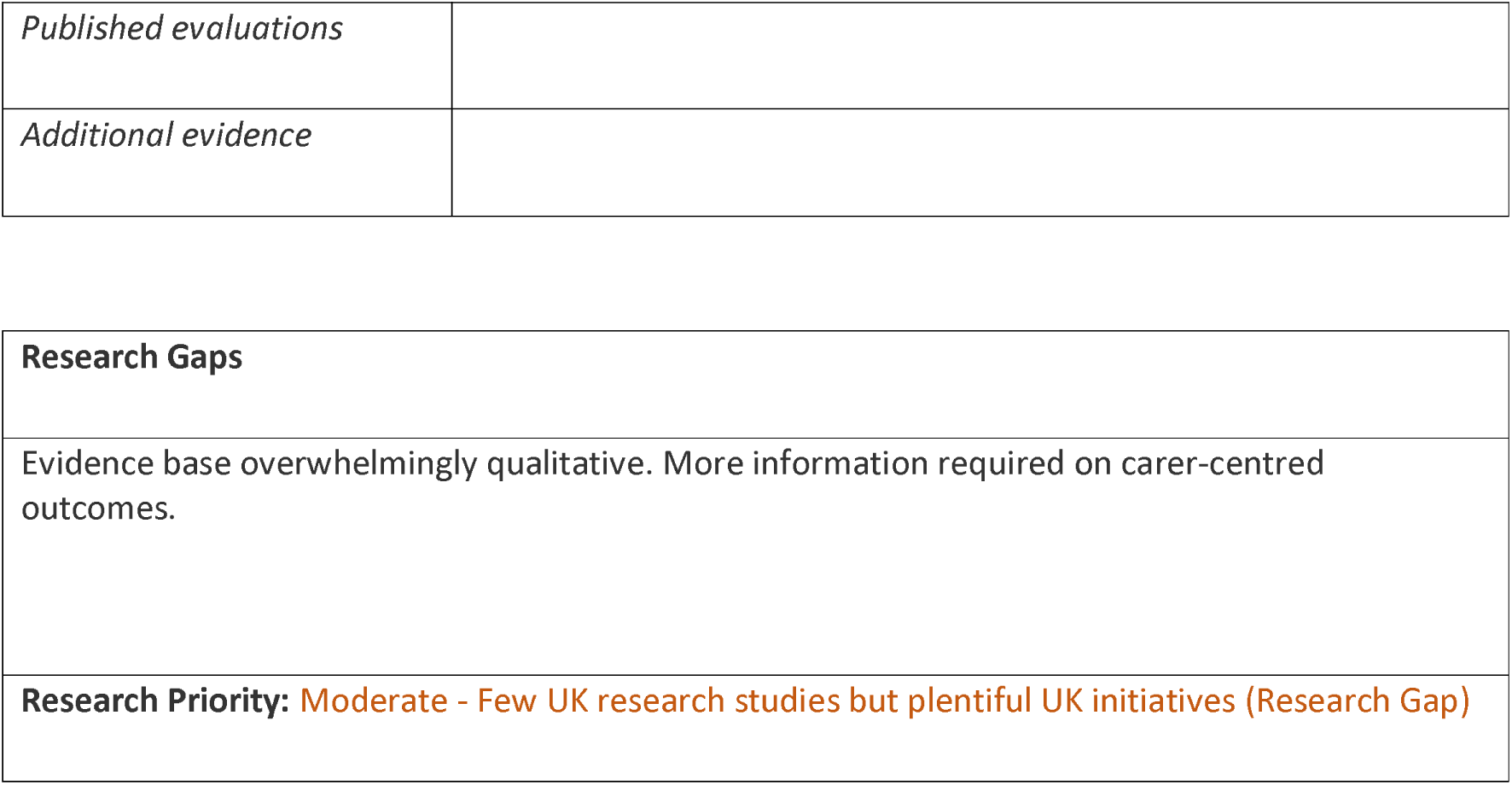
Targeting carers: Interventions and Supporting Evidence.

| Supporting evidence | Results/Findings/Recommendations (from Abstract) |
| --- | --- |
| *Winter pressure specific* | None specified |
| *Systematic reviews*<br>Collins et al (2022) <sup>142</sup><br>Qualitative Evidence Synthesis | Four synthesized findings from 69 findings in sixteen papers. Patients/carers generally unprepared to manage at home post discharge after major trauma injury. Need identified for better information delivery and preparation of patients. Proposes a systematic approach to empowering patients/family caregivers through their recovery and rehabilitation. Adoption of patient centred principles may improve patient/carer experiences of discharge planning. Discharge planning that is organised, prepared and collaborative leads to a positive patient experience. Discharge information should be individualised and in an easily accessible format for patients/carers. |
| *Systematic reviews*<br>Rodakowski et al (2017) <sup>143</sup><br>(Meta-analysis) | 15 included studies; eleven with readmission rates. Discharge planning interventions with caregiver integration associated with 25% fewer readmissions at 90 days (relative risk (RR) = 0.75, 95% confidence interval (CI) = 0.62-0.91) and 24% fewer readmissions at 180 days (RR = 0.76, 95% CI = 0.64-0.90).<br>Most studies reported statistically significant shorter time to readmission, shorter rehospitalization, and lower costs of postdischarge care among discharge planning interventions with caregiver integration. For older adults discharged to a community setting, integration of caregivers into discharge planning reduces risk of hospital readmission. |
| *Systematic reviews*<br>Mockford et al (2015) <sup>144</sup> | Little evidence documents carer and patient perspectives of health and social care services during and after hospital discharge, little published evidence to guide best practice for people with dementia who leave hospital and return home. Little evidence that discharge planning meets best practice. Absence of evidence on good outcomes for carers of people with dementia at hospital discharge. |
| Systematic review |  |
| Other reviews<br>Stockwell-Smith et al (2018)<sup>145</sup><br>Integrated literature review | For people with dementia, significant role of family carers in healthcare/social care decision-making has been reinforced via UK clinical guidelines (National Institute for Health and Care Excellence) |
| Other reviews<br>Pearson et al (2015)<sup>133</sup><br>Realist Review | Involving service users/carers in collaborative decision-making is pivotal and relies on practitioners, service managers and commissioners being aware of the impact that organisational structures at the local level have on enabling/inhibiting collaborative decision-making and care co-ordination. Through all interactions with service users/care networks, health/ social care professionals should establish what alternative care environments mean for different service users. Doing so means decisions about the best place of care are better informed and gives service users choice. This in turn should support psychological and social stability, and attainment of functional goals. |
| Other major studies<br>Ewing et al (2018)<br>(Qualitative) | Three NHS Trusts in England: focus groups and interviews on experiences of carer support during hospital discharge. Barriers to supporting carers at hospital discharge were an organisational focus on patients' needs, what practitioners perceived as carers' often 'unrealistic expectations' of end-of-life caregiving at home and lack of awareness of patients' end-of-life situation.<br>Enhanced carer support requires assessment and support (hospital with community follow-up) using a carer-held record to manage transition. Expanding discharge planning to include assessment of carers' support needs at transition, potentially prevents breakdown of care at home and patient readmissions to hospital. |
| Other major studies<br>Davis et al (2019)<sup>146</sup> | Priorities for discharge included remaining independent despite often feeling lonely at home; to remain in hospital if needed; and for services to ensure effective communication with families.<br>The main research priority was facilitating independence, whilst establishing a permanent PPIE panel involving older adults was viewed favourably |
| *Published evaluations* |  |
| *Additional evidence* |  |

|  |
| --- |
| **Research Gaps** |
| Evidence base overwhelmingly qualitative. More information required on carer-centred outcomes. |
| **Research Priority:** Moderate - Few UK research studies but plentiful UK initiatives (Research Gap) |

The sheer breadth and scope of this taxonomy section made a comprehensive survey of research prohibitive. The heading is included in the taxonomy for completeness, as mentioned in the discussion targeting carers sustains its own research agenda. Nevertheless we have identified the four key areas required within targeting carers and have identified limited initiatives or research studies. The role of carers has not only been identified as pivotal but it also represents a hidden cost alongside formal systems of care, often compensating for pressures within the overall health system. However, reliance on carers exerts its own impact on the carers themselves. “Winter pressures and the implications on carers” would seem to constitute a promising area for further exploration. It remains to be seen whether benefits of, for example, carer respite during less-pressured times can produce a dividend during subsequent pressure points in terms of carer resilience and community support. Because of its often invisible nature carer contribution can prove very challenging to measure.

## Discussion

### Overview of Discussion

The Discussion starts with an overall summary of common findings over all intervention types. It then proceeds to brief summaries of the five overarching types of interventions; (i) structural interventions, (ii) changing staff behaviour, (iii) changing community provision, (iv) integrated care and (v) targeting carers. Finally, the Discussion summarises the research priorities and the potential for a realist review.

### Overall summary

Undoubtedly the foremost limitation of this review is the absolute shortage of studies that describe and evaluate winter pressures in a specific way. As mentioned above there is no shortage of studies that describe achievements against a backdrop of winter pressures or that describe quality improvements with multiple collateral benefits including an impact against winter demands. However, as a review team we agreed that this is substantively different from interventions that specifically target winter pressures. Even following winter funding to identify specific targets for development could not guarantee that each intervention was conceived against the specific requirements particular to winter pressures as opposed to relieving capacity pressures more broadly. As a team we found ourselves asking should “winter” simply privilege the time when initiatives take place, conventionally “winter pressures” have been attributed to a specific time period or even a specific date!, or are we looking for something related to specific pressures on the system as characterised by the pressures observed at winter. This discussion is particularly relevant given recent acknowledgement that in recent years winter pressures seem to be taking place “all the year round”. This realisation had a considerable bearing on our decision, following the preliminary mapping specific to winter pressures, to expand to the evidence base on wider responses to health and social care system pressures.

Overall, the evidence base is characterised by large numbers of case studies, often published online or presented at conferences, and relatively few peer-reviewed journal articles. Case studies are often accompanied by guidance to support implementation of changes to services. This distribution of evidence probably reflects the urgent need to develop and implement solutions to the ever increasing winter (and increasingly year-round) pressures on the health and care system. The majority of evaluations report positive effects on important outcomes such as length of hospital stay but many are uncontrolled or based on small samples, meaning that they need to be interpreted with caution.

A further interpretation of the evidence base, fuelled by the expansion of winter pressures to other times of the year, acknowledges that “winter pressures”, although undoubtedly real, tend to assume extra narrative significance by presenting a continuous systemic challenge in binary form. Politically “winter pressures” may become a device used to spotlight specific attention on health system under-resourcing so that these issues assume special significance over and above general awareness as highlighted by politicians, the popular media and public campaigns.

### Summary – Structural Interventions

Structural responses to winter pressures were classified as ‘hospital avoidance’, ‘alternative delivery site’, ‘facilitated discharge’ and ‘cross-cutting’. In line with the review’s emphasis on timely hospital discharge and integrated care post-discharge, most taxonomy headings fell into the latter two categories. Nevertheless, it is clear that avoiding admission in the first place (where clinically appropriate and feasible) and exploring care delivery in different settings both play important roles in the overall response.

In the area of hospital avoidance, we identified many UK initiatives around same day emergency care, mostly reported as case studies or evaluations of low methodological quality, suggesting a research gap. Surgical hubs are a more recent development and we have identified research and implementation gaps in this area. Extra service delivery takes a variety of forms and some successes have been reported in both emergency and elective care but the evidence base remains limited.

In terms of alternative delivery sites, acute medical units were defined by the Royal College of Physicians, London as long ago as 2007. Recent systematic reviews have demonstrated their effectiveness. Other initiatives have involved developing specialist units using winter funding but these remain to be fully evaluated.

Some aspects of facilitated discharge are relatively well researched, particularly models based on ‘discharge to assess’ (D2A, also ‘Home First’ and others). In addition to case studies and guidance, we identified a range of Healthwatch reports, providing a patient/carer perspective on how D2A services operate in real-life clinical practice. Interventions with little UK evidence were sometimes supported by systematic reviews, although the quality of these varied. Some taxonomy headings, e.g. ‘bed management’ and ‘discharge co-ordinators’, were often evaluated as part of a broader process of ‘discharge planning’. ‘Patient flow’ is another broad heading with some overlap with both ‘bed management’ and discharge planning. The concept of ‘patient flow’ is also broader than facilitated discharge, although its ultimate goal is ensuring safe discharge as soon as is clinically appropriate. This highlights an issue we found with the taxonomy of the overlap within the taxonomy headings.

Evidence support for initiatives defined as ‘cross-cutting’ varied widely. The winter pressure search identified UK initiatives to support improved teamwork, together with associated guidance (Table 19). Various initiatives have applied data sharing and digital technology, but we found a relative lack of research studies. Actions at the organisational level, e.g. involving changes in governance or formulation and implementation of policy, play an important part in responding to winter pressures but this was not reflected in research. Further research studies are needed in the area of seven day services although promising case studies exist. Staff redeployment has achieved successes within the pharmacy profession and research involving other staff groups would be useful. Volunteers have been used within the NHS in many different areas but there needs to be further evaluation of these initiatives in relation to winter pressures.

We identified significant numbers of recent systematic reviews relevant to, but not targeted at, responses to winter pressures. These are potentially valuable but the review questions rarely overlap completely with the needs of addressing winter pressures, suggesting that further targeted evidence synthesis (perhaps using realist approaches to focus on the UK context) could be valuable. The diverse range of literature identified by this mapping review suggest that such an approach would be feasible for at least some approaches, e.g. ‘discharge to assess’, reablement, virtual wards and optimising patient flow.

### Summary - Changing Staff Behaviour

Changing staff behaviour responses to winter pressures were all classified as ‘cross-cutting’.

There was recent guidance available for documenting local clinical audit from the Healthcare Quality Improvement Partnership. A case study from SaTH used clinical audit to investigate compliance with the discharge process and found that in 2019 patients were still been kept waiting for the same reasons in the discharge lounge and few patients were transferred to the discharge lounge.

Within the Education of Staff heading there was one small research study about teaching staff about patient flow which found that multimodal teaching approaches were the most successful at the ward level to make it relevant to staff. Patient flow is one of the heading within structural interventions and the interventions introduced also could include education staff which again highlights the overlap within the taxonomy headings. Education staff could be part of larger initiatives with multiple components but might not be the outcome that is been measured, more likely that the number of patients discharged or patient length of stay would be main outcomes measured. The importance of looking after workforce and creating the best possible conditions for staff to do their best for patients was highlighted in a recent poster of a rapid review.

There was no evidence for the use of protocols/guidelines specifically to mitigate winter pressures. A systematic review from 2015 researching how to improve the quality of discharge summaries found that guidelines were useful for providing the key components of discharge summaries.

The lack of evidence for the use of quality improvement programmes to mitigate winter pressures could be due to the nature of winter pressures with a need for implementation of rapid solutions without the time for thorough evaluation. However, the NHS Confederation has drawn six lessons from the pandemic experiences of the Q community that would be useful to apply as winter pressures increase. Quality improvement programmes could be useful for introducing initiatives to mitigate winter pressures and would allow for more thorough planning and evaluation.

There was no evidence of the use of quality management systems to mitigate winter pressures. Within Scotland there is a move from the use of quality improvement to the use of quality management to support better quality health and social care indicating could become more important.

Changing staff behaviour can take time and initiatives to minimise winter pressures may need to be implemented rapidly without the time for thorough evaluation or to measure all potential outcomes. Initiatives could potentially be too short to make changes to staff behaviour and they might not measure staff behaviour as an outcome. Further research studies could plan evaluations that consider a variety of outcomes including changes in staff behaviour, although it can be difficult to measure.

### Summary – Changing Community Provision

Numerous approaches were classified as Changing Community Provision. Basically community responses either seek to keep those who need not be admitted to a hospital within the community, whether through preventive or early warning measures or through substituting community for acute services as the point of delivery, or they seek to facilitate and accelerate hospital discharge. In the latter case, two overall approaches can be observed; either by co-ordinating the patient’s needs to remove obstacles to discharge or, again, by offering services traditionally delivered within acute care from a community setting. ‘Hospital avoidance’ from the community has been artificially separated from “hospital avoidance” within the Structural category simply on the basis of the setting from which avoidance is enacted. We considered that attributing initiatives and evidence to the setting is more helpful than aggregating all hospital avoidance schemes together. So, for example, a scheme that seeks to avoid hospital admission by meeting patient needs as an outpatient within acute services is distinguished from a scheme that seeks to avoid placing any demand on acute services.

Findings are presented in detail in Tables 45 to 76 above and associated summary paragraphs. As with Structural interventions (above) we found considerable overlap between definitions and interventions. In some cases we found ourselves placing artificial demarcations between interventions e.g. in focusing telecare on “non-health” care needs and in distinguishing “virtual wards” where a single set of related functions (e.g. rehabilitation) take place from a “virtual hospital” which seeks to replicate the full range of functions offered by a physical hospital Equally common are packages that can be coded for multiple components.

As with the King’s Fund report Reimagining community services: Making the most of our assets (2018) we have found that many areas are trying to improve co-ordination through introducing single points of access or trusted assessor arrangements. The intention is to reduce duplication in referral and assessment processes. Sometimes these changes are associated with creation of new roles with a specific care colZIordination function (care navigators or care co-ordinators). The King’s Fund report highlights how these roles are intended to act as a “workaround” to help people navigate complex and uncolZIordinated services and highlights how “a better solution would be to make services less fragmented, so that this additional layer of complexity is not needed”.

Similarly, we identified commonality in interventions that seek to better co-ordinate care by bringing professionals together in integrated community teams. We discuss team-based approaches in the community under “community teams” while whole system approaches are explored under a separate Integrated Care section. These multidisciplinary community teams typically assemble diverse of community health and social care professionals working alongside groups of GPs. Teams often target older people or other groups with relatively high health and care needs (Naylor et al 2017). Teams are usually locality-based and include multiple professionals. These may include district nurses, community matrons, social workers, mental health professionals, therapists, GPs and voluntary sector workers. The core teams interact with a wide network of local services to meet the full range of people’s needs: “By working together in one team, staff across different disciplines can communicate regularly, share knowledge and expertise and co-ordinate care planning and delivery”. ^109^.

Some aspects of changing community provision are comparatively well-researched, particularly hospital at home services for a variety of health care needs where several systematic reviews exist. Other less-researched initiatives, particularly trusted assessors, step down facilities and single point of access for social care are well represented by UK initiatives. Indeed so many local constituencies are experimenting with such models that it is questionable whether it is meaningful to single specific initiatives for particular attention. Clearly, it could be valuable to explore variations across different models of the same initiative and to undertake robust evaluations.

Almost all of the initiatives are not specifically “winter pressures” approaches. However we have identified their use as winter pressure measures or as recipients for winter funding. This fact has been used to identify the candidacy of initiatives in this report for which we have subsequently sought more substantive and plentiful evidence. A key evaluation issue relates to whether to undertake evaluation specifically of initiatives within a winter pressures context or whether to evaluate them more widely on the understanding that they may be particularly beneficial when the health system is under particular strain as is exemplified by winter pressures.

As with Structural interventions, the evidence base is characterised by large numbers of case studies, often published online or presented at conferences, and relatively few peer-reviewed journal articles. The imperative to tackle urgent issues and to adopt initiatives that have proved promising elsewhere can be detected across the evidence base. We have identified a need for extensive and robust evaluation which could then be targeted for consolidated learning through reviews of case studies or even systematic reviews. Again there is methodological cause for concern in the likely presence of multiple biases such as early adopter bias, hero innovator bias and reporting bias. A further possibility relates to the fact that organisations that are encountering particularly severe problems may adopt multiple responses meaning that the strength of the effect is not replicated across more uniform sites or that positive impact may be achieved by a “kitchen sink effect” of multiple indistinguishable interventions that are introduced simultaneously.

As with the discharge to assess literature within Structural Interventions, the hospital at home literature is associated with significant numbers of recent systematic reviews. Their value is again dissipated by the lack of a specific focus on addressing winter pressures, suggesting that further targeted evidence synthesis (perhaps using realist approaches to focus on the UK context) could be valuable. Certainly the plethora of different initiatives with diverse labels suggests that a focus on common mechanisms would prove more useful than a diffuse investigation based on interventions.

### Summary - Integrated Care

Approaches to Integrated Care operate at multiple organisational levels. Evidence from Scotland allows exploration of vertical integration between health and social care services and its impact on delayed discharges. Early experience from the Vanguards also allows comparison of system-wide integration. Less transformative experiences of integration are seen when health and social care agencies seek to deliver approaches to discharge planning in a joint approach that works across the respective organisational boundaries. Finally, front-line instances of integrated care can be achieved when health and social care agencies are represented in joint operations. This final approach is exemplified by, for example, social care and voluntary agencies being represented at a local level in discharge huddles. Other approaches involve placing staff within a different organisational environment; for example by placing social workers within a hospital or by providing community access to acute hospital expertise. These types of initiative have tended to be discussed under the delivery site for the initiative, whether an acute hospital (covered under structural) or community provision.

Integrated care poses challenges in both achieving and measuring an impact. The complexity of interventions that work across organisational boundaries means that it can take some time to organise new pathways of lasting permanence. Alternatively, short term initiatives may be able to achieve short wins but this impact is either not sustained or, more typically, is not reported over longer time periods making evaluation problematic. Generally, evaluation of organisational approaches to improved discharge planning is neither as wide-ranging nor as sustained as it needs to be to provide reliable evidence. Measurement of long-term outcomes is therefore required.

Positive reporting bias is particularly likely to occur within new initiatives for integrated care. Where multiple measures are reported accounts focus on those indicators that have achieved a positive outcome. Where initiatives are able to report less demonstrable achievements they may focus on qualitative aspects of the integration, in terms of both structure and outcome. In particular, reports of integrated care initiatives focus little attention on the hidden costs in terms of staff time in setting up initiatives and in managing organisational change.

Notwithstanding such caveats the pursuit of integrated care is supported by several studies that have demonstrated positive results and by a wealth of anecdotal experience of the breakdown of health and social care communications between separate agencies. Simplifying communications and procedures carries an implicit driver towards improvement, whether or not achieved short term or sustained over a longer time period.

The specific challenge in relation to integrated care once more lies in the difficulty in attributing the impact of integrated care interventions and initiatives specifically to benefits to acute care utilisation during times of winter pressure. By implication care has to be integrated as a permanent feature across all seasons and patterns of demand. Commentators have observed that the health and social care systems now face “winter pressures” all the year around making integrated care interventions more apposite as a response to endemic health service challenges. The other implication relates to the scale of change often required for change to take place; particularly involving multiple stakeholders and organisations. While any attempts at improving information flow and communication and operation of shared processes are to be welcomed the capacity of organisations to be able to invest time and effort in organisational change at a time of systemic overload has to be questioned. A form of “natural selection” may therefore take place whereby the more easily achieved small-scale adaptation of existing systems may be privileged over more radical system-wide transformation.

A more detailed analytical review could analyse initiatives against a tightly defined and recognised classification of integration. More significantly a realist synthesis could focus on shared mechanisms by which different levels of integration achieve improvements in outcomes relating to admission avoidance or reduction in delayed discharge.

### Summary – Targeting Carers

Approaches to Targeting Carers tend to fall within two main types of response; reducing either demand per se or inappropriate demand by improving carer navigation across the community/acute margins or increasing carer resource and resilience by equipping them to handle demands that might otherwise draw upon hospital-based resources. One of the challenges of the latter approach lies in the fact that carers themselves have their own health and care needs. Placing extra demands on those who are already finding it challenging to cope may represent a short-term solution that reaps later negative dividends e.g. in the carer requiring extra support at another time. At best this may simply spread demands upon the health and social care system more evenly across the year – for example when the carer requires respite services at other times of the year. Whether such a “transferred need” presents acutely or is managed and planned exerts a considerable difference in the success of such measures. Of more concern is when placing an extra burden on a carer results in an immediate breakdown of informal support, meaning that both patient and their carer make demands on the system at a time of winter crisis and perhaps for an extended period in contrast to an otherwise brief episode.

Carer Education and Carer Preparation are generally targeted at the carer in advance of any challenging situation. Education may involve knowledge of what to do when faced with a particular crisis, thereby increasing carer resilience and reducing demand upon acute services or it may involve increasing carer capacity to navigate the care system appropriately, leading to better targeting of need. Preparation may involve practical skills or competencies that reduce demand on formal carers for example in being able to change dressings, administer medication etcetera. In both cases education and preparation typically take place at a time of “normal” demand on the health and care system and needs can be planned and anticipated.

Carer respite may occur in response to an acute need or crisis or may form part of a planned or managed response. Respite can take place on a very short term basis e.g. allowing carers an hour or two away from the demands of their informal care or it can take place over an extended period, as with accommodating an annual one-week or two-week holiday. Again, the assumption behind such interventions is that a planned and managed period of time will not only place less demand on the system than an acute crisis but that its overall duration will typically be shorter than otherwise accumulated. Carer support may either be episodic or ongoing. Carers may offer mutual and reciprocal support developing their own resources to relieve demands on formal care or carer support may be provided by health professionals, with the assumption that these may have more capacity, for example if located within the community, than is available within the acute care system.

The challenge in relation to targeting carers lies in the difficulty in attributing the impact of interventions specifically to benefits to acute care during times of winter pressure. The value of informal care in general to the health and social care system is well recognised and has been quantified. However, this contribution is typically conceived as relating to the whole year around not specifically to times of system pressure. Carer interventions can be attributed to winter pressures where specific winter pressure funding is targeted at carers as beneficiaries. The challenge for evaluation may lie in the reverse direction with benefits from specific-winter pressure interventions for carers delivering benefits that persist beyond the critical winter pressure period. The artificial separation of budgets, both across purposes and across organisations makes attribution of benefit to carer interventions particularly challenging.

Targeting Carers has its own separate research agenda aside from winter pressure responses. Almost all of the initiatives are not specifically “winter pressures” approaches. However, we have identified their use as winter pressure measures or as recipients for winter funding. This fact has been used to identify the candidacy of initiatives in this report for which we have subsequently sought more substantive and plentiful evidence. As for other initiatives, a key evaluation issue relates to whether to undertake evaluation specifically of carer initiatives within a winter pressures context or whether to evaluate them more widely on the understanding that they may be particularly beneficial when the health system is under particular strain as exemplified by winter pressures.

### Research Priorities

#### An overarching research agenda

As suspected from the outset we identified few initiatives specifically identified as a response to winter pressures. Conversely many initiatives tackling admission avoidance or reduction of length of stay and delayed discharge attributed the response to numerous factors which included “winter pressures” alongside multiple contextual factors including an increasingly ageing population and increased demands on health services. The lack of specificity of these responses suggests that an exploration of underpinning mechanisms and how these might contribute to tackling winter pressures, and indeed, other peaks of demand, as with a realist synthesis may be particularly useful. Methodologically, one of the most effective ways of identifying interventions or initiatives in response to winter pressures was in “following the money” i.e. looking at reports of how health organisations allocated funds directly attributed as “winter funding”. We then worked from a list of cursorily identified initiatives and interventions to match to the underpinning evidence base of systematic reviews and major studies. Even here, though, it was challenging to separate interventions targeted at winter pressures from more opportunistic approaches to allocating extra monies. We also observed that evidence coverage was very uneven and variable with some heavily used interventions being well frequently supported (e.g. hospital at home) and yet other, equally utilised responses being based on hardly any evidence.

Taken as a whole the evidence base, as briefly reviewed under different intervention types in relation to discharge planning and integrated care more generally, confirms the research priorities identified in a recent scoping review. Across the range of interventions and initiatives we found a dearth of studies that explored patient, family and provider needs and experiences particularly during the development and implementation of initiatives aimed at improving delayed discharges. This lack has similarly been highlighted in a review of patient and caregiver experience of delayed discharge. These perspectives are typically gathered in relation to experiencing the issues rather than gauging the response to specific interventions. The exploration of discharge to assess is a notable exception to this. As a consequence of this omission the patient and carer perspective is often excluded from the development and documentation of best practice.

We also observed a shortage of studies that measured impact of initiatives and interventions over a long-term. The evidence base performs more strongly in terms of promising “candidate” interventions than it does in terms of tried and tested approaches. As a consequence short-term results may appear promising, while the drivers are apparent and the staff committed to change. However, little evidence is available on longer term sustainability. Not only is the evaluation period short but the evaluation frame is narrow with a focus on more readily observable targeted change rather than on the wider impact across other sectors such as primary and community care. In this connection, our evidence synthesis extends the exclusive hospital focus of the scoping review^3^ to include primary and community care, voluntary agencies, social care and, specifically, integrated care. However, in relation to integrated care, as mentioned previously, our review had to effect an artificial separation between initiatives that harnessed some degree of integration at any of a number of levels (grouped by setting) and those explicitly badged under “integrated care”. The value of the research priorities identified for each intervention derives more from the collective picture rather than any one set of recommendations.

The evidence base sees increasing recognition that hospital avoidance and reduction in delayed discharge is a “wicked” whole system problem and therefore any lens of evaluation benefits from being targeted at interventions that are implemented and integrated across sectors. However our evidence summaries extend beyond the relatively well-trodden sectors of hospital, primary care and home and community care) to include other parties including voluntary agencies, the private sector and agencies such as housing and the fire brigade. Such an approach is essential if one is to “get at the root of the problem and ensure the implementation of an initiative in one setting does not simply shift the problem to another”.

Similarly our evidence summaries seeks to address the suggestion to include “initiatives that are more upstream in nature (eg, hospital admission avoidance, emergency department diversion and delivery models that proactively address the health and social care needs of individuals in community settings)”.

#### Stratified evidence priorities

Specific research gaps are identified under each intervention category in the taxonomy. For convenience these are listed in categories below.

Main priorities are documented in Table 84.

**Table 84.**
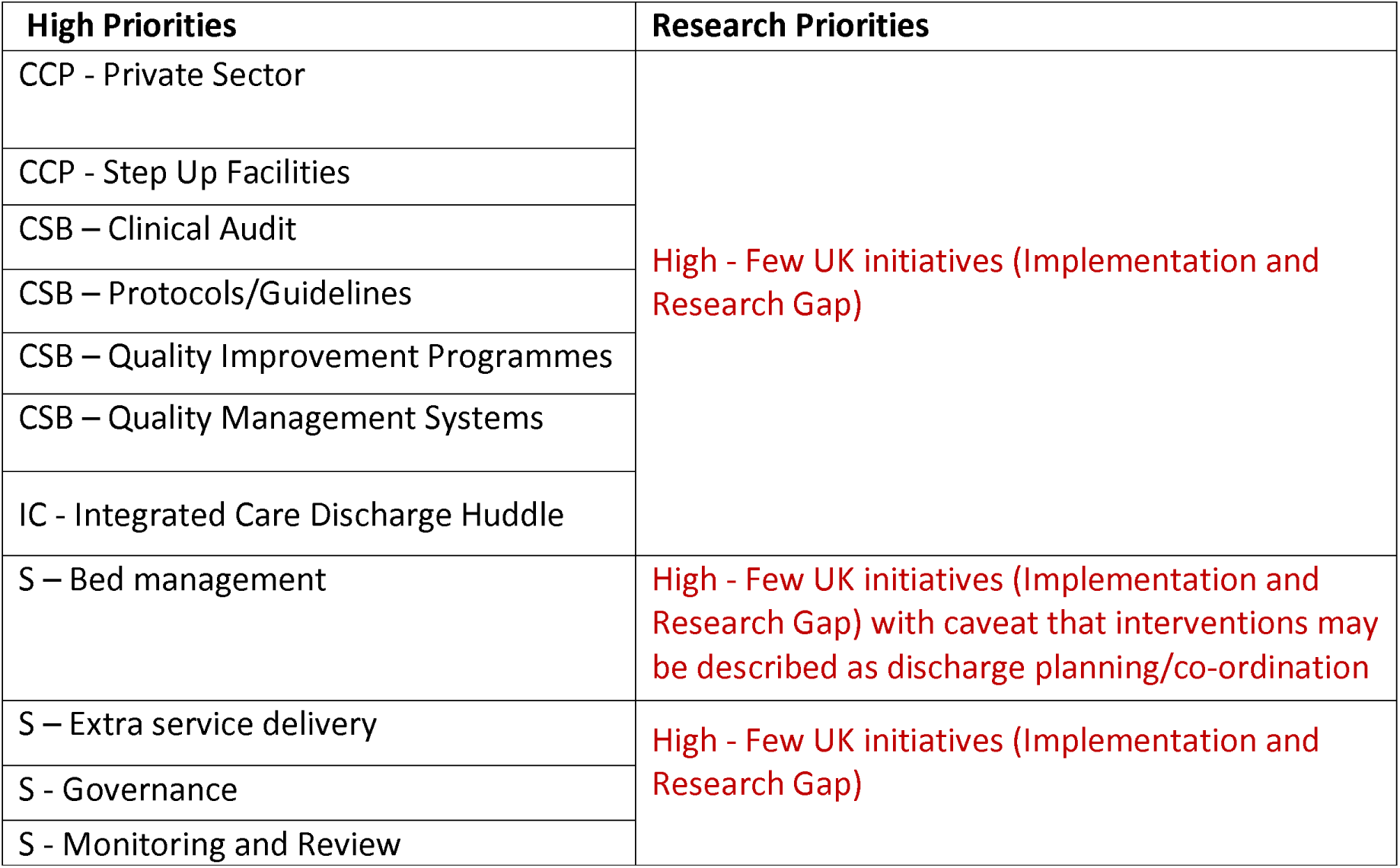
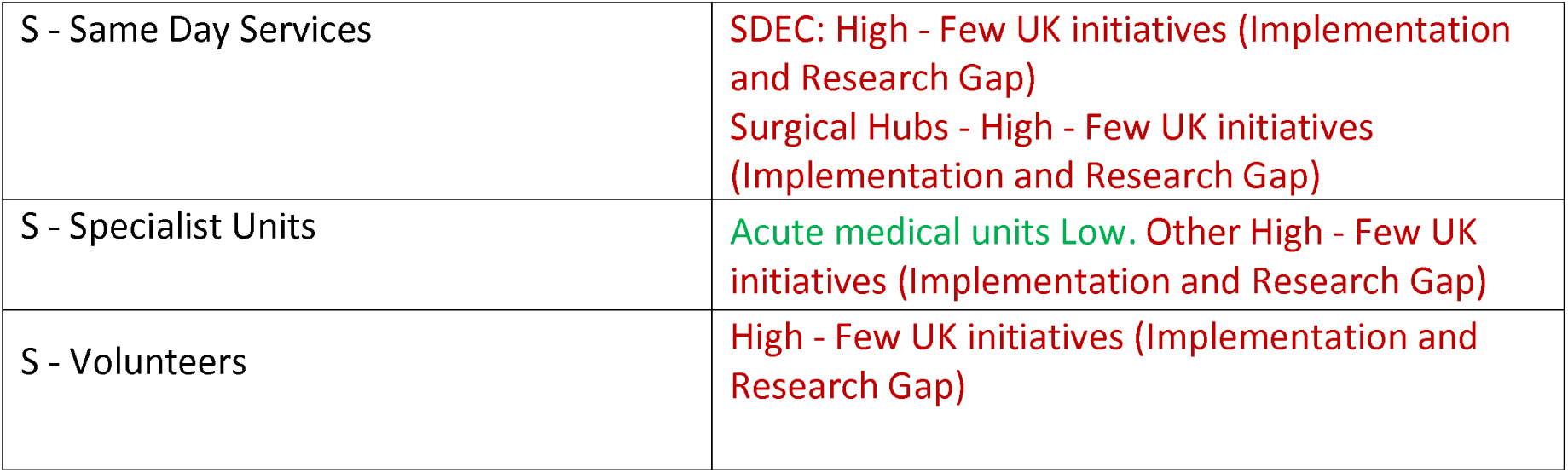
High Priorities (Implementation and Research Gap)

Other priorities are documented in Tables 85 and 86.

**Table 85.**
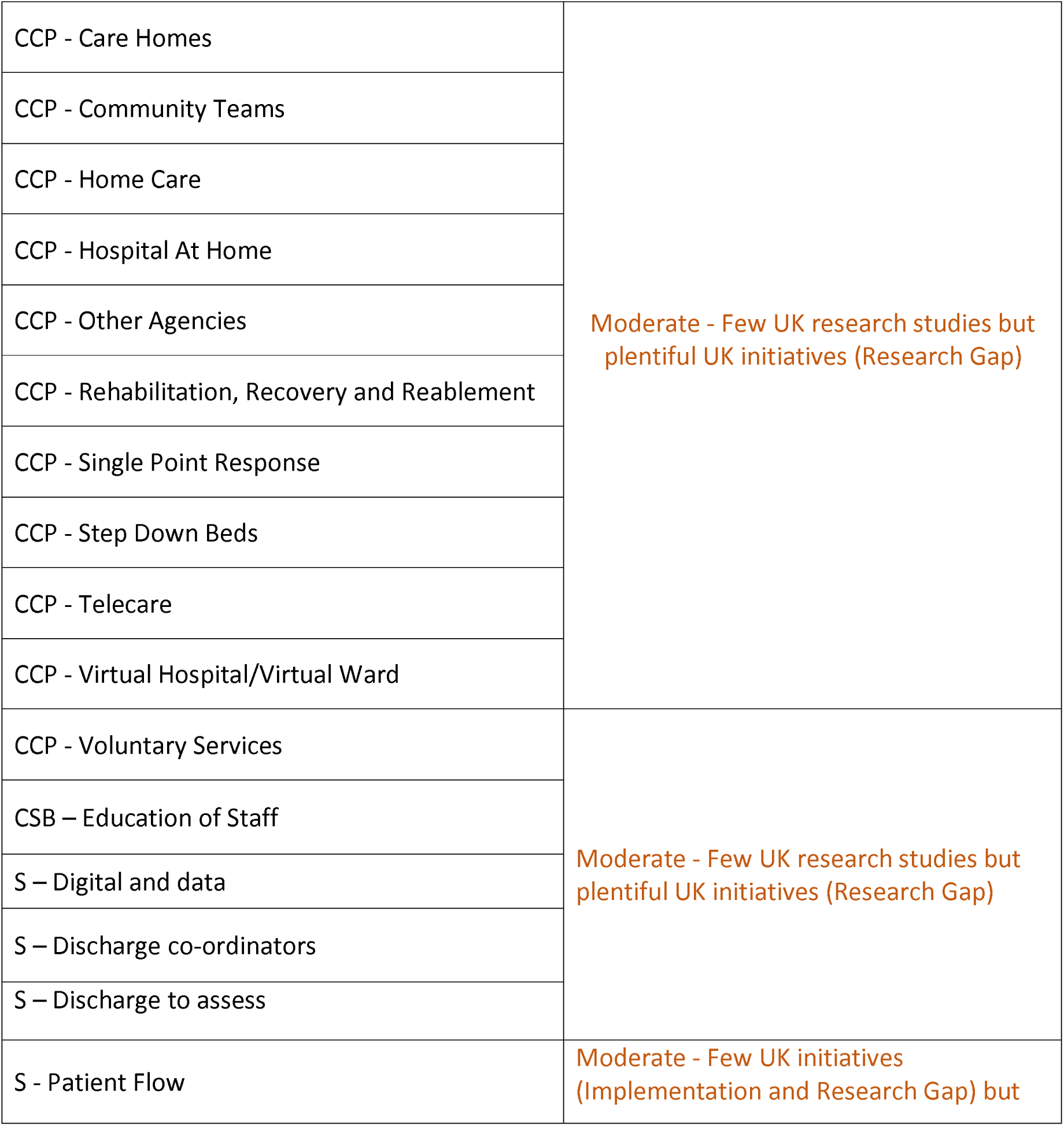
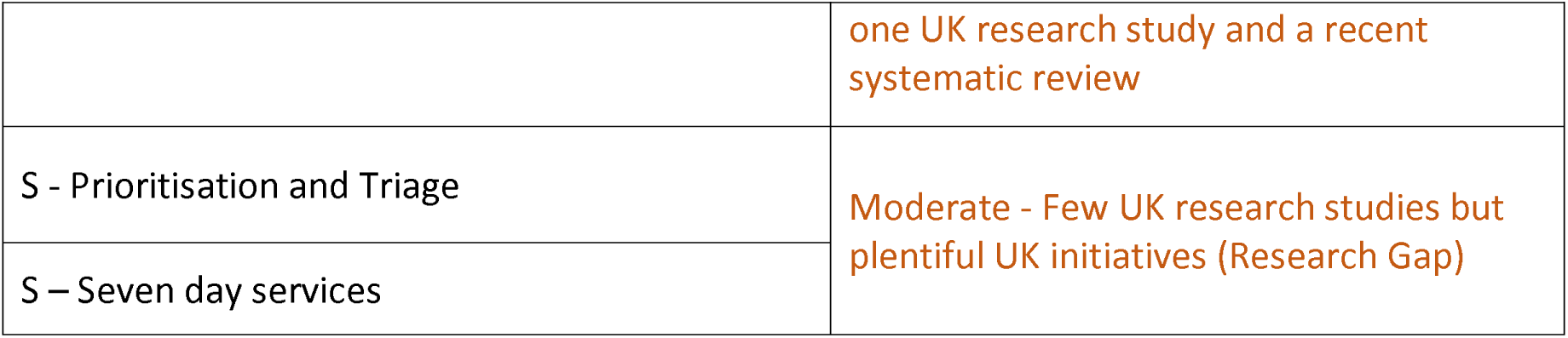
Moderate Priorities (Research Gap)

**Table 86.** Moderate Priorities (Synthesis Gap)

|  |  |
| --- | --- |
|  | **Research Priorities** |
| CCP - Rapid Response/See and Treat | Moderate - No/poor Systematic Review but plentiful international research studies (Synthesis Gap) |

Lowest priorities include CCP - Hospital Avoidance; CCP- Social Care; IC – Integrated Care and S – Specialist Units - Acute medical units where the research priority for all four is: Low - Good quality Systematic Review(s) and plentiful UK research studies (No Gap). S – Policies is difficult to assign a level of priority as most policies will cover one or more headings from elsewhere in the taxonomy.

### Towards a realist review

The scoping review, published in 2021, concludes by identifying a potential need for “future research to consider a realist review of the literature on delayed hospital discharge to understand the context, mechanisms of impact, outcomes and theories of change, given that addressing a delayed discharge is a complex problem”. Our own exploration has confirmed both this need and its potential contribution. In providing brief summaries of how the individual interventions and initiatives might contribute to relieving winter pressures we have identified certain common underpinning mechanisms, such as reassurance of the older person, establishment of clinical need, provision of a second/specialist opinion, transfer of venue etcetera that could be operationalised well within a realist review.

We would add that rather than focus on a “single sector and reactive approach to addressing delayed discharge” our evidence summaries provide initial justification for including upstream anticipatory approaches viewed holistically across the widest possible array of sectors as they contribute within a whole system evaluation frame. However, it must be noted that, to remain both feasible and productive, the holistic lens required will necessitate a clear targeting and prioritisation of a focus for synthesis. The potential contribution of a realist approach is further attested to by identification of an existing realist synthesis in the specific area of intermediate care^133^.

### Strengths and Limitations

This review benefitted from ongoing stakeholder involvement throughout the project to ensure that the deliverable is closely targeted to the requirements of the commissioner. Winter pressures is a broad topic that runs the full gamut of public health and health service provision from upstream public health approaches through to delayed discharge and overwhelmed capacity of social care provision. Detailed consultation helped to focus on the specific issues that have been found to be of strategic importance, as blockages to the entire system, that relate to health services reconfiguration and that command ongoing press and public interest and concern. Therefore, a strength of this report is its specific focus on discharge arrangements and on integrated care.

However, it soon became apparent that health service and social care utilisation takes place within a complex whole system. Focusing attention only a model that assumes admission as an inpatient as a default fails to capitalise on developments such as same day services that seek to bypass any form of overnight stay. Furthermore, assuming presentation to an emergency department fails to take into account utilisation of other points of access, particularly those that aim at community based hospital avoidance. A strength of this review is its facility to extend beyond hospital-based interventions, a limitation openly acknowledged for previous scoping reviews. Indeed, inclusion of a significant body of community and voluntary service initiatives moves much closer to the concept of integrated care, although not necessarily badged as such. Broadening the coverage of the taxonomy, even given exclusion of any workforce or modelling interventions, necessarily holds consequences in the inevitable trade-off between breadth and depth. The review team was only able to focus on indicative items of evidence, namely systematic reviews and major UK studies. Between these items of evidence, privileged for rigour and relevance respectively, and the many identified case studies, privileged for currency and creativity, lies an extensive unacknowledged supply of studies exploring various aspects of admission avoidance, hospital discharge and integrated care provision. To this extent the picture afforded by the individual intervention summaries is more a guide to evidence availability rather than a comprehensive snapshot of research activity. We encourage any research team that follows up any individual intervention summary to invest significant time in developing a more comprehensive picture of that topic. Nevertheless, the purpose of this report is to indicate promising areas for further research activity and so does not require a comparably comprehensive approach to a review that identifies effective interventions.

This review was conducted by three experienced reviewers with a collective portfolio across multiple health services and delivery review topics. This expertise was particularly important in this context because of the premium that the methodology placed on rapid identification and summary of indicative items – a particular strength of all three individuals. The three reviewers possessed the skillset to evaluate and appraise individual reviews and studies as identified but the rapid timescale and broad coverage of the review meant that this could not be performed formally and comprehensively for all retrieved items and could only be carried out opportunistically as reading of specific individual studies took place. The short timescale also meant that the team had to identify a collective point at which to draw a line under identification of further initiatives and to move on to taking stock of items already found and documenting their characteristics. In addition, time spent on each intervention type could not be standardised and so variability in coverage and depth reflects both different levels of research activity and different degrees of ease of study identification.

Finally, we must acknowledge the limitations of the taxonomy. In seeking to characterise each intervention within at least one place within the taxonomy we have often simplified their characteristics. Interventions are frequently multi-component and therefore typically include multiple headings from the taxonomy. In a few cases, where interventions possessed multiple defining characteristics or where discrete evidence on the intervention was less plentiful we would duplicate entries across the taxonomy. However, our general intention was to try to make the intervention categories self-defining and discrete; a notable exception was the cross-boundary work on hospital avoidance which necessarily appears in both structural interventions and community provision – in the former instance having the principal interpretation of admission avoidance and in the latter having the broader interpretation of acute hospital avoidance (both emergency departments and inpatient admission). In pursuing the further application of the taxonomy we would encourage indexing interventions exhaustively across multiple taxonomy headings, potentially as a prequel to further work unpacking the mechanisms that underpin multiple intervention types attesting to the likely value of a planned realist approach.

### Conclusions

We conducted a mapping review of evidence specifically related to winter pressures. It was challenging to attribute initiatives as a specific response to the winter pressures agenda, given both the lack of specificity and the artificiality of winter pressures as a binary, time-defined construct. This conception becomes progressively less useful as system pressures become a perennial issue, not a seasonal one. Few initiatives identified were specifically implemented as a response to winter pressures.

In seeking an evidence base we extended a process of evidence summaries to general initiatives used to relieve pressures on admission and discharge. Hospital at home and discharge to assess were well-supported initiatives but other responses, while also heavily used, were based on limited evidence. Across all intervention groups we confirmed the lack of studies considering patient, family and provider needs when developing interventions aimed at improving delayed discharge; identified from previous reviews of discharge and of patient and care involvement. This is a particularly important omission given the heavy dependence of formal health care systems on the contribution of informal caregivers, whether when in hospital or in the community.

Additionally, few studies measure the impact of interventions over a long time, short-term results can appear promising but evidence for longer-term sustainability is notably absent. Hospital avoidance and delayed discharge requires a whole system approach. We conclude, alongside a previous scoping review that it is imperative to consider the whole system to ensure that implementing an initiative in one setting does not simply move the problem to another setting.

### Patient and Public Involvement

Stakeholder involvement was undertaken to ensure that the review was informed by the views and experiences of stakeholders at the Department of Health and Social Care and the National Institute for Health and Care Research as well as with a standing advisory group of patients and cares.

The consultation aimed to:

- highlight the practical and personal challenges experienced by those planning or using health services during times of exceptional demand.
- to inform the second phase of our searches by identifying specific strategies, initiatives and interventions that target discharge planning and integrated care in the context of winter pressures
- to identify gaps in the literature with a view to compiling a future research agenda.

In the review proposal, we aimed to consult with stakeholders from the Department of Health and Social Care and the National Institute for Health and Care Research and a standing advisory group of patient and carers.

The key stakeholders were involved in the proposal for review and commented on a draft map when they asked for findings to be included from research.

A standing advisory group of patients and carers met once during the project to discuss the groups knowledge about different interventions that could be used to mitigate winter pressures and the research team found it especially interesting to elicit group opinions on which interventions might require research funding. The rapid nature of the project meant that discussions with the standing advisory group had to be concentrated within a single meeting. For a long project would include the standing advisory group earlier enabling them to input into the scope of the review, discuss review findings, in particular through discussing the research gaps identified.

### Equality, Diversity and Inclusion

As a secondary data study our review did not include any research participants. We were however inclusive in the studies we selected.

### Disclaimer

This project was funded by the Health and Social Care Delivery Research Programme (project number NIHR 135773) and will be published in full in the NIHR Journals Library. The views expressed are those of the author(s) and not necessarily those of the NIHR or the Department of Health and Social Care.

### Contribution of authors

Anna Cantrell (Research Fellow): Lead Reviewer, protocol development, information retrieval, study selection, data extraction, report writing.

Duncan Chambers (Research Fellow): Study selection, data extraction, report writing.

Andrew Booth (Professor in Evidence Synthesis): Methodological adviser, lead protocol developer, study selection, data extraction, report writing, guarantor of the review.

All authors commented on drafts of the protocol and report.

## Data Availability

Any additional data not included in this report and its appendices are available on request. All queries should be submitted to the corresponding author.

## Abbreviations

AA: Admission Avoidance
A&E: Accident and Emergency
ACU: Ambulatory Care Unit
AECU: Ambulatory Emergency Care Unit
BRI: Bristol Royal Infirmary
CCP: Changing Community Provision
C & RC: Care & Repair Cymru
CSB: Changing Staff Behaviour
CTT: Community Treatment team
D2A: Discharge to Assess
DHSC: Department for Health and Social Care
ED: Emergency Department
EPOC: Effective Practice and Organisation of Care
EPR: Electronic Patient Record
ESD: Early-supported discharge
FRS: Falls Response Service
GP: General Practitioner
H2HH: Hospital to a Healthier Home
HART: Hospital Avoidance Response Team
HMIC: Health Management Information Consortium
HIU: High Impact User
IC: Integrated Care
ICU: Intensive Care Unit
NICE: National Institute for Health and Care Excellence
PDSA: Plan-Do-Study-Act
PECOS: Population-Exposure-Comparative-Outcome(s)-Study Types
PPI: Patient and Public Involvement
SaTH: Shrewsbury and Telford Hospital
SDEC: Same Day Emergency care
S: Structural
SSCI: Social Sciences Citation Index
UK: United Kingdom
TC: Targeting Carers

## Acknowledgements

Ethical approval: This review did not involve the collection or analysis of any data that was not included in previously published research in the public domain. Therefore, it was exempt from formal ethical review by the University of Sheffield Ethics Committee.

Data sharing: Any additional data not included in this report and its appendices are available on request. All queries should be submitted to the corresponding author.

Information Governance: NIHR is committed to handling all personal information in line with the UK Data Protection Act (2018) and the General Data Protection Regulation (EU GDPR) 2016/679. Under Data Protection legislation NIHR is the Data Processor; the Department for Health and Social Care (DHSC) is the Data Controller, and we process personal data in accordance with their instructions. You can find out more about how we handle personal data, including how to exercise your individual rights and the contact details for DHSC’s Data Protection Officer *<u>here</u>*.

## Appendices

## Appendix 1: Medline Search Strategy

Database: Ovid MEDLINE(R) and Epub Ahead of Print, In-Process, In-Data-Review & Other Non-Indexed Citations, Daily and Versions <1946 to October 11, 2022>

Search Strategy:

--------------------------------------------------------------------------------

1. (“winter plan” or “winter planning”).tw. (7)
2. “winter pressure*”.tw. (90)
3. “winter pressures”.kw. (4)
4. “winter resilience”.tw. (1)
5. “winter surge*”.tw. (27)
6. “winter demand*”.tw. (4)
7. or/1-6 (130)
8. limit 7 to yr=”2018 - 2022” (54)
9. limit 8 to english language (54)

NB. To optimize sensitivity our experienced methodologist advised that judgements on discharge planning/integrated care context be operationalised during title/abstract and full-text screening stages rather than rely on uneven application of database indexing. Therefore we do not use specific discharge/integrated care terminology in the search strategy.

## Appendix 2: PPI Scenarios

### 1. Discharge to Assess (D2A)

Susie has recently suffered a fall and was admitted to her local general hospital for a couple of days. Her hospital has identified funding to support people to leave hospital as quickly as possible, when safe and appropriate to do so. Susie’s ongoing care and assessment will now take place out of hospital. Instead of getting a “snapshot” of her long-term needs while Susie is in hospital she will be assessed over the next six weeks while she is back in her own home.

### 2. Same Day Emergency Care (SDEC)

Asif’s GP is concerned about his dizzy spells, sometimes leading to falls. He arranges for Asif to be transported to a Same Day Emergency Care Unit located in his local hospital. Asif is assessed within a short time of his arrival at the Unit and undergoes a series of tests. At the end of the same day Asif returns to his own home with some new medication. He will be contacted for follow up if required.

Also see video at: https://youtu.be/rokhVD_1XSE

### 3. Hospital Avoidance Response Team

Dave phones 999 after a serious attack of acute bronchitis. He is put in contact with a specialist hub for severe breathing difficulties. Instead of an ambulance being sent to him he receives a visit from a paramedic. The paramedic listens to what Dave has to say about his condition. After brief contact with the specialist hub the paramedic arranges for a change in medication and a follow up visit.

### 4. Care & Repair

Rosie wants to leave hospital as soon as possible. However, her health and social care workers are concerned that her home is currently neither warm enough nor safe enough for her to return home. They contact a local charity called Care & Repair which arranges for volunteers to visit her house and prepare the house for Rosie’s return.

### 5. Telecare

Jawinta has been discharged from hospital to her care home. Her ongoing heart condition will be monitored by the hospital from a portable transmitter beside her bed. She will also have a consultation over Skype or Zoom with a diabetic nurse who will contact her online every three to four weeks.

## Footnotes

1 For the purposes of this review “integrated care” involves partnerships of organisations that come together to plan and deliver joined up health and care services (NHS England).

